# Development of a Multivariable Model for COVID-19 Risk Stratification Based on Gradient Boosting Decision Trees

**DOI:** 10.1101/2020.12.23.20248783

**Authors:** Jahir M. Gutierrez, Maksims Volkovs, Tomi Poutanen, Tristan Watson, Laura Rosella

## Abstract

**Importance:** Population stratification of the adult population in Ontario, Canada by their risk of COVID-19 complications can support rapid pandemic response, resource allocation, and decision making.

**Objective:** To develop and validate a multivariable model to predict risk of hospitalization due to COVID-19 severity from routinely collected health records of the entire adult population of Ontario, Canada.

**Design, Setting, and Participants:** This cohort study included 36,323 adult patients (age ≥ 18 years) from the province of Ontario, Canada, who tested positive for SARS-CoV-2 nucleic acid by polymerase chain reaction between February 2 and October 5, 2020, and followed up through November 5, 2020. Patients living in long-term care facilities were excluded from the analysis.

**Main Outcomes and Measures:** Risk of hospitalization within 30 days of COVID-19 diagnosis was estimated via Gradient Boosting Decision Trees, and risk factor importance was examined via Shapley values.

**Results:** The study cohort included 36,323 patients with majority female sex (18,895 [52.02%]) and median (IQR) age of 45 (31-58) years. The cohort had a hospitalization rate of 7.11% (2,583 hospitalizations) with median (IQR) time to hospitalization of 1 (0-5) days, and a mortality rate of 2.49% (906 deaths) with median (IQR) time to death of 12 (6-27) days. In contrast to patients who were not hospitalized, those who were hospitalized had a higher median age (64 years vs 43 years, p-value < 0.001), majority male (56.25% vs 47.35%, p-value<0.001), and had a higher median [IQR] number of comorbidities (3 [2-6] vs 1 [0-3], p-value<0.001). Patients were randomly split into development (n=29,058, 80%) and held-out validation (n=7,265, 20%) cohorts. The final Gradient Boosting model was built using the XGBoost algorithm and achieved high discrimination (development cohort: mean area under the receiver operating characteristic curve across the five folds of 0.852; held-out validation cohort: 0.8475) as well as excellent calibration (R^2^=0.998, slope=1.01, intercept=-0.01). The patients who scored at the top 10% in the validation cohort captured 47.41% of the actual hospitalizations, whereas those scored at the top 30% captured 80.56%. Patients in the held-out validation cohort (n=7,265) with a score of at least 0.5 (n=2,149, 29.58%) had a 20.29% hospitalization rate (positive predictive value 20.29%) compared with 2.2% hospitalization rate for those with a score less than 0.5 (n=5,116, 70.42%; negative predictive value 97.8%). Aside from age, gender and number of comorbidities, the features that most contribute to model predictions were: history of abnormal blood levels of creatinine, neutrophils and leukocytes, geography and chronic kidney disease.

**Conclusions:** A risk stratification model has been developed and validated using unique, de-identified, and linked routinely collected health administrative data available in Ontario, Canada. The final XGBoost model showed a high discrimination rate, with the potential utility to stratify patients at risk of serious COVID-19 outcomes. This model demonstrates that routinely collected health system data can be successfully leveraged as a proxy for the potential risk of severe COVID-19 complications. Specifically, past laboratory results and demographic factors provide a strong signal for identifying patients who are susceptible to complications. The model can support population risk stratification that informs patients’ protection most at risk for severe COVID-19 complications.

## Introduction

As of 5 October 2020, over 35 million cases of confirmed coronavirus disease 2019 (COVID-19) and at least 1 million deaths had been reported worldwide [1]. The COVID-19 outbreak has led to an increased demand for healthcare resources and a shortage of medical equipment and staff. Governments and healthcare organizations around the globe are currently working on containing and slowing down the spread of infections while trying to understand the risk factors associated with severe complications of COVID-19. It remains unclear which and how risk factors contribute to COVID-19 severity. Such understanding is crucial to help mitigate the healthcare system’s burden by prioritizing testing and resource allocation for those patients at the highest risk. Furthermore, now that vaccines are available [2], the ability to accurately estimate population risk can guide vaccine rollout strategies and return-to-work prioritization.

A number of diagnostic and prognostic models for COVID-19 have been developed to support medical decision making [3]. The majority of these models depend on clinical data obtained upon hospital admission (e.g. X-ray images, blood tests) as well as on demographic and medical records (age, comorbidity history) to make a prediction [4],[5],[6],[7]. Since these models can only be applied to patients already hospitalized for COVID-19, it is not possible to extend their use for the general population to identify individuals with the highest potential risk of hospitalization or death from COVID-19. Therefore, risk stratification models that depend only on historical medical records are necessary to fill this gap. Such models are particularly effective in countries with single-payer healthcare systems such as Canada, the United Kingdom and Australia since single-payer systems facilitate access to population-wide medical records. Access to extensive medical records is not limited to single-payer countries. However, databases of commercial insurance claim data are also available for large portions of the population in countries with private healthcare systems such as the United States. Consequently, we believe that with sufficient adaptation, our proposed model has wide applicability for assessing the risk of severe COVID-19 complications in the population using routinely collected data.

The province of Ontario in Canada is one of a number of jurisdictions in the world that has linked medical records on its entire population due to its single-payer health system and robust infrastructure that links all residents through a unique identifier. Analyzing the medical records of Ontario’s population is particularly interesting due to its high diversity. Almost three in ten Ontarians identify as members of visible minorities, with most of these individuals living in large metropolitan areas, such as the city of Toronto [8]. Here, we leveraged this extensive and comprehensive data source from COVID-19 positive patients in Ontario to develop a machine learning model to predict the risk of COVID-19 hospitalization. Our methodology utilizes general medical and demographic attributes commonly collected in claims data in other countries, thus facilitating its repurposing in other jurisdictions.

## Methods

### Data Source and Study Population

We obtained health administrative records from a comprehensive data repository held at ICES, a not-for-profit research institute in Ontario. To support COVID-19 research, ICES developed a high-performance infrastructure, the Health AI Data Analytics Platform (HAIDAP), as well as a continually-updated COVID-19 data resource. Using this resource, we identified all patients at least 18 years of age who were enrolled in the Ontario Health Insurance Plan (OHIP), which covers all Ontario residents, and had nasopharyngeal swabs tested for severe acute respiratory syndrome coronavirus 2 (SARS-CoV-2) between February 2, 2020, and October 5 2020 (Figure 1). Patients were followed up through November 5, 2020 to allow a follow-up period of 30 days.

**Figure 1.**
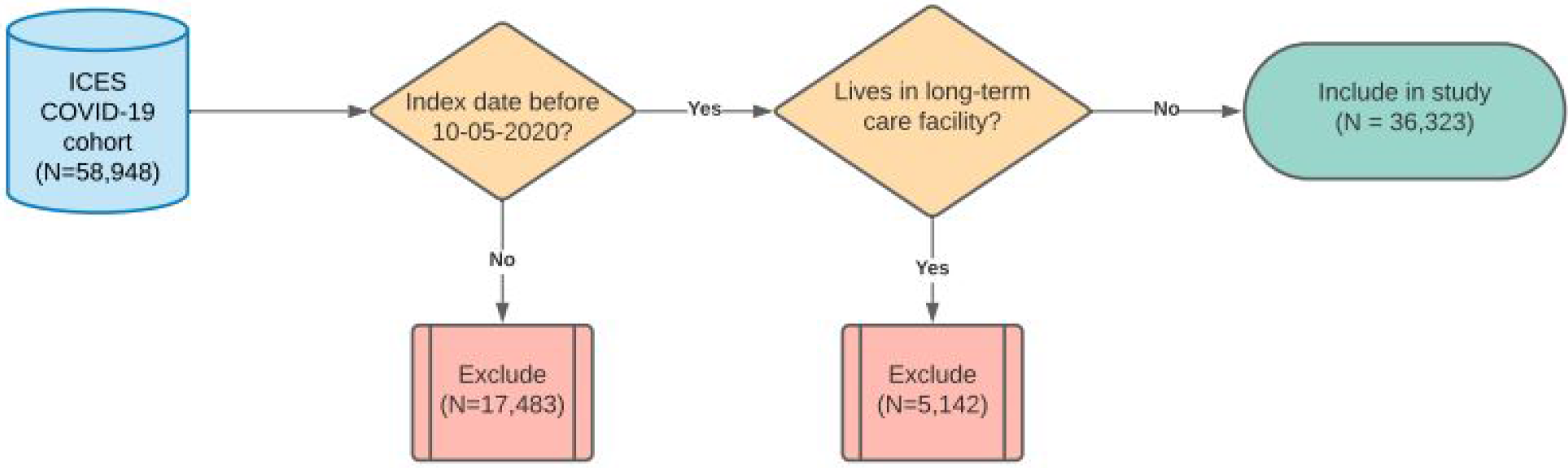
Study design. The ICES COVID-19 cohort was last updated on November 7th 2020 and it includes patients with index (diagnosis) dates between February 2 2020 and November 5th 2020. Patients with an index date after October 5, 2020 or currently living in a long-term care facility were excluded. Included patients were followed up for 30 days for the outcome of hospitalization due to COVID-19.

Individual laboratory data from the Ontario Laboratories Information System (OLIS) to relevant datasets containing healthcare use, demographic, and geographic information using unique encoders and held at ICES. It is important to note that the OLIS data captures most SARS-CoV-2 but maybe missing results from certain private laboratories in the province, which may result in discrepancies between the number of cases in our study and those officially reported [9]. This study was approved by the ICES Privacy and Compliance Office. This study followed the Transparent Reporting of a multivariable prediction model for Individual Prognosis Or Diagnosis (TRIPOD) reporting guideline [10].

### Definition of Index Date and Positive COVID-19 status

Patients were defined as positive for COVID-19 if they had one viral RNA positive polymerase chain reaction (PCR) test during the observation period. The index date for all analyses was defined as the date of the first recorded positive test.

### Adverse Outcome and Baseline Characteristics

We determined the adverse outcome as hospitalization due to COVID-19 (ICD10 code U071) at or within 30 days of the index date, since the median time to event from the index date and interquartile range (IQR) were 1 day and 0-5 days, respectively. We included baseline sociodemographic and clinical characteristics such as age, sex and comorbidity history (Table 1).

**Table 1.**
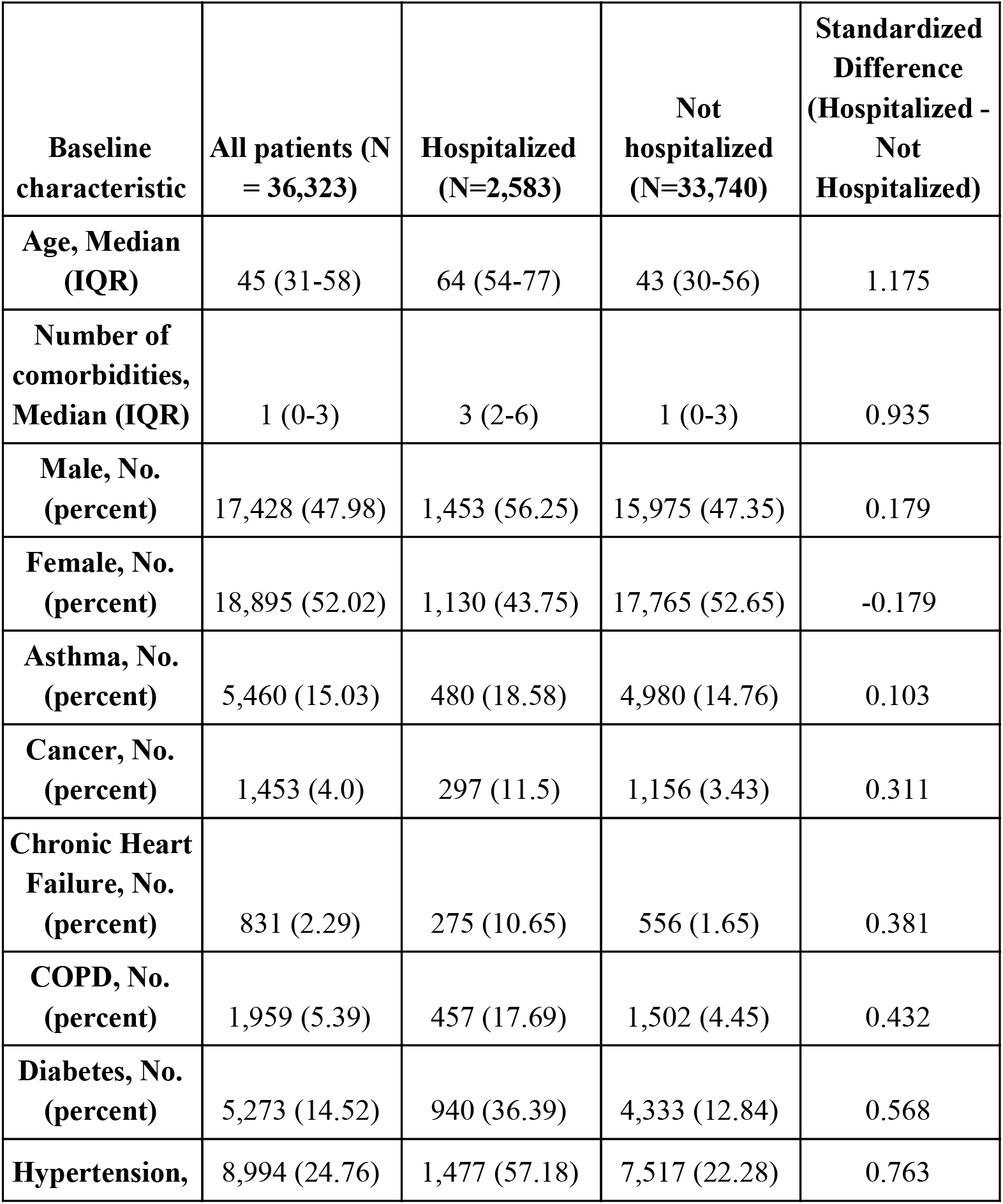
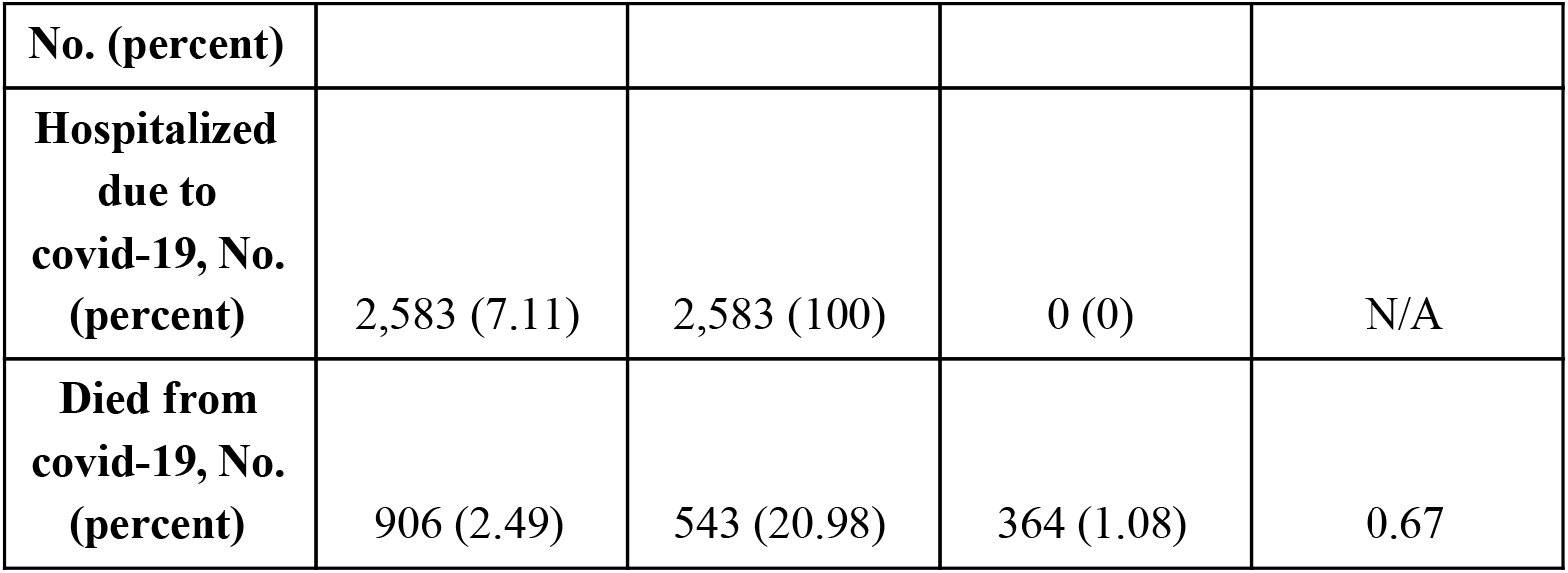
Baseline characteristics of patients included in this study.

### Model development

To ensure that only the most recent data prior to COVID-19 diagnosis were included in our model, we included medical records dated no later than 30 days prior to the index date and not earlier than 2 years before the index date (Figure 2). The 2-year window was selected due to the fact that the majority (>92%) of patients in our study cohort had at least two years of recorded clinical history in our database. Other windows were considered (3, 4, and 5 years), but we discarded them for not covering more than 90% of our patients. The 30-day buffer before the index date was applied to ensure that only historical medical records are used to make a prediction for each patient and not tests are done as a result of the COVID-19 infection.

**Figure 2.**
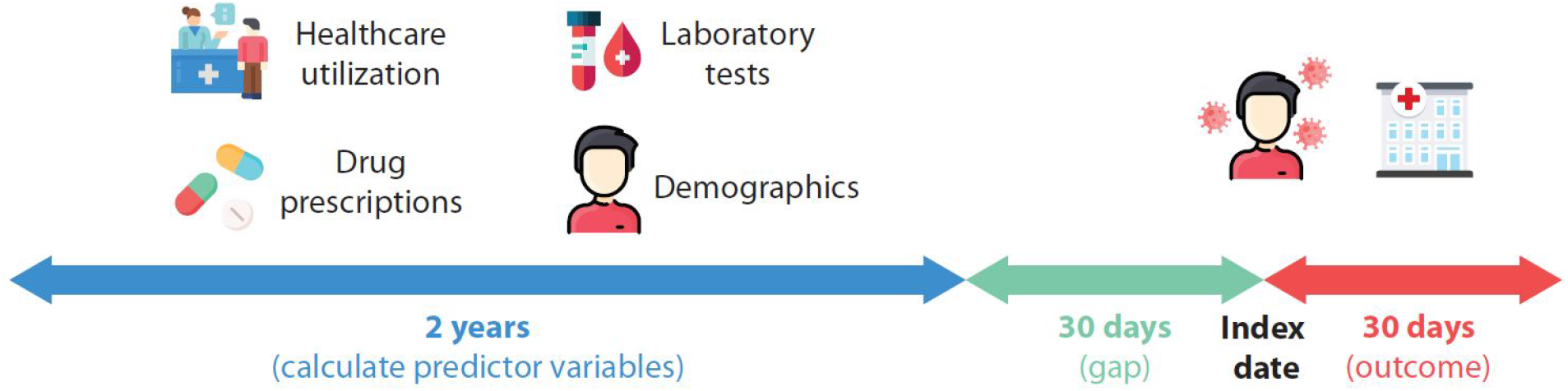
Electronic medical records used for model development. The date of COVID-19 diagnosis is used as the index date. From this date, a look-ahead period of 30 days is used to look for the outcome of hospitalization related to COVID-19. Besides including demographics, independent predictor variables were constructed by aggregating two years of medical records (e.g. past healthcare utilization, laboratory results, drug prescriptions) up to 30 days before the index date. The complete list of predictor variables calculated can be found in Supplementary Table 1.

In addition to hospitalizations, we aggregated historical records of doctor visits, outpatient services, drug prescriptions, and laboratory results for each individual. For each type of laboratory result (blood and urine), we calculated absolute deviations from normal ranges and counted the number of abnormally high or abnormally low measurements recorded in the 2-year window prior to the index date. We excluded variables with records for less than 50% of the patients in our cohort. For example, visits to a nephrologist are only recorded for those patients seeking kidney care; thus if the variable “number of visits to a nephrologist in the last 2 years” is recorded for less than 50% of the patients, then this variable would be discarded from our model. The full list of independent predictor variables extracted from the COVID-19 data source, as well as the fraction of patients lacking observations for each variable can be found in the Supplementary Table 1.

An 80%/20% random split of the dataset (where each example corresponds to one patient) was used to define development and validation sets. The validation dataset was held back and not used for model training or tuning. For the final model, we built a Gradient Boosted Trees model using the XGBoost algorithm [11]. The set of variables included in our XGBoost model were selected using a backward search approach, and hyperparameter tuning (learning rate, maximum tree depth, number of trees, alpha and gamma) was done with the grid-search algorithm to maximize the cross validation area under the receiver operating characteristic curve (AUCROC). XGBoost allows explicit handling of missing values and thus we did not perform data imputation in our model. Finally, discrimination was evaluated by using the AUCROC, in which a value of 0.5 indicates no predictive ability and 1.0 indicates perfect discrimination.

## Results

The ICES COVID-19 data source included 58,948 patients who tested positive for COVID-19 between February 2 and November 5, 2020. From these, we excluded patients with index date after October 5, 2020 as well as those patients currently living in a long-term care facility. After exclusions, 36,323 patients were included in our study cohort and followed up for 30 days (Figure 1). The hospitalization rate was 7.11% (2,583 hospitalizations) with median (IQR) time to event of 1 (0-5) day(s), and the mortality rate was 2.49% (906 deaths) with median (IQR) time to death of 12 (6-27) days after the index date. The median age of patients in the cohort was 45 years (IQR of 31-58). Table 1 shows the baseline characteristics for all patients in the cohort, and Table 2 shows the same characteristics for the development (29,058; 80%) and validation (7,265; 20%) datasets.

**Table 2.** Baseline characteristics of patients in the training and test sets.

| **Baseline characteristic** | **Development set (N=29,058)** | **Validation set (N=7,265)** | **Standardized Difference** |
| --- | --- | --- | --- |
| **Age, Median (IQR)** | 44 (31-58) | 45 (31-58) | -0.015 |
| **Number of comorbidities, Median (IQR)** | 1 (0-3) | 1 (0-3) | -0.009 |
| **Male, No. (percent)** | 13,995 (48.16) | 3,433 (47.25) | 0.018 |
| **Female, No. (percent)** | 15,063 (51.84) | 3,832 (52.75) | 0.003 |
| **Asthma, No. (percent)** | 4,376 (15.06) | 1,084 (14.92) | 0.004 |
| **Cancer, No. (percent)** | 1,163 (4) | 290 (3.99) | 0.001 |
| **Chronic Heart Failure, No. (percent)** | 668 (2.3) | 163 (2.24) | 0.004 |
| **COPD, No. (percent)** | 1,549 (5.33) | 410 (5.64) | -0.014 |
| **Diabetes, No. (percent)** | 4,202 (14.46) | 1,071 (14.74) | -0.008 |
| **Hypertension, No. (percent)** | 7,181 (24.71) | 1,813 (24.96) | -0.006 |
| **Hospitalized**<br>**due to**<br>**covid-19, No.**<br>**(percent)** | 2,043 (7.03) | 540 (7.43) | -0.016 |
| **Died from**<br>**covid-19, No.**<br>**(percent)** | 719 (2.47) | 187 (2.57) | -0.006 |

### XGBoost model

We identified 18 important predictor variables of COVID-19 hospitalization and ranked them by their Shapley contribution score which measures the impact of the variable value on the model output [12] (Table 3 and Supplementary Figure 1). These variables are age, days since the last creatinine blood test, geographical latitude, days since the last basophils blood test, gender, number of doctor visits in the last 2 years, number of comorbidities, number of different unique subclasses of drugs taken in the last 2 years, highest value of creatinine recorded in the last 2 years, number of radiology studies in the last 2 years, average value of neutrophils in blood in the last 2 years, the average value of leukocytes in blood in the last 2 years, number of creatinine blood test in the last 2 years, highest value of hemoglobin recorded in the last 2 years, history of chronic kidney disease, and days since the last mean corpuscular hemoglobin test in the last 2 years.

**Table 3.** Variables included in final XGBoost model ranked by Shapley values of importance.

| **Predictor variable** | **Shapley value** |
| --- | --- |
| Age | 0.7567 |
| Days since last creatinine blood test | 0.1320 |
| Geographical latitude | 0.1299 |
| Days since last basophils test | 0.1196 |
| Male | 0.1196 |
| Number of family doctor visits in the last 2 years | 0.1165 |
| Number of comorbidities | 0.1072 |
| Number of unique drug subclasses taken in the last 2 years | 0.0845 |
| Highest recorded level of creatinine in the last 2 years | 0.0773 |
| Number of diagnostic radiology studies received in the last 2 years | 0.0381 |
| Average measurement of neutrophils in blood in the last 2 years | 0.0289 |
| Number of doctor visits in the last 2 years | 0.0237 |
| Median level of neutrophils in the last 2 years | 0.0165 |
| Average level of leukocytes in the last 2 years | 0.0144 |
| Number of creatinine tests in the last 2 years | 0.0144 |
| Highest recorded level of hemoglobin in blood in the last 2 years | 0.0021 |
| History of chronic kidney disease | 0.0021 |
| Days since last mean corpuscular hemoglobin test | 0.0010 |

The final XGBoost model achieved high discrimination in the 5-fold cross-validation setting with a mean AUCROC of 0.852, and an AUCROC of 0.8475 in the held-out validation cohort. Figure 3A shows the ROC curve of the final model. The model also shows excellent calibration with R^2^=0.998, slope=1.01, and intercept=-0.01 (Figure 3B). Patients in the validation cohort with a score of at least 0.5 (n=2149, 29.58%) had a 20.29% hospitalization rate compared with 2.2% hospitalization rate for those patients with a score less than 0.5 (n=5116, 70.42%). Furthermore, patients in the validation cohort scored at the top 10% represent 47.41% of actual hospitalizations, and those scored at the top 30% capture 80.56% of hospitalizations (Figure 4, top bar). Interestingly, geographical latitude and laboratory test history were ranked among the top 10 predictors in our model (Supplementary Figure 1). Of note, blood biomarkers such as basophils, creatinine, and leukocytes were identified as important predictors, despite the fact that these are historical values and not measurements taken at admission [13].

**Figure 3.**
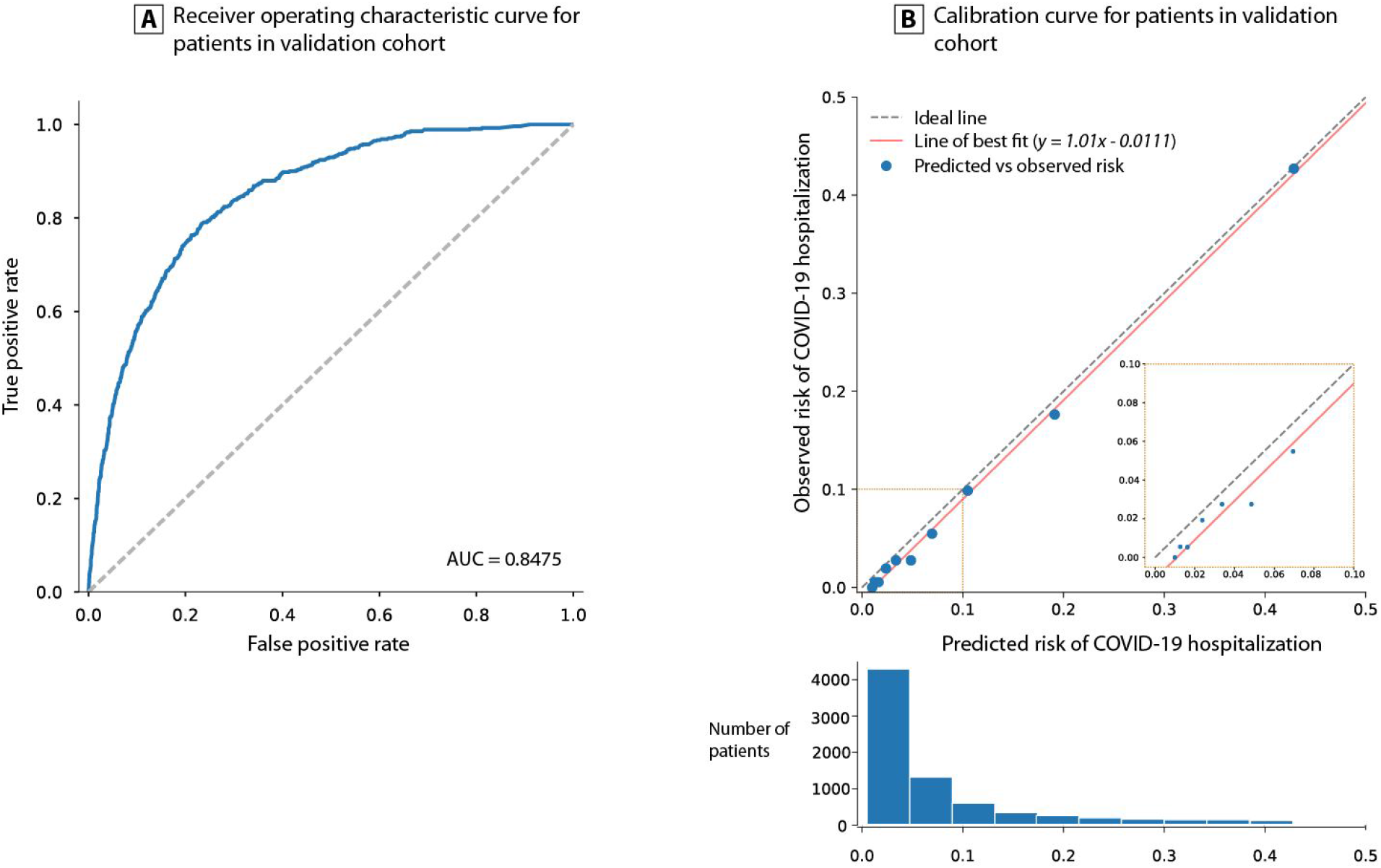
XGBoost model performance. The final model was trained with 18 features extracted from the ICES COVID-19 data source. (A) The blue line shows the receiver operating characteristic curve (ROC). (B) Calibration curve of final XGBoost model on the validation dataset. AUC: area under the curve

**Figure 4.**
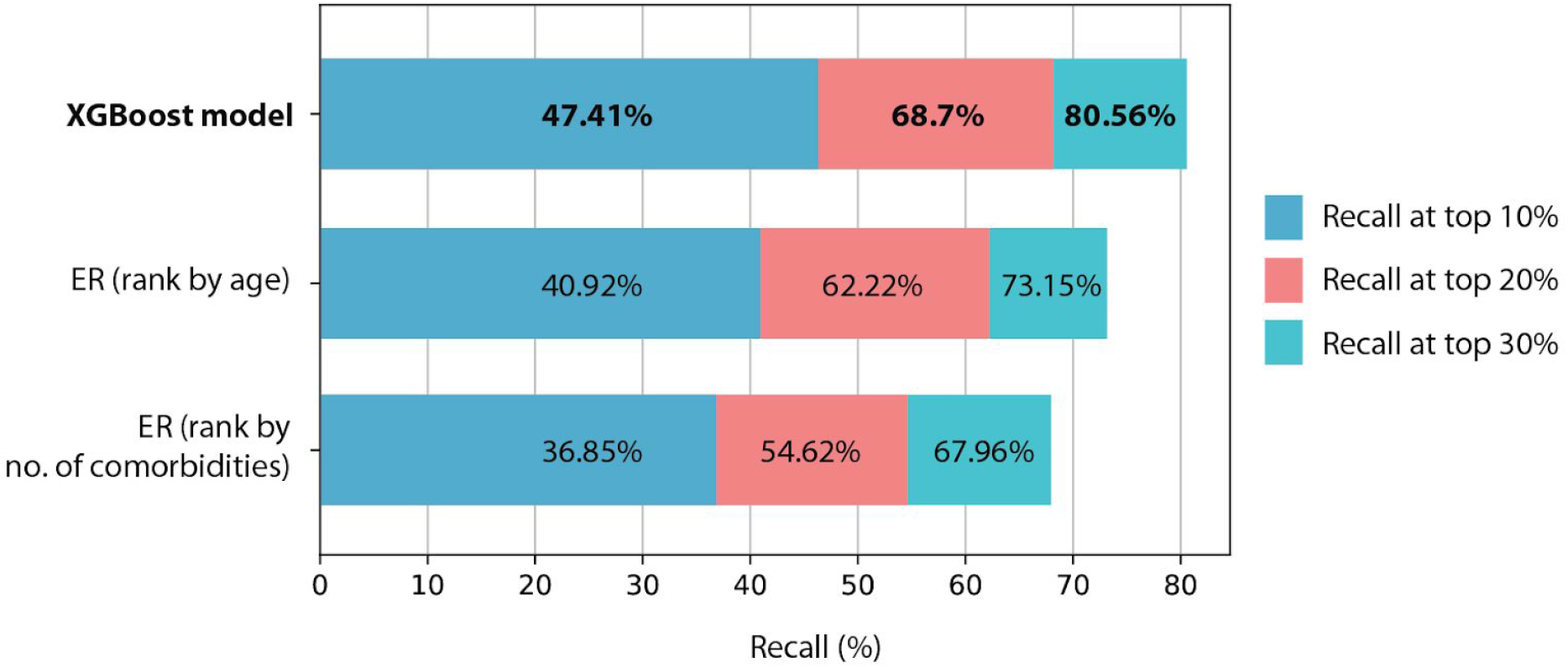
Comparison of recall at top percentiles. The final XGBoost model recall (percentage of true hospitalizations recovered in validation dataset) was compared against two empirical rules (ER).

### Comparison of XGBoost model recall against two empirical rules

After vaccines with sufficient efficacy were announced in October and November of 2020[14], governments of virtually all affected countries started to actively develop vaccine rollout and prioritization schedules[15,16]. The most prevalent approach to assessing vaccine risk is to start with the oldest patients (especially those in the long term care facilities) that account for the majority of reported deaths, and healthcare workers that have a high risk of exposure[17]. However, after these two groups, no agreement has been reached on how to proceed with the rest of the population. Two factors that commonly influence expert recommendations are age and pre-existing conditions (comorbidities)[18].

Our model provides an alternative approach to leverage machine learning to predict a risk score for every patient. These risk scores can then be used to rank patients and prioritize vaccination. To compare commonly recommended risk factors, we constructed two empirical rules and applied them to the held-out validation cohort to see how many actual hospitalizations we could capture. These two empirical rules are: (1) rank patients by age and select the oldest patients; (2) rank patients by the number of comorbidities and select patients with the most comorbidities.

We split the held-out validation cohort (n=7,265) into percentiles and calculated the recall at the top 10th (n=726), 20th (n=1,453), and 30th (n=2,180) percentiles after ranking the cohort according to our model or each empirical rule. The results of this comparison are shown in Figure 4. The final XGBoost model outperformed these rules across the top three percentiles with relative gains between 10% and 30%. These results indicate that our model can more accurately identify people at high risk of severe complications from the COVID-19 infection.

## Discussion

### Principal findings

We have developed and validated a Gradient Boosted Trees model for predicting the risk of COVID-19 hospitalization in a cohort of patients in Ontario. Our model showed excellent calibration and a high discrimination performance consistent across 5-fold cross-validation cohorts, which was comparably superior to two empirical rules. We envision our model to be deployed and utilized by public health organizations, as opposed to by clinicians directly, to effectively plan resource allocation and vaccination campaigns. Interestingly, past laboratory test results contributed to model predictions and suggests that legacy blood tests can be leveraged as a proxy for future risk of COVID-19 hospitalization. We identified past neutrophil counts in blood as a strong contributor to our model predictions. These findings are consistent with recent studies documenting the role of excessive neutrophil counts in severe COVID-19 pneumonia[19,20].

The availability of variables included in the final model is not limited to Ontario’s region, as these are variables readily available in most medical record and insurance claim databases around the world. Thus, our methodology could be extended for scoring populations and informing decision making in other jurisdictions outside of Ontario, Canada.

### Strengths and limitations

Many recently developed prognostic models for COVID-19 rely on information that must be collected post-infection or at admission into a hospital [21, 22]. A key strength of our model is that it depends only on historical medical records and demographic variables available before infection. These are variables that are routinely collected and readily available in both public and private medical claims databases used across many countries. Furthermore, Ontario has a diverse population that covers a range of population groups and thus will likely have applicability outside of Ontario or could be easily adapted to score other populations. Although future work with an external dataset would be required to validate the model performance in other geographies, we have observed that models developed on these data can usually be repurposed to other jurisdictions[23–25]. An important strength of our study is the use of Gradient Boosted trees, which allow for highly interpretable models to yield novel insights and relationships among predictor variables.

Our study has limitations. Although we have a diverse data source that captures all healthcare interactions, we are limited to some data elements that are not collected in routine data holdings. For example risk factors such as diet and physical activity associated with disease immunity [26] and not captured in our data. Furthermore, recent studies have identified genetic [27], transcriptomic [28], and proteomic [29] markers that play an important role in COVID-19 disease progression and outcome, but these data are not routinely collected at the population level and thus not included in our study. The incorporation of such factors, if available, could boost both accuracy and robustness across subgroups of patients in the population.

## Conclusions

Our model has the potential utility to directly inform public health decision making and vaccination campaigns without relying on empirical measures [30][31][32]. Our model has considerable strengths, including the ability to perform risk stratification at a population-wide level, for being based on an accurate and explainable algorithm, and finally for demonstrating the potential use of legacy laboratory data as a proxy for potential risk of severe COVID-19 complications. These risk stratification models are currently not in practice in our setting to support health system decision-making for COVID-19. We envision our model to provide a more effective way to use routinely collected data to support strategies that protect patients most at risk for serious COVID-19 complications and more careful and precise management for those at low risk.

## Data Availability

(Not applicable)

## Acknowledgements

We thank Elisa Candido and Keng-Yuan Liu for providing technical support and giving relevant advice on handling the data. The icons used in Figure 2 were made by Freepik, Monkik, and Vitaly Gorbachev from www.flaticon.com.

## Ethics

ICES has obtained ethical approval (and repeats this review tri-annually) for its privacy and security policies, procedures, and practices. Each research project that is conducted at ICES is also subject to internal ethical review by the ICES Privacy and Compliance Office. Please find attached to this submission a letter with more information regarding the ethical review and approval process for this research; please do not hesitate to contact me with any questions. ICES is a prescribed entity under section 45 of Ontario’s Personal Health Information Protection Act (PHIPA). Section 45 is the provision that enables analysis and compilation of statistical information related to the management, evaluation, and monitoring of, allocation of resources to, and planning for the health system. Section 45 authorizes health information custodians to disclose personal health information to a prescribed entity, like ICES, without consent for such purposes. Projects conducted wholly under section 45, by definition, do not require review by a Research Ethics Board. As a prescribed entity, ICES must submit to trio-annual review and approval of its privacy and security policies, procedures and practices by Ontario’s Information and Privacy Commissioner. These include policies, practices and procedures that require internal review and approval of every project by ICES’ Privacy and Compliance Office.

## TRIPOD checklist

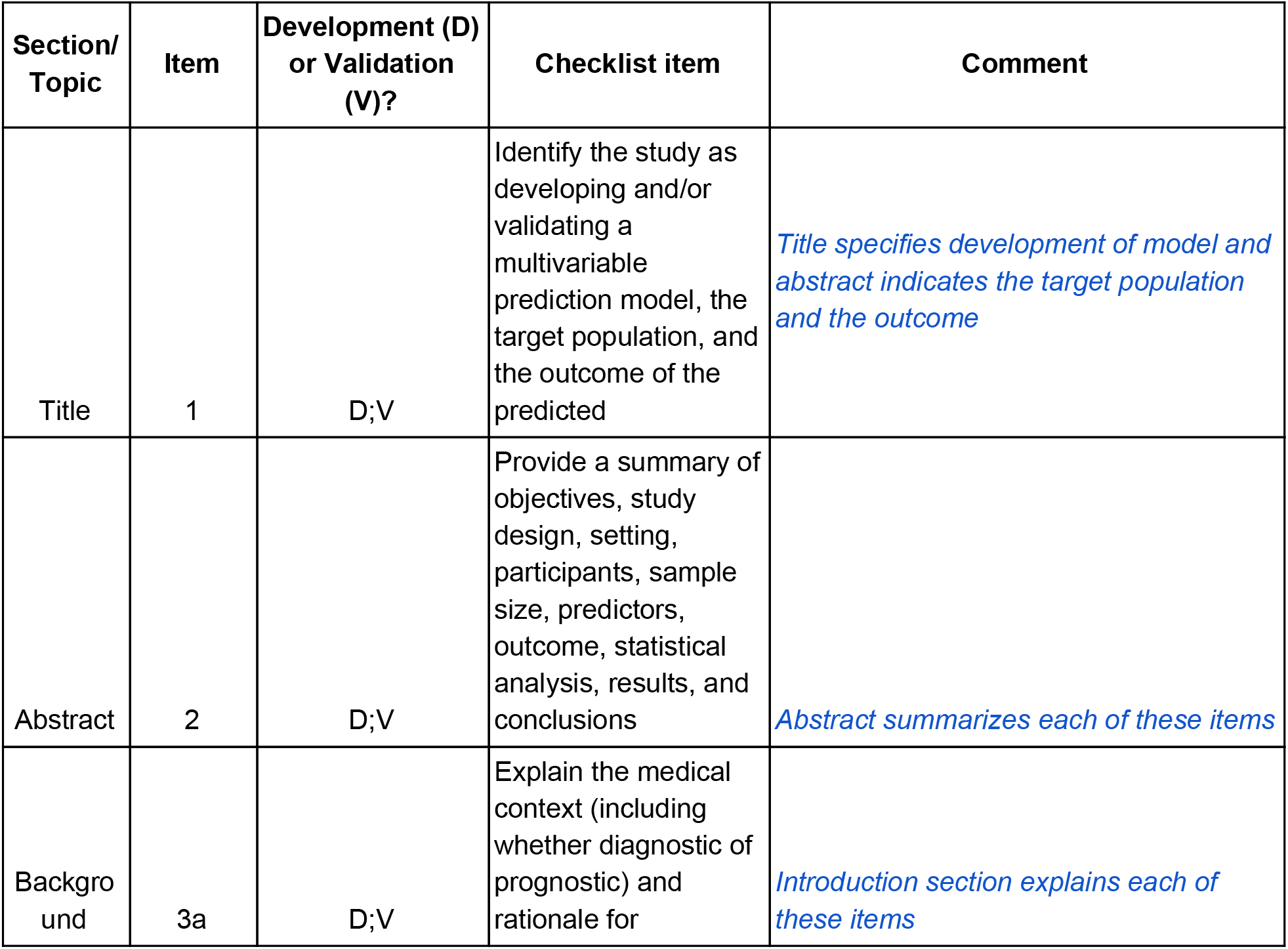
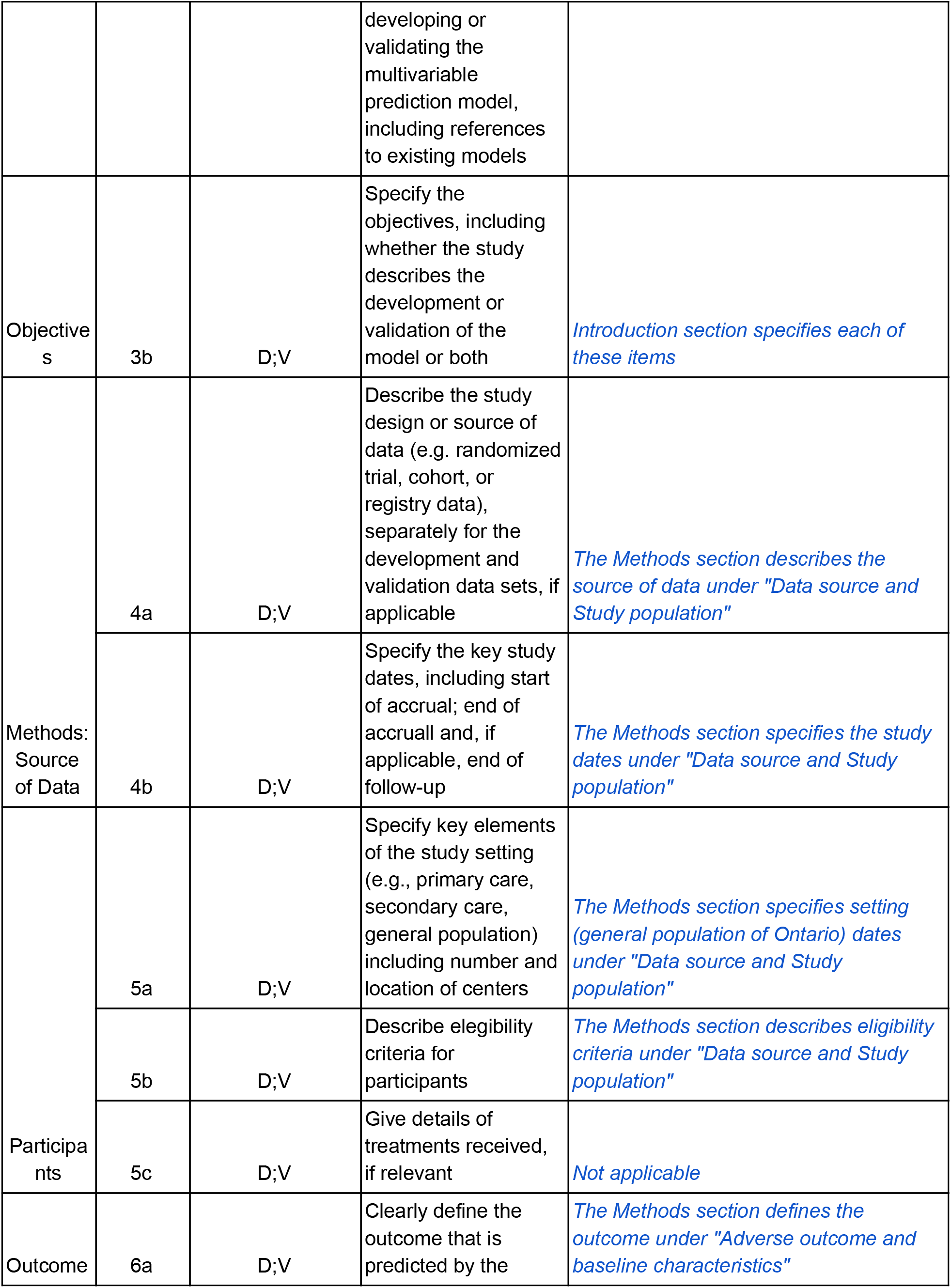
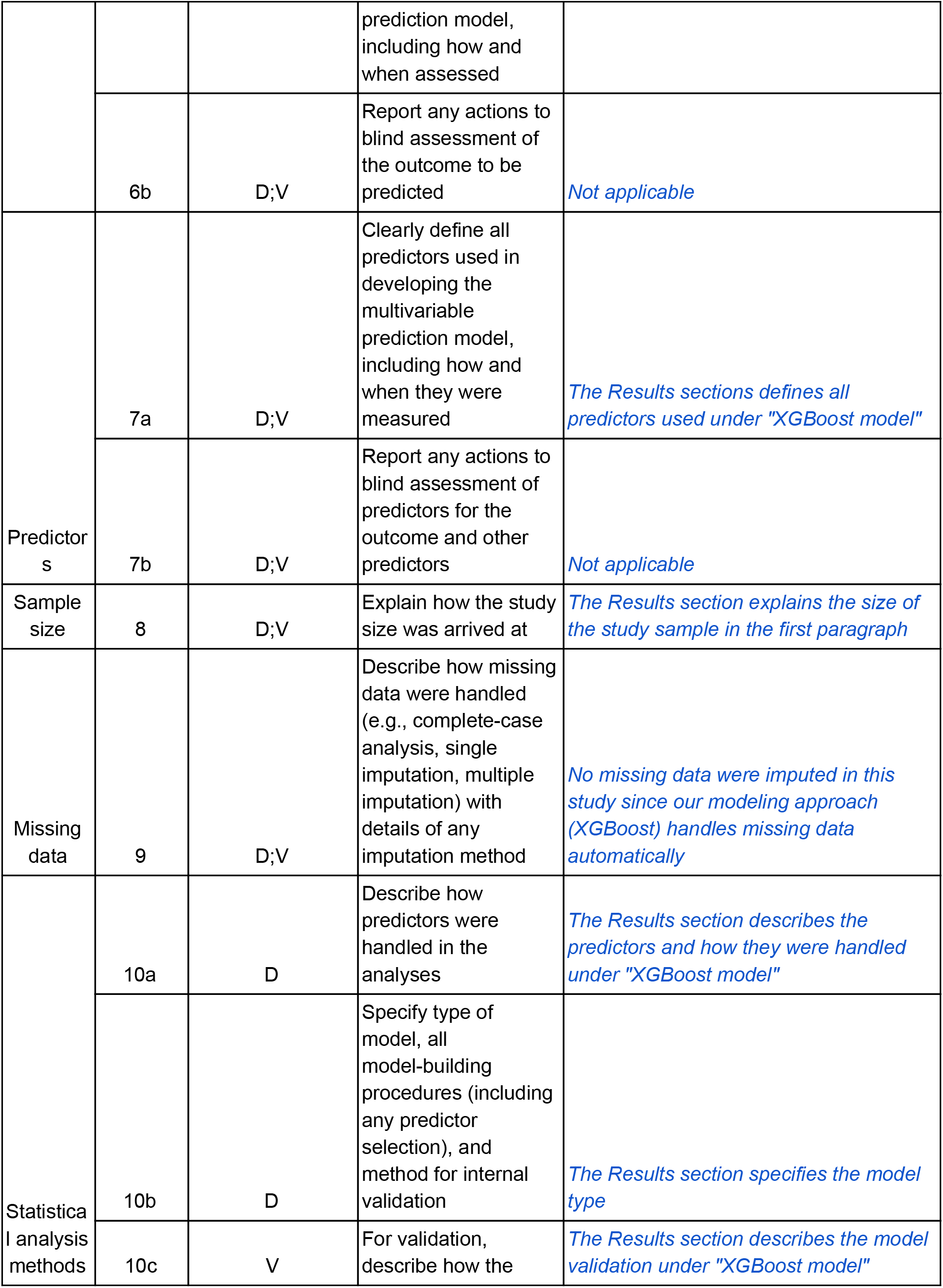
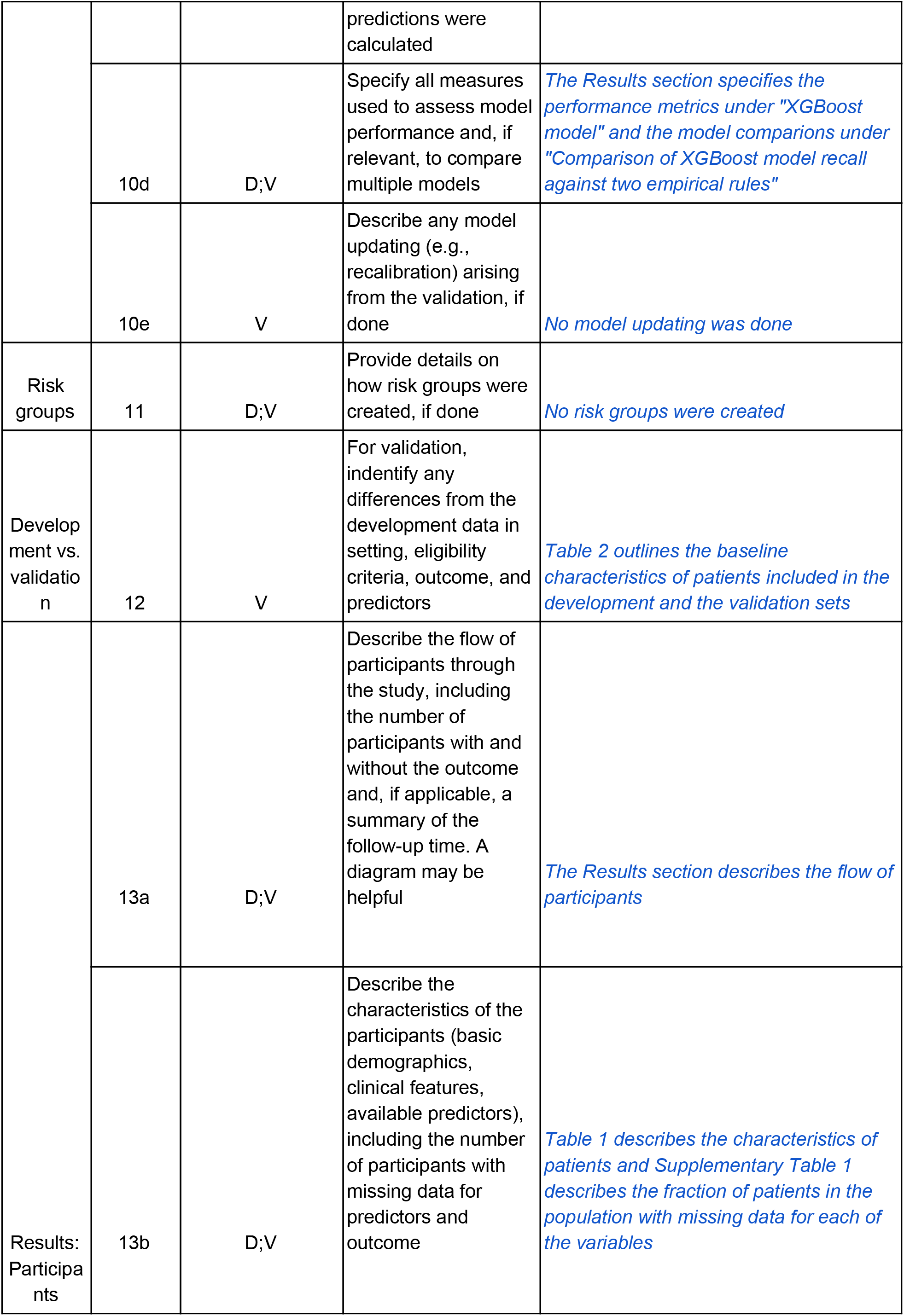
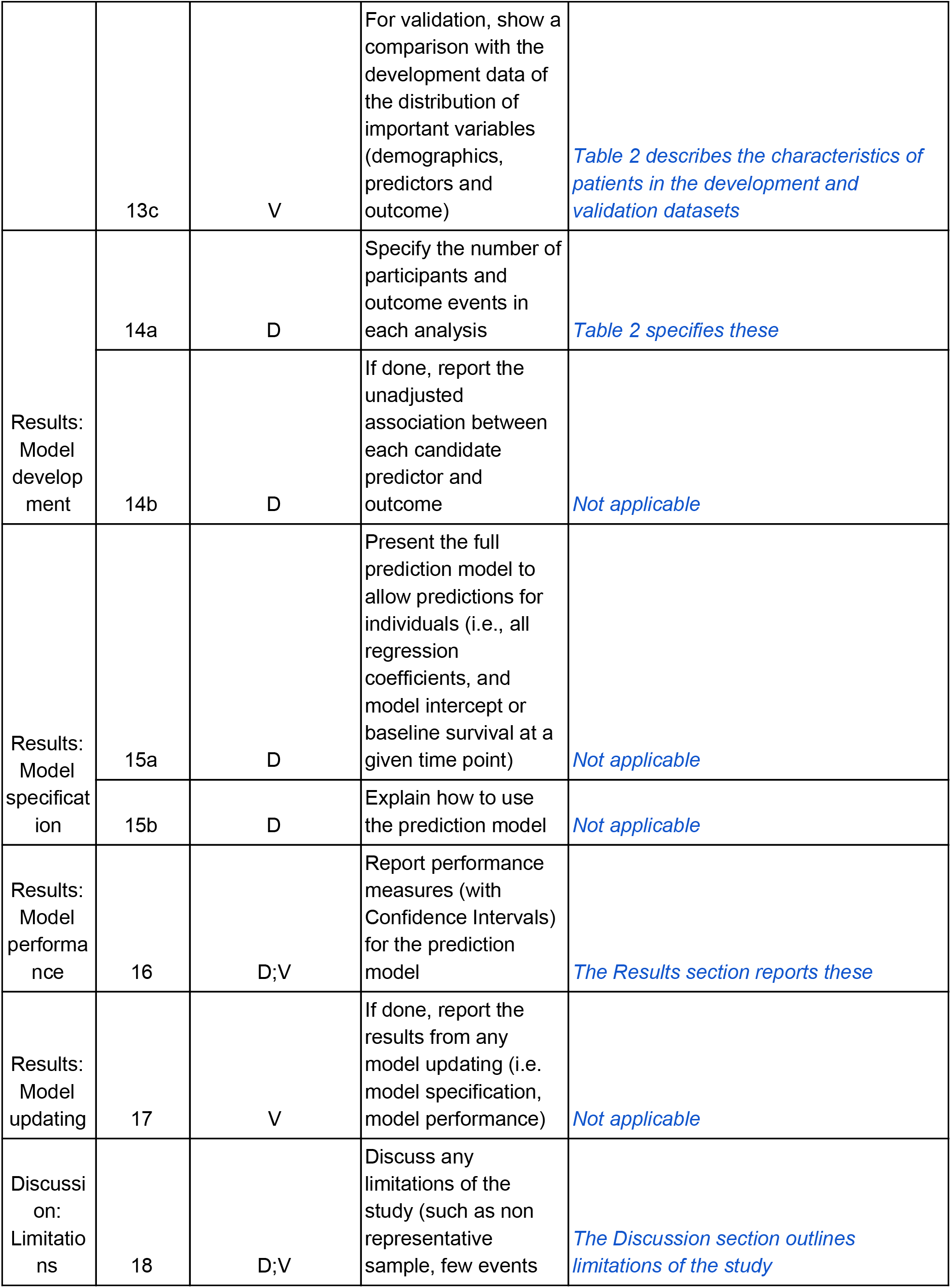
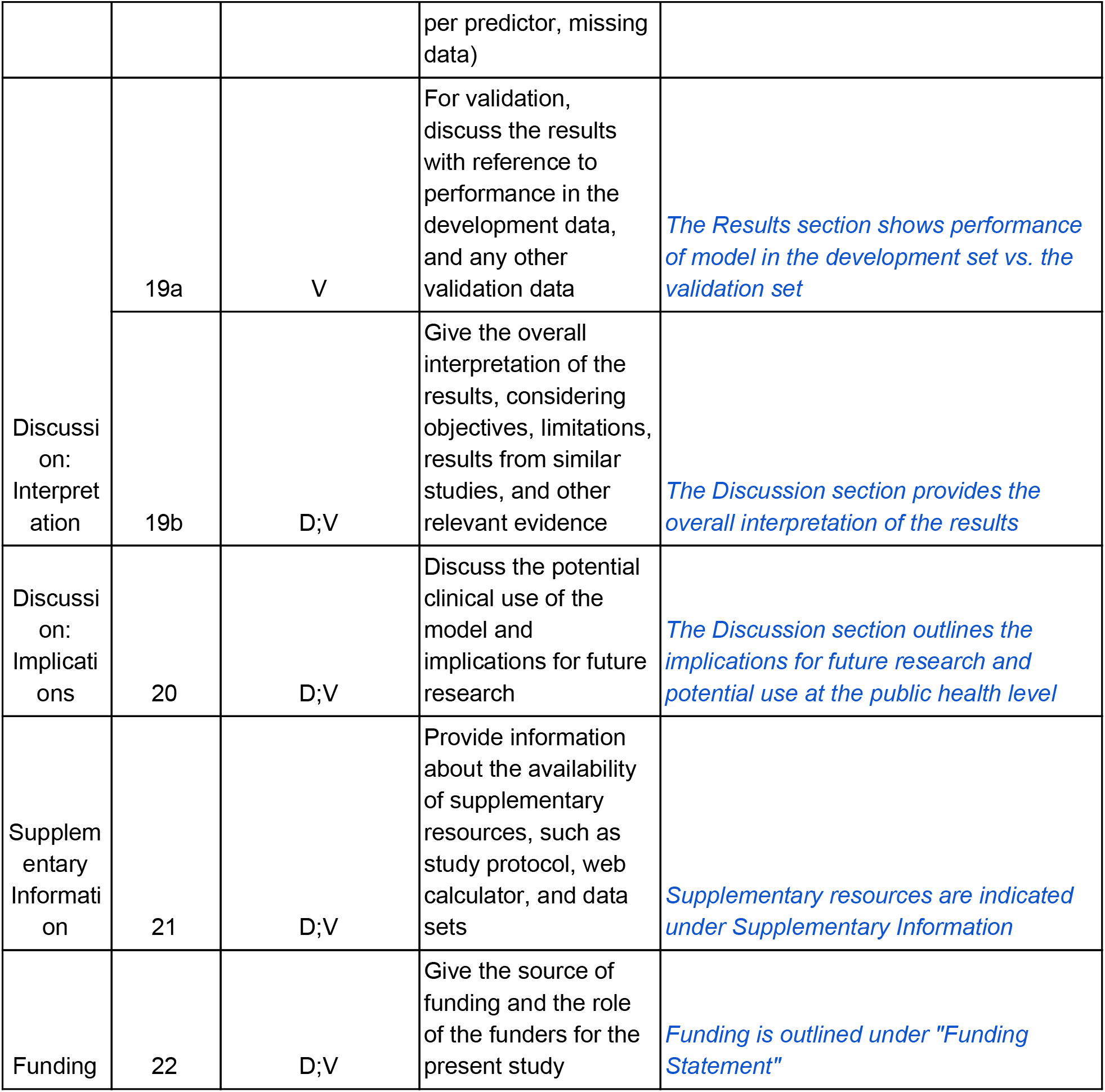

## References

1. COVID-19 Map - Johns Hopkins Coronavirus Resource Center. [cited 5 Oct 2020]. Available: https://coronavirus.jhu.edu/map.html

2. Health Canada. Pfizer-BioNTech COVID-19 vaccine: What you should know. 9 Dec 2020 [cited 11 Dec 2020]. Available: https://www.canada.ca/en/health-canada/services/drugs-health-products/covid19-industry/drugs-vaccines-treatments/vaccines/pfizer-biontech.html

3. Wynants L, Van Calster B, Collins GS, Riley RD, Heinze G, Schuit E, et al. Prediction models for diagnosis and prognosis of covid-19: systematic review and critical appraisal. BMJ. 2020;369. doi: 10.1136/bmj.m1328

4. Liang W, Yao J, Chen A, Lv Q, Zanin M, Liu J, et al. Early triage of critically ill COVID-19 patients using deep learning. Nat Commun. 2020;11: 1–7.

5. Clift AK, Coupland CAC, Keogh RH, Diaz-Ordaz K, Williamson E, Harrison EM, et al. Living risk prediction algorithm (QCOVID) for risk of hospital admission and mortality from coronavirus 19 in adults: national derivation and validation cohort study. BMJ. 2020;371. doi: 10.1136/bmj.m3731” 10.1136/bmj.m3731

6. Yan L, Zhang H-T, Goncalves J, Xiao Y, Wang M, Guo Y, et al. An interpretable mortality prediction model for COVID-19 patients. Nature Machine Intelligence. 2020;2: 283–288.

7. Knight SR, Ho A, Pius R, Buchan I, Carson G, Drake TM, et al. Risk stratification of patients admitted to hospital with covid-19 using the ISARIC WHO Clinical Characterisation Protocol: development and validation of the 4C Mortality Score. BMJ. 2020;370. doi: 10.1136/bmj.m3339

8. Government of Ontario, Ministry of Finance. 2016 CENSUS HIGHLIGHTS: Factsheet 9. [cited 11 Dec 2020]. Available: https://www.fin.gov.on.ca/en/economy/demographics/census/cenhi16-9.html

9. COVID-19 (coronavirus) in Ontario. [cited 8 Dec 2020]. Available: https://covid-19.ontario.ca/index.html

10. Tripod statement. [cited 6 Oct 2020]. Available: https://www.tripod-statement.org/

11. Chen T, Guestrin C. XGBoost: A Scalable Tree Boosting System. Proceedings of the 22nd ACM SIGKDD International Conference on Knowledge Discovery and Data Mining. New York, NY, USA: ACM; 2016. pp. 785–794.

12. Lundberg SM, Erion G, Chen H, DeGrave A, Prutkin JM, Nair B, et al. From local explanations to global understanding with explainable AI for trees. Nature Machine Intelligence. 2020;2: 56–67.

13. Ioannou GN, Locke E, Green P, Berry K, O’Hare AM, Shah JA, et al. Risk Factors for Hospitalization, Mechanical Ventilation, or Death Among 10 131 US Veterans With SARS-CoV-2 Infection. JAMA Netw Open. 2020;3: e2022310–e2022310.

14. Website. [cited 16 Dec 2020]. Available: https://www.who.int/news-room/q-a-detail/coronavirus-disease-(covid-19)-vaccines?adgroupsurvey={adgroupsurvey}&gclid=CjwKCAiA_eb-BRB2EiwAGBnXXjQv6sqIimzrxNg-lERY5E6wf3rPfy8H5_t3fXrHRqnQdSmBLSW24RoCRCYQAvD_BwE

15. Subbaraman N. Who gets a COVID vaccine first? Access plans are taking shape. Nature. 2020;585: 492–493.

16. Mullard A. How COVID vaccines are being divvied up around the world. Nature. 2020 [cited 16 Dec 2020]. doi: 10.1038/d41586-020-03370-6

17. Jeyanathan M, Afkhami S, Smaill F, Miller MS, Lichty BD, Xing Z. Immunological considerations for COVID-19 vaccine strategies. Nat Rev Immunol. 2020;20: 615–632.

18. Austen I. A Vaccine Is on Its Way to Canada. Who Will Get It First? The New York Times. 12 Dec 2020. Available: https://www.nytimes.com/2020/12/11/world/canada/vaccine-canada.html. Accessed 16 Dec 2020.

19. Wang J, Li Q, Yin Y, Zhang Y, Cao Y, Lin X, et al. Excessive Neutrophils and Neutrophil Extracellular Traps in COVID-19. Front Immunol. 2020;11. doi: 10.3389/fimmu.2020.02063

20. Arcanjo A, Logullo J, Menezes CCB, de Souza Carvalho Giangiarulo TC, dos Reis MC, de Castro GMM, et al. The emerging role of neutrophil extracellular traps in severe acute respiratory syndrome coronavirus 2 (COVID-19). Sci Rep. 2020;10: 1–11.

21. Xie J, Hungerford D, Chen H, Abrams ST, Li S, Wang G, et al. Development and external validation of a prognostic multivariable model on admission for hospitalized patients with COVID-19. medRxiv. 2020; 2020.03.28.20045997.

22. Lu J, Hu S, Fan R, Liu Z, Yin X, Wang Q, et al. ACP risk grade: a simple mortality index for patients with confirmed or suspected severe acute respiratory syndrome coronavirus 2 disease (COVID-19) during the early stage of outbreak in Wuhan, China. medRxiv. 2020; 2020.02.20.20025510.

23. Rosella LC, Manuel DG, Burchill C, Stukel TA, for the PHIAT-DM team. A population-based risk algorithm for the development of diabetes: development and validation of the Diabetes Population Risk Tool (DPoRT). Journal of Epidemiology & Community Health. 2011;65: 613–620.

24. Ng R, Sutradhar R, Kornas K, Wodchis WP, Sarkar J, Fransoo R, et al. Development and Validation of the Chronic Disease Population Risk Tool (CDPoRT) to Predict Incidence of Adult Chronic Disease. JAMA Netw Open. 2020;3: e204669–e204669.

25. Manuel DG, Perez R, Sanmartin C, Taljaard M, Hennessy D, Wilson K, et al. Measuring Burden of Unhealthy Behaviours Using a Multivariable Predictive Approach: Life Expectancy Lost in Canada Attributable to Smoking, Alcohol, Physical Inactivity, and Diet. PLoS Med. 2016;13: e1002082.

26. Lifestyle factors in the prevention of COVID-19. Global Health Journal. y2020 [cited 9 Dec 2020]. doi: 10.1016/j.glohj.2020.11.002

27. Zeberg H, Pääbo S. The major genetic risk factor for severe COVID-19 is inherited from Neanderthals. Nature. 2020;587: 610–612.

28. Longitudinal Multi-omics Analyses Identify Responses of Megakaryocytes, Erythroid Cells, and Plasmablasts as Hallmarks of Severe COVID-19. Immunity. 2020 [cited 11 Dec 2020]. doi: 10.1016/j.immuni.2020.11.017

29. Demichev V, Tober-Lau P, Nazarenko T, Thibeault C, Whitwell H, Lemke O, et al. A time-resolved proteomic and diagnostic map characterizes COVID-19 disease progression and predicts outcome. medRxiv. 2020; 2020.11.09.20228015.

30. Lurie N, Experton B. How to Leverage the Medicare Program for a COVID-19 Vaccination Campaign. JAMA. 2020 [cited 25 Nov 2020]. doi: 10.1001/jama.2020.22720

31. Experton B, Tetteh HA, Lurie N, Walker P, Carroll CJ, Elena A, et al. A Multi-Factor Risk Model for Severe Covid-19, Vaccine Prioritization and Monitoring Based on a 16 Million Medicare Cohort. medRxiv. 2020; 2020.10.28.20219816.

32. Tasker JP. Seniors, long-term care workers should be first in line for COVID-19 vaccine, committee says. In: CBC [Internet]. 4 Dec 2020 [cited 9 Dec 2020]. Available: https://www.cbc.ca/news/politics/seniors-long-term-care-workers-first-in-line-1.5828720

