## Supplemental Table 1 for "Development of a Multivariable Model for COVID-19 Risk Stratification Based on Gradient Boosting Decision Trees"

| Predictor variable | Fraction of patients without observations for variable | Decision (discard if fraction > 0.5) |
|---|---|---|
| 1-year summary (Drug prescriptions) - unique drug subclasses | 0.000000 | INCLUDE |
| 1-year summary (Doctor visits) - COUNTS | 0.000000 | INCLUDE |
| 2-year summary (Drug prescriptions) - unique drug subclasses | 0.000000 | INCLUDE |
| 2-year summary (Doctor visits) - COUNTS | 0.000000 | INCLUDE |
| (Demographics) - Age | 0.000000 | INCLUDE |
| (Demographics) - History of acute myocardial infarction | 0.000000 | INCLUDE |
| (Demographics) - History of arrhythmia | 0.000000 | INCLUDE |
| (Demographics) - History of arthritis | 0.000000 | INCLUDE |
| (Demographics) - History of asthma | 0.000000 | INCLUDE |
| (Demographics) - History of cancer | 0.000000 | INCLUDE |
| (Demographics) - History of chronic heart failure | 0.000000 | INCLUDE |
| (Demographics) - History of colitis | 0.000000 | INCLUDE |
| (Demographics) - History of chronic obstructive pulmonary disease | 0.000000 | INCLUDE |
| (Demographics) - History of coronary disease | 0.000000 | INCLUDE |
| (Demographics) - History of dementia | 0.000000 | INCLUDE |
| (Demographics) - History of diabetes | 0.000000 | INCLUDE |
| (Demographics) - History of hypertension | 0.000000 | INCLUDE |
| (Demographics) - Sex | 0.000000 | INCLUDE |
| (Demographics) - History of mental illness | 0.000000 | INCLUDE |
| (Demographics) - History of mood disorder | 0.000000 | INCLUDE |
| (Demographics) - Number comorbidities | 0.000000 | INCLUDE |
| (Demographics) - History of ostarthritis | 0.000000 | INCLUDE |
| (Demographics) - History of osteoporosis | 0.000000 | INCLUDE |
| (Demographics) - History of kidney disease | 0.000000 | INCLUDE |
| (Demographics) - History of stroke | 0.000000 | INCLUDE |
| (Geographical) - Education quantile | 0.075655 | INCLUDE |
| (Geographical) - Lives in Ontario eastern region | 0.075655 | INCLUDE |
| (Geographical) - Lives in Ontario GTHA region | 0.075655 | INCLUDE |
| (Geographical) - Lives in Toronto | 0.075655 | INCLUDE |
| (Demographics) - Marginalized dependencies indicator | 0.075655 | INCLUDE |
| (Demographics) - Marginalized deprivation indicator | 0.075655 | INCLUDE |
| (Demographics) - Marginalized ethnicity indicator | 0.075655 | INCLUDE |
| (Demographics) - Marginalized instability quintile | 0.075655 | INCLUDE |
| (Geographical) - Lives in Ontario western region | 0.075655 | INCLUDE |

| | | | |
|---|---|---|---|
| (Geographical) - Lives in Ontario northern region | | 0.075655 | INCLUDE |
| (Geographical) - LHIN number | | 0.075957 | INCLUDE |
| (Geographical) - Latitude | | 0.077829 | INCLUDE |
| (Geographical) - Longitude | | 0.077829 | INCLUDE |
| (Demographics) - Income quantile | | 0.078408 | INCLUDE |
| (Geographical) - Lives in rural community | | 0.078408 | INCLUDE |
| 2-year summary (Doctor visits) - FAMILY PRACTICE AND GENERAL PRACTICE | | 0.079812 | INCLUDE |
| 1-year summary (Doctor visits) - FAMILY PRACTICE AND GENERAL PRACTICE | | 0.168103 | INCLUDE |
| 2-year summary (Laboratory tests) - number of tests for Neutrophils | | 0.354541 | INCLUDE |
| 2-year summary (Laboratory tests) - days since earliest test for Neutrophils | | 0.354541 | INCLUDE |
| 2-year summary (Laboratory tests) - days since latest test for Neutrophils | | 0.354541 | INCLUDE |
| 2-year summary (Laboratory tests) - number of high values of Neutrophils | | 0.354541 | INCLUDE |
| 2-year summary (Laboratory tests) - highest value of Neutrophils | | 0.354541 | INCLUDE |
| 2-year summary (Laboratory tests) - number of low values of Neutrophils | | 0.354541 | INCLUDE |
| 2-year summary (Laboratory tests) - lowest value of Neutrophils | | 0.354541 | INCLUDE |
| 2-year summary (Laboratory tests) - average value of Neutrophils | | 0.354541 | INCLUDE |
| 2-year summary (Laboratory tests) - median value of Neutrophils | | 0.354541 | INCLUDE |
| 2-year summary (Laboratory tests) - count of normal tests for Neutrophils | | 0.354541 | INCLUDE |
| 2-year summary (Laboratory tests) - number of tests for Eosinophils | | 0.354872 | INCLUDE |
| 2-year summary (Laboratory tests) - days since earliest test for Eosinophils | | 0.354872 | INCLUDE |
| 2-year summary (Laboratory tests) - days since latest test for Eosinophils | | 0.354872 | INCLUDE |
| 2-year summary (Laboratory tests) - number of high values of Eosinophils | | 0.354872 | INCLUDE |
| 2-year summary (Laboratory tests) - highest value of Eosinophils | | 0.354872 | INCLUDE |
| 2-year summary (Laboratory tests) - number of low values of Eosinophils | | 0.354872 | INCLUDE |
| 2-year summary (Laboratory tests) - lowest value of Eosinophils | | 0.354872 | INCLUDE |
| 2-year summary (Laboratory tests) - average value of Eosinophils | | 0.354872 | INCLUDE |
| 2-year summary (Laboratory tests) - median value of Eosinophils | | 0.354872 | INCLUDE |
| 2-year summary (Laboratory tests) - count of normal tests for Eosinophils | | 0.354872 | INCLUDE |
| 2-year summary (Laboratory tests) - number of tests for Leukocytes | | 0.354899 | INCLUDE |
| 2-year summary (Laboratory tests) - days since earliest test for Leukocytes | | 0.354899 | INCLUDE |
| 2-year summary (Laboratory tests) - days since latest test for Leukocytes | | 0.354899 | INCLUDE |
| 2-year summary (Laboratory tests) - number of high values of Leukocytes | | 0.354899 | INCLUDE |
| 2-year summary (Laboratory tests) - highest value of Leukocytes | | 0.354899 | INCLUDE |
| 2-year summary (Laboratory tests) - number of low values of Leukocytes | | 0.354899 | INCLUDE |
| 2-year summary (Laboratory tests) - lowest value of Leukocytes | | 0.354899 | INCLUDE |
| 2-year summary (Laboratory tests) - average value of Leukocytes | | 0.354899 | INCLUDE |
| 2-year summary (Laboratory tests) - median value of Leukocytes | | 0.354899 | INCLUDE |

| | | | |
|---|---|---|---|
| 2-year summary (Laboratory tests) - count of normal tests for Leukocytes | | 0.354899 | INCLUDE |
| 2-year summary (Laboratory tests) - number of tests for Platelets | | 0.355119 | INCLUDE |
| 2-year summary (Laboratory tests) - days since earliest test for Platelets | | 0.355119 | INCLUDE |
| 2-year summary (Laboratory tests) - days since latest test for Platelets | | 0.355119 | INCLUDE |
| 2-year summary (Laboratory tests) - number of high values of Platelets | | 0.355119 | INCLUDE |
| 2-year summary (Laboratory tests) - highest value of Platelets | | 0.355119 | INCLUDE |
| 2-year summary (Laboratory tests) - number of low values of Platelets | | 0.355119 | INCLUDE |
| 2-year summary (Laboratory tests) - lowest value of Platelets | | 0.355119 | INCLUDE |
| 2-year summary (Laboratory tests) - average value of Platelets | | 0.355119 | INCLUDE |
| 2-year summary (Laboratory tests) - median value of Platelets | | 0.355119 | INCLUDE |
| 2-year summary (Laboratory tests) - count of normal tests for Platelets | | 0.355119 | INCLUDE |
| 2-year summary (Laboratory tests) - number of tests for Hemoglobin | | 0.355945 | INCLUDE |
| 2-year summary (Laboratory tests) - number of tests for Monocytes | | 0.355945 | INCLUDE |
| 2-year summary (Laboratory tests) - days since earliest test for Hemoglobin | | 0.355945 | INCLUDE |
| 2-year summary (Laboratory tests) - days since earliest test for Monocytes | | 0.355945 | INCLUDE |
| 2-year summary (Laboratory tests) - days since latest test for Hemoglobin | | 0.355945 | INCLUDE |
| 2-year summary (Laboratory tests) - days since latest test for Monocytes | | 0.355945 | INCLUDE |
| 2-year summary (Laboratory tests) - number of high values of Hemoglobin | | 0.355945 | INCLUDE |
| 2-year summary (Laboratory tests) - number of high values of Monocytes | | 0.355945 | INCLUDE |
| 2-year summary (Laboratory tests) - highest value of Hemoglobin | | 0.355945 | INCLUDE |
| 2-year summary (Laboratory tests) - highest value of Monocytes | | 0.355945 | INCLUDE |
| 2-year summary (Laboratory tests) - number of low values of Hemoglobin | | 0.355945 | INCLUDE |
| 2-year summary (Laboratory tests) - number of low values of Monocytes | | 0.355945 | INCLUDE |
| 2-year summary (Laboratory tests) - lowest value of Hemoglobin | | 0.355945 | INCLUDE |
| 2-year summary (Laboratory tests) - lowest value of Monocytes | | 0.355945 | INCLUDE |
| 2-year summary (Laboratory tests) - average value of Hemoglobin | | 0.355945 | INCLUDE |
| 2-year summary (Laboratory tests) - average value of Monocytes | | 0.355945 | INCLUDE |
| 2-year summary (Laboratory tests) - median value of Hemoglobin | | 0.355945 | INCLUDE |
| 2-year summary (Laboratory tests) - median value of Monocytes | | 0.355945 | INCLUDE |
| 2-year summary (Laboratory tests) - count of normal tests for Hemoglobin | | 0.355945 | INCLUDE |
| 2-year summary (Laboratory tests) - count of normal tests for Monocytes | | 0.355945 | INCLUDE |
| 2-year summary (Laboratory tests) - number of tests for Basophils | | 0.356110 | INCLUDE |
| 2-year summary (Laboratory tests) - days since earliest test for Basophils | | 0.356110 | INCLUDE |
| 2-year summary (Laboratory tests) - days since latest test for Basophils | | 0.356110 | INCLUDE |
| 2-year summary (Laboratory tests) - number of high values of Basophils | | 0.356110 | INCLUDE |
| 2-year summary (Laboratory tests) - highest value of Basophils | | 0.356110 | INCLUDE |
| 2-year summary (Laboratory tests) - number of low values of Basophils | | 0.356110 | INCLUDE |

| | | | |
|---|---|---|---|
| 2-year summary (Laboratory tests) - lowest value of Basophils | | 0.356110 | INCLUDE |
| 2-year summary (Laboratory tests) - average value of Basophils | | 0.356110 | INCLUDE |
| 2-year summary (Laboratory tests) - median value of Basophils | | 0.356110 | INCLUDE |
| 2-year summary (Laboratory tests) - count of normal tests for Basophils | | 0.356110 | INCLUDE |
| 2-year summary (Doctor visits) - DIAGNOSTIC RADIOLOGY | | 0.360240 | INCLUDE |
| 2-year summary (Laboratory tests) - number of tests for Creatinine | | 0.377750 | INCLUDE |
| 2-year summary (Laboratory tests) - days since earliest test for Creatinine | | 0.377750 | INCLUDE |
| 2-year summary (Laboratory tests) - days since latest test for Creatinine | | 0.377750 | INCLUDE |
| 2-year summary (Laboratory tests) - number of high values of Creatinine | | 0.377750 | INCLUDE |
| 2-year summary (Laboratory tests) - highest value of Creatinine | | 0.377750 | INCLUDE |
| 2-year summary (Laboratory tests) - number of low values of Creatinine | | 0.377750 | INCLUDE |
| 2-year summary (Laboratory tests) - lowest value of Creatinine | | 0.377750 | INCLUDE |
| 2-year summary (Laboratory tests) - average value of Creatinine | | 0.377750 | INCLUDE |
| 2-year summary (Laboratory tests) - median value of Creatinine | | 0.377750 | INCLUDE |
| 2-year summary (Laboratory tests) - count of normal tests for Creatinine | | 0.377750 | INCLUDE |
| 2-year summary (Laboratory tests) - number of tests for Erythrocyte_mean_corpuscular_hemoglobin | | 0.397462 | INCLUDE |
| 2-year summary (Laboratory tests) - days since earliest test for Erythrocyte_mean_corpuscular_hemoglobin | | 0.397462 | INCLUDE |
| 2-year summary (Laboratory tests) - days since latest test for Erythrocyte_mean_corpuscular_hemoglobin | | 0.397462 | INCLUDE |
| 2-year summary (Laboratory tests) - number of high values of Erythrocyte_mean_corpuscular_hemoglobin | | 0.397462 | INCLUDE |
| 2-year summary (Laboratory tests) - highest value of Erythrocyte_mean_corpuscular_hemoglobin | | 0.397462 | INCLUDE |
| 2-year summary (Laboratory tests) - number of low values of Erythrocyte_mean_corpuscular_hemoglobin | | 0.397462 | INCLUDE |
| 2-year summary (Laboratory tests) - lowest value of Erythrocyte_mean_corpuscular_hemoglobin | | 0.397462 | INCLUDE |
| 2-year summary (Laboratory tests) - average value of Erythrocyte_mean_corpuscular_hemoglobin | | 0.397462 | INCLUDE |
| 2-year summary (Laboratory tests) - median value of Erythrocyte_mean_corpuscular_hemoglobin | | 0.397462 | INCLUDE |
| 2-year summary (Laboratory tests) - count of normal tests for Erythrocyte_mean_corpuscular_hemoglobin | | 0.397462 | INCLUDE |
| 1-year summary (Doctor visits) - DIAGNOSTIC RADIOLOGY | | 0.546265 | DISCARD |
| 1-year summary (Laboratory tests) - number of tests for Neutrophils | | 0.630647 | DISCARD |
| 1-year summary (Laboratory tests) - days since earliest test for Neutrophils | | 0.630647 | DISCARD |
| 1-year summary (Laboratory tests) - days since latest test for Neutrophils | | 0.630647 | DISCARD |
| 1-year summary (Laboratory tests) - number of high values of Neutrophils | | 0.630647 | DISCARD |
| 1-year summary (Laboratory tests) - highest value of Neutrophils | | 0.630647 | DISCARD |
| 1-year summary (Laboratory tests) - number of low values of Neutrophils | | 0.630647 | DISCARD |
| 1-year summary (Laboratory tests) - lowest value of Neutrophils | | 0.630647 | DISCARD |
| 1-year summary (Laboratory tests) - average value of Neutrophils | | 0.630647 | DISCARD |
| 1-year summary (Laboratory tests) - median value of Neutrophils | | 0.630647 | DISCARD |
| 1-year summary (Laboratory tests) - count of normal tests for Neutrophils | | 0.630647 | DISCARD |
| 1-year summary (Laboratory tests) - number of tests for Eosinophils | | 0.630923 | DISCARD |

| | | | |
|---|---|---|---|
| 1-year summary (Laboratory tests) - days since earliest test for Eosinophils | | 0.630923 | DISCARD |
| 1-year summary (Laboratory tests) - days since latest test for Eosinophils | | 0.630923 | DISCARD |
| 1-year summary (Laboratory tests) - number of high values of Eosinophils | | 0.630923 | DISCARD |
| 1-year summary (Laboratory tests) - highest value of Eosinophils | | 0.630923 | DISCARD |
| 1-year summary (Laboratory tests) - number of low values of Eosinophils | | 0.630923 | DISCARD |
| 1-year summary (Laboratory tests) - lowest value of Eosinophils | | 0.630923 | DISCARD |
| 1-year summary (Laboratory tests) - average value of Eosinophils | | 0.630923 | DISCARD |
| 1-year summary (Laboratory tests) - median value of Eosinophils | | 0.630923 | DISCARD |
| 1-year summary (Laboratory tests) - count of normal tests for Eosinophils | | 0.630923 | DISCARD |
| 1-year summary (Laboratory tests) - number of tests for Platelets | | 0.631170 | DISCARD |
| 1-year summary (Laboratory tests) - days since earliest test for Platelets | | 0.631170 | DISCARD |
| 1-year summary (Laboratory tests) - days since latest test for Platelets | | 0.631170 | DISCARD |
| 1-year summary (Laboratory tests) - number of high values of Platelets | | 0.631170 | DISCARD |
| 1-year summary (Laboratory tests) - highest value of Platelets | | 0.631170 | DISCARD |
| 1-year summary (Laboratory tests) - number of low values of Platelets | | 0.631170 | DISCARD |
| 1-year summary (Laboratory tests) - lowest value of Platelets | | 0.631170 | DISCARD |
| 1-year summary (Laboratory tests) - average value of Platelets | | 0.631170 | DISCARD |
| 1-year summary (Laboratory tests) - median value of Platelets | | 0.631170 | DISCARD |
| 1-year summary (Laboratory tests) - count of normal tests for Platelets | | 0.631170 | DISCARD |
| 1-year summary (Laboratory tests) - number of tests for Leukocytes | | 0.631280 | DISCARD |
| 1-year summary (Laboratory tests) - days since earliest test for Leukocytes | | 0.631280 | DISCARD |
| 1-year summary (Laboratory tests) - days since latest test for Leukocytes | | 0.631280 | DISCARD |
| 1-year summary (Laboratory tests) - number of high values of Leukocytes | | 0.631280 | DISCARD |
| 1-year summary (Laboratory tests) - highest value of Leukocytes | | 0.631280 | DISCARD |
| 1-year summary (Laboratory tests) - number of low values of Leukocytes | | 0.631280 | DISCARD |
| 1-year summary (Laboratory tests) - lowest value of Leukocytes | | 0.631280 | DISCARD |
| 1-year summary (Laboratory tests) - average value of Leukocytes | | 0.631280 | DISCARD |
| 1-year summary (Laboratory tests) - median value of Leukocytes | | 0.631280 | DISCARD |
| 1-year summary (Laboratory tests) - count of normal tests for Leukocytes | | 0.631280 | DISCARD |
| 1-year summary (Laboratory tests) - number of tests for Hemoglobin | | 0.631996 | DISCARD |
| 1-year summary (Laboratory tests) - days since earliest test for Hemoglobin | | 0.631996 | DISCARD |
| 1-year summary (Laboratory tests) - days since latest test for Hemoglobin | | 0.631996 | DISCARD |
| 1-year summary (Laboratory tests) - number of high values of Hemoglobin | | 0.631996 | DISCARD |
| 1-year summary (Laboratory tests) - highest value of Hemoglobin | | 0.631996 | DISCARD |
| 1-year summary (Laboratory tests) - number of low values of Hemoglobin | | 0.631996 | DISCARD |
| 1-year summary (Laboratory tests) - lowest value of Hemoglobin | | 0.631996 | DISCARD |
| 1-year summary (Laboratory tests) - average value of Hemoglobin | | 0.631996 | DISCARD |

| | | |
|---|---|---|
| 1-year summary (Laboratory tests) - median value of Hemoglobin | 0.631996 | DISCARD |
| 1-year summary (Laboratory tests) - count of normal tests for Hemoglobin | 0.631996 | DISCARD |
| 1-year summary (Laboratory tests) - number of tests for Basophils | 0.632051 | DISCARD |
| 1-year summary (Laboratory tests) - days since earliest test for Basophils | 0.632051 | DISCARD |
| 1-year summary (Laboratory tests) - days since latest test for Basophils | 0.632051 | DISCARD |
| 1-year summary (Laboratory tests) - number of high values of Basophils | 0.632051 | DISCARD |
| 1-year summary (Laboratory tests) - highest value of Basophils | 0.632051 | DISCARD |
| 1-year summary (Laboratory tests) - number of low values of Basophils | 0.632051 | DISCARD |
| 1-year summary (Laboratory tests) - lowest value of Basophils | 0.632051 | DISCARD |
| 1-year summary (Laboratory tests) - average value of Basophils | 0.632051 | DISCARD |
| 1-year summary (Laboratory tests) - median value of Basophils | 0.632051 | DISCARD |
| 1-year summary (Laboratory tests) - count of normal tests for Basophils | 0.632051 | DISCARD |
| 1-year summary (Laboratory tests) - number of tests for Monocytes | 0.632437 | DISCARD |
| 1-year summary (Laboratory tests) - days since earliest test for Monocytes | 0.632437 | DISCARD |
| 1-year summary (Laboratory tests) - days since latest test for Monocytes | 0.632437 | DISCARD |
| 1-year summary (Laboratory tests) - number of high values of Monocytes | 0.632437 | DISCARD |
| 1-year summary (Laboratory tests) - highest value of Monocytes | 0.632437 | DISCARD |
| 1-year summary (Laboratory tests) - number of low values of Monocytes | 0.632437 | DISCARD |
| 1-year summary (Laboratory tests) - lowest value of Monocytes | 0.632437 | DISCARD |
| 1-year summary (Laboratory tests) - average value of Monocytes | 0.632437 | DISCARD |
| 1-year summary (Laboratory tests) - median value of Monocytes | 0.632437 | DISCARD |
| 1-year summary (Laboratory tests) - count of normal tests for Monocytes | 0.632437 | DISCARD |
| 1-year summary (Laboratory tests) - number of tests for Creatinine | 0.637530 | DISCARD |
| 1-year summary (Laboratory tests) - days since earliest test for Creatinine | 0.637530 | DISCARD |
| 1-year summary (Laboratory tests) - days since latest test for Creatinine | 0.637530 | DISCARD |
| 1-year summary (Laboratory tests) - number of high values of Creatinine | 0.637530 | DISCARD |
| 1-year summary (Laboratory tests) - highest value of Creatinine | 0.637530 | DISCARD |
| 1-year summary (Laboratory tests) - number of low values of Creatinine | 0.637530 | DISCARD |
| 1-year summary (Laboratory tests) - lowest value of Creatinine | 0.637530 | DISCARD |
| 1-year summary (Laboratory tests) - average value of Creatinine | 0.637530 | DISCARD |
| 1-year summary (Laboratory tests) - median value of Creatinine | 0.637530 | DISCARD |
| 1-year summary (Laboratory tests) - count of normal tests for Creatinine | 0.637530 | DISCARD |
| 2-year summary (Laboratory tests) - number of tests for Glucose | 0.641604 | DISCARD |
| 2-year summary (Laboratory tests) - days since earliest test for Glucose | 0.641604 | DISCARD |
| 2-year summary (Laboratory tests) - days since latest test for Glucose | 0.641604 | DISCARD |
| 2-year summary (Laboratory tests) - number of high values of Glucose | 0.641604 | DISCARD |
| 2-year summary (Laboratory tests) - highest value of Glucose | 0.641604 | DISCARD |

| | | | |
|---|---|---|---|
| 2-year summary (Laboratory tests) - number of low values of Glucose | | 0.641604 | DISCARD |
| 2-year summary (Laboratory tests) - lowest value of Glucose | | 0.641604 | DISCARD |
| 2-year summary (Laboratory tests) - average value of Glucose | | 0.641604 | DISCARD |
| 2-year summary (Laboratory tests) - median value of Glucose | | 0.641604 | DISCARD |
| 2-year summary (Laboratory tests) - count of normal tests for Glucose | | 0.641604 | DISCARD |
| 1-year summary (Laboratory tests) - number of tests for Erythrocyte_mean_corpuscular_hemoglobin | | 0.662060 | DISCARD |
| 1-year summary (Laboratory tests) - days since earliest test for Erythrocyte_mean_corpuscular_hemoglobin | | 0.662060 | DISCARD |
| 1-year summary (Laboratory tests) - days since latest test for Erythrocyte_mean_corpuscular_hemoglobin | | 0.662060 | DISCARD |
| 1-year summary (Laboratory tests) - number of high values of Erythrocyte_mean_corpuscular_hemoglobin | | 0.662060 | DISCARD |
| 1-year summary (Laboratory tests) - highest value of Erythrocyte_mean_corpuscular_hemoglobin | | 0.662060 | DISCARD |
| 1-year summary (Laboratory tests) - number of low values of Erythrocyte_mean_corpuscular_hemoglobin | | 0.662060 | DISCARD |
| 1-year summary (Laboratory tests) - lowest value of Erythrocyte_mean_corpuscular_hemoglobin | | 0.662060 | DISCARD |
| 1-year summary (Laboratory tests) - average value of Erythrocyte_mean_corpuscular_hemoglobin | | 0.662060 | DISCARD |
| 1-year summary (Laboratory tests) - median value of Erythrocyte_mean_corpuscular_hemoglobin | | 0.662060 | DISCARD |
| 1-year summary (Laboratory tests) - count of normal tests for Erythrocyte_mean_corpuscular_hemoglobin | | 0.662060 | DISCARD |
| 2-year summary (Laboratory tests) - number of tests for Glucose^post_CFst | | 0.667594 | DISCARD |
| 2-year summary (Laboratory tests) - days since earliest test for Glucose^post_CFst | | 0.667594 | DISCARD |
| 2-year summary (Laboratory tests) - days since latest test for Glucose^post_CFst | | 0.667594 | DISCARD |
| 2-year summary (Laboratory tests) - number of high values of Glucose^post_CFst | | 0.667594 | DISCARD |
| 2-year summary (Laboratory tests) - highest value of Glucose^post_CFst | | 0.667594 | DISCARD |
| 2-year summary (Laboratory tests) - number of low values of Glucose^post_CFst | | 0.667594 | DISCARD |
| 2-year summary (Laboratory tests) - lowest value of Glucose^post_CFst | | 0.667594 | DISCARD |
| 2-year summary (Laboratory tests) - average value of Glucose^post_CFst | | 0.667594 | DISCARD |
| 2-year summary (Laboratory tests) - median value of Glucose^post_CFst | | 0.667594 | DISCARD |
| 2-year summary (Laboratory tests) - count of normal tests for Glucose^post_CFst | | 0.667594 | DISCARD |
| 2-year summary (Doctor visits) - INTERNAL MEDICINE | | 0.703549 | DISCARD |
| 2-year summary (Laboratory tests) - number of tests for Cholesterol.in_HDL | | 0.765713 | DISCARD |
| 2-year summary (Laboratory tests) - days since earliest test for Cholesterol.in_HDL | | 0.765713 | DISCARD |
| 2-year summary (Laboratory tests) - days since latest test for Cholesterol.in_HDL | | 0.765713 | DISCARD |
| 2-year summary (Laboratory tests) - number of high values of Cholesterol.in_HDL | | 0.765713 | DISCARD |
| 2-year summary (Laboratory tests) - highest value of Cholesterol.in_HDL | | 0.765713 | DISCARD |
| 2-year summary (Laboratory tests) - number of low values of Cholesterol.in_HDL | | 0.765713 | DISCARD |
| 2-year summary (Laboratory tests) - lowest value of Cholesterol.in_HDL | | 0.765713 | DISCARD |
| 2-year summary (Laboratory tests) - average value of Cholesterol.in_HDL | | 0.765713 | DISCARD |
| 2-year summary (Laboratory tests) - median value of Cholesterol.in_HDL | | 0.765713 | DISCARD |
| 2-year summary (Laboratory tests) - count of normal tests for Cholesterol.in_HDL | | 0.765713 | DISCARD |
| 2-year summary (Laboratory tests) - number of tests for Cholesterol | | 0.766787 | DISCARD |

| | | | |
|---|---|---|---|
| 2-year summary (Laboratory tests) - days since earliest test for Cholesterol | | 0.766787 | DISCARD |
| 2-year summary (Laboratory tests) - days since latest test for Cholesterol | | 0.766787 | DISCARD |
| 2-year summary (Laboratory tests) - number of high values of Cholesterol | | 0.766787 | DISCARD |
| 2-year summary (Laboratory tests) - highest value of Cholesterol | | 0.766787 | DISCARD |
| 2-year summary (Laboratory tests) - number of low values of Cholesterol | | 0.766787 | DISCARD |
| 2-year summary (Laboratory tests) - lowest value of Cholesterol | | 0.766787 | DISCARD |
| 2-year summary (Laboratory tests) - average value of Cholesterol | | 0.766787 | DISCARD |
| 2-year summary (Laboratory tests) - median value of Cholesterol | | 0.766787 | DISCARD |
| 2-year summary (Laboratory tests) - count of normal tests for Cholesterol | | 0.766787 | DISCARD |
| 2-year summary (Doctor visits) - CARDIOLOGY | | 0.771164 | DISCARD |
| 2-year summary (Doctor visits) - OPTOMETRISTS | | 0.806954 | DISCARD |
| 1-year summary (Doctor visits) - INTERNAL MEDICINE | | 0.809102 | DISCARD |
| 1-year summary (Laboratory tests) - number of tests for Glucose | | 0.820004 | DISCARD |
| 1-year summary (Laboratory tests) - days since earliest test for Glucose | | 0.820004 | DISCARD |
| 1-year summary (Laboratory tests) - days since latest test for Glucose | | 0.820004 | DISCARD |
| 1-year summary (Laboratory tests) - number of high values of Glucose | | 0.820004 | DISCARD |
| 1-year summary (Laboratory tests) - highest value of Glucose | | 0.820004 | DISCARD |
| 1-year summary (Laboratory tests) - number of low values of Glucose | | 0.820004 | DISCARD |
| 1-year summary (Laboratory tests) - lowest value of Glucose | | 0.820004 | DISCARD |
| 1-year summary (Laboratory tests) - average value of Glucose | | 0.820004 | DISCARD |
| 1-year summary (Laboratory tests) - median value of Glucose | | 0.820004 | DISCARD |
| 1-year summary (Laboratory tests) - count of normal tests for Glucose | | 0.820004 | DISCARD |
| 1-year summary (Laboratory tests) - number of tests for Glucose^post_CFst | | 0.840735 | DISCARD |
| 1-year summary (Laboratory tests) - days since earliest test for Glucose^post_CFst | | 0.840735 | DISCARD |
| 1-year summary (Laboratory tests) - days since latest test for Glucose^post_CFst | | 0.840735 | DISCARD |
| 1-year summary (Laboratory tests) - number of high values of Glucose^post_CFst | | 0.840735 | DISCARD |
| 1-year summary (Laboratory tests) - highest value of Glucose^post_CFst | | 0.840735 | DISCARD |
| 1-year summary (Laboratory tests) - number of low values of Glucose^post_CFst | | 0.840735 | DISCARD |
| 1-year summary (Laboratory tests) - lowest value of Glucose^post_CFst | | 0.840735 | DISCARD |
| 1-year summary (Laboratory tests) - average value of Glucose^post_CFst | | 0.840735 | DISCARD |
| 1-year summary (Laboratory tests) - median value of Glucose^post_CFst | | 0.840735 | DISCARD |
| 1-year summary (Laboratory tests) - count of normal tests for Glucose^post_CFst | | 0.840735 | DISCARD |
| 2-year summary (Laboratory tests) - number of tests for Albumin/Creatinine | | 0.841147 | DISCARD |
| 2-year summary (Laboratory tests) - days since earliest test for Albumin/Creatinine | | 0.841147 | DISCARD |
| 2-year summary (Laboratory tests) - days since latest test for Albumin/Creatinine | | 0.841147 | DISCARD |
| 2-year summary (Laboratory tests) - number of high values of Albumin/Creatinine | | 0.841147 | DISCARD |
| 2-year summary (Laboratory tests) - highest value of Albumin/Creatinine | | 0.841147 | DISCARD |

| | | | |
|---|---|---|---|
| 2-year summary (Laboratory tests) - number of low values of Albumin/Creatinine | | 0.841147 | DISCARD |
| 2-year summary (Laboratory tests) - lowest value of Albumin/Creatinine | | 0.841147 | DISCARD |
| 2-year summary (Laboratory tests) - average value of Albumin/Creatinine | | 0.841147 | DISCARD |
| 2-year summary (Laboratory tests) - median value of Albumin/Creatinine | | 0.841147 | DISCARD |
| 2-year summary (Laboratory tests) - count of normal tests for Albumin/Creatinine | | 0.841147 | DISCARD |
| 1-year summary (Doctor visits) - CARDIOLOGY | | 0.859098 | DISCARD |
| 2-year summary (Doctor visits) - ANAESTHESIA | | 0.864824 | DISCARD |
| 2-year summary (Doctor visits) - OBSTETRICS AND GYNAECOLOGY | | 0.874157 | DISCARD |
| 2-year summary (Doctor visits) - EMERGENCY MEDICINE | | 0.878810 | DISCARD |
| 1-year summary (Doctor visits) - OPTOMETRISTS | | 0.879333 | DISCARD |
| 2-year summary (Drug prescriptions) - CORTICOSTEROIDS, PLAIN Days since first prescription | | 0.884976 | DISCARD |
| 2-year summary (Drug prescriptions) - CORTICOSTEROIDS, PLAIN Days since last prescription | | 0.884976 | DISCARD |
| 2-year summary (Drug prescriptions) - CORTICOSTEROIDS, PLAIN Prescription count | | 0.884976 | DISCARD |
| 1-year summary (Laboratory tests) - number of tests for Cholesterol.in_HDL | | 0.888693 | DISCARD |
| 1-year summary (Laboratory tests) - days since earliest test for Cholesterol.in_HDL | | 0.888693 | DISCARD |
| 1-year summary (Laboratory tests) - days since latest test for Cholesterol.in_HDL | | 0.888693 | DISCARD |
| 1-year summary (Laboratory tests) - number of high values of Cholesterol.in_HDL | | 0.888693 | DISCARD |
| 1-year summary (Laboratory tests) - highest value of Cholesterol.in_HDL | | 0.888693 | DISCARD |
| 1-year summary (Laboratory tests) - number of low values of Cholesterol.in_HDL | | 0.888693 | DISCARD |
| 1-year summary (Laboratory tests) - lowest value of Cholesterol.in_HDL | | 0.888693 | DISCARD |
| 1-year summary (Laboratory tests) - average value of Cholesterol.in_HDL | | 0.888693 | DISCARD |
| 1-year summary (Laboratory tests) - median value of Cholesterol.in_HDL | | 0.888693 | DISCARD |
| 1-year summary (Laboratory tests) - count of normal tests for Cholesterol.in_HDL | | 0.888693 | DISCARD |
| 1-year summary (Laboratory tests) - number of tests for Cholesterol | | 0.889079 | DISCARD |
| 1-year summary (Laboratory tests) - days since earliest test for Cholesterol | | 0.889079 | DISCARD |
| 1-year summary (Laboratory tests) - days since latest test for Cholesterol | | 0.889079 | DISCARD |
| 1-year summary (Laboratory tests) - number of high values of Cholesterol | | 0.889079 | DISCARD |
| 1-year summary (Laboratory tests) - highest value of Cholesterol | | 0.889079 | DISCARD |
| 1-year summary (Laboratory tests) - number of low values of Cholesterol | | 0.889079 | DISCARD |
| 1-year summary (Laboratory tests) - lowest value of Cholesterol | | 0.889079 | DISCARD |
| 1-year summary (Laboratory tests) - average value of Cholesterol | | 0.889079 | DISCARD |
| 1-year summary (Laboratory tests) - median value of Cholesterol | | 0.889079 | DISCARD |
| 1-year summary (Laboratory tests) - count of normal tests for Cholesterol | | 0.889079 | DISCARD |
| 2-year summary (Doctor visits) - OPHTHALMOLOGY | | 0.891501 | DISCARD |
| 2-year summary (Drug prescriptions) - VACCINES Days since first prescription | | 0.893263 | DISCARD |
| 2-year summary (Drug prescriptions) - VACCINES Days since last prescription | | 0.893263 | DISCARD |
| 2-year summary (Drug prescriptions) - VACCINES Prescription count | | 0.893263 | DISCARD |

| | | | |
|---|---|---|---|
| 2-year summary (Drug prescriptions) - NOT IDENTIFIED Days since first prescription | | 0.895190 | DISCARD |
| 2-year summary (Drug prescriptions) - NOT IDENTIFIED Days since last prescription | | 0.895190 | DISCARD |
| 2-year summary (Drug prescriptions) - NOT IDENTIFIED Prescription count | | 0.895190 | DISCARD |
| 2-year summary (Drug prescriptions) - ANTILIPEMIC: STATINS Days since first prescription | | 0.896870 | DISCARD |
| 2-year summary (Drug prescriptions) - ANTILIPEMIC: STATINS Days since last prescription | | 0.896870 | DISCARD |
| 2-year summary (Drug prescriptions) - ANTILIPEMIC: STATINS Prescription count | | 0.896870 | DISCARD |
| 2-year summary (Doctor visits) - GENERAL SURGERY | | 0.898935 | DISCARD |
| 2-year summary (Drug prescriptions) - PENICILLINS Days since first prescription | | 0.901522 | DISCARD |
| 2-year summary (Drug prescriptions) - PENICILLINS Days since last prescription | | 0.901522 | DISCARD |
| 2-year summary (Drug prescriptions) - PENICILLINS Prescription count | | 0.901522 | DISCARD |
| 2-year summary (Drug prescriptions) - ANTILIPEMIC: STATINS Average frequency (days) | | 0.901770 | DISCARD |
| 2-year summary (Drug prescriptions) - ANTILIPEMIC: STATINS Max frequency (days) | | 0.901770 | DISCARD |
| 2-year summary (Drug prescriptions) - ANTILIPEMIC: STATINS Min frequency (days) | | 0.901770 | DISCARD |
| 1-year summary (Drug prescriptions) - ANTILIPEMIC: STATINS Days since first prescription | | 0.903477 | DISCARD |
| 1-year summary (Drug prescriptions) - ANTILIPEMIC: STATINS Days since last prescription | | 0.903477 | DISCARD |
| 1-year summary (Drug prescriptions) - ANTILIPEMIC: STATINS Prescription count | | 0.903477 | DISCARD |
| 2-year summary (Hospitalizations) - Days-since-last-hospitalization | | 0.907304 | DISCARD |
| 2-year summary (Hospitalizations) - Hospitalization-counts | | 0.907304 | DISCARD |
| 2-year summary (Hospitalizations) - Hospitalization-days | | 0.907304 | DISCARD |
| 2-year summary (Doctor visits) - DERMATOLOGY | | 0.907579 | DISCARD |
| 2-year summary (Drug prescriptions) - PROTON PUMP INHIBITORS Days since first prescription | | 0.909011 | DISCARD |
| 2-year summary (Drug prescriptions) - PROTON PUMP INHIBITORS Days since last prescription | | 0.909011 | DISCARD |
| 2-year summary (Drug prescriptions) - PROTON PUMP INHIBITORS Prescription count | | 0.909011 | DISCARD |
| 1-year summary (Drug prescriptions) - ANTILIPEMIC: STATINS Average frequency (days) | | 0.909727 | DISCARD |
| 1-year summary (Drug prescriptions) - ANTILIPEMIC: STATINS Max frequency (days) | | 0.909727 | DISCARD |
| 1-year summary (Drug prescriptions) - ANTILIPEMIC: STATINS Min frequency (days) | | 0.909727 | DISCARD |
| 2-year summary (Doctor visits) - ORTHOPAEDIC SURGERY | | 0.915205 | DISCARD |
| 1-year summary (Laboratory tests) - number of tests for Albumin/Creatinine | | 0.918702 | DISCARD |
| 1-year summary (Laboratory tests) - days since earliest test for Albumin/Creatinine | | 0.918702 | DISCARD |
| 1-year summary (Laboratory tests) - days since latest test for Albumin/Creatinine | | 0.918702 | DISCARD |
| 1-year summary (Laboratory tests) - number of high values of Albumin/Creatinine | | 0.918702 | DISCARD |
| 1-year summary (Laboratory tests) - highest value of Albumin/Creatinine | | 0.918702 | DISCARD |
| 1-year summary (Laboratory tests) - number of low values of Albumin/Creatinine | | 0.918702 | DISCARD |
| 1-year summary (Laboratory tests) - lowest value of Albumin/Creatinine | | 0.918702 | DISCARD |
| 1-year summary (Laboratory tests) - average value of Albumin/Creatinine | | 0.918702 | DISCARD |
| 1-year summary (Laboratory tests) - median value of Albumin/Creatinine | | 0.918702 | DISCARD |
| 1-year summary (Laboratory tests) - count of normal tests for Albumin/Creatinine | | 0.918702 | DISCARD |

| | | | |
|---|---|---|---|
| 2-year summary (Doctor visits) - PATHOLOGY, MICROBIOLOGY, CLINICAL BIOCHEMISTRY | | 0.920464 | DISCARD |
| 1-year summary (Doctor visits) - OBSTETRICS AND GYNAECOLOGY | | 0.921647 | DISCARD |
| 2-year summary (Drug prescriptions) - NON-STEROIDAL ANTI-INFLAMMATORY: NON-ASA BASE Days since first prescription | | 0.925116 | DISCARD |
| 2-year summary (Drug prescriptions) - NON-STEROIDAL ANTI-INFLAMMATORY: NON-ASA BASE Days since last prescription | | 0.925116 | DISCARD |
| 2-year summary (Drug prescriptions) - NON-STEROIDAL ANTI-INFLAMMATORY: NON-ASA BASE Prescription count | | 0.925116 | DISCARD |
| 1-year summary (Drug prescriptions) - PROTON PUMP INHIBITORS Days since first prescription | | 0.926631 | DISCARD |
| 1-year summary (Drug prescriptions) - PROTON PUMP INHIBITORS Days since last prescription | | 0.926631 | DISCARD |
| 1-year summary (Drug prescriptions) - PROTON PUMP INHIBITORS Prescription count | | 0.926631 | DISCARD |
| 1-year summary (Doctor visits) - OPHTHALMOLOGY | | 0.926631 | DISCARD |
| 1-year summary (Drug prescriptions) - CORTICOSTEROIDS, PLAIN Days since first prescription | | 0.927511 | DISCARD |
| 1-year summary (Drug prescriptions) - CORTICOSTEROIDS, PLAIN Days since last prescription | | 0.927511 | DISCARD |
| 1-year summary (Drug prescriptions) - CORTICOSTEROIDS, PLAIN Prescription count | | 0.927511 | DISCARD |
| 1-year summary (Drug prescriptions) - VACCINES Days since first prescription | | 0.927704 | DISCARD |
| 1-year summary (Drug prescriptions) - VACCINES Days since last prescription | | 0.927704 | DISCARD |
| 1-year summary (Drug prescriptions) - VACCINES Prescription count | | 0.927704 | DISCARD |
| 2-year summary (Doctor visits) - OTOLARYNGOLOGY | | 0.928310 | DISCARD |
| 2-year summary (Drug prescriptions) - PROTON PUMP INHIBITORS Average frequency (days) | | 0.928943 | DISCARD |
| 2-year summary (Drug prescriptions) - PROTON PUMP INHIBITORS Max frequency (days) | | 0.928943 | DISCARD |
| 2-year summary (Drug prescriptions) - PROTON PUMP INHIBITORS Min frequency (days) | | 0.928943 | DISCARD |
| 1-year summary (Doctor visits) - EMERGENCY MEDICINE | | 0.929852 | DISCARD |
| 1-year summary (Doctor visits) - ANAESTHESIA | | 0.931889 | DISCARD |
| 2-year summary (Drug prescriptions) - NARCOTICS: OPIATE AGONISTS Days since first prescription | | 0.932027 | DISCARD |
| 2-year summary (Drug prescriptions) - NARCOTICS: OPIATE AGONISTS Days since last prescription | | 0.932027 | DISCARD |
| 2-year summary (Drug prescriptions) - NARCOTICS: OPIATE AGONISTS Prescription count | | 0.932027 | DISCARD |
| 2-year summary (Drug prescriptions) - CORTICOSTEROIDS, PLAIN Average frequency (days) | | 0.934174 | DISCARD |
| 2-year summary (Drug prescriptions) - CORTICOSTEROIDS, PLAIN Max frequency (days) | | 0.934174 | DISCARD |
| 2-year summary (Drug prescriptions) - CORTICOSTEROIDS, PLAIN Min frequency (days) | | 0.934174 | DISCARD |
| 1-year summary (Drug prescriptions) - NOT IDENTIFIED Days since first prescription | | 0.935908 | DISCARD |
| 1-year summary (Drug prescriptions) - NOT IDENTIFIED Days since last prescription | | 0.935908 | DISCARD |
| 1-year summary (Drug prescriptions) - NOT IDENTIFIED Prescription count | | 0.935908 | DISCARD |
| 2-year summary (Drug prescriptions) - ANALGESICS AND ANTIPYRETICS: MISCELLANEOUS Days since first prescription | | 0.938276 | DISCARD |
| 2-year summary (Drug prescriptions) - ANALGESICS AND ANTIPYRETICS: MISCELLANEOUS Days since last prescription | | 0.938276 | DISCARD |
| 2-year summary (Drug prescriptions) - ANALGESICS AND ANTIPYRETICS: MISCELLANEOUS Prescription count | | 0.938276 | DISCARD |
| 2-year summary (Doctor visits) - GASTROENTEROLOGY | | 0.939295 | DISCARD |
| 2-year summary (Doctor visits) - NEUROLOGY | | 0.939515 | DISCARD |
| 2-year summary (Doctor visits) - UROLOGY | | 0.939708 | DISCARD |
| 2-year summary (Drug prescriptions) - CALCIUM BLOCKERS Days since first prescription | | 0.940038 | DISCARD |

| | | | |
|---|---|---|---|
| 2-year summary (Drug prescriptions) - CALCIUM BLOCKERS Days since last prescription | | 0.940038 | DISCARD |
| 2-year summary (Drug prescriptions) - CALCIUM BLOCKERS Prescription count | | 0.940038 | DISCARD |
| 1-year summary (Drug prescriptions) - PROTON PUMP INHIBITORS Average frequency (days) | | 0.941194 | DISCARD |
| 1-year summary (Drug prescriptions) - PROTON PUMP INHIBITORS Max frequency (days) | | 0.941194 | DISCARD |
| 1-year summary (Drug prescriptions) - PROTON PUMP INHIBITORS Min frequency (days) | | 0.941194 | DISCARD |
| 1-year summary (Doctor visits) - GENERAL SURGERY | | 0.942103 | DISCARD |
| 2-year summary (Drug prescriptions) - ORAL ANTI-GLYCEMICS Days since first prescription | | 0.942268 | DISCARD |
| 2-year summary (Drug prescriptions) - ORAL ANTI-GLYCEMICS Days since last prescription | | 0.942268 | DISCARD |
| 2-year summary (Drug prescriptions) - ORAL ANTI-GLYCEMICS Prescription count | | 0.942268 | DISCARD |
| 2-year summary (Drug prescriptions) - CALCIUM BLOCKERS Average frequency (days) | | 0.943892 | DISCARD |
| 2-year summary (Drug prescriptions) - CALCIUM BLOCKERS Max frequency (days) | | 0.943892 | DISCARD |
| 2-year summary (Drug prescriptions) - CALCIUM BLOCKERS Min frequency (days) | | 0.943892 | DISCARD |
| 2-year summary (Drug prescriptions) - SELECTIVE BETA2-ADRENERGIC AGONISTS Days since first prescription | | 0.944140 | DISCARD |
| 2-year summary (Drug prescriptions) - SELECTIVE BETA2-ADRENERGIC AGONISTS Days since last prescription | | 0.944140 | DISCARD |
| 2-year summary (Drug prescriptions) - SELECTIVE BETA2-ADRENERGIC AGONISTS Prescription count | | 0.944140 | DISCARD |
| 2-year summary (Doctor visits) - PSYCHIATRY | | 0.944801 | DISCARD |
| 2-year summary (Drug prescriptions) - ORAL ANTI-GLYCEMICS Average frequency (days) | | 0.944828 | DISCARD |
| 2-year summary (Drug prescriptions) - ORAL ANTI-GLYCEMICS Max frequency (days) | | 0.944828 | DISCARD |
| 2-year summary (Drug prescriptions) - ORAL ANTI-GLYCEMICS Min frequency (days) | | 0.944828 | DISCARD |
| 1-year summary (Drug prescriptions) - ORAL ANTI-GLYCEMICS Days since first prescription | | 0.945792 | DISCARD |
| 1-year summary (Drug prescriptions) - ORAL ANTI-GLYCEMICS Days since last prescription | | 0.945792 | DISCARD |
| 1-year summary (Drug prescriptions) - ORAL ANTI-GLYCEMICS Prescription count | | 0.945792 | DISCARD |
| 2-year summary (Doctor visits) - ENDOCRINOLOGY | | 0.945985 | DISCARD |
| 1-year summary (Drug prescriptions) - CALCIUM BLOCKERS Days since first prescription | | 0.946122 | DISCARD |
| 1-year summary (Drug prescriptions) - CALCIUM BLOCKERS Days since last prescription | | 0.946122 | DISCARD |
| 1-year summary (Drug prescriptions) - CALCIUM BLOCKERS Prescription count | | 0.946122 | DISCARD |
| 1-year summary (Doctor visits) - DERMATOLOGY | | 0.946590 | DISCARD |
| 2-year summary (Doctor visits) - RESPIRATORY DISEASE | | 0.946645 | DISCARD |
| 2-year summary (Drug prescriptions) - MACROLIDES Days since first prescription | | 0.948105 | DISCARD |
| 2-year summary (Drug prescriptions) - MACROLIDES Days since last prescription | | 0.948105 | DISCARD |
| 2-year summary (Drug prescriptions) - MACROLIDES Prescription count | | 0.948105 | DISCARD |
| 1-year summary (Hospitalizations) - Days-since-last-hospitalization | | 0.948242 | DISCARD |
| 1-year summary (Hospitalizations) - Hospitalization-counts | | 0.948242 | DISCARD |
| 1-year summary (Hospitalizations) - Hospitalization-days | | 0.948242 | DISCARD |
| 2-year summary (Drug prescriptions) - BETA-BLOCKERS Days since first prescription | | 0.948655 | DISCARD |
| 2-year summary (Drug prescriptions) - BETA-BLOCKERS Days since last prescription | | 0.948655 | DISCARD |
| 2-year summary (Drug prescriptions) - BETA-BLOCKERS Prescription count | | 0.948655 | DISCARD |

| | | | |
|---|---|---|---|
| 2-year summary (Hospitalizations) - Class - Factors influencing health status and contact with health services (Z00-Z99) | | 0.948930 | DISCARD |
| 1-year summary (Drug prescriptions) - PENICILLINS Days since first prescription | | 0.948985 | DISCARD |
| 1-year summary (Drug prescriptions) - PENICILLINS Days since last prescription | | 0.948985 | DISCARD |
| 1-year summary (Drug prescriptions) - PENICILLINS Prescription count | | 0.948985 | DISCARD |
| 1-year summary (Drug prescriptions) - ORAL ANTI-GLYCEMICS Average frequency (days) | | 0.949013 | DISCARD |
| 1-year summary (Drug prescriptions) - ORAL ANTI-GLYCEMICS Max frequency (days) | | 0.949013 | DISCARD |
| 1-year summary (Drug prescriptions) - ORAL ANTI-GLYCEMICS Min frequency (days) | | 0.949013 | DISCARD |
| 2-year summary (Drug prescriptions) - ACE INHIBITORS Days since first prescription | | 0.949619 | DISCARD |
| 2-year summary (Drug prescriptions) - ACE INHIBITORS Days since last prescription | | 0.949619 | DISCARD |
| 2-year summary (Drug prescriptions) - ACE INHIBITORS Prescription count | | 0.949619 | DISCARD |
| 1-year summary (Drug prescriptions) - CALCIUM BLOCKERS Average frequency (days) | | 0.950224 | DISCARD |
| 1-year summary (Drug prescriptions) - CALCIUM BLOCKERS Max frequency (days) | | 0.950224 | DISCARD |
| 1-year summary (Drug prescriptions) - CALCIUM BLOCKERS Min frequency (days) | | 0.950224 | DISCARD |
| 1-year summary (Doctor visits) - ORTHOPAEDIC SURGERY | | 0.950334 | DISCARD |
| 2-year summary (Drug prescriptions) - CEPHALOSPORIN Days since first prescription | | 0.950472 | DISCARD |
| 2-year summary (Drug prescriptions) - CEPHALOSPORIN Days since last prescription | | 0.950472 | DISCARD |
| 2-year summary (Drug prescriptions) - CEPHALOSPORIN Prescription count | | 0.950472 | DISCARD |
| 2-year summary (Drug prescriptions) - CATHARTICS AND LAXATIVES Days since first prescription | | 0.950692 | DISCARD |
| 2-year summary (Drug prescriptions) - CATHARTICS AND LAXATIVES Days since last prescription | | 0.950692 | DISCARD |
| 2-year summary (Drug prescriptions) - CATHARTICS AND LAXATIVES Prescription count | | 0.950692 | DISCARD |
| 2-year summary (Drug prescriptions) - INHALED CORTICOSTEROIDS Days since first prescription | | 0.951105 | DISCARD |
| 2-year summary (Drug prescriptions) - INHALED CORTICOSTEROIDS Days since last prescription | | 0.951105 | DISCARD |
| 2-year summary (Drug prescriptions) - INHALED CORTICOSTEROIDS Prescription count | | 0.951105 | DISCARD |
| 2-year summary (Drug prescriptions) - DIURETICS Days since first prescription | | 0.951711 | DISCARD |
| 2-year summary (Drug prescriptions) - DIURETICS Days since last prescription | | 0.951711 | DISCARD |
| 2-year summary (Drug prescriptions) - DIURETICS Prescription count | | 0.951711 | DISCARD |
| 2-year summary (Drug prescriptions) - BETA-BLOCKERS Average frequency (days) | | 0.952041 | DISCARD |
| 2-year summary (Drug prescriptions) - BETA-BLOCKERS Max frequency (days) | | 0.952041 | DISCARD |
| 2-year summary (Drug prescriptions) - BETA-BLOCKERS Min frequency (days) | | 0.952041 | DISCARD |
| 2-year summary (Drug prescriptions) - DIABETES DIAGNOSTIC Days since first prescription | | 0.952096 | DISCARD |
| 2-year summary (Drug prescriptions) - DIABETES DIAGNOSTIC Days since last prescription | | 0.952096 | DISCARD |
| 2-year summary (Drug prescriptions) - DIABETES DIAGNOSTIC Prescription count | | 0.952096 | DISCARD |
| 1-year summary (Drug prescriptions) - BETA-BLOCKERS Days since first prescription | | 0.953170 | DISCARD |
| 1-year summary (Drug prescriptions) - BETA-BLOCKERS Days since last prescription | | 0.953170 | DISCARD |
| 1-year summary (Drug prescriptions) - BETA-BLOCKERS Prescription count | | 0.953170 | DISCARD |
| 1-year summary (Drug prescriptions) - ANALGESICS AND ANTIPYRETICS: MISCELLANEOUS Days since first prescription | | 0.953418 | DISCARD |
| 1-year summary (Drug prescriptions) - ANALGESICS AND ANTIPYRETICS: MISCELLANEOUS Days since last prescription | | 0.953418 | DISCARD |

| | | |
|---|---|---|
| 1-year summary (Drug prescriptions) - ANALGESICS AND ANTIPYRETICS: MISCELLANEOUS Prescription count | 0.953418 | DISCARD |
| 2-year summary (Drug prescriptions) - ACE INHIBITORS Average frequency (days) | 0.953831 | DISCARD |
| 2-year summary (Drug prescriptions) - ACE INHIBITORS Max frequency (days) | 0.953831 | DISCARD |
| 2-year summary (Drug prescriptions) - ACE INHIBITORS Min frequency (days) | 0.953831 | DISCARD |
| 2-year summary (Laboratory tests) - number of tests for Cholesterol.in_LDL | 0.955841 | DISCARD |
| 2-year summary (Laboratory tests) - days since earliest test for Cholesterol.in_LDL | 0.955841 | DISCARD |
| 2-year summary (Laboratory tests) - days since latest test for Cholesterol.in_LDL | 0.955841 | DISCARD |
| 2-year summary (Laboratory tests) - number of high values of Cholesterol.in_LDL | 0.955841 | DISCARD |
| 2-year summary (Laboratory tests) - highest value of Cholesterol.in_LDL | 0.955841 | DISCARD |
| 2-year summary (Laboratory tests) - number of low values of Cholesterol.in_LDL | 0.955841 | DISCARD |
| 2-year summary (Laboratory tests) - lowest value of Cholesterol.in_LDL | 0.955841 | DISCARD |
| 2-year summary (Laboratory tests) - average value of Cholesterol.in_LDL | 0.955841 | DISCARD |
| 2-year summary (Laboratory tests) - median value of Cholesterol.in_LDL | 0.955841 | DISCARD |
| 2-year summary (Laboratory tests) - count of normal tests for Cholesterol.in_LDL | 0.955841 | DISCARD |
| 1-year summary (Drug prescriptions) - BETA-BLOCKERS Average frequency (days) | 0.956529 | DISCARD |
| 1-year summary (Drug prescriptions) - BETA-BLOCKERS Max frequency (days) | 0.956529 | DISCARD |
| 1-year summary (Drug prescriptions) - BETA-BLOCKERS Min frequency (days) | 0.956529 | DISCARD |
| 1-year summary (Drug prescriptions) - ACE INHIBITORS Days since first prescription | 0.956639 | DISCARD |
| 1-year summary (Drug prescriptions) - ACE INHIBITORS Days since last prescription | 0.956639 | DISCARD |
| 1-year summary (Drug prescriptions) - ACE INHIBITORS Prescription count | 0.956639 | DISCARD |
| 2-year summary (Laboratory tests) - number of tests for Prostate_specific_Ag | 0.957080 | DISCARD |
| 2-year summary (Laboratory tests) - days since earliest test for Prostate_specific_Ag | 0.957080 | DISCARD |
| 2-year summary (Laboratory tests) - days since latest test for Prostate_specific_Ag | 0.957080 | DISCARD |
| 2-year summary (Laboratory tests) - number of high values of Prostate_specific_Ag | 0.957080 | DISCARD |
| 2-year summary (Laboratory tests) - highest value of Prostate_specific_Ag | 0.957080 | DISCARD |
| 2-year summary (Laboratory tests) - number of low values of Prostate_specific_Ag | 0.957080 | DISCARD |
| 2-year summary (Laboratory tests) - lowest value of Prostate_specific_Ag | 0.957080 | DISCARD |
| 2-year summary (Laboratory tests) - average value of Prostate_specific_Ag | 0.957080 | DISCARD |
| 2-year summary (Laboratory tests) - median value of Prostate_specific_Ag | 0.957080 | DISCARD |
| 2-year summary (Laboratory tests) - count of normal tests for Prostate_specific_Ag | 0.957080 | DISCARD |
| 2-year summary (Drug prescriptions) - DIURETICS Average frequency (days) | 0.957327 | DISCARD |
| 2-year summary (Drug prescriptions) - DIURETICS Max frequency (days) | 0.957327 | DISCARD |
| 2-year summary (Drug prescriptions) - DIURETICS Min frequency (days) | 0.957327 | DISCARD |
| 2-year summary (Drug prescriptions) - FLUOROQUINOLONES Days since first prescription | 0.958429 | DISCARD |
| 2-year summary (Drug prescriptions) - FLUOROQUINOLONES Days since last prescription | 0.958429 | DISCARD |
| 2-year summary (Drug prescriptions) - FLUOROQUINOLONES Prescription count | 0.958429 | DISCARD |
| 1-year summary (Drug prescriptions) - DIURETICS Days since first prescription | 0.958456 | DISCARD |

| | | | |
|---|---|---|---|
| 1-year summary (Drug prescriptions) - DIURETICS Days since last prescription | | 0.958456 | DISCARD |
| 1-year summary (Drug prescriptions) - DIURETICS Prescription count | | 0.958456 | DISCARD |
| 1-year summary (Doctor visits) - OTOLARYNGOLOGY | | 0.958484 | DISCARD |
| 1-year summary (Doctor visits) - ENDOCRINOLOGY | | 0.958759 | DISCARD |
| 1-year summary (Drug prescriptions) - NARCOTICS: OPIATE AGONISTS Days since first prescription | | 0.958869 | DISCARD |
| 1-year summary (Drug prescriptions) - NARCOTICS: OPIATE AGONISTS Days since last prescription | | 0.958869 | DISCARD |
| 1-year summary (Drug prescriptions) - NARCOTICS: OPIATE AGONISTS Prescription count | | 0.958869 | DISCARD |
| 2-year summary (Drug prescriptions) - FUNGICIDES Days since first prescription | | 0.958979 | DISCARD |
| 2-year summary (Drug prescriptions) - FUNGICIDES Days since last prescription | | 0.958979 | DISCARD |
| 2-year summary (Drug prescriptions) - FUNGICIDES Prescription count | | 0.958979 | DISCARD |
| 1-year summary (Doctor visits) - UROLOGY | | 0.959089 | DISCARD |
| 2-year summary (Doctor visits) - PLASTIC SURGERY | | 0.959695 | DISCARD |
| 1-year summary (Drug prescriptions) - NON-STEROIDAL ANTI-INFLAMMATORY: NON-ASA BASE Days since first prescription | | 0.959888 | DISCARD |
| 1-year summary (Drug prescriptions) - NON-STEROIDAL ANTI-INFLAMMATORY: NON-ASA BASE Days since last prescription | | 0.959888 | DISCARD |
| 1-year summary (Drug prescriptions) - NON-STEROIDAL ANTI-INFLAMMATORY: NON-ASA BASE Prescription count | | 0.959888 | DISCARD |
| 2-year summary (Drug prescriptions) - ANGIOTENSIN II ANTAGONIST Days since first prescription | | 0.960080 | DISCARD |
| 2-year summary (Drug prescriptions) - ANGIOTENSIN II ANTAGONIST Days since last prescription | | 0.960080 | DISCARD |
| 2-year summary (Drug prescriptions) - ANGIOTENSIN II ANTAGONIST Prescription count | | 0.960080 | DISCARD |
| 2-year summary (Laboratory tests) - number of tests for Hemoglobin_A1c/Hemoglobin.total | | 0.960080 | DISCARD |
| 2-year summary (Laboratory tests) - days since earliest test for Hemoglobin_A1c/Hemoglobin.total | | 0.960080 | DISCARD |
| 2-year summary (Laboratory tests) - days since latest test for Hemoglobin_A1c/Hemoglobin.total | | 0.960080 | DISCARD |
| 2-year summary (Laboratory tests) - number of high values of Hemoglobin_A1c/Hemoglobin.total | | 0.960080 | DISCARD |
| 2-year summary (Laboratory tests) - highest value of Hemoglobin_A1c/Hemoglobin.total | | 0.960080 | DISCARD |
| 2-year summary (Laboratory tests) - number of low values of Hemoglobin_A1c/Hemoglobin.total | | 0.960080 | DISCARD |
| 2-year summary (Laboratory tests) - lowest value of Hemoglobin_A1c/Hemoglobin.total | | 0.960080 | DISCARD |
| 2-year summary (Laboratory tests) - average value of Hemoglobin_A1c/Hemoglobin.total | | 0.960080 | DISCARD |
| 2-year summary (Laboratory tests) - median value of Hemoglobin_A1c/Hemoglobin.total | | 0.960080 | DISCARD |
| 2-year summary (Laboratory tests) - count of normal tests for Hemoglobin_A1c/Hemoglobin.total | | 0.960080 | DISCARD |
| 1-year summary (Drug prescriptions) - ACE INHIBITORS Average frequency (days) | | 0.960108 | DISCARD |
| 1-year summary (Drug prescriptions) - ACE INHIBITORS Max frequency (days) | | 0.960108 | DISCARD |
| 1-year summary (Drug prescriptions) - ACE INHIBITORS Min frequency (days) | | 0.960108 | DISCARD |
| 1-year summary (Doctor visits) - PATHOLOGY, MICROBIOLOGY, CLINICAL BIOCHEMISTRY | | 0.960191 | DISCARD |
| 2-year summary (Drug prescriptions) - ANALGESICS AND ANTIPYRETICS: MISCELLANEOUS Average frequency (days) | | 0.960356 | DISCARD |
| 2-year summary (Drug prescriptions) - ANALGESICS AND ANTIPYRETICS: MISCELLANEOUS Max frequency (days) | | 0.960356 | DISCARD |
| 2-year summary (Drug prescriptions) - ANALGESICS AND ANTIPYRETICS: MISCELLANEOUS Min frequency (days) | | 0.960356 | DISCARD |
| 2-year summary (Drug prescriptions) - SELECTIVE SEROTONIN REUPTAKE INHIBITORS Days since first prescription | | 0.960659 | DISCARD |
| 2-year summary (Drug prescriptions) - SELECTIVE SEROTONIN REUPTAKE INHIBITORS Days since last prescription | | 0.960659 | DISCARD |

| | | | |
|---|---|---|---|
| 2-year summary (Drug prescriptions) - SELECTIVE SEROTONIN REUPTAKE INHIBITORS Prescription count | | 0.960659 | DISCARD |
| 2-year summary (Drug prescriptions) - NOT IDENTIFIED Average frequency (days) | | 0.960796 | DISCARD |
| 2-year summary (Drug prescriptions) - NOT IDENTIFIED Max frequency (days) | | 0.960796 | DISCARD |
| 2-year summary (Drug prescriptions) - NOT IDENTIFIED Min frequency (days) | | 0.960796 | DISCARD |
| 1-year summary (Doctor visits) - PSYCHIATRY | | 0.961264 | DISCARD |
| 1-year summary (Drug prescriptions) - SELECTIVE BETA2-ADRENERGIC AGONISTS Days since first prescription | | 0.962035 | DISCARD |
| 1-year summary (Drug prescriptions) - SELECTIVE BETA2-ADRENERGIC AGONISTS Days since last prescription | | 0.962035 | DISCARD |
| 1-year summary (Drug prescriptions) - SELECTIVE BETA2-ADRENERGIC AGONISTS Prescription count | | 0.962035 | DISCARD |
| 2-year summary (Hospitalizations) - Class - Diseases of the circulatory system (I00-I99) | | 0.962668 | DISCARD |
| 2-year summary (Drug prescriptions) - ANGIOTENSIN II ANTAGONIST Average frequency (days) | | 0.962889 | DISCARD |
| 2-year summary (Drug prescriptions) - ANGIOTENSIN II ANTAGONIST Max frequency (days) | | 0.962889 | DISCARD |
| 2-year summary (Drug prescriptions) - ANGIOTENSIN II ANTAGONIST Min frequency (days) | | 0.962889 | DISCARD |
| 1-year summary (Doctor visits) - NEUROLOGY | | 0.963026 | DISCARD |
| 1-year summary (Drug prescriptions) - DIABETES DIAGNOSTIC Days since first prescription | | 0.963054 | DISCARD |
| 1-year summary (Drug prescriptions) - DIABETES DIAGNOSTIC Days since last prescription | | 0.963054 | DISCARD |
| 1-year summary (Drug prescriptions) - DIABETES DIAGNOSTIC Prescription count | | 0.963054 | DISCARD |
| 1-year summary (Drug prescriptions) - CORTICOSTEROIDS, PLAIN Average frequency (days) | | 0.963219 | DISCARD |
| 1-year summary (Drug prescriptions) - CORTICOSTEROIDS, PLAIN Max frequency (days) | | 0.963219 | DISCARD |
| 1-year summary (Drug prescriptions) - CORTICOSTEROIDS, PLAIN Min frequency (days) | | 0.963219 | DISCARD |
| 1-year summary (Drug prescriptions) - DIURETICS Average frequency (days) | | 0.963219 | DISCARD |
| 1-year summary (Drug prescriptions) - DIURETICS Max frequency (days) | | 0.963219 | DISCARD |
| 1-year summary (Drug prescriptions) - DIURETICS Min frequency (days) | | 0.963219 | DISCARD |
| 2-year summary (Drug prescriptions) - PENICILLINS Average frequency (days) | | 0.963301 | DISCARD |
| 2-year summary (Drug prescriptions) - PENICILLINS Max frequency (days) | | 0.963301 | DISCARD |
| 2-year summary (Drug prescriptions) - PENICILLINS Min frequency (days) | | 0.963301 | DISCARD |
| 2-year summary (Drug prescriptions) - ANTIBIOTICS Days since first prescription | | 0.963329 | DISCARD |
| 2-year summary (Drug prescriptions) - ANTIBIOTICS Days since last prescription | | 0.963329 | DISCARD |
| 2-year summary (Drug prescriptions) - ANTIBIOTICS Prescription count | | 0.963329 | DISCARD |
| 1-year summary (Doctor visits) - GASTROENTEROLOGY | | 0.963357 | DISCARD |
| 2-year summary (Drug prescriptions) - NON-STEROIDAL ANTI-INFLAMMATORY: NON-ASA BASE Average frequency (days) | | 0.964265 | DISCARD |
| 2-year summary (Drug prescriptions) - NON-STEROIDAL ANTI-INFLAMMATORY: NON-ASA BASE Max frequency (days) | | 0.964265 | DISCARD |
| 2-year summary (Drug prescriptions) - NON-STEROIDAL ANTI-INFLAMMATORY: NON-ASA BASE Min frequency (days) | | 0.964265 | DISCARD |
| 1-year summary (Drug prescriptions) - CATHARTICS AND LAXATIVES Days since first prescription | | 0.964568 | DISCARD |
| 1-year summary (Drug prescriptions) - CATHARTICS AND LAXATIVES Days since last prescription | | 0.964568 | DISCARD |
| 1-year summary (Drug prescriptions) - CATHARTICS AND LAXATIVES Prescription count | | 0.964568 | DISCARD |
| 1-year summary (Drug prescriptions) - ANGIOTENSIN II ANTAGONIST Days since first prescription | | 0.964871 | DISCARD |
| 1-year summary (Drug prescriptions) - ANGIOTENSIN II ANTAGONIST Days since last prescription | | 0.964871 | DISCARD |

| | | | |
|---|---|---|---|
| 1-year summary (Drug prescriptions) - ANGIOTENSIN II ANTAGONIST Prescription count | | 0.964871 | DISCARD |
| 2-year summary (Hospitalizations) - Class - Endocrine, nutritional and metabolic diseases (E00-E90) | | 0.964926 | DISCARD |
| 2-year summary (Drug prescriptions) - DIABETES DIAGNOSTIC Average frequency (days) | | 0.965449 | DISCARD |
| 2-year summary (Drug prescriptions) - DIABETES DIAGNOSTIC Max frequency (days) | | 0.965449 | DISCARD |
| 2-year summary (Drug prescriptions) - DIABETES DIAGNOSTIC Min frequency (days) | | 0.965449 | DISCARD |
| 1-year summary (Doctor visits) - RESPIRATORY DISEASE | | 0.965476 | DISCARD |
| 2-year summary (Drug prescriptions) - URINARY ANTI-INFECTIVES Days since first prescription | | 0.965587 | DISCARD |
| 2-year summary (Drug prescriptions) - URINARY ANTI-INFECTIVES Days since last prescription | | 0.965587 | DISCARD |
| 2-year summary (Drug prescriptions) - URINARY ANTI-INFECTIVES Prescription count | | 0.965587 | DISCARD |
| 2-year summary (Drug prescriptions) - ANTITUSSIVE Days since first prescription | | 0.966110 | DISCARD |
| 2-year summary (Drug prescriptions) - ANTITUSSIVE Days since last prescription | | 0.966110 | DISCARD |
| 2-year summary (Drug prescriptions) - ANTITUSSIVE Prescription count | | 0.966110 | DISCARD |
| 2-year summary (Drug prescriptions) - BENZODIAZEPINE DERIVATIVES Days since first prescription | | 0.966302 | DISCARD |
| 2-year summary (Drug prescriptions) - BENZODIAZEPINE DERIVATIVES Days since last prescription | | 0.966302 | DISCARD |
| 2-year summary (Drug prescriptions) - BENZODIAZEPINE DERIVATIVES Prescription count | | 0.966302 | DISCARD |
| 2-year summary (Drug prescriptions) - NARCOTICS: OPIATE AGONISTS Average frequency (days) | | 0.966440 | DISCARD |
| 2-year summary (Drug prescriptions) - NARCOTICS: OPIATE AGONISTS Max frequency (days) | | 0.966440 | DISCARD |
| 2-year summary (Drug prescriptions) - NARCOTICS: OPIATE AGONISTS Min frequency (days) | | 0.966440 | DISCARD |
| 2-year summary (Drug prescriptions) - SELECTIVE SEROTONIN REUPTAKE INHIBITORS Average frequency (days) | | 0.966468 | DISCARD |
| 2-year summary (Drug prescriptions) - SELECTIVE SEROTONIN REUPTAKE INHIBITORS Max frequency (days) | | 0.966468 | DISCARD |
| 2-year summary (Drug prescriptions) - SELECTIVE SEROTONIN REUPTAKE INHIBITORS Min frequency (days) | | 0.966468 | DISCARD |
| 2-year summary (Doctor visits) - RHEUMATOLOGY | | 0.966991 | DISCARD |
| 1-year summary (Drug prescriptions) - ANGIOTENSIN II ANTAGONIST Average frequency (days) | | 0.967872 | DISCARD |
| 1-year summary (Drug prescriptions) - ANGIOTENSIN II ANTAGONIST Max frequency (days) | | 0.967872 | DISCARD |
| 1-year summary (Drug prescriptions) - ANGIOTENSIN II ANTAGONIST Min frequency (days) | | 0.967872 | DISCARD |
| 2-year summary (Drug prescriptions) - HYPOTHYROIDISM THERAPY Days since first prescription | | 0.968257 | DISCARD |
| 2-year summary (Drug prescriptions) - HYPOTHYROIDISM THERAPY Days since last prescription | | 0.968257 | DISCARD |
| 2-year summary (Drug prescriptions) - HYPOTHYROIDISM THERAPY Prescription count | | 0.968257 | DISCARD |
| 2-year summary (Hospitalizations) - Class - Symptoms, signs and abnormal clinical and laboratory findings, not elsewhere classified (I | | 0.968450 | DISCARD |
| 2-year summary (Drug prescriptions) - VACCINES Average frequency (days) | | 0.969303 | DISCARD |
| 2-year summary (Drug prescriptions) - VACCINES Max frequency (days) | | 0.969303 | DISCARD |
| 2-year summary (Drug prescriptions) - VACCINES Min frequency (days) | | 0.969303 | DISCARD |
| 2-year summary (Drug prescriptions) - SELECTIVE BETA2-ADRENERGIC AGONISTS Average frequency (days) | | 0.969331 | DISCARD |
| 2-year summary (Drug prescriptions) - SELECTIVE BETA2-ADRENERGIC AGONISTS Max frequency (days) | | 0.969331 | DISCARD |
| 2-year summary (Drug prescriptions) - SELECTIVE BETA2-ADRENERGIC AGONISTS Min frequency (days) | | 0.969331 | DISCARD |
| 2-year summary (Drug prescriptions) - CATHARTICS AND LAXATIVES Average frequency (days) | | 0.969413 | DISCARD |
| 2-year summary (Drug prescriptions) - CATHARTICS AND LAXATIVES Max frequency (days) | | 0.969413 | DISCARD |

| | | |
|---|---|---|
| 2-year summary (Drug prescriptions) - CATHARTICS AND LAXATIVES Min frequency (days) | 0.969413 | DISCARD |
| 2-year summary (Drug prescriptions) - HYPOTHYROIDISM THERAPY Average frequency (days) | 0.969468 | DISCARD |
| 2-year summary (Drug prescriptions) - HYPOTHYROIDISM THERAPY Max frequency (days) | 0.969468 | DISCARD |
| 2-year summary (Drug prescriptions) - HYPOTHYROIDISM THERAPY Min frequency (days) | 0.969468 | DISCARD |
| 1-year summary (Drug prescriptions) - INHALED CORTICOSTEROIDS Days since first prescription | 0.970047 | DISCARD |
| 1-year summary (Drug prescriptions) - INHALED CORTICOSTEROIDS Days since last prescription | 0.970047 | DISCARD |
| 1-year summary (Drug prescriptions) - INHALED CORTICOSTEROIDS Prescription count | 0.970047 | DISCARD |
| 1-year summary (Drug prescriptions) - HYPOTHYROIDISM THERAPY Days since first prescription | 0.970459 | DISCARD |
| 1-year summary (Drug prescriptions) - HYPOTHYROIDISM THERAPY Days since last prescription | 0.970459 | DISCARD |
| 1-year summary (Drug prescriptions) - HYPOTHYROIDISM THERAPY Prescription count | 0.970459 | DISCARD |
| 1-year summary (Drug prescriptions) - SELECTIVE SEROTONIN REUPTAKE INHIBITORS Days since first prescription | 0.970570 | DISCARD |
| 1-year summary (Drug prescriptions) - SELECTIVE SEROTONIN REUPTAKE INHIBITORS Days since last prescription | 0.970570 | DISCARD |
| 1-year summary (Drug prescriptions) - SELECTIVE SEROTONIN REUPTAKE INHIBITORS Prescription count | 0.970570 | DISCARD |
| 1-year summary (Drug prescriptions) - ANALGESICS AND ANTIPYRETICS: MISCELLANEOUS Average frequency (days) | 0.971065 | DISCARD |
| 1-year summary (Drug prescriptions) - ANALGESICS AND ANTIPYRETICS: MISCELLANEOUS Max frequency (days) | 0.971065 | DISCARD |
| 1-year summary (Drug prescriptions) - ANALGESICS AND ANTIPYRETICS: MISCELLANEOUS Min frequency (days) | 0.971065 | DISCARD |
| 2-year summary (Drug prescriptions) - ANTIDEPRESSANTS: MISCELLANEOUS Days since first prescription | 0.971175 | DISCARD |
| 2-year summary (Drug prescriptions) - ANTIDEPRESSANTS: MISCELLANEOUS Days since last prescription | 0.971175 | DISCARD |
| 2-year summary (Drug prescriptions) - ANTIDEPRESSANTS: MISCELLANEOUS Prescription count | 0.971175 | DISCARD |
| 1-year summary (Drug prescriptions) - HYPOTHYROIDISM THERAPY Average frequency (days) | 0.971588 | DISCARD |
| 1-year summary (Drug prescriptions) - HYPOTHYROIDISM THERAPY Max frequency (days) | 0.971588 | DISCARD |
| 1-year summary (Drug prescriptions) - HYPOTHYROIDISM THERAPY Min frequency (days) | 0.971588 | DISCARD |
| 1-year summary (Hospitalizations) - Class - Factors influencing health status and contact with health services (Z00-Z99) | 0.971864 | DISCARD |
| 2-year summary (Hospitalizations) - Class - Diseases of the genitourinary system (N00-N99) | 0.972662 | DISCARD |
| 1-year summary (Drug prescriptions) - SELECTIVE SEROTONIN REUPTAKE INHIBITORS Average frequency (days) | 0.974479 | DISCARD |
| 1-year summary (Drug prescriptions) - SELECTIVE SEROTONIN REUPTAKE INHIBITORS Max frequency (days) | 0.974479 | DISCARD |
| 1-year summary (Drug prescriptions) - SELECTIVE SEROTONIN REUPTAKE INHIBITORS Min frequency (days) | 0.974479 | DISCARD |
| 1-year summary (Drug prescriptions) - CEPHALOSPORIN Days since first prescription | 0.974507 | DISCARD |
| 1-year summary (Drug prescriptions) - CEPHALOSPORIN Days since last prescription | 0.974507 | DISCARD |
| 1-year summary (Drug prescriptions) - CEPHALOSPORIN Prescription count | 0.974507 | DISCARD |
| 1-year summary (Drug prescriptions) - MACROLIDES Days since first prescription | 0.974562 | DISCARD |
| 1-year summary (Drug prescriptions) - MACROLIDES Days since last prescription | 0.974562 | DISCARD |
| 1-year summary (Drug prescriptions) - MACROLIDES Prescription count | 0.974562 | DISCARD |
| 2-year summary (Hospitalizations) - Class - External causes of morbidity and mortality (V01-Y98) | 0.974727 | DISCARD |
| 1-year summary (Drug prescriptions) - BENZODIAZEPINE DERIVATIVES Days since first prescription | 0.974919 | DISCARD |
| 1-year summary (Drug prescriptions) - BENZODIAZEPINE DERIVATIVES Days since last prescription | 0.974919 | DISCARD |
| 1-year summary (Drug prescriptions) - BENZODIAZEPINE DERIVATIVES Prescription count | 0.974919 | DISCARD |

| | | | |
|---|---|---|---|
| ground_truth_death_T | | 0.975057 | DISCARD |
| 2-year summary (Drug prescriptions) - ANTIPSYCHOTIC AGENTS Days since first prescription | | 0.975085 | DISCARD |
| 2-year summary (Drug prescriptions) - ANTIPSYCHOTIC AGENTS Days since last prescription | | 0.975085 | DISCARD |
| 2-year summary (Drug prescriptions) - ANTIPSYCHOTIC AGENTS Prescription count | | 0.975085 | DISCARD |
| 2-year summary (Drug prescriptions) - IRON PREPARATIONS Days since first prescription | | 0.975112 | DISCARD |
| 2-year summary (Drug prescriptions) - IRON PREPARATIONS Days since last prescription | | 0.975112 | DISCARD |
| 2-year summary (Drug prescriptions) - IRON PREPARATIONS Prescription count | | 0.975112 | DISCARD |
| 2-year summary (Drug prescriptions) - ANTICONVULSANTS: MISCELLANEOUS Days since first prescription | | 0.975525 | DISCARD |
| 2-year summary (Drug prescriptions) - ANTICONVULSANTS: MISCELLANEOUS Days since last prescription | | 0.975525 | DISCARD |
| 2-year summary (Drug prescriptions) - ANTICONVULSANTS: MISCELLANEOUS Prescription count | | 0.975525 | DISCARD |
| 1-year summary (Drug prescriptions) - FUNGICIDES Days since first prescription | | 0.975828 | DISCARD |
| 1-year summary (Drug prescriptions) - FUNGICIDES Days since last prescription | | 0.975828 | DISCARD |
| 1-year summary (Drug prescriptions) - FUNGICIDES Prescription count | | 0.975828 | DISCARD |
| 2-year summary (Drug prescriptions) - BENZODIAZEPINE DERIVATIVES Average frequency (days) | | 0.975993 | DISCARD |
| 2-year summary (Drug prescriptions) - BENZODIAZEPINE DERIVATIVES Max frequency (days) | | 0.975993 | DISCARD |
| 2-year summary (Drug prescriptions) - BENZODIAZEPINE DERIVATIVES Min frequency (days) | | 0.975993 | DISCARD |
| 2-year summary (Hospitalizations) - Class - Diseases of the digestive system (K00-K93) | | 0.976021 | DISCARD |
| 1-year summary (Drug prescriptions) - FLUOROQUINOLONES Days since first prescription | | 0.976213 | DISCARD |
| 1-year summary (Drug prescriptions) - FLUOROQUINOLONES Days since last prescription | | 0.976213 | DISCARD |
| 1-year summary (Drug prescriptions) - FLUOROQUINOLONES Prescription count | | 0.976213 | DISCARD |
| 1-year summary (Hospitalizations) - Class - Diseases of the circulatory system (I00-I99) | | 0.976461 | DISCARD |
| 1-year summary (Drug prescriptions) - CATHARTICS AND LAXATIVES Average frequency (days) | | 0.977232 | DISCARD |
| 1-year summary (Drug prescriptions) - CATHARTICS AND LAXATIVES Max frequency (days) | | 0.977232 | DISCARD |
| 1-year summary (Drug prescriptions) - CATHARTICS AND LAXATIVES Min frequency (days) | | 0.977232 | DISCARD |
| 2-year summary (Laboratory tests) - number of tests for Cholesterol.non_HDL | | 0.977535 | DISCARD |
| 2-year summary (Laboratory tests) - days since earliest test for Cholesterol.non_HDL | | 0.977535 | DISCARD |
| 2-year summary (Laboratory tests) - days since latest test for Cholesterol.non_HDL | | 0.977535 | DISCARD |
| 2-year summary (Laboratory tests) - number of high values of Cholesterol.non_HDL | | 0.977535 | DISCARD |
| 2-year summary (Laboratory tests) - highest value of Cholesterol.non_HDL | | 0.977535 | DISCARD |
| 2-year summary (Laboratory tests) - number of low values of Cholesterol.non_HDL | | 0.977535 | DISCARD |
| 2-year summary (Laboratory tests) - lowest value of Cholesterol.non_HDL | | 0.977535 | DISCARD |
| 2-year summary (Laboratory tests) - average value of Cholesterol.non_HDL | | 0.977535 | DISCARD |
| 2-year summary (Laboratory tests) - median value of Cholesterol.non_HDL | | 0.977535 | DISCARD |
| 2-year summary (Laboratory tests) - count of normal tests for Cholesterol.non_HDL | | 0.977535 | DISCARD |
| 1-year summary (Doctor visits) - RHEUMATOLOGY | | 0.977617 | DISCARD |
| 1-year summary (Drug prescriptions) - ANTIDEPRESSANTS: MISCELLANEOUS Days since first prescription | | 0.977645 | DISCARD |
| 1-year summary (Drug prescriptions) - ANTIDEPRESSANTS: MISCELLANEOUS Days since last prescription | | 0.977645 | DISCARD |

| | | | |
|---|---|---|---|
| 1-year summary (Drug prescriptions) - ANTIDEPRESSANTS: MISCELLANEOUS Prescription count | | 0.977645 | DISCARD |
| 1-year summary (Doctor visits) - PLASTIC SURGERY | | 0.977865 | DISCARD |
| 1-year summary (Hospitalizations) - Class - Endocrine, nutritional and metabolic diseases (E00-E90) | | 0.977893 | DISCARD |
| 2-year summary (Drug prescriptions) - INHALED CORTICOSTEROIDS Average frequency (days) | | 0.977893 | DISCARD |
| 2-year summary (Drug prescriptions) - INHALED CORTICOSTEROIDS Max frequency (days) | | 0.977893 | DISCARD |
| 2-year summary (Drug prescriptions) - INHALED CORTICOSTEROIDS Min frequency (days) | | 0.977893 | DISCARD |
| 2-year summary (Drug prescriptions) - ANTIDEPRESSANTS: MISCELLANEOUS Average frequency (days) | | 0.977920 | DISCARD |
| 2-year summary (Drug prescriptions) - ANTIDEPRESSANTS: MISCELLANEOUS Max frequency (days) | | 0.977920 | DISCARD |
| 2-year summary (Drug prescriptions) - ANTIDEPRESSANTS: MISCELLANEOUS Min frequency (days) | | 0.977920 | DISCARD |
| 2-year summary (Doctor visits) - VASCULAR SURGERY | | 0.978003 | DISCARD |
| 2-year summary (Drug prescriptions) - PROSTATIC HYPERPLASIA Days since first prescription | | 0.978251 | DISCARD |
| 2-year summary (Drug prescriptions) - PROSTATIC HYPERPLASIA Days since last prescription | | 0.978251 | DISCARD |
| 2-year summary (Drug prescriptions) - PROSTATIC HYPERPLASIA Prescription count | | 0.978251 | DISCARD |
| 1-year summary (Drug prescriptions) - NARCOTICS: OPIATE AGONISTS Average frequency (days) | | 0.978306 | DISCARD |
| 1-year summary (Drug prescriptions) - NARCOTICS: OPIATE AGONISTS Max frequency (days) | | 0.978306 | DISCARD |
| 1-year summary (Drug prescriptions) - NARCOTICS: OPIATE AGONISTS Min frequency (days) | | 0.978306 | DISCARD |
| 2-year summary (Doctor visits) - PHYSICAL MEDICINE | | 0.978306 | DISCARD |
| 2-year summary (Doctor visits) - NEPHROLOGY | | 0.978361 | DISCARD |
| 2-year summary (Drug prescriptions) - ANTIPSYCHOTIC AGENTS Average frequency (days) | | 0.978388 | DISCARD |
| 2-year summary (Drug prescriptions) - ANTIPSYCHOTIC AGENTS Max frequency (days) | | 0.978388 | DISCARD |
| 2-year summary (Drug prescriptions) - ANTIPSYCHOTIC AGENTS Min frequency (days) | | 0.978388 | DISCARD |
| 1-year summary (Drug prescriptions) - ANTIPSYCHOTIC AGENTS Days since first prescription | | 0.979049 | DISCARD |
| 1-year summary (Drug prescriptions) - ANTIPSYCHOTIC AGENTS Days since last prescription | | 0.979049 | DISCARD |
| 1-year summary (Drug prescriptions) - ANTIPSYCHOTIC AGENTS Prescription count | | 0.979049 | DISCARD |
| 1-year summary (Drug prescriptions) - DIABETES DIAGNOSTIC Average frequency (days) | | 0.979352 | DISCARD |
| 1-year summary (Drug prescriptions) - DIABETES DIAGNOSTIC Max frequency (days) | | 0.979352 | DISCARD |
| 1-year summary (Drug prescriptions) - DIABETES DIAGNOSTIC Min frequency (days) | | 0.979352 | DISCARD |
| 1-year summary (Laboratory tests) - number of tests for Cholesterol.in_LDL | | 0.979490 | DISCARD |
| 1-year summary (Laboratory tests) - days since earliest test for Cholesterol.in_LDL | | 0.979490 | DISCARD |
| 1-year summary (Laboratory tests) - days since latest test for Cholesterol.in_LDL | | 0.979490 | DISCARD |
| 1-year summary (Laboratory tests) - number of high values of Cholesterol.in_LDL | | 0.979490 | DISCARD |
| 1-year summary (Laboratory tests) - highest value of Cholesterol.in_LDL | | 0.979490 | DISCARD |
| 1-year summary (Laboratory tests) - number of low values of Cholesterol.in_LDL | | 0.979490 | DISCARD |
| 1-year summary (Laboratory tests) - lowest value of Cholesterol.in_LDL | | 0.979490 | DISCARD |
| 1-year summary (Laboratory tests) - average value of Cholesterol.in_LDL | | 0.979490 | DISCARD |
| 1-year summary (Laboratory tests) - median value of Cholesterol.in_LDL | | 0.979490 | DISCARD |
| 1-year summary (Laboratory tests) - count of normal tests for Cholesterol.in_LDL | | 0.979490 | DISCARD |

| | | | |
|---|---|---|---|
| 1-year summary (Laboratory tests) - number of tests for Hemoglobin_A1c/Hemoglobin.total | | 0.979710 | DISCARD |
| 1-year summary (Laboratory tests) - days since earliest test for Hemoglobin_A1c/Hemoglobin.total | | 0.979710 | DISCARD |
| 1-year summary (Laboratory tests) - days since latest test for Hemoglobin_A1c/Hemoglobin.total | | 0.979710 | DISCARD |
| 1-year summary (Laboratory tests) - number of high values of Hemoglobin_A1c/Hemoglobin.total | | 0.979710 | DISCARD |
| 1-year summary (Laboratory tests) - highest value of Hemoglobin_A1c/Hemoglobin.total | | 0.979710 | DISCARD |
| 1-year summary (Laboratory tests) - number of low values of Hemoglobin_A1c/Hemoglobin.total | | 0.979710 | DISCARD |
| 1-year summary (Laboratory tests) - lowest value of Hemoglobin_A1c/Hemoglobin.total | | 0.979710 | DISCARD |
| 1-year summary (Laboratory tests) - average value of Hemoglobin_A1c/Hemoglobin.total | | 0.979710 | DISCARD |
| 1-year summary (Laboratory tests) - median value of Hemoglobin_A1c/Hemoglobin.total | | 0.979710 | DISCARD |
| 1-year summary (Laboratory tests) - count of normal tests for Hemoglobin_A1c/Hemoglobin.total | | 0.979710 | DISCARD |
| 2-year summary (Drug prescriptions) - STOOL SOFTENERS Days since first prescription | | 0.979792 | DISCARD |
| 2-year summary (Drug prescriptions) - STOOL SOFTENERS Days since last prescription | | 0.979792 | DISCARD |
| 2-year summary (Drug prescriptions) - STOOL SOFTENERS Prescription count | | 0.979792 | DISCARD |
| 1-year summary (Drug prescriptions) - ANTIBIOTICS Days since first prescription | | 0.980178 | DISCARD |
| 1-year summary (Drug prescriptions) - ANTIBIOTICS Days since last prescription | | 0.980178 | DISCARD |
| 1-year summary (Drug prescriptions) - ANTIBIOTICS Prescription count | | 0.980178 | DISCARD |
| 2-year summary (Doctor visits) - INFECTIOUS DISEASE | | 0.980233 | DISCARD |
| 2-year summary (Hospitalizations) - Class - Pregnancy, childbirth and the puerperium (O00-O99) | | 0.980260 | DISCARD |
| 2-year summary (Drug prescriptions) - IRON PREPARATIONS Average frequency (days) | | 0.980371 | DISCARD |
| 2-year summary (Drug prescriptions) - IRON PREPARATIONS Max frequency (days) | | 0.980371 | DISCARD |
| 2-year summary (Drug prescriptions) - IRON PREPARATIONS Min frequency (days) | | 0.980371 | DISCARD |
| 1-year summary (Drug prescriptions) - PROSTATIC HYPERPLASIA Days since first prescription | | 0.980398 | DISCARD |
| 1-year summary (Drug prescriptions) - PROSTATIC HYPERPLASIA Days since last prescription | | 0.980398 | DISCARD |
| 1-year summary (Drug prescriptions) - PROSTATIC HYPERPLASIA Prescription count | | 0.980398 | DISCARD |
| 2-year summary (Doctor visits) - PAEDIATRICS | | 0.980426 | DISCARD |
| 2-year summary (Drug prescriptions) - PROSTATIC HYPERPLASIA Average frequency (days) | | 0.980563 | DISCARD |
| 2-year summary (Drug prescriptions) - PROSTATIC HYPERPLASIA Max frequency (days) | | 0.980563 | DISCARD |
| 2-year summary (Drug prescriptions) - PROSTATIC HYPERPLASIA Min frequency (days) | | 0.980563 | DISCARD |
| 2-year summary (Drug prescriptions) - ESTROGENS-PROGESTERONE FIXED COMBINATIONS Days since first prescription | | 0.980591 | DISCARD |
| 2-year summary (Drug prescriptions) - ESTROGENS-PROGESTERONE FIXED COMBINATIONS Days since last prescription | | 0.980591 | DISCARD |
| 2-year summary (Drug prescriptions) - ESTROGENS-PROGESTERONE FIXED COMBINATIONS Prescription count | | 0.980591 | DISCARD |
| 2-year summary (Drug prescriptions) - INSULINS Days since first prescription | | 0.980591 | DISCARD |
| 2-year summary (Drug prescriptions) - INSULINS Days since last prescription | | 0.980591 | DISCARD |
| 2-year summary (Drug prescriptions) - INSULINS Prescription count | | 0.980591 | DISCARD |
| 1-year summary (Drug prescriptions) - SELECTIVE BETA2-ADRENERGIC AGONISTS Average frequency (days) | | 0.980646 | DISCARD |
| 1-year summary (Drug prescriptions) - SELECTIVE BETA2-ADRENERGIC AGONISTS Max frequency (days) | | 0.980646 | DISCARD |
| 1-year summary (Drug prescriptions) - SELECTIVE BETA2-ADRENERGIC AGONISTS Min frequency (days) | | 0.980646 | DISCARD |

| | | |
|---|---|---|
| 2-year summary (Doctor visits) - HAEMATOLOGY | 0.980646 | DISCARD |
| 2-year summary (Drug prescriptions) - BRONCHODILATOR AND ANTI-INFLAMMATORY COMBINATION Days since first prescriptio | 0.980756 | DISCARD |
| 2-year summary (Drug prescriptions) - BRONCHODILATOR AND ANTI-INFLAMMATORY COMBINATION Days since last prescriptio | 0.980756 | DISCARD |
| 2-year summary (Drug prescriptions) - BRONCHODILATOR AND ANTI-INFLAMMATORY COMBINATION Prescription count | 0.980756 | DISCARD |
| 1-year summary (Hospitalizations) - Class - Symptoms, signs and abnormal clinical and laboratory findings, not elsewhere classified (\| | 0.980784 | DISCARD |
| 1-year summary (Drug prescriptions) - ANTITUSSIVE Days since first prescription | 0.980811 | DISCARD |
| 1-year summary (Drug prescriptions) - ANTITUSSIVE Days since last prescription | 0.980811 | DISCARD |
| 1-year summary (Drug prescriptions) - ANTITUSSIVE Prescription count | 0.980811 | DISCARD |
| 1-year summary (Drug prescriptions) - ANTICONVULSANTS: MISCELLANEOUS Days since first prescription | 0.981086 | DISCARD |
| 1-year summary (Drug prescriptions) - ANTICONVULSANTS: MISCELLANEOUS Days since last prescription | 0.981086 | DISCARD |
| 1-year summary (Drug prescriptions) - ANTICONVULSANTS: MISCELLANEOUS Prescription count | 0.981086 | DISCARD |
| 2-year summary (Drug prescriptions) - ANTICONVULSANTS: MISCELLANEOUS Average frequency (days) | 0.981169 | DISCARD |
| 2-year summary (Drug prescriptions) - ANTICONVULSANTS: MISCELLANEOUS Max frequency (days) | 0.981169 | DISCARD |
| 2-year summary (Drug prescriptions) - ANTICONVULSANTS: MISCELLANEOUS Min frequency (days) | 0.981169 | DISCARD |
| 1-year summary (Drug prescriptions) - ANTIPSYCHOTIC AGENTS Average frequency (days) | 0.981196 | DISCARD |
| 1-year summary (Drug prescriptions) - ANTIPSYCHOTIC AGENTS Max frequency (days) | 0.981196 | DISCARD |
| 1-year summary (Drug prescriptions) - ANTIPSYCHOTIC AGENTS Min frequency (days) | 0.981196 | DISCARD |
| 1-year summary (Laboratory tests) - number of tests for Cholesterol.non_HDL | 0.981279 | DISCARD |
| 1-year summary (Laboratory tests) - days since earliest test for Cholesterol.non_HDL | 0.981279 | DISCARD |
| 1-year summary (Laboratory tests) - days since latest test for Cholesterol.non_HDL | 0.981279 | DISCARD |
| 1-year summary (Laboratory tests) - number of high values of Cholesterol.non_HDL | 0.981279 | DISCARD |
| 1-year summary (Laboratory tests) - highest value of Cholesterol.non_HDL | 0.981279 | DISCARD |
| 1-year summary (Laboratory tests) - number of low values of Cholesterol.non_HDL | 0.981279 | DISCARD |
| 1-year summary (Laboratory tests) - lowest value of Cholesterol.non_HDL | 0.981279 | DISCARD |
| 1-year summary (Laboratory tests) - average value of Cholesterol.non_HDL | 0.981279 | DISCARD |
| 1-year summary (Laboratory tests) - median value of Cholesterol.non_HDL | 0.981279 | DISCARD |
| 1-year summary (Laboratory tests) - count of normal tests for Cholesterol.non_HDL | 0.981279 | DISCARD |
| 1-year summary (Drug prescriptions) - URINARY ANTI-INFECTIVES Days since first prescription | 0.981334 | DISCARD |
| 1-year summary (Drug prescriptions) - URINARY ANTI-INFECTIVES Days since last prescription | 0.981334 | DISCARD |
| 1-year summary (Drug prescriptions) - URINARY ANTI-INFECTIVES Prescription count | 0.981334 | DISCARD |
| 1-year summary (Drug prescriptions) - BENZODIAZEPINE DERIVATIVES Average frequency (days) | 0.981692 | DISCARD |
| 1-year summary (Drug prescriptions) - BENZODIAZEPINE DERIVATIVES Max frequency (days) | 0.981692 | DISCARD |
| 1-year summary (Drug prescriptions) - BENZODIAZEPINE DERIVATIVES Min frequency (days) | 0.981692 | DISCARD |
| 1-year summary (Drug prescriptions) - NON-STEROIDAL ANTI-INFLAMMATORY: NON-ASA BASE Average frequency (days) | 0.981692 | DISCARD |
| 1-year summary (Drug prescriptions) - NON-STEROIDAL ANTI-INFLAMMATORY: NON-ASA BASE Max frequency (days) | 0.981692 | DISCARD |
| 1-year summary (Drug prescriptions) - NON-STEROIDAL ANTI-INFLAMMATORY: NON-ASA BASE Min frequency (days) | 0.981692 | DISCARD |
| 2-year summary (Hospitalizations) - Class - Diseases of the respiratory system (J00-J99) | 0.981885 | DISCARD |

| | | | |
|---|---|---|---|
| 2-year summary (Hospitalizations) - Class - Mental and behavioural disorders (F00-F99) | | 0.981885 | DISCARD |
| 1-year summary (Drug prescriptions) - ANTIDEPRESSANTS: MISCELLANEOUS Average frequency (days) | | 0.981967 | DISCARD |
| 1-year summary (Drug prescriptions) - ANTIDEPRESSANTS: MISCELLANEOUS Max frequency (days) | | 0.981967 | DISCARD |
| 1-year summary (Drug prescriptions) - ANTIDEPRESSANTS: MISCELLANEOUS Min frequency (days) | | 0.981967 | DISCARD |
| 1-year summary (Drug prescriptions) - IRON PREPARATIONS Days since first prescription | | 0.981967 | DISCARD |
| 1-year summary (Drug prescriptions) - IRON PREPARATIONS Days since last prescription | | 0.981967 | DISCARD |
| 1-year summary (Drug prescriptions) - IRON PREPARATIONS Prescription count | | 0.981967 | DISCARD |
| 2-year summary (Hospitalizations) - Class - Injury, poisoning and certain other consequences of external causes (S00-T98) | | 0.981995 | DISCARD |
| 1-year summary (Drug prescriptions) - PROSTATIC HYPERPLASIA Average frequency (days) | | 0.982133 | DISCARD |
| 1-year summary (Drug prescriptions) - PROSTATIC HYPERPLASIA Max frequency (days) | | 0.982133 | DISCARD |
| 1-year summary (Drug prescriptions) - PROSTATIC HYPERPLASIA Min frequency (days) | | 0.982133 | DISCARD |
| 2-year summary (Hospitalizations) - Class - Diseases of the musculoskeletal system and connective tissue (M00-M99) | | 0.982133 | DISCARD |
| 2-year summary (Doctor visits) - NUCLEAR MEDICINE | | 0.982243 | DISCARD |
| 2-year summary (Drug prescriptions) - INSULINS Average frequency (days) | | 0.982298 | DISCARD |
| 2-year summary (Drug prescriptions) - INSULINS Max frequency (days) | | 0.982298 | DISCARD |
| 2-year summary (Drug prescriptions) - INSULINS Min frequency (days) | | 0.982298 | DISCARD |
| 1-year summary (Drug prescriptions) - INSULINS Days since first prescription | | 0.982463 | DISCARD |
| 1-year summary (Drug prescriptions) - INSULINS Days since last prescription | | 0.982463 | DISCARD |
| 1-year summary (Drug prescriptions) - INSULINS Prescription count | | 0.982463 | DISCARD |
| 2-year summary (Drug prescriptions) - ANTIBACTERIAL/ANTIPROTOZOAL Days since first prescription | | 0.982490 | DISCARD |
| 2-year summary (Drug prescriptions) - ANTIBACTERIAL/ANTIPROTOZOAL Days since last prescription | | 0.982490 | DISCARD |
| 2-year summary (Drug prescriptions) - ANTIBACTERIAL/ANTIPROTOZOAL Prescription count | | 0.982490 | DISCARD |
| 2-year summary (Drug prescriptions) - BISPHOSPHONATES Days since first prescription | | 0.982683 | DISCARD |
| 2-year summary (Drug prescriptions) - BISPHOSPHONATES Days since last prescription | | 0.982683 | DISCARD |
| 2-year summary (Drug prescriptions) - BISPHOSPHONATES Prescription count | | 0.982683 | DISCARD |
| 1-year summary (Doctor visits) - NEPHROLOGY | | 0.982793 | DISCARD |
| 2-year summary (Drug prescriptions) - FUNGICIDES Average frequency (days) | | 0.982931 | DISCARD |
| 2-year summary (Drug prescriptions) - FUNGICIDES Max frequency (days) | | 0.982931 | DISCARD |
| 2-year summary (Drug prescriptions) - FUNGICIDES Min frequency (days) | | 0.982931 | DISCARD |
| 1-year summary (Hospitalizations) - Class - Diseases of the genitourinary system (N00-N99) | | 0.983069 | DISCARD |
| 2-year summary (Drug prescriptions) - CORTICOSTEROIDS AND ANTI-INFECTIVE COMBINATIONS Days since first prescription | | 0.983371 | DISCARD |
| 2-year summary (Drug prescriptions) - CORTICOSTEROIDS AND ANTI-INFECTIVE COMBINATIONS Days since last prescription | | 0.983371 | DISCARD |
| 2-year summary (Drug prescriptions) - CORTICOSTEROIDS AND ANTI-INFECTIVE COMBINATIONS Prescription count | | 0.983371 | DISCARD |
| 2-year summary (Drug prescriptions) - ANTICOAGULANTS Days since first prescription | | 0.983702 | DISCARD |
| 2-year summary (Drug prescriptions) - ANTICOAGULANTS Days since last prescription | | 0.983702 | DISCARD |
| 2-year summary (Drug prescriptions) - ANTICOAGULANTS Prescription count | | 0.983702 | DISCARD |
| 2-year summary (Drug prescriptions) - BISPHOSPHONATES Average frequency (days) | | 0.984087 | DISCARD |

| | | | |
|---|---|---|---|
| 2-year summary (Drug prescriptions) - BISPHOSPHONATES Max frequency (days) | | 0.984087 | DISCARD |
| 2-year summary (Drug prescriptions) - BISPHOSPHONATES Min frequency (days) | | 0.984087 | DISCARD |
| 1-year summary (Drug prescriptions) - INSULINS Average frequency (days) | | 0.984445 | DISCARD |
| 1-year summary (Drug prescriptions) - INSULINS Max frequency (days) | | 0.984445 | DISCARD |
| 1-year summary (Drug prescriptions) - INSULINS Min frequency (days) | | 0.984445 | DISCARD |
| 2-year summary (Drug prescriptions) - SULFONAMIDES, TRIMETROPRIM AND COMBINATION Days since first prescription | | 0.984528 | DISCARD |
| 2-year summary (Drug prescriptions) - SULFONAMIDES, TRIMETROPRIM AND COMBINATION Days since last prescription | | 0.984528 | DISCARD |
| 2-year summary (Drug prescriptions) - SULFONAMIDES, TRIMETROPRIM AND COMBINATION Prescription count | | 0.984528 | DISCARD |
| 2-year summary (Drug prescriptions) - ESTROGENS-PROGESTERONE FIXED COMBINATIONS Average frequency (days) | | 0.984638 | DISCARD |
| 2-year summary (Drug prescriptions) - ESTROGENS-PROGESTERONE FIXED COMBINATIONS Max frequency (days) | | 0.984638 | DISCARD |
| 2-year summary (Drug prescriptions) - ESTROGENS-PROGESTERONE FIXED COMBINATIONS Min frequency (days) | | 0.984638 | DISCARD |
| 1-year summary (Drug prescriptions) - ANTICONVULSANTS: MISCELLANEOUS Average frequency (days) | | 0.984748 | DISCARD |
| 1-year summary (Drug prescriptions) - ANTICONVULSANTS: MISCELLANEOUS Max frequency (days) | | 0.984748 | DISCARD |
| 1-year summary (Drug prescriptions) - ANTICONVULSANTS: MISCELLANEOUS Min frequency (days) | | 0.984748 | DISCARD |
| 2-year summary (Drug prescriptions) - GABA DERIVATIVES Days since first prescription | | 0.984913 | DISCARD |
| 2-year summary (Drug prescriptions) - GABA DERIVATIVES Days since last prescription | | 0.984913 | DISCARD |
| 2-year summary (Drug prescriptions) - GABA DERIVATIVES Prescription count | | 0.984913 | DISCARD |
| 2-year summary (Hospitalizations) - Class - Certain infectious and parasitic diseases (A00-B99) | | 0.984968 | DISCARD |
| 2-year summary (Drug prescriptions) - GAMMA-AMINOBUTYRIC ACID (GABA) DERIVATIVES Days since first prescription | | 0.984968 | DISCARD |
| 2-year summary (Drug prescriptions) - GAMMA-AMINOBUTYRIC ACID (GABA) DERIVATIVES Days since last prescription | | 0.984968 | DISCARD |
| 2-year summary (Drug prescriptions) - GAMMA-AMINOBUTYRIC ACID (GABA) DERIVATIVES Prescription count | | 0.984968 | DISCARD |
| 2-year summary (Doctor visits) - GERIATRICS | | 0.984968 | DISCARD |
| 2-year summary (Drug prescriptions) - CEPHALOSPORIN Average frequency (days) | | 0.985161 | DISCARD |
| 2-year summary (Drug prescriptions) - CEPHALOSPORIN Max frequency (days) | | 0.985161 | DISCARD |
| 2-year summary (Drug prescriptions) - CEPHALOSPORIN Min frequency (days) | | 0.985161 | DISCARD |
| 1-year summary (Hospitalizations) - Class - External causes of morbidity and mortality (V01-Y98) | | 0.985216 | DISCARD |
| 1-year summary (Drug prescriptions) - PENICILLINS Average frequency (days) | | 0.985464 | DISCARD |
| 1-year summary (Drug prescriptions) - PENICILLINS Max frequency (days) | | 0.985464 | DISCARD |
| 1-year summary (Drug prescriptions) - PENICILLINS Min frequency (days) | | 0.985464 | DISCARD |
| 2-year summary (Drug prescriptions) - MACROLIDES Average frequency (days) | | 0.985491 | DISCARD |
| 2-year summary (Drug prescriptions) - MACROLIDES Max frequency (days) | | 0.985491 | DISCARD |
| 2-year summary (Drug prescriptions) - MACROLIDES Min frequency (days) | | 0.985491 | DISCARD |
| 2-year summary (Doctor visits) - PHYSIOTHERAPY (HOME/OFFICE) | | 0.985546 | DISCARD |
| 1-year summary (Drug prescriptions) - BRONCHODILATOR AND ANTI-INFLAMMATORY COMBINATION Days since first prescriptio | | 0.985574 | DISCARD |
| 1-year summary (Drug prescriptions) - BRONCHODILATOR AND ANTI-INFLAMMATORY COMBINATION Days since last prescriptio | | 0.985574 | DISCARD |
| 1-year summary (Drug prescriptions) - BRONCHODILATOR AND ANTI-INFLAMMATORY COMBINATION Prescription count | | 0.985574 | DISCARD |
| 2-year summary (Drug prescriptions) - HISTAMINE H2 RECEPTOR ANTAGONIST Days since first prescription | | 0.985574 | DISCARD |

| | | |
|---|---|---|
| 2-year summary (Drug prescriptions) - HISTAMINE H2 RECEPTOR ANTAGONIST Days since last prescription | 0.985574 | DISCARD |
| 2-year summary (Drug prescriptions) - HISTAMINE H2 RECEPTOR ANTAGONIST Prescription count | 0.985574 | DISCARD |
| 1-year summary (Drug prescriptions) - BISPHOSPHONATES Days since first prescription | 0.985601 | DISCARD |
| 1-year summary (Drug prescriptions) - BISPHOSPHONATES Days since last prescription | 0.985601 | DISCARD |
| 1-year summary (Drug prescriptions) - BISPHOSPHONATES Prescription count | 0.985601 | DISCARD |
| 1-year summary (Drug prescriptions) - IRON PREPARATIONS Average frequency (days) | 0.985629 | DISCARD |
| 1-year summary (Drug prescriptions) - IRON PREPARATIONS Max frequency (days) | 0.985629 | DISCARD |
| 1-year summary (Drug prescriptions) - IRON PREPARATIONS Min frequency (days) | 0.985629 | DISCARD |
| 2-year summary (Drug prescriptions) - ANTIFUNGALS Days since first prescription | 0.985794 | DISCARD |
| 2-year summary (Drug prescriptions) - ANTIFUNGALS Days since last prescription | 0.985794 | DISCARD |
| 2-year summary (Drug prescriptions) - ANTIFUNGALS Prescription count | 0.985794 | DISCARD |
| 2-year summary (Drug prescriptions) - FLUOROQUINOLONES Average frequency (days) | 0.985877 | DISCARD |
| 2-year summary (Drug prescriptions) - FLUOROQUINOLONES Max frequency (days) | 0.985877 | DISCARD |
| 2-year summary (Drug prescriptions) - FLUOROQUINOLONES Min frequency (days) | 0.985877 | DISCARD |
| 2-year summary (Hospitalizations) - Days-since-last-icu | 0.986014 | DISCARD |
| 2-year summary (Hospitalizations) - ICU-counts | 0.986014 | DISCARD |
| 2-year summary (Hospitalizations) - ICU-days | 0.986014 | DISCARD |
| 1-year summary (Doctor visits) - HAEMATOLOGY | 0.986042 | DISCARD |
| 2-year summary (Drug prescriptions) - ANTICOAGULANTS Average frequency (days) | 0.986152 | DISCARD |
| 2-year summary (Drug prescriptions) - ANTICOAGULANTS Max frequency (days) | 0.986152 | DISCARD |
| 2-year summary (Drug prescriptions) - ANTICOAGULANTS Min frequency (days) | 0.986152 | DISCARD |
| 2-year summary (Drug prescriptions) - TETRACYCLINES Days since first prescription | 0.986152 | DISCARD |
| 2-year summary (Drug prescriptions) - TETRACYCLINES Days since last prescription | 0.986152 | DISCARD |
| 2-year summary (Drug prescriptions) - TETRACYCLINES Prescription count | 0.986152 | DISCARD |
| 1-year summary (Drug prescriptions) - ANTICOAGULANTS Days since first prescription | 0.986207 | DISCARD |
| 1-year summary (Drug prescriptions) - ANTICOAGULANTS Days since last prescription | 0.986207 | DISCARD |
| 1-year summary (Drug prescriptions) - ANTICOAGULANTS Prescription count | 0.986207 | DISCARD |
| 1-year summary (Hospitalizations) - Class - Diseases of the digestive system (K00-K93) | 0.986317 | DISCARD |
| 2-year summary (Drug prescriptions) - ANGIOTENSIN II COMBINATION Days since first prescription | 0.986400 | DISCARD |
| 2-year summary (Drug prescriptions) - ANGIOTENSIN II COMBINATION Days since last prescription | 0.986400 | DISCARD |
| 2-year summary (Drug prescriptions) - ANGIOTENSIN II COMBINATION Prescription count | 0.986400 | DISCARD |
| 1-year summary (Drug prescriptions) - NOT IDENTIFIED Average frequency (days) | 0.986620 | DISCARD |
| 1-year summary (Drug prescriptions) - NOT IDENTIFIED Max frequency (days) | 0.986620 | DISCARD |
| 1-year summary (Drug prescriptions) - NOT IDENTIFIED Min frequency (days) | 0.986620 | DISCARD |
| 2-year summary (Hospitalizations) - Class - Provisional codes for research and temporary assignments in Canada | 0.986730 | DISCARD |
| 1-year summary (Doctor visits) - PHYSICAL MEDICINE | 0.986785 | DISCARD |
| 2-year summary (Drug prescriptions) - BRONCHODILATOR AND ANTI-INFLAMMATORY COMBINATION Average frequency (days) | 0.986813 | DISCARD |

| | | | |
|---|---|---|---|
| 2-year summary (Drug prescriptions) - BRONCHODILATOR AND ANTI-INFLAMMATORY COMBINATION Max frequency (days) | | 0.986813 | DISCARD |
| 2-year summary (Drug prescriptions) - BRONCHODILATOR AND ANTI-INFLAMMATORY COMBINATION Min frequency (days) | | 0.986813 | DISCARD |
| 1-year summary (Drug prescriptions) - STOOL SOFTENERS Days since first prescription | | 0.986840 | DISCARD |
| 1-year summary (Drug prescriptions) - STOOL SOFTENERS Days since last prescription | | 0.986840 | DISCARD |
| 1-year summary (Drug prescriptions) - STOOL SOFTENERS Prescription count | | 0.986840 | DISCARD |
| 2-year summary (Hospitalizations) - Class - Diseases of the blood and blood-forming organs and certain disorders involving the immu | | 0.986840 | DISCARD |
| 2-year summary (Drug prescriptions) - TRICYCLIC ANTIDEPRESSANT Days since first prescription | | 0.986978 | DISCARD |
| 2-year summary (Drug prescriptions) - TRICYCLIC ANTIDEPRESSANT Days since last prescription | | 0.986978 | DISCARD |
| 2-year summary (Drug prescriptions) - TRICYCLIC ANTIDEPRESSANT Prescription count | | 0.986978 | DISCARD |
| 2-year summary (Drug prescriptions) - STOOL SOFTENERS Average frequency (days) | | 0.987033 | DISCARD |
| 2-year summary (Drug prescriptions) - STOOL SOFTENERS Max frequency (days) | | 0.987033 | DISCARD |
| 2-year summary (Drug prescriptions) - STOOL SOFTENERS Min frequency (days) | | 0.987033 | DISCARD |
| 1-year summary (Drug prescriptions) - INHALED CORTICOSTEROIDS Average frequency (days) | | 0.987116 | DISCARD |
| 1-year summary (Drug prescriptions) - INHALED CORTICOSTEROIDS Max frequency (days) | | 0.987116 | DISCARD |
| 1-year summary (Drug prescriptions) - INHALED CORTICOSTEROIDS Min frequency (days) | | 0.987116 | DISCARD |
| 2-year summary (Drug prescriptions) - ANTITUSSIVE Average frequency (days) | | 0.987116 | DISCARD |
| 2-year summary (Drug prescriptions) - ANTITUSSIVE Max frequency (days) | | 0.987116 | DISCARD |
| 2-year summary (Drug prescriptions) - ANTITUSSIVE Min frequency (days) | | 0.987116 | DISCARD |
| 1-year summary (Drug prescriptions) - BISPHOSPHONATES Average frequency (days) | | 0.987171 | DISCARD |
| 1-year summary (Drug prescriptions) - BISPHOSPHONATES Max frequency (days) | | 0.987171 | DISCARD |
| 1-year summary (Drug prescriptions) - BISPHOSPHONATES Min frequency (days) | | 0.987171 | DISCARD |
| 2-year summary (Drug prescriptions) - GASTROINTESTINAL DRUGS: MISCELLANEOUS Days since first prescription | | 0.987281 | DISCARD |
| 2-year summary (Drug prescriptions) - GASTROINTESTINAL DRUGS: MISCELLANEOUS Days since last prescription | | 0.987281 | DISCARD |
| 2-year summary (Drug prescriptions) - GASTROINTESTINAL DRUGS: MISCELLANEOUS Prescription count | | 0.987281 | DISCARD |
| 2-year summary (Drug prescriptions) - ANGIOTENSIN II COMBINATION Average frequency (days) | | 0.987308 | DISCARD |
| 2-year summary (Drug prescriptions) - ANGIOTENSIN II COMBINATION Max frequency (days) | | 0.987308 | DISCARD |
| 2-year summary (Drug prescriptions) - ANGIOTENSIN II COMBINATION Min frequency (days) | | 0.987308 | DISCARD |
| 1-year summary (Doctor visits) - INFECTIOUS DISEASE | | 0.987584 | DISCARD |
| 1-year summary (Hospitalizations) - Class - Mental and behavioural disorders (F00-F99) | | 0.987639 | DISCARD |
| 1-year summary (Doctor visits) - VASCULAR SURGERY | | 0.987639 | DISCARD |
| 2-year summary (Hospitalizations) - Class - Neoplasms (C00-D48) | | 0.987749 | DISCARD |
| 2-year summary (Hospitalizations) - Class - Diseases of the nervous system (G00-G99) | | 0.987831 | DISCARD |
| 1-year summary (Drug prescriptions) - ANTICOAGULANTS Average frequency (days) | | 0.987859 | DISCARD |
| 1-year summary (Drug prescriptions) - ANTICOAGULANTS Max frequency (days) | | 0.987859 | DISCARD |
| 1-year summary (Drug prescriptions) - ANTICOAGULANTS Min frequency (days) | | 0.987859 | DISCARD |
| 2-year summary (Drug prescriptions) - VITAMIN B12 PREP Days since first prescription | | 0.988024 | DISCARD |
| 2-year summary (Drug prescriptions) - VITAMIN B12 PREP Days since last prescription | | 0.988024 | DISCARD |

| | | | |
|---|---|---|---|
| 2-year summary (Drug prescriptions) - VITAMIN B12 PREP Prescription count | | 0.988024 | DISCARD |
| 2-year summary (Drug prescriptions) - CORONARY VASODILATORS Days since first prescription | | 0.988217 | DISCARD |
| 2-year summary (Drug prescriptions) - CORONARY VASODILATORS Days since last prescription | | 0.988217 | DISCARD |
| 2-year summary (Drug prescriptions) - CORONARY VASODILATORS Prescription count | | 0.988217 | DISCARD |
| 2-year summary (Drug prescriptions) - URINARY ANTI-INFECTIVES Average frequency (days) | | 0.988244 | DISCARD |
| 2-year summary (Drug prescriptions) - URINARY ANTI-INFECTIVES Max frequency (days) | | 0.988244 | DISCARD |
| 2-year summary (Drug prescriptions) - URINARY ANTI-INFECTIVES Min frequency (days) | | 0.988244 | DISCARD |
| 2-year summary (Drug prescriptions) - AMYNOGLYCOSIDES Days since first prescription | | 0.988272 | DISCARD |
| 2-year summary (Drug prescriptions) - AMYNOGLYCOSIDES Days since last prescription | | 0.988272 | DISCARD |
| 2-year summary (Drug prescriptions) - AMYNOGLYCOSIDES Prescription count | | 0.988272 | DISCARD |
| 1-year summary (Drug prescriptions) - ANGIOTENSIN II COMBINATION Days since first prescription | | 0.988685 | DISCARD |
| 1-year summary (Drug prescriptions) - ANGIOTENSIN II COMBINATION Days since last prescription | | 0.988685 | DISCARD |
| 1-year summary (Drug prescriptions) - ANGIOTENSIN II COMBINATION Prescription count | | 0.988685 | DISCARD |
| 2-year summary (Drug prescriptions) - ADENOSINE DIPHOSPHATE INHIBITORS Days since first prescription | | 0.988795 | DISCARD |
| 2-year summary (Drug prescriptions) - ADENOSINE DIPHOSPHATE INHIBITORS Days since last prescription | | 0.988795 | DISCARD |
| 2-year summary (Drug prescriptions) - ADENOSINE DIPHOSPHATE INHIBITORS Prescription count | | 0.988795 | DISCARD |
| 2-year summary (Drug prescriptions) - ANTI-CHOLINERGICS Days since first prescription | | 0.988795 | DISCARD |
| 2-year summary (Drug prescriptions) - ANTI-CHOLINERGICS Days since last prescription | | 0.988795 | DISCARD |
| 2-year summary (Drug prescriptions) - ANTI-CHOLINERGICS Prescription count | | 0.988795 | DISCARD |
| 2-year summary (Doctor visits) - MEDICAL ONCOLOGY | | 0.988823 | DISCARD |
| 2-year summary (Drug prescriptions) - GAMMA-AMINOBUTYRIC ACID (GABA) DERIVATIVES Average frequency (days) | | 0.988850 | DISCARD |
| 2-year summary (Drug prescriptions) - GAMMA-AMINOBUTYRIC ACID (GABA) DERIVATIVES Max frequency (days) | | 0.988850 | DISCARD |
| 2-year summary (Drug prescriptions) - GAMMA-AMINOBUTYRIC ACID (GABA) DERIVATIVES Min frequency (days) | | 0.988850 | DISCARD |
| 1-year summary (Drug prescriptions) - GAMMA-AMINOBUTYRIC ACID (GABA) DERIVATIVES Days since first prescription | | 0.988960 | DISCARD |
| 1-year summary (Drug prescriptions) - GAMMA-AMINOBUTYRIC ACID (GABA) DERIVATIVES Days since last prescription | | 0.988960 | DISCARD |
| 1-year summary (Drug prescriptions) - GAMMA-AMINOBUTYRIC ACID (GABA) DERIVATIVES Prescription count | | 0.988960 | DISCARD |
| 1-year summary (Hospitalizations) - Class - Diseases of the respiratory system (J00-J99) | | 0.988988 | DISCARD |
| 2-year summary (Drug prescriptions) - ACNE THERAPY Days since first prescription | | 0.989153 | DISCARD |
| 2-year summary (Drug prescriptions) - ACNE THERAPY Days since last prescription | | 0.989153 | DISCARD |
| 2-year summary (Drug prescriptions) - ACNE THERAPY Prescription count | | 0.989153 | DISCARD |
| 2-year summary (Drug prescriptions) - ANTIBIOTICS Average frequency (days) | | 0.989208 | DISCARD |
| 2-year summary (Drug prescriptions) - ANTIBIOTICS Max frequency (days) | | 0.989208 | DISCARD |
| 2-year summary (Drug prescriptions) - ANTIBIOTICS Min frequency (days) | | 0.989208 | DISCARD |
| 2-year summary (Drug prescriptions) - VITAMINS & MINERALS Days since first prescription | | 0.989428 | DISCARD |
| 2-year summary (Drug prescriptions) - VITAMINS & MINERALS Days since last prescription | | 0.989428 | DISCARD |
| 2-year summary (Drug prescriptions) - VITAMINS & MINERALS Prescription count | | 0.989428 | DISCARD |
| 1-year summary (Doctor visits) - GERIATRICS | | 0.989483 | DISCARD |

| | | | |
|---|---|---|---|
| 1-year summary (Drug prescriptions) - ANGIOTENSIN II COMBINATION Average frequency (days) | | 0.989759 | DISCARD |
| 1-year summary (Drug prescriptions) - ANGIOTENSIN II COMBINATION Max frequency (days) | | 0.989759 | DISCARD |
| 1-year summary (Drug prescriptions) - ANGIOTENSIN II COMBINATION Min frequency (days) | | 0.989759 | DISCARD |
| 1-year summary (Hospitalizations) - Class - Injury, poisoning and certain other consequences of external causes (S00-T98) | | 0.989786 | DISCARD |
| 1-year summary (Doctor visits) - PAEDIATRICS | | 0.989786 | DISCARD |
| 2-year summary (Drug prescriptions) - ADENOSINE DIPHOSPHATE INHIBITORS Average frequency (days) | | 0.989869 | DISCARD |
| 2-year summary (Drug prescriptions) - ADENOSINE DIPHOSPHATE INHIBITORS Max frequency (days) | | 0.989869 | DISCARD |
| 2-year summary (Drug prescriptions) - ADENOSINE DIPHOSPHATE INHIBITORS Min frequency (days) | | 0.989869 | DISCARD |
| 1-year summary (Hospitalizations) - Class - Diseases of the musculoskeletal system and connective tissue (M00-M99) | | 0.989896 | DISCARD |
| 2-year summary (Drug prescriptions) - XANTHINE OXIDASE INHIBITOR Days since first prescription | | 0.990337 | DISCARD |
| 2-year summary (Drug prescriptions) - XANTHINE OXIDASE INHIBITOR Days since last prescription | | 0.990337 | DISCARD |
| 2-year summary (Drug prescriptions) - XANTHINE OXIDASE INHIBITOR Prescription count | | 0.990337 | DISCARD |
| 1-year summary (Drug prescriptions) - BRONCHODILATOR AND ANTI-INFLAMMATORY COMBINATION Average frequency (days) | | 0.990419 | DISCARD |
| 1-year summary (Drug prescriptions) - BRONCHODILATOR AND ANTI-INFLAMMATORY COMBINATION Max frequency (days) | | 0.990419 | DISCARD |
| 1-year summary (Drug prescriptions) - BRONCHODILATOR AND ANTI-INFLAMMATORY COMBINATION Min frequency (days) | | 0.990419 | DISCARD |
| 2-year summary (Laboratory tests) - number of tests for Albumin | | 0.990447 | DISCARD |
| 2-year summary (Laboratory tests) - days since earliest test for Albumin | | 0.990447 | DISCARD |
| 2-year summary (Laboratory tests) - days since latest test for Albumin | | 0.990447 | DISCARD |
| 2-year summary (Laboratory tests) - number of high values of Albumin | | 0.990447 | DISCARD |
| 2-year summary (Laboratory tests) - highest value of Albumin | | 0.990447 | DISCARD |
| 2-year summary (Laboratory tests) - number of low values of Albumin | | 0.990447 | DISCARD |
| 2-year summary (Laboratory tests) - lowest value of Albumin | | 0.990447 | DISCARD |
| 2-year summary (Laboratory tests) - average value of Albumin | | 0.990447 | DISCARD |
| 2-year summary (Laboratory tests) - median value of Albumin | | 0.990447 | DISCARD |
| 2-year summary (Laboratory tests) - count of normal tests for Albumin | | 0.990447 | DISCARD |
| 2-year summary (Doctor visits) - NON MEDICAL PROFESSIONLS FOR IHF | | 0.990529 | DISCARD |
| 1-year summary (Drug prescriptions) - STOOL SOFTENERS Average frequency (days) | | 0.990557 | DISCARD |
| 1-year summary (Drug prescriptions) - STOOL SOFTENERS Max frequency (days) | | 0.990557 | DISCARD |
| 1-year summary (Drug prescriptions) - STOOL SOFTENERS Min frequency (days) | | 0.990557 | DISCARD |
| 1-year summary (Drug prescriptions) - ADENOSINE DIPHOSPHATE INHIBITORS Days since first prescription | | 0.990640 | DISCARD |
| 1-year summary (Drug prescriptions) - ADENOSINE DIPHOSPHATE INHIBITORS Days since last prescription | | 0.990640 | DISCARD |
| 1-year summary (Drug prescriptions) - ADENOSINE DIPHOSPHATE INHIBITORS Prescription count | | 0.990640 | DISCARD |
| 2-year summary (Drug prescriptions) - DIURETICS (POTASSIUM-SPARING) Days since first prescription | | 0.990640 | DISCARD |
| 2-year summary (Drug prescriptions) - DIURETICS (POTASSIUM-SPARING) Days since last prescription | | 0.990640 | DISCARD |
| 2-year summary (Drug prescriptions) - DIURETICS (POTASSIUM-SPARING) Prescription count | | 0.990640 | DISCARD |
| 2-year summary (Doctor visits) - THERAPEUTIC RADIOLOGY | | 0.990640 | DISCARD |
| 1-year summary (Hospitalizations) - Class - Certain infectious and parasitic diseases (A00-B99) | | 0.990695 | DISCARD |

| | | | |
|---|---|---|---|
| 2-year summary (Drug prescriptions) - ANTILIPEMIC: OTHER Days since first prescription | | 0.990777 | DISCARD |
| 2-year summary (Drug prescriptions) - ANTILIPEMIC: OTHER Days since last prescription | | 0.990777 | DISCARD |
| 2-year summary (Drug prescriptions) - ANTILIPEMIC: OTHER Prescription count | | 0.990777 | DISCARD |
| 1-year summary (Doctor visits) - NUCLEAR MEDICINE | | 0.990832 | DISCARD |
| 2-year summary (Drug prescriptions) - ASTHMA, COPD AND RELATED THERAPIES Days since first prescription | | 0.990832 | DISCARD |
| 2-year summary (Drug prescriptions) - ASTHMA, COPD AND RELATED THERAPIES Days since last prescription | | 0.990832 | DISCARD |
| 2-year summary (Drug prescriptions) - ASTHMA, COPD AND RELATED THERAPIES Prescription count | | 0.990832 | DISCARD |
| 2-year summary (Doctor visits) - NEUROSURGERY | | 0.990832 | DISCARD |
| 1-year summary (Drug prescriptions) - FUNGICIDES Average frequency (days) | | 0.990915 | DISCARD |
| 1-year summary (Drug prescriptions) - FUNGICIDES Max frequency (days) | | 0.990915 | DISCARD |
| 1-year summary (Drug prescriptions) - FUNGICIDES Min frequency (days) | | 0.990915 | DISCARD |
| 2-year summary (Drug prescriptions) - LINCOSAMIDES Days since first prescription | | 0.991053 | DISCARD |
| 2-year summary (Drug prescriptions) - LINCOSAMIDES Days since last prescription | | 0.991053 | DISCARD |
| 2-year summary (Drug prescriptions) - LINCOSAMIDES Prescription count | | 0.991053 | DISCARD |
| 1-year summary (Drug prescriptions) - TRICYCLIC ANTIDEPRESSANT Days since first prescription | | 0.991080 | DISCARD |
| 1-year summary (Drug prescriptions) - TRICYCLIC ANTIDEPRESSANT Days since last prescription | | 0.991080 | DISCARD |
| 1-year summary (Drug prescriptions) - TRICYCLIC ANTIDEPRESSANT Prescription count | | 0.991080 | DISCARD |
| 1-year summary (Drug prescriptions) - ANTIBACTERIAL/ANTIPROTOZOAL Days since first prescription | | 0.991108 | DISCARD |
| 1-year summary (Drug prescriptions) - ANTIBACTERIAL/ANTIPROTOZOAL Days since last prescription | | 0.991108 | DISCARD |
| 1-year summary (Drug prescriptions) - ANTIBACTERIAL/ANTIPROTOZOAL Prescription count | | 0.991108 | DISCARD |
| 2-year summary (Drug prescriptions) - TRICYCLIC ANTIDEPRESSANT Average frequency (days) | | 0.991135 | DISCARD |
| 2-year summary (Drug prescriptions) - TRICYCLIC ANTIDEPRESSANT Max frequency (days) | | 0.991135 | DISCARD |
| 2-year summary (Drug prescriptions) - TRICYCLIC ANTIDEPRESSANT Min frequency (days) | | 0.991135 | DISCARD |
| 2-year summary (Drug prescriptions) - XANTHINE OXIDASE INHIBITOR Average frequency (days) | | 0.991245 | DISCARD |
| 2-year summary (Drug prescriptions) - XANTHINE OXIDASE INHIBITOR Max frequency (days) | | 0.991245 | DISCARD |
| 2-year summary (Drug prescriptions) - XANTHINE OXIDASE INHIBITOR Min frequency (days) | | 0.991245 | DISCARD |
| 1-year summary (Drug prescriptions) - XANTHINE OXIDASE INHIBITOR Days since first prescription | | 0.991300 | DISCARD |
| 1-year summary (Drug prescriptions) - XANTHINE OXIDASE INHIBITOR Days since last prescription | | 0.991300 | DISCARD |
| 1-year summary (Drug prescriptions) - XANTHINE OXIDASE INHIBITOR Prescription count | | 0.991300 | DISCARD |
| 1-year summary (Doctor visits) - MEDICAL ONCOLOGY | | 0.991300 | DISCARD |
| 2-year summary (Drug prescriptions) - BONE CALCIUM REGULATOR Days since first prescription | | 0.991300 | DISCARD |
| 2-year summary (Drug prescriptions) - BONE CALCIUM REGULATOR Days since last prescription | | 0.991300 | DISCARD |
| 2-year summary (Drug prescriptions) - BONE CALCIUM REGULATOR Prescription count | | 0.991300 | DISCARD |
| 1-year summary (Drug prescriptions) - ANTI-CHOLINERGICS Days since first prescription | | 0.991328 | DISCARD |
| 1-year summary (Drug prescriptions) - ANTI-CHOLINERGICS Days since last prescription | | 0.991328 | DISCARD |
| 1-year summary (Drug prescriptions) - ANTI-CHOLINERGICS Prescription count | | 0.991328 | DISCARD |
| 1-year summary (Drug prescriptions) - GABA DERIVATIVES Days since first prescription | | 0.991383 | DISCARD |

| | | | |
|---|---|---|---|
| 1-year summary (Drug prescriptions) - GABA DERIVATIVES Days since last prescription | | 0.991383 | DISCARD |
| 1-year summary (Drug prescriptions) - GABA DERIVATIVES Prescription count | | 0.991383 | DISCARD |
| 1-year summary (Drug prescriptions) - VITAMINS & MINERALS Days since first prescription | | 0.991438 | DISCARD |
| 1-year summary (Drug prescriptions) - VITAMINS & MINERALS Days since last prescription | | 0.991438 | DISCARD |
| 1-year summary (Drug prescriptions) - VITAMINS & MINERALS Prescription count | | 0.991438 | DISCARD |
| 1-year summary (Drug prescriptions) - GAMMA-AMINOBUTYRIC ACID (GABA) DERIVATIVES Average frequency (days) | | 0.991465 | DISCARD |
| 1-year summary (Drug prescriptions) - GAMMA-AMINOBUTYRIC ACID (GABA) DERIVATIVES Max frequency (days) | | 0.991465 | DISCARD |
| 1-year summary (Drug prescriptions) - GAMMA-AMINOBUTYRIC ACID (GABA) DERIVATIVES Min frequency (days) | | 0.991465 | DISCARD |
| 2-year summary (Drug prescriptions) - ANTIDEPRESSANTS Days since first prescription | | 0.991493 | DISCARD |
| 2-year summary (Drug prescriptions) - ANTIDEPRESSANTS Days since last prescription | | 0.991493 | DISCARD |
| 2-year summary (Drug prescriptions) - ANTIDEPRESSANTS Prescription count | | 0.991493 | DISCARD |
| 2-year summary (Drug prescriptions) - CONTACT LENS SOLN, TEARS AND LUBRICANTS Days since first prescription | | 0.991521 | DISCARD |
| 2-year summary (Drug prescriptions) - CONTACT LENS SOLN, TEARS AND LUBRICANTS Days since last prescription | | 0.991521 | DISCARD |
| 2-year summary (Drug prescriptions) - CONTACT LENS SOLN, TEARS AND LUBRICANTS Prescription count | | 0.991521 | DISCARD |
| 2-year summary (Doctor visits) - CLINICAL IMMUNOLOGY | | 0.991521 | DISCARD |
| 1-year summary (Drug prescriptions) - ADENOSINE DIPHOSPHATE INHIBITORS Average frequency (days) | | 0.991548 | DISCARD |
| 1-year summary (Drug prescriptions) - ADENOSINE DIPHOSPHATE INHIBITORS Max frequency (days) | | 0.991548 | DISCARD |
| 1-year summary (Drug prescriptions) - ADENOSINE DIPHOSPHATE INHIBITORS Min frequency (days) | | 0.991548 | DISCARD |
| 2-year summary (Drug prescriptions) - ANTILIPEMIC: OTHER Average frequency (days) | | 0.991548 | DISCARD |
| 2-year summary (Drug prescriptions) - ANTILIPEMIC: OTHER Max frequency (days) | | 0.991548 | DISCARD |
| 2-year summary (Drug prescriptions) - ANTILIPEMIC: OTHER Min frequency (days) | | 0.991548 | DISCARD |
| 1-year summary (Drug prescriptions) - ANTILIPEMIC: OTHER Days since first prescription | | 0.991576 | DISCARD |
| 1-year summary (Drug prescriptions) - ANTILIPEMIC: OTHER Days since last prescription | | 0.991576 | DISCARD |
| 1-year summary (Drug prescriptions) - ANTILIPEMIC: OTHER Prescription count | | 0.991576 | DISCARD |
| 2-year summary (Drug prescriptions) - ORAL CONTRACEPTIVES Days since first prescription | | 0.991576 | DISCARD |
| 2-year summary (Drug prescriptions) - ORAL CONTRACEPTIVES Days since last prescription | | 0.991576 | DISCARD |
| 2-year summary (Drug prescriptions) - ORAL CONTRACEPTIVES Prescription count | | 0.991576 | DISCARD |
| 2-year summary (Drug prescriptions) - ANTI-CHOLINERGICS Average frequency (days) | | 0.991631 | DISCARD |
| 2-year summary (Drug prescriptions) - ANTI-CHOLINERGICS Max frequency (days) | | 0.991631 | DISCARD |
| 2-year summary (Drug prescriptions) - ANTI-CHOLINERGICS Min frequency (days) | | 0.991631 | DISCARD |
| 2-year summary (Drug prescriptions) - DIURETICS (POTASSIUM-SPARING) Average frequency (days) | | 0.991686 | DISCARD |
| 2-year summary (Drug prescriptions) - DIURETICS (POTASSIUM-SPARING) Max frequency (days) | | 0.991686 | DISCARD |
| 2-year summary (Drug prescriptions) - DIURETICS (POTASSIUM-SPARING) Min frequency (days) | | 0.991686 | DISCARD |
| 1-year summary (Drug prescriptions) - CORTICOSTEROIDS AND ANTI-INFECTIVE COMBINATIONS Days since first prescription | | 0.991713 | DISCARD |
| 1-year summary (Drug prescriptions) - CORTICOSTEROIDS AND ANTI-INFECTIVE COMBINATIONS Days since last prescription | | 0.991713 | DISCARD |
| 1-year summary (Drug prescriptions) - CORTICOSTEROIDS AND ANTI-INFECTIVE COMBINATIONS Prescription count | | 0.991713 | DISCARD |
| 2-year summary (Drug prescriptions) - HISTAMINE H2 RECEPTOR ANTAGONIST Average frequency (days) | | 0.991741 | DISCARD |

| | | | |
|---|---|---|---|
| 2-year summary (Drug prescriptions) - HISTAMINE H2 RECEPTOR ANTAGONIST Max frequency (days) | | 0.991741 | DISCARD |
| 2-year summary (Drug prescriptions) - HISTAMINE H2 RECEPTOR ANTAGONIST Min frequency (days) | | 0.991741 | DISCARD |
| 2-year summary (Drug prescriptions) - DIABETES MELLITUS Days since first prescription | | 0.991796 | DISCARD |
| 2-year summary (Drug prescriptions) - DIABETES MELLITUS Days since last prescription | | 0.991796 | DISCARD |
| 2-year summary (Drug prescriptions) - DIABETES MELLITUS Prescription count | | 0.991796 | DISCARD |
| 1-year summary (Drug prescriptions) - SULFONAMIDES, TRIMETROPRIM AND COMBINATION Days since first prescription | | 0.991823 | DISCARD |
| 1-year summary (Drug prescriptions) - SULFONAMIDES, TRIMETROPRIM AND COMBINATION Days since last prescription | | 0.991823 | DISCARD |
| 1-year summary (Drug prescriptions) - SULFONAMIDES, TRIMETROPRIM AND COMBINATION Prescription count | | 0.991823 | DISCARD |
| 2-year summary (Drug prescriptions) - CHOLINESTERASE INHIBITORS Days since first prescription | | 0.991933 | DISCARD |
| 2-year summary (Drug prescriptions) - CHOLINESTERASE INHIBITORS Days since last prescription | | 0.991933 | DISCARD |
| 2-year summary (Drug prescriptions) - CHOLINESTERASE INHIBITORS Prescription count | | 0.991933 | DISCARD |
| 1-year summary (Hospitalizations) - Class - Provisional codes for research and temporary assignments in Canada | | 0.992044 | DISCARD |
| 1-year summary (Hospitalizations) - Days-since-last-icu | | 0.992071 | DISCARD |
| 1-year summary (Hospitalizations) - ICU-counts | | 0.992071 | DISCARD |
| 1-year summary (Hospitalizations) - ICU-days | | 0.992071 | DISCARD |
| 1-year summary (Drug prescriptions) - GASTROINTESTINAL DRUGS: MISCELLANEOUS Days since first prescription | | 0.992071 | DISCARD |
| 1-year summary (Drug prescriptions) - GASTROINTESTINAL DRUGS: MISCELLANEOUS Days since last prescription | | 0.992071 | DISCARD |
| 1-year summary (Drug prescriptions) - GASTROINTESTINAL DRUGS: MISCELLANEOUS Prescription count | | 0.992071 | DISCARD |
| 1-year summary (Drug prescriptions) - XANTHINE OXIDASE INHIBITOR Average frequency (days) | | 0.992071 | DISCARD |
| 1-year summary (Drug prescriptions) - XANTHINE OXIDASE INHIBITOR Max frequency (days) | | 0.992071 | DISCARD |
| 1-year summary (Drug prescriptions) - XANTHINE OXIDASE INHIBITOR Min frequency (days) | | 0.992071 | DISCARD |
| 2-year summary (Drug prescriptions) - VITAMINS & MINERALS Average frequency (days) | | 0.992071 | DISCARD |
| 2-year summary (Drug prescriptions) - VITAMINS & MINERALS Max frequency (days) | | 0.992071 | DISCARD |
| 2-year summary (Drug prescriptions) - VITAMINS & MINERALS Min frequency (days) | | 0.992071 | DISCARD |
| 1-year summary (Drug prescriptions) - ANTILIPEMIC: OTHER Average frequency (days) | | 0.992099 | DISCARD |
| 1-year summary (Drug prescriptions) - ANTILIPEMIC: OTHER Max frequency (days) | | 0.992099 | DISCARD |
| 1-year summary (Drug prescriptions) - ANTILIPEMIC: OTHER Min frequency (days) | | 0.992099 | DISCARD |
| 2-year summary (Drug prescriptions) - ACE INHIBITORS COMBINATION Days since first prescription | | 0.992099 | DISCARD |
| 2-year summary (Drug prescriptions) - ACE INHIBITORS COMBINATION Days since last prescription | | 0.992099 | DISCARD |
| 2-year summary (Drug prescriptions) - ACE INHIBITORS COMBINATION Prescription count | | 0.992099 | DISCARD |
| 1-year summary (Hospitalizations) - Class - Pregnancy, childbirth and the puerperium (O00-O99) | | 0.992154 | DISCARD |
| 2-year summary (Drug prescriptions) - CHOLINESTERASE INHIBITORS Average frequency (days) | | 0.992181 | DISCARD |
| 2-year summary (Drug prescriptions) - CHOLINESTERASE INHIBITORS Max frequency (days) | | 0.992181 | DISCARD |
| 2-year summary (Drug prescriptions) - CHOLINESTERASE INHIBITORS Min frequency (days) | | 0.992181 | DISCARD |
| 1-year summary (Drug prescriptions) - BONE CALCIUM REGULATOR Days since first prescription | | 0.992209 | DISCARD |
| 1-year summary (Drug prescriptions) - BONE CALCIUM REGULATOR Days since last prescription | | 0.992209 | DISCARD |
| 1-year summary (Drug prescriptions) - BONE CALCIUM REGULATOR Prescription count | | 0.992209 | DISCARD |

| | | | |
|---|---|---|---|
| 1-year summary (Drug prescriptions) - ASTHMA, COPD AND RELATED THERAPIES Days since first prescription | | 0.992374 | DISCARD |
| 1-year summary (Drug prescriptions) - ASTHMA, COPD AND RELATED THERAPIES Days since last prescription | | 0.992374 | DISCARD |
| 1-year summary (Drug prescriptions) - ASTHMA, COPD AND RELATED THERAPIES Prescription count | | 0.992374 | DISCARD |
| 1-year summary (Drug prescriptions) - DIABETES MELLITUS Days since first prescription | | 0.992374 | DISCARD |
| 1-year summary (Drug prescriptions) - DIABETES MELLITUS Days since last prescription | | 0.992374 | DISCARD |
| 1-year summary (Drug prescriptions) - DIABETES MELLITUS Prescription count | | 0.992374 | DISCARD |
| 1-year summary (Doctor visits) - PHYSIOTHERAPY (HOME/OFFICE) | | 0.992402 | DISCARD |
| 1-year summary (Hospitalizations) - Class - Diseases of the blood and blood-forming organs and certain disorders involving the immu | | 0.992429 | DISCARD |
| 2-year summary (Drug prescriptions) - GENITOURINARY SMOOTH MUSCLE RELAXANTS Days since first prescription | | 0.992457 | DISCARD |
| 2-year summary (Drug prescriptions) - GENITOURINARY SMOOTH MUSCLE RELAXANTS Days since last prescription | | 0.992457 | DISCARD |
| 2-year summary (Drug prescriptions) - GENITOURINARY SMOOTH MUSCLE RELAXANTS Prescription count | | 0.992457 | DISCARD |
| 1-year summary (Drug prescriptions) - DIURETICS (POTASSIUM-SPARING) Days since first prescription | | 0.992512 | DISCARD |
| 1-year summary (Drug prescriptions) - DIURETICS (POTASSIUM-SPARING) Days since last prescription | | 0.992512 | DISCARD |
| 1-year summary (Drug prescriptions) - DIURETICS (POTASSIUM-SPARING) Prescription count | | 0.992512 | DISCARD |
| 2-year summary (Doctor visits) - CARDIOTHORACIC SURGERY | | 0.992512 | DISCARD |
| 2-year summary (Drug prescriptions) - BETA BLOCKING AGENTS Days since first prescription | | 0.992594 | DISCARD |
| 2-year summary (Drug prescriptions) - BETA BLOCKING AGENTS Days since last prescription | | 0.992594 | DISCARD |
| 2-year summary (Drug prescriptions) - BETA BLOCKING AGENTS Prescription count | | 0.992594 | DISCARD |
| 1-year summary (Drug prescriptions) - ESTROGENS-PROGESTERONE FIXED COMBINATIONS Days since first prescription | | 0.992677 | DISCARD |
| 1-year summary (Drug prescriptions) - ESTROGENS-PROGESTERONE FIXED COMBINATIONS Days since last prescription | | 0.992677 | DISCARD |
| 1-year summary (Drug prescriptions) - ESTROGENS-PROGESTERONE FIXED COMBINATIONS Prescription count | | 0.992677 | DISCARD |
| 2-year summary (Drug prescriptions) - REPLACEMENT AGENTS Days since first prescription | | 0.992704 | DISCARD |
| 2-year summary (Drug prescriptions) - REPLACEMENT AGENTS Days since last prescription | | 0.992704 | DISCARD |
| 2-year summary (Drug prescriptions) - REPLACEMENT AGENTS Prescription count | | 0.992704 | DISCARD |
| 1-year summary (Hospitalizations) - Class - Diseases of the nervous system (G00-G99) | | 0.992732 | DISCARD |
| 1-year summary (Drug prescriptions) - CORONARY VASODILATORS Days since first prescription | | 0.992787 | DISCARD |
| 1-year summary (Drug prescriptions) - CORONARY VASODILATORS Days since last prescription | | 0.992787 | DISCARD |
| 1-year summary (Drug prescriptions) - CORONARY VASODILATORS Prescription count | | 0.992787 | DISCARD |
| 1-year summary (Drug prescriptions) - HISTAMINE H2 RECEPTOR ANTAGONIST Days since first prescription | | 0.992787 | DISCARD |
| 1-year summary (Drug prescriptions) - HISTAMINE H2 RECEPTOR ANTAGONIST Days since last prescription | | 0.992787 | DISCARD |
| 1-year summary (Drug prescriptions) - HISTAMINE H2 RECEPTOR ANTAGONIST Prescription count | | 0.992787 | DISCARD |
| 2-year summary (Drug prescriptions) - GASTROINTESTINAL DRUGS: MISCELLANEOUS Average frequency (days) | | 0.992787 | DISCARD |
| 2-year summary (Drug prescriptions) - GASTROINTESTINAL DRUGS: MISCELLANEOUS Max frequency (days) | | 0.992787 | DISCARD |
| 2-year summary (Drug prescriptions) - GASTROINTESTINAL DRUGS: MISCELLANEOUS Min frequency (days) | | 0.992787 | DISCARD |
| 2-year summary (Drug prescriptions) - OTHER ANTI-INFLMMATORY AGENTS Days since first prescription | | 0.992842 | DISCARD |
| 2-year summary (Drug prescriptions) - OTHER ANTI-INFLMMATORY AGENTS Days since last prescription | | 0.992842 | DISCARD |
| 2-year summary (Drug prescriptions) - OTHER ANTI-INFLMMATORY AGENTS Prescription count | | 0.992842 | DISCARD |

| | | | |
|---|---|---|---|
| 1-year summary (Drug prescriptions) - ANTIFUNGALS Days since first prescription | | 0.992870 | DISCARD |
| 1-year summary (Drug prescriptions) - ANTIFUNGALS Days since last prescription | | 0.992870 | DISCARD |
| 1-year summary (Drug prescriptions) - ANTIFUNGALS Prescription count | | 0.992870 | DISCARD |
| 1-year summary (Drug prescriptions) - CHOLINESTERASE INHIBITORS Days since first prescription | | 0.992925 | DISCARD |
| 1-year summary (Drug prescriptions) - CHOLINESTERASE INHIBITORS Days since last prescription | | 0.992925 | DISCARD |
| 1-year summary (Drug prescriptions) - CHOLINESTERASE INHIBITORS Prescription count | | 0.992925 | DISCARD |
| 2-year summary (Drug prescriptions) - ACE INHIBITORS COMBINATION Average frequency (days) | | 0.992980 | DISCARD |
| 2-year summary (Drug prescriptions) - ACE INHIBITORS COMBINATION Max frequency (days) | | 0.992980 | DISCARD |
| 2-year summary (Drug prescriptions) - ACE INHIBITORS COMBINATION Min frequency (days) | | 0.992980 | DISCARD |
| 1-year summary (Hospitalizations) - Class - Neoplasms (C00-D48) | | 0.993007 | DISCARD |
| 2-year summary (Drug prescriptions) - ANTIDEPRESSANTS Average frequency (days) | | 0.993007 | DISCARD |
| 2-year summary (Drug prescriptions) - ANTIDEPRESSANTS Max frequency (days) | | 0.993007 | DISCARD |
| 2-year summary (Drug prescriptions) - ANTIDEPRESSANTS Min frequency (days) | | 0.993007 | DISCARD |
| 1-year summary (Drug prescriptions) - VITAMIN B12 PREP Days since first prescription | | 0.993090 | DISCARD |
| 1-year summary (Drug prescriptions) - VITAMIN B12 PREP Days since last prescription | | 0.993090 | DISCARD |
| 1-year summary (Drug prescriptions) - VITAMIN B12 PREP Prescription count | | 0.993090 | DISCARD |
| 2-year summary (Drug prescriptions) - ASTHMA, COPD AND RELATED THERAPIES Average frequency (days) | | 0.993090 | DISCARD |
| 2-year summary (Drug prescriptions) - ASTHMA, COPD AND RELATED THERAPIES Max frequency (days) | | 0.993090 | DISCARD |
| 2-year summary (Drug prescriptions) - ASTHMA, COPD AND RELATED THERAPIES Min frequency (days) | | 0.993090 | DISCARD |
| 2-year summary (Drug prescriptions) - PROSTAGLANDIN ANALOGS Days since first prescription | | 0.993090 | DISCARD |
| 2-year summary (Drug prescriptions) - PROSTAGLANDIN ANALOGS Days since last prescription | | 0.993090 | DISCARD |
| 2-year summary (Drug prescriptions) - PROSTAGLANDIN ANALOGS Prescription count | | 0.993090 | DISCARD |
| 2-year summary (Drug prescriptions) - ANTIEMETICS AND ANTINAUSEANTS Days since first prescription | | 0.993172 | DISCARD |
| 2-year summary (Drug prescriptions) - ANTIEMETICS AND ANTINAUSEANTS Days since last prescription | | 0.993172 | DISCARD |
| 2-year summary (Drug prescriptions) - ANTIEMETICS AND ANTINAUSEANTS Prescription count | | 0.993172 | DISCARD |
| 1-year summary (Drug prescriptions) - VITAMINS & MINERALS Average frequency (days) | | 0.993200 | DISCARD |
| 1-year summary (Drug prescriptions) - VITAMINS & MINERALS Max frequency (days) | | 0.993200 | DISCARD |
| 1-year summary (Drug prescriptions) - VITAMINS & MINERALS Min frequency (days) | | 0.993200 | DISCARD |
| 1-year summary (Drug prescriptions) - CHOLINESTERASE INHIBITORS Average frequency (days) | | 0.993255 | DISCARD |
| 1-year summary (Drug prescriptions) - CHOLINESTERASE INHIBITORS Max frequency (days) | | 0.993255 | DISCARD |
| 1-year summary (Drug prescriptions) - CHOLINESTERASE INHIBITORS Min frequency (days) | | 0.993255 | DISCARD |
| 1-year summary (Doctor visits) - THERAPEUTIC RADIOLOGY | | 0.993255 | DISCARD |
| 1-year summary (Drug prescriptions) - ACE INHIBITORS COMBINATION Days since first prescription | | 0.993282 | DISCARD |
| 1-year summary (Drug prescriptions) - ACE INHIBITORS COMBINATION Days since last prescription | | 0.993282 | DISCARD |
| 1-year summary (Drug prescriptions) - ACE INHIBITORS COMBINATION Prescription count | | 0.993282 | DISCARD |
| 1-year summary (Drug prescriptions) - ANTI-CHOLINERGICS Average frequency (days) | | 0.993282 | DISCARD |
| 1-year summary (Drug prescriptions) - ANTI-CHOLINERGICS Max frequency (days) | | 0.993282 | DISCARD |

| | | | |
|---|---|---|---|
| 1-year summary (Drug prescriptions) - ANTI-CHOLINERGICS Min frequency (days) | | 0.993282 | DISCARD |
| 1-year summary (Drug prescriptions) - DIURETICS (POTASSIUM-SPARING) Average frequency (days) | | 0.993338 | DISCARD |
| 1-year summary (Drug prescriptions) - DIURETICS (POTASSIUM-SPARING) Max frequency (days) | | 0.993338 | DISCARD |
| 1-year summary (Drug prescriptions) - DIURETICS (POTASSIUM-SPARING) Min frequency (days) | | 0.993338 | DISCARD |
| 2-year summary (Drug prescriptions) - SELECTIVE SEROTONIN REUPTAKE INHIBITORS - OTHER Days since first prescription | | 0.993338 | DISCARD |
| 2-year summary (Drug prescriptions) - SELECTIVE SEROTONIN REUPTAKE INHIBITORS - OTHER Days since last prescription | | 0.993338 | DISCARD |
| 2-year summary (Drug prescriptions) - SELECTIVE SEROTONIN REUPTAKE INHIBITORS - OTHER Prescription count | | 0.993338 | DISCARD |
| 1-year summary (Drug prescriptions) - FLUOROQUINOLONES Average frequency (days) | | 0.993365 | DISCARD |
| 1-year summary (Drug prescriptions) - FLUOROQUINOLONES Max frequency (days) | | 0.993365 | DISCARD |
| 1-year summary (Drug prescriptions) - FLUOROQUINOLONES Min frequency (days) | | 0.993365 | DISCARD |
| 1-year summary (Drug prescriptions) - TRICYCLIC ANTIDEPRESSANT Average frequency (days) | | 0.993393 | DISCARD |
| 1-year summary (Drug prescriptions) - TRICYCLIC ANTIDEPRESSANT Max frequency (days) | | 0.993393 | DISCARD |
| 1-year summary (Drug prescriptions) - TRICYCLIC ANTIDEPRESSANT Min frequency (days) | | 0.993393 | DISCARD |
| 1-year summary (Drug prescriptions) - TETRACYCLINES Days since first prescription | | 0.993420 | DISCARD |
| 1-year summary (Drug prescriptions) - TETRACYCLINES Days since last prescription | | 0.993420 | DISCARD |
| 1-year summary (Drug prescriptions) - TETRACYCLINES Prescription count | | 0.993420 | DISCARD |
| 2-year summary (Drug prescriptions) - GENITOURINARY SMOOTH MUSCLE RELAXANTS Average frequency (days) | | 0.993420 | DISCARD |
| 2-year summary (Drug prescriptions) - GENITOURINARY SMOOTH MUSCLE RELAXANTS Max frequency (days) | | 0.993420 | DISCARD |
| 2-year summary (Drug prescriptions) - GENITOURINARY SMOOTH MUSCLE RELAXANTS Min frequency (days) | | 0.993420 | DISCARD |
| 1-year summary (Drug prescriptions) - ANTIDEPRESSANTS Days since first prescription | | 0.993503 | DISCARD |
| 1-year summary (Drug prescriptions) - ANTIDEPRESSANTS Days since last prescription | | 0.993503 | DISCARD |
| 1-year summary (Drug prescriptions) - ANTIDEPRESSANTS Prescription count | | 0.993503 | DISCARD |
| 2-year summary (Drug prescriptions) - ESTROGENS, PLAIN Days since first prescription | | 0.993530 | DISCARD |
| 2-year summary (Drug prescriptions) - ESTROGENS, PLAIN Days since last prescription | | 0.993530 | DISCARD |
| 2-year summary (Drug prescriptions) - ESTROGENS, PLAIN Prescription count | | 0.993530 | DISCARD |
| 2-year summary (Drug prescriptions) - BONE CALCIUM REGULATOR Average frequency (days) | | 0.993558 | DISCARD |
| 2-year summary (Drug prescriptions) - BONE CALCIUM REGULATOR Max frequency (days) | | 0.993558 | DISCARD |
| 2-year summary (Drug prescriptions) - BONE CALCIUM REGULATOR Min frequency (days) | | 0.993558 | DISCARD |
| 2-year summary (Doctor visits) - CHIROPODISTS | | 0.993640 | DISCARD |
| 1-year summary (Drug prescriptions) - CEPHALOSPORIN Average frequency (days) | | 0.993668 | DISCARD |
| 1-year summary (Drug prescriptions) - CEPHALOSPORIN Max frequency (days) | | 0.993668 | DISCARD |
| 1-year summary (Drug prescriptions) - CEPHALOSPORIN Min frequency (days) | | 0.993668 | DISCARD |
| 2-year summary (Drug prescriptions) - BETA BLOCKING AGENTS Average frequency (days) | | 0.993695 | DISCARD |
| 2-year summary (Drug prescriptions) - BETA BLOCKING AGENTS Max frequency (days) | | 0.993695 | DISCARD |
| 2-year summary (Drug prescriptions) - BETA BLOCKING AGENTS Min frequency (days) | | 0.993695 | DISCARD |
| 2-year summary (Drug prescriptions) - DIABETES MELLITUS Average frequency (days) | | 0.993695 | DISCARD |
| 2-year summary (Drug prescriptions) - DIABETES MELLITUS Max frequency (days) | | 0.993695 | DISCARD |

| | | | |
|---|---|---|---|
| 2-year summary (Drug prescriptions) - DIABETES MELLITUS Min frequency (days) | | 0.993695 | DISCARD |
| 2-year summary (Doctor visits) - NURSE PRACTITIONERS | | 0.993695 | DISCARD |
| 1-year summary (Drug prescriptions) - GENITOURINARY SMOOTH MUSCLE RELAXANTS Days since first prescription | | 0.993778 | DISCARD |
| 1-year summary (Drug prescriptions) - GENITOURINARY SMOOTH MUSCLE RELAXANTS Days since last prescription | | 0.993778 | DISCARD |
| 1-year summary (Drug prescriptions) - GENITOURINARY SMOOTH MUSCLE RELAXANTS Prescription count | | 0.993778 | DISCARD |
| 2-year summary (Drug prescriptions) - PROSTAGLANDIN ANALOGS Average frequency (days) | | 0.993806 | DISCARD |
| 2-year summary (Drug prescriptions) - PROSTAGLANDIN ANALOGS Max frequency (days) | | 0.993806 | DISCARD |
| 2-year summary (Drug prescriptions) - PROSTAGLANDIN ANALOGS Min frequency (days) | | 0.993806 | DISCARD |
| 2-year summary (Hospitalizations) - Class - Diseases of the skin and subcutaneous tissue (L00-L99) | | 0.993888 | DISCARD |
| 2-year summary (Drug prescriptions) - ALPHA ADRENERGIC BLOCKING AGENTS Days since first prescription | | 0.993888 | DISCARD |
| 2-year summary (Drug prescriptions) - ALPHA ADRENERGIC BLOCKING AGENTS Days since last prescription | | 0.993888 | DISCARD |
| 2-year summary (Drug prescriptions) - ALPHA ADRENERGIC BLOCKING AGENTS Prescription count | | 0.993888 | DISCARD |
| 1-year summary (Drug prescriptions) - ANTITUSSIVE Average frequency (days) | | 0.993916 | DISCARD |
| 1-year summary (Drug prescriptions) - ANTITUSSIVE Max frequency (days) | | 0.993916 | DISCARD |
| 1-year summary (Drug prescriptions) - ANTITUSSIVE Min frequency (days) | | 0.993916 | DISCARD |
| 1-year summary (Drug prescriptions) - PROSTAGLANDIN ANALOGS Days since first prescription | | 0.993971 | DISCARD |
| 1-year summary (Drug prescriptions) - PROSTAGLANDIN ANALOGS Days since last prescription | | 0.993971 | DISCARD |
| 1-year summary (Drug prescriptions) - PROSTAGLANDIN ANALOGS Prescription count | | 0.993971 | DISCARD |
| 2-year summary (Drug prescriptions) - ANTIVIRAL AGENTS Days since first prescription | | 0.993971 | DISCARD |
| 2-year summary (Drug prescriptions) - ANTIVIRAL AGENTS Days since last prescription | | 0.993971 | DISCARD |
| 2-year summary (Drug prescriptions) - ANTIVIRAL AGENTS Prescription count | | 0.993971 | DISCARD |
| 1-year summary (Drug prescriptions) - AMYNOGLYCOSIDES Days since first prescription | | 0.994053 | DISCARD |
| 1-year summary (Drug prescriptions) - AMYNOGLYCOSIDES Days since last prescription | | 0.994053 | DISCARD |
| 1-year summary (Drug prescriptions) - AMYNOGLYCOSIDES Prescription count | | 0.994053 | DISCARD |
| 1-year summary (Drug prescriptions) - BETA BLOCKING AGENTS Days since first prescription | | 0.994053 | DISCARD |
| 1-year summary (Drug prescriptions) - BETA BLOCKING AGENTS Days since last prescription | | 0.994053 | DISCARD |
| 1-year summary (Drug prescriptions) - BETA BLOCKING AGENTS Prescription count | | 0.994053 | DISCARD |
| 1-year summary (Drug prescriptions) - DIABETES MELLITUS Average frequency (days) | | 0.994081 | DISCARD |
| 1-year summary (Drug prescriptions) - DIABETES MELLITUS Max frequency (days) | | 0.994081 | DISCARD |
| 1-year summary (Drug prescriptions) - DIABETES MELLITUS Min frequency (days) | | 0.994081 | DISCARD |
| 1-year summary (Drug prescriptions) - ACE INHIBITORS COMBINATION Average frequency (days) | | 0.994163 | DISCARD |
| 1-year summary (Drug prescriptions) - ACE INHIBITORS COMBINATION Max frequency (days) | | 0.994163 | DISCARD |
| 1-year summary (Drug prescriptions) - ACE INHIBITORS COMBINATION Min frequency (days) | | 0.994163 | DISCARD |
| 2-year summary (Drug prescriptions) - GABA DERIVATIVES Average frequency (days) | | 0.994219 | DISCARD |
| 2-year summary (Drug prescriptions) - GABA DERIVATIVES Max frequency (days) | | 0.994219 | DISCARD |
| 2-year summary (Drug prescriptions) - GABA DERIVATIVES Min frequency (days) | | 0.994219 | DISCARD |
| 1-year summary (Doctor visits) - NEUROSURGERY | | 0.994246 | DISCARD |

| | | | |
|---|---|---|---|
| 2-year summary (Drug prescriptions) - ORAL CONTRACEPTIVES Average frequency (days) | | 0.994274 | DISCARD |
| 2-year summary (Drug prescriptions) - ORAL CONTRACEPTIVES Max frequency (days) | | 0.994274 | DISCARD |
| 2-year summary (Drug prescriptions) - ORAL CONTRACEPTIVES Min frequency (days) | | 0.994274 | DISCARD |
| 1-year summary (Drug prescriptions) - ASTHMA, COPD AND RELATED THERAPIES Average frequency (days) | | 0.994329 | DISCARD |
| 1-year summary (Drug prescriptions) - ASTHMA, COPD AND RELATED THERAPIES Max frequency (days) | | 0.994329 | DISCARD |
| 1-year summary (Drug prescriptions) - ASTHMA, COPD AND RELATED THERAPIES Min frequency (days) | | 0.994329 | DISCARD |
| 2-year summary (Drug prescriptions) - SELECTIVE SEROTONIN REUPTAKE INHIBITORS - OTHER Average frequency (days) | | 0.994356 | DISCARD |
| 2-year summary (Drug prescriptions) - SELECTIVE SEROTONIN REUPTAKE INHIBITORS - OTHER Max frequency (days) | | 0.994356 | DISCARD |
| 2-year summary (Drug prescriptions) - SELECTIVE SEROTONIN REUPTAKE INHIBITORS - OTHER Min frequency (days) | | 0.994356 | DISCARD |
| 1-year summary (Drug prescriptions) - ANTIDEPRESSANTS Average frequency (days) | | 0.994384 | DISCARD |
| 1-year summary (Drug prescriptions) - ANTIDEPRESSANTS Max frequency (days) | | 0.994384 | DISCARD |
| 1-year summary (Drug prescriptions) - ANTIDEPRESSANTS Min frequency (days) | | 0.994384 | DISCARD |
| 1-year summary (Drug prescriptions) - ESTROGENS-PROGESTERONE FIXED COMBINATIONS Average frequency (days) | | 0.994439 | DISCARD |
| 1-year summary (Drug prescriptions) - ESTROGENS-PROGESTERONE FIXED COMBINATIONS Max frequency (days) | | 0.994439 | DISCARD |
| 1-year summary (Drug prescriptions) - ESTROGENS-PROGESTERONE FIXED COMBINATIONS Min frequency (days) | | 0.994439 | DISCARD |
| 1-year summary (Drug prescriptions) - GENITOURINARY SMOOTH MUSCLE RELAXANTS Average frequency (days) | | 0.994521 | DISCARD |
| 1-year summary (Drug prescriptions) - GENITOURINARY SMOOTH MUSCLE RELAXANTS Max frequency (days) | | 0.994521 | DISCARD |
| 1-year summary (Drug prescriptions) - GENITOURINARY SMOOTH MUSCLE RELAXANTS Min frequency (days) | | 0.994521 | DISCARD |
| 2-year summary (Drug prescriptions) - ALPHA ADRENERGIC BLOCKING AGENTS Average frequency (days) | | 0.994521 | DISCARD |
| 2-year summary (Drug prescriptions) - ALPHA ADRENERGIC BLOCKING AGENTS Max frequency (days) | | 0.994521 | DISCARD |
| 2-year summary (Drug prescriptions) - ALPHA ADRENERGIC BLOCKING AGENTS Min frequency (days) | | 0.994521 | DISCARD |
| 2-year summary (Drug prescriptions) - CORONARY VASODILATORS Average frequency (days) | | 0.994521 | DISCARD |
| 2-year summary (Drug prescriptions) - CORONARY VASODILATORS Max frequency (days) | | 0.994521 | DISCARD |
| 2-year summary (Drug prescriptions) - CORONARY VASODILATORS Min frequency (days) | | 0.994521 | DISCARD |
| 1-year summary (Drug prescriptions) - URINARY ANTI-INFECTIVES Average frequency (days) | | 0.994549 | DISCARD |
| 1-year summary (Drug prescriptions) - URINARY ANTI-INFECTIVES Max frequency (days) | | 0.994549 | DISCARD |
| 1-year summary (Drug prescriptions) - URINARY ANTI-INFECTIVES Min frequency (days) | | 0.994549 | DISCARD |
| 2-year summary (Drug prescriptions) - UNCLASSIFIED THERAPEUTIC AGENTS Days since first prescription | | 0.994576 | DISCARD |
| 2-year summary (Drug prescriptions) - UNCLASSIFIED THERAPEUTIC AGENTS Days since last prescription | | 0.994576 | DISCARD |
| 2-year summary (Drug prescriptions) - UNCLASSIFIED THERAPEUTIC AGENTS Prescription count | | 0.994576 | DISCARD |
| 1-year summary (Drug prescriptions) - PROSTAGLANDIN ANALOGS Average frequency (days) | | 0.994714 | DISCARD |
| 1-year summary (Drug prescriptions) - PROSTAGLANDIN ANALOGS Max frequency (days) | | 0.994714 | DISCARD |
| 1-year summary (Drug prescriptions) - PROSTAGLANDIN ANALOGS Min frequency (days) | | 0.994714 | DISCARD |
| 1-year summary (Drug prescriptions) - CONTACT LENS SOLN, TEARS AND LUBRICANTS Days since first prescription | | 0.994742 | DISCARD |
| 1-year summary (Drug prescriptions) - CONTACT LENS SOLN, TEARS AND LUBRICANTS Days since last prescription | | 0.994742 | DISCARD |
| 1-year summary (Drug prescriptions) - CONTACT LENS SOLN, TEARS AND LUBRICANTS Prescription count | | 0.994742 | DISCARD |
| 1-year summary (Drug prescriptions) - ACNE THERAPY Days since first prescription | | 0.994824 | DISCARD |

| | | | |
|---|---|---|---|
| 1-year summary (Drug prescriptions) - ACNE THERAPY Days since last prescription | | 0.994824 | DISCARD |
| 1-year summary (Drug prescriptions) - ACNE THERAPY Prescription count | | 0.994824 | DISCARD |
| 2-year summary (Drug prescriptions) - ACNE THERAPY Average frequency (days) | | 0.994824 | DISCARD |
| 2-year summary (Drug prescriptions) - ACNE THERAPY Max frequency (days) | | 0.994824 | DISCARD |
| 2-year summary (Drug prescriptions) - ACNE THERAPY Min frequency (days) | | 0.994824 | DISCARD |
| 2-year summary (Drug prescriptions) - VITAMIN B12 PREP Average frequency (days) | | 0.994824 | DISCARD |
| 2-year summary (Drug prescriptions) - VITAMIN B12 PREP Max frequency (days) | | 0.994824 | DISCARD |
| 2-year summary (Drug prescriptions) - VITAMIN B12 PREP Min frequency (days) | | 0.994824 | DISCARD |
| 2-year summary (Drug prescriptions) - C.N.S. STIMULANTS Days since first prescription | | 0.994852 | DISCARD |
| 2-year summary (Drug prescriptions) - C.N.S. STIMULANTS Days since last prescription | | 0.994852 | DISCARD |
| 2-year summary (Drug prescriptions) - C.N.S. STIMULANTS Prescription count | | 0.994852 | DISCARD |
| 1-year summary (Drug prescriptions) - ALPHA ADRENERGIC BLOCKING AGENTS Days since first prescription | | 0.994879 | DISCARD |
| 1-year summary (Drug prescriptions) - ALPHA ADRENERGIC BLOCKING AGENTS Days since last prescription | | 0.994879 | DISCARD |
| 1-year summary (Drug prescriptions) - ALPHA ADRENERGIC BLOCKING AGENTS Prescription count | | 0.994879 | DISCARD |
| 1-year summary (Drug prescriptions) - GASTROINTESTINAL DRUGS: MISCELLANEOUS Average frequency (days) | | 0.994907 | DISCARD |
| 1-year summary (Drug prescriptions) - GASTROINTESTINAL DRUGS: MISCELLANEOUS Max frequency (days) | | 0.994907 | DISCARD |
| 1-year summary (Drug prescriptions) - GASTROINTESTINAL DRUGS: MISCELLANEOUS Min frequency (days) | | 0.994907 | DISCARD |
| 1-year summary (Drug prescriptions) - SELECTIVE SEROTONIN REUPTAKE INHIBITORS - OTHER Days since first prescription | | 0.994907 | DISCARD |
| 1-year summary (Drug prescriptions) - SELECTIVE SEROTONIN REUPTAKE INHIBITORS - OTHER Days since last prescription | | 0.994907 | DISCARD |
| 1-year summary (Drug prescriptions) - SELECTIVE SEROTONIN REUPTAKE INHIBITORS - OTHER Prescription count | | 0.994907 | DISCARD |
| 1-year summary (Drug prescriptions) - MACROLIDES Average frequency (days) | | 0.994962 | DISCARD |
| 1-year summary (Drug prescriptions) - MACROLIDES Max frequency (days) | | 0.994962 | DISCARD |
| 1-year summary (Drug prescriptions) - MACROLIDES Min frequency (days) | | 0.994962 | DISCARD |
| 2-year summary (Laboratory tests) - number of tests for Glucose^1H_post_50_g_glucose_PO | | 0.994989 | DISCARD |
| 2-year summary (Laboratory tests) - days since earliest test for Glucose^1H_post_50_g_glucose_PO | | 0.994989 | DISCARD |
| 2-year summary (Laboratory tests) - days since latest test for Glucose^1H_post_50_g_glucose_PO | | 0.994989 | DISCARD |
| 2-year summary (Laboratory tests) - number of high values of Glucose^1H_post_50_g_glucose_PO | | 0.994989 | DISCARD |
| 2-year summary (Laboratory tests) - highest value of Glucose^1H_post_50_g_glucose_PO | | 0.994989 | DISCARD |
| 2-year summary (Laboratory tests) - number of low values of Glucose^1H_post_50_g_glucose_PO | | 0.994989 | DISCARD |
| 2-year summary (Laboratory tests) - lowest value of Glucose^1H_post_50_g_glucose_PO | | 0.994989 | DISCARD |
| 2-year summary (Laboratory tests) - average value of Glucose^1H_post_50_g_glucose_PO | | 0.994989 | DISCARD |
| 2-year summary (Laboratory tests) - median value of Glucose^1H_post_50_g_glucose_PO | | 0.994989 | DISCARD |
| 2-year summary (Laboratory tests) - count of normal tests for Glucose^1H_post_50_g_glucose_PO | | 0.994989 | DISCARD |
| 2-year summary (Drug prescriptions) - ANTIVIRAL AGENTS - INFLUENZA VIRUS SPECIFIC Days since first prescription | | 0.995017 | DISCARD |
| 2-year summary (Drug prescriptions) - ANTIVIRAL AGENTS - INFLUENZA VIRUS SPECIFIC Days since last prescription | | 0.995017 | DISCARD |
| 2-year summary (Drug prescriptions) - ANTIVIRAL AGENTS - INFLUENZA VIRUS SPECIFIC Prescription count | | 0.995017 | DISCARD |
| 2-year summary (Drug prescriptions) - UNCLASSIFIED THERAPEUTIC AGENTS Average frequency (days) | | 0.995017 | DISCARD |

| | | | |
|---|---|---|---|
| 2-year summary (Drug prescriptions) - UNCLASSIFIED THERAPEUTIC AGENTS Max frequency (days) | | 0.995017 | DISCARD |
| 2-year summary (Drug prescriptions) - UNCLASSIFIED THERAPEUTIC AGENTS Min frequency (days) | | 0.995017 | DISCARD |
| 1-year summary (Drug prescriptions) - BETA BLOCKING AGENTS Average frequency (days) | | 0.995044 | DISCARD |
| 1-year summary (Drug prescriptions) - BETA BLOCKING AGENTS Max frequency (days) | | 0.995044 | DISCARD |
| 1-year summary (Drug prescriptions) - BETA BLOCKING AGENTS Min frequency (days) | | 0.995044 | DISCARD |
| 2-year summary (Drug prescriptions) - VITAMIN K ANTAGONISTS Days since first prescription | | 0.995072 | DISCARD |
| 2-year summary (Drug prescriptions) - VITAMIN K ANTAGONISTS Days since last prescription | | 0.995072 | DISCARD |
| 2-year summary (Drug prescriptions) - VITAMIN K ANTAGONISTS Prescription count | | 0.995072 | DISCARD |
| 1-year summary (Drug prescriptions) - ESTROGENS, PLAIN Days since first prescription | | 0.995155 | DISCARD |
| 1-year summary (Drug prescriptions) - ESTROGENS, PLAIN Days since last prescription | | 0.995155 | DISCARD |
| 1-year summary (Drug prescriptions) - ESTROGENS, PLAIN Prescription count | | 0.995155 | DISCARD |
| 1-year summary (Drug prescriptions) - UNCLASSIFIED THERAPEUTIC AGENTS Days since first prescription | | 0.995155 | DISCARD |
| 1-year summary (Drug prescriptions) - UNCLASSIFIED THERAPEUTIC AGENTS Days since last prescription | | 0.995155 | DISCARD |
| 1-year summary (Drug prescriptions) - UNCLASSIFIED THERAPEUTIC AGENTS Prescription count | | 0.995155 | DISCARD |
| 1-year summary (Drug prescriptions) - ANTIBIOTICS Average frequency (days) | | 0.995210 | DISCARD |
| 1-year summary (Drug prescriptions) - ANTIBIOTICS Max frequency (days) | | 0.995210 | DISCARD |
| 1-year summary (Drug prescriptions) - ANTIBIOTICS Min frequency (days) | | 0.995210 | DISCARD |
| 2-year summary (Drug prescriptions) - ANTI-HERPESVIRUS INFECTION Days since first prescription | | 0.995210 | DISCARD |
| 2-year summary (Drug prescriptions) - ANTI-HERPESVIRUS INFECTION Days since last prescription | | 0.995210 | DISCARD |
| 2-year summary (Drug prescriptions) - ANTI-HERPESVIRUS INFECTION Prescription count | | 0.995210 | DISCARD |
| 2-year summary (Drug prescriptions) - CONTACT LENS SOLN, TEARS AND LUBRICANTS Average frequency (days) | | 0.995237 | DISCARD |
| 2-year summary (Drug prescriptions) - CONTACT LENS SOLN, TEARS AND LUBRICANTS Max frequency (days) | | 0.995237 | DISCARD |
| 2-year summary (Drug prescriptions) - CONTACT LENS SOLN, TEARS AND LUBRICANTS Min frequency (days) | | 0.995237 | DISCARD |
| 2-year summary (Drug prescriptions) - CORTICOSTEROIDS AND ANTI-INFECTIVE COMBINATIONS Average frequency (days) | | 0.995237 | DISCARD |
| 2-year summary (Drug prescriptions) - CORTICOSTEROIDS AND ANTI-INFECTIVE COMBINATIONS Max frequency (days) | | 0.995237 | DISCARD |
| 2-year summary (Drug prescriptions) - CORTICOSTEROIDS AND ANTI-INFECTIVE COMBINATIONS Min frequency (days) | | 0.995237 | DISCARD |
| 2-year summary (Drug prescriptions) - VASOPRESSORS Days since first prescription | | 0.995237 | DISCARD |
| 2-year summary (Drug prescriptions) - VASOPRESSORS Days since last prescription | | 0.995237 | DISCARD |
| 2-year summary (Drug prescriptions) - VASOPRESSORS Prescription count | | 0.995237 | DISCARD |
| 1-year summary (Doctor visits) - CLINICAL IMMUNOLOGY | | 0.995265 | DISCARD |
| 1-year summary (Drug prescriptions) - ALPHA ADRENERGIC BLOCKING AGENTS Average frequency (days) | | 0.995292 | DISCARD |
| 1-year summary (Drug prescriptions) - ALPHA ADRENERGIC BLOCKING AGENTS Max frequency (days) | | 0.995292 | DISCARD |
| 1-year summary (Drug prescriptions) - ALPHA ADRENERGIC BLOCKING AGENTS Min frequency (days) | | 0.995292 | DISCARD |
| 2-year summary (Drug prescriptions) - VITAMIN K ANTAGONISTS Average frequency (days) | | 0.995292 | DISCARD |
| 2-year summary (Drug prescriptions) - VITAMIN K ANTAGONISTS Max frequency (days) | | 0.995292 | DISCARD |
| 2-year summary (Drug prescriptions) - VITAMIN K ANTAGONISTS Min frequency (days) | | 0.995292 | DISCARD |
| 1-year summary (Drug prescriptions) - OTHER ANTI-INFLMMATORY AGENTS Days since first prescription | | 0.995347 | DISCARD |

| | | | |
|---|---|---|---|
| 1-year summary (Drug prescriptions) - OTHER ANTI-INFLMMATORY AGENTS Days since last prescription | | 0.995347 | DISCARD |
| 1-year summary (Drug prescriptions) - OTHER ANTI-INFLMMATORY AGENTS Prescription count | | 0.995347 | DISCARD |
| 1-year summary (Laboratory tests) - number of tests for Albumin | | 0.995347 | DISCARD |
| 1-year summary (Laboratory tests) - days since earliest test for Albumin | | 0.995347 | DISCARD |
| 1-year summary (Laboratory tests) - days since latest test for Albumin | | 0.995347 | DISCARD |
| 1-year summary (Laboratory tests) - number of high values of Albumin | | 0.995347 | DISCARD |
| 1-year summary (Laboratory tests) - highest value of Albumin | | 0.995347 | DISCARD |
| 1-year summary (Laboratory tests) - number of low values of Albumin | | 0.995347 | DISCARD |
| 1-year summary (Laboratory tests) - lowest value of Albumin | | 0.995347 | DISCARD |
| 1-year summary (Laboratory tests) - average value of Albumin | | 0.995347 | DISCARD |
| 1-year summary (Laboratory tests) - median value of Albumin | | 0.995347 | DISCARD |
| 1-year summary (Laboratory tests) - count of normal tests for Albumin | | 0.995347 | DISCARD |
| 2-year summary (Drug prescriptions) - ANTIMETABOLITES Days since first prescription | | 0.995347 | DISCARD |
| 2-year summary (Drug prescriptions) - ANTIMETABOLITES Days since last prescription | | 0.995347 | DISCARD |
| 2-year summary (Drug prescriptions) - ANTIMETABOLITES Prescription count | | 0.995347 | DISCARD |
| 2-year summary (Drug prescriptions) - VITAMIN D Days since first prescription | | 0.995347 | DISCARD |
| 2-year summary (Drug prescriptions) - VITAMIN D Days since last prescription | | 0.995347 | DISCARD |
| 2-year summary (Drug prescriptions) - VITAMIN D Prescription count | | 0.995347 | DISCARD |
| 1-year summary (Doctor visits) - NON MEDICAL PROFESSIONLS FOR IHF | | 0.995402 | DISCARD |
| 2-year summary (Drug prescriptions) - ESTROGENS, PLAIN Average frequency (days) | | 0.995402 | DISCARD |
| 2-year summary (Drug prescriptions) - ESTROGENS, PLAIN Max frequency (days) | | 0.995402 | DISCARD |
| 2-year summary (Drug prescriptions) - ESTROGENS, PLAIN Min frequency (days) | | 0.995402 | DISCARD |
| 2-year summary (Drug prescriptions) - OVER-ACTIVE BLADDER Days since first prescription | | 0.995430 | DISCARD |
| 2-year summary (Drug prescriptions) - OVER-ACTIVE BLADDER Days since last prescription | | 0.995430 | DISCARD |
| 2-year summary (Drug prescriptions) - OVER-ACTIVE BLADDER Prescription count | | 0.995430 | DISCARD |
| 2-year summary (Drug prescriptions) - NEEDLE/SYRINGE/LANCET Days since first prescription | | 0.995457 | DISCARD |
| 2-year summary (Drug prescriptions) - NEEDLE/SYRINGE/LANCET Days since last prescription | | 0.995457 | DISCARD |
| 2-year summary (Drug prescriptions) - NEEDLE/SYRINGE/LANCET Prescription count | | 0.995457 | DISCARD |
| 1-year summary (Doctor visits) - CARDIOTHORACIC SURGERY | | 0.995512 | DISCARD |
| 1-year summary (Drug prescriptions) - SELECTIVE SEROTONIN REUPTAKE INHIBITORS - OTHER Average frequency (days) | | 0.995540 | DISCARD |
| 1-year summary (Drug prescriptions) - SELECTIVE SEROTONIN REUPTAKE INHIBITORS - OTHER Max frequency (days) | | 0.995540 | DISCARD |
| 1-year summary (Drug prescriptions) - SELECTIVE SEROTONIN REUPTAKE INHIBITORS - OTHER Min frequency (days) | | 0.995540 | DISCARD |
| 2-year summary (Drug prescriptions) - SULFONAMIDES, TRIMETROPRIM AND COMBINATION Average frequency (days) | | 0.995540 | DISCARD |
| 2-year summary (Drug prescriptions) - SULFONAMIDES, TRIMETROPRIM AND COMBINATION Max frequency (days) | | 0.995540 | DISCARD |
| 2-year summary (Drug prescriptions) - SULFONAMIDES, TRIMETROPRIM AND COMBINATION Min frequency (days) | | 0.995540 | DISCARD |
| 1-year summary (Drug prescriptions) - UNCLASSIFIED THERAPEUTIC AGENTS Average frequency (days) | | 0.995568 | DISCARD |
| 1-year summary (Drug prescriptions) - UNCLASSIFIED THERAPEUTIC AGENTS Max frequency (days) | | 0.995568 | DISCARD |

| | | | |
|---|---|---|---|
| 1-year summary (Drug prescriptions) - UNCLASSIFIED THERAPEUTIC AGENTS Min frequency (days) | | 0.995568 | DISCARD |
| 2-year summary (Drug prescriptions) - CARBONIC ANHYDRASE INHIBITORS Days since first prescription | | 0.995568 | DISCARD |
| 2-year summary (Drug prescriptions) - CARBONIC ANHYDRASE INHIBITORS Days since last prescription | | 0.995568 | DISCARD |
| 2-year summary (Drug prescriptions) - CARBONIC ANHYDRASE INHIBITORS Prescription count | | 0.995568 | DISCARD |
| 2-year summary (Drug prescriptions) - ANTI-HEMORRHOIDAL AGENTS WITHOUT CORTICOSTEROIDS Days since first prescriptic | | 0.995595 | DISCARD |
| 2-year summary (Drug prescriptions) - ANTI-HEMORRHOIDAL AGENTS WITHOUT CORTICOSTEROIDS Days since last prescriptic | | 0.995595 | DISCARD |
| 2-year summary (Drug prescriptions) - ANTI-HEMORRHOIDAL AGENTS WITHOUT CORTICOSTEROIDS Prescription count | | 0.995595 | DISCARD |
| 2-year summary (Drug prescriptions) - NON-STEROIDAL ANTI-INFLAMMATORY Days since first prescription | | 0.995623 | DISCARD |
| 2-year summary (Drug prescriptions) - NON-STEROIDAL ANTI-INFLAMMATORY Days since last prescription | | 0.995623 | DISCARD |
| 2-year summary (Drug prescriptions) - NON-STEROIDAL ANTI-INFLAMMATORY Prescription count | | 0.995623 | DISCARD |
| 1-year summary (Drug prescriptions) - ANTIEMETICS AND ANTINAUSEANTS Days since first prescription | | 0.995678 | DISCARD |
| 1-year summary (Drug prescriptions) - ANTIEMETICS AND ANTINAUSEANTS Days since last prescription | | 0.995678 | DISCARD |
| 1-year summary (Drug prescriptions) - ANTIEMETICS AND ANTINAUSEANTS Prescription count | | 0.995678 | DISCARD |
| 2-year summary (Drug prescriptions) - ANTIBACTERIAL/ANTIPROTOZOAL Average frequency (days) | | 0.995678 | DISCARD |
| 2-year summary (Drug prescriptions) - ANTIBACTERIAL/ANTIPROTOZOAL Max frequency (days) | | 0.995678 | DISCARD |
| 2-year summary (Drug prescriptions) - ANTIBACTERIAL/ANTIPROTOZOAL Min frequency (days) | | 0.995678 | DISCARD |
| 2-year summary (Drug prescriptions) - ANTIFUNGALS Average frequency (days) | | 0.995705 | DISCARD |
| 2-year summary (Drug prescriptions) - ANTIFUNGALS Max frequency (days) | | 0.995705 | DISCARD |
| 2-year summary (Drug prescriptions) - ANTIFUNGALS Min frequency (days) | | 0.995705 | DISCARD |
| 2-year summary (Drug prescriptions) - C.N.S. STIMULANTS Average frequency (days) | | 0.995705 | DISCARD |
| 2-year summary (Drug prescriptions) - C.N.S. STIMULANTS Max frequency (days) | | 0.995705 | DISCARD |
| 2-year summary (Drug prescriptions) - C.N.S. STIMULANTS Min frequency (days) | | 0.995705 | DISCARD |
| 2-year summary (Drug prescriptions) - VITAMIN B COMPLEX Days since first prescription | | 0.995788 | DISCARD |
| 2-year summary (Drug prescriptions) - VITAMIN B COMPLEX Days since last prescription | | 0.995788 | DISCARD |
| 2-year summary (Drug prescriptions) - VITAMIN B COMPLEX Prescription count | | 0.995788 | DISCARD |
| 1-year summary (Hospitalizations) - Class - Diseases of the skin and subcutaneous tissue (L00-L99) | | 0.995870 | DISCARD |
| 1-year summary (Drug prescriptions) - VITAMIN K ANTAGONISTS Days since first prescription | | 0.995870 | DISCARD |
| 1-year summary (Drug prescriptions) - VITAMIN K ANTAGONISTS Days since last prescription | | 0.995870 | DISCARD |
| 1-year summary (Drug prescriptions) - VITAMIN K ANTAGONISTS Prescription count | | 0.995870 | DISCARD |
| 2-year summary (Drug prescriptions) - GLAUCOMA THERAPY: OTHER Days since first prescription | | 0.995870 | DISCARD |
| 2-year summary (Drug prescriptions) - GLAUCOMA THERAPY: OTHER Days since last prescription | | 0.995870 | DISCARD |
| 2-year summary (Drug prescriptions) - GLAUCOMA THERAPY: OTHER Prescription count | | 0.995870 | DISCARD |
| 2-year summary (Drug prescriptions) - TETRACYCLINES Average frequency (days) | | 0.995870 | DISCARD |
| 2-year summary (Drug prescriptions) - TETRACYCLINES Max frequency (days) | | 0.995870 | DISCARD |
| 2-year summary (Drug prescriptions) - TETRACYCLINES Min frequency (days) | | 0.995870 | DISCARD |
| 1-year summary (Drug prescriptions) - LINCOSAMIDES Days since first prescription | | 0.995898 | DISCARD |
| 1-year summary (Drug prescriptions) - LINCOSAMIDES Days since last prescription | | 0.995898 | DISCARD |

| | | | |
|---|---|---|---|
| 1-year summary (Drug prescriptions) - LINCOSAMIDES Prescription count | | 0.995898 | DISCARD |
| 1-year summary (Drug prescriptions) - HISTAMINE H2 RECEPTOR ANTAGONIST Average frequency (days) | | 0.995925 | DISCARD |
| 1-year summary (Drug prescriptions) - HISTAMINE H2 RECEPTOR ANTAGONIST Max frequency (days) | | 0.995925 | DISCARD |
| 1-year summary (Drug prescriptions) - HISTAMINE H2 RECEPTOR ANTAGONIST Min frequency (days) | | 0.995925 | DISCARD |
| 1-year summary (Drug prescriptions) - BONE CALCIUM REGULATOR Average frequency (days) | | 0.995953 | DISCARD |
| 1-year summary (Drug prescriptions) - BONE CALCIUM REGULATOR Max frequency (days) | | 0.995953 | DISCARD |
| 1-year summary (Drug prescriptions) - BONE CALCIUM REGULATOR Min frequency (days) | | 0.995953 | DISCARD |
| 2-year summary (Drug prescriptions) - VITAMIN D Average frequency (days) | | 0.996008 | DISCARD |
| 2-year summary (Drug prescriptions) - VITAMIN D Max frequency (days) | | 0.996008 | DISCARD |
| 2-year summary (Drug prescriptions) - VITAMIN D Min frequency (days) | | 0.996008 | DISCARD |
| 1-year summary (Drug prescriptions) - VITAMIN D Days since first prescription | | 0.996063 | DISCARD |
| 1-year summary (Drug prescriptions) - VITAMIN D Days since last prescription | | 0.996063 | DISCARD |
| 1-year summary (Drug prescriptions) - VITAMIN D Prescription count | | 0.996063 | DISCARD |
| 2-year summary (Drug prescriptions) - UNCLASSIFIED THERAPEUTIC AGENTS: OTHER Days since first prescription | | 0.996063 | DISCARD |
| 2-year summary (Drug prescriptions) - UNCLASSIFIED THERAPEUTIC AGENTS: OTHER Days since last prescription | | 0.996063 | DISCARD |
| 2-year summary (Drug prescriptions) - UNCLASSIFIED THERAPEUTIC AGENTS: OTHER Prescription count | | 0.996063 | DISCARD |
| 1-year summary (Doctor visits) - CHIROPODISTS | | 0.996146 | DISCARD |
| 2-year summary (Drug prescriptions) - GLAUCOMA THERAPY: OTHER Average frequency (days) | | 0.996146 | DISCARD |
| 2-year summary (Drug prescriptions) - GLAUCOMA THERAPY: OTHER Max frequency (days) | | 0.996146 | DISCARD |
| 2-year summary (Drug prescriptions) - GLAUCOMA THERAPY: OTHER Min frequency (days) | | 0.996146 | DISCARD |
| 2-year summary (Drug prescriptions) - ANTIEMETICS AND ANTINAUSEANTS Average frequency (days) | | 0.996173 | DISCARD |
| 2-year summary (Drug prescriptions) - ANTIEMETICS AND ANTINAUSEANTS Max frequency (days) | | 0.996173 | DISCARD |
| 2-year summary (Drug prescriptions) - ANTIEMETICS AND ANTINAUSEANTS Min frequency (days) | | 0.996173 | DISCARD |
| 2-year summary (Drug prescriptions) - OVER-ACTIVE BLADDER Average frequency (days) | | 0.996173 | DISCARD |
| 2-year summary (Drug prescriptions) - OVER-ACTIVE BLADDER Max frequency (days) | | 0.996173 | DISCARD |
| 2-year summary (Drug prescriptions) - OVER-ACTIVE BLADDER Min frequency (days) | | 0.996173 | DISCARD |
| 1-year summary (Drug prescriptions) - GLAUCOMA THERAPY: OTHER Days since first prescription | | 0.996201 | DISCARD |
| 1-year summary (Drug prescriptions) - GLAUCOMA THERAPY: OTHER Days since last prescription | | 0.996201 | DISCARD |
| 1-year summary (Drug prescriptions) - GLAUCOMA THERAPY: OTHER Prescription count | | 0.996201 | DISCARD |
| 1-year summary (Drug prescriptions) - VITAMIN K ANTAGONISTS Average frequency (days) | | 0.996256 | DISCARD |
| 1-year summary (Drug prescriptions) - VITAMIN K ANTAGONISTS Max frequency (days) | | 0.996256 | DISCARD |
| 1-year summary (Drug prescriptions) - VITAMIN K ANTAGONISTS Min frequency (days) | | 0.996256 | DISCARD |
| 2-year summary (Drug prescriptions) - PROGESTOGENS Days since first prescription | | 0.996283 | DISCARD |
| 2-year summary (Drug prescriptions) - PROGESTOGENS Days since last prescription | | 0.996283 | DISCARD |
| 2-year summary (Drug prescriptions) - PROGESTOGENS Prescription count | | 0.996283 | DISCARD |
| 2-year summary (Drug prescriptions) - ANTIDIARRHEA Days since first prescription | | 0.996311 | DISCARD |
| 2-year summary (Drug prescriptions) - ANTIDIARRHEA Days since last prescription | | 0.996311 | DISCARD |

| | | | |
|---|---|---|---|
| 2-year summary (Drug prescriptions) - ANTIDIARRHEA Prescription count | | 0.996311 | DISCARD |
| 1-year summary (Drug prescriptions) - ORAL CONTRACEPTIVES Days since first prescription | | 0.996366 | DISCARD |
| 1-year summary (Drug prescriptions) - ORAL CONTRACEPTIVES Days since last prescription | | 0.996366 | DISCARD |
| 1-year summary (Drug prescriptions) - ORAL CONTRACEPTIVES Prescription count | | 0.996366 | DISCARD |
| 2-year summary (Laboratory tests) - number of tests for Cholesterol.total/Cholesterol.in_HDL | | 0.996366 | DISCARD |
| 2-year summary (Laboratory tests) - days since earliest test for Cholesterol.total/Cholesterol.in_HDL | | 0.996366 | DISCARD |
| 2-year summary (Laboratory tests) - days since latest test for Cholesterol.total/Cholesterol.in_HDL | | 0.996366 | DISCARD |
| 2-year summary (Laboratory tests) - number of high values of Cholesterol.total/Cholesterol.in_HDL | | 0.996366 | DISCARD |
| 2-year summary (Laboratory tests) - highest value of Cholesterol.total/Cholesterol.in_HDL | | 0.996366 | DISCARD |
| 2-year summary (Laboratory tests) - number of low values of Cholesterol.total/Cholesterol.in_HDL | | 0.996366 | DISCARD |
| 2-year summary (Laboratory tests) - lowest value of Cholesterol.total/Cholesterol.in_HDL | | 0.996366 | DISCARD |
| 2-year summary (Laboratory tests) - average value of Cholesterol.total/Cholesterol.in_HDL | | 0.996366 | DISCARD |
| 2-year summary (Laboratory tests) - median value of Cholesterol.total/Cholesterol.in_HDL | | 0.996366 | DISCARD |
| 2-year summary (Laboratory tests) - count of normal tests for Cholesterol.total/Cholesterol.in_HDL | | 0.996366 | DISCARD |
| 1-year summary (Drug prescriptions) - ANTIVIRAL AGENTS Days since first prescription | | 0.996393 | DISCARD |
| 1-year summary (Drug prescriptions) - ANTIVIRAL AGENTS Days since last prescription | | 0.996393 | DISCARD |
| 1-year summary (Drug prescriptions) - ANTIVIRAL AGENTS Prescription count | | 0.996393 | DISCARD |
| 2-year summary (Laboratory tests) - number of tests for Glucose^2H_post_75_g_glucose_PO | | 0.996393 | DISCARD |
| 2-year summary (Laboratory tests) - days since earliest test for Glucose^2H_post_75_g_glucose_PO | | 0.996393 | DISCARD |
| 2-year summary (Laboratory tests) - days since latest test for Glucose^2H_post_75_g_glucose_PO | | 0.996393 | DISCARD |
| 2-year summary (Laboratory tests) - number of high values of Glucose^2H_post_75_g_glucose_PO | | 0.996393 | DISCARD |
| 2-year summary (Laboratory tests) - highest value of Glucose^2H_post_75_g_glucose_PO | | 0.996393 | DISCARD |
| 2-year summary (Laboratory tests) - number of low values of Glucose^2H_post_75_g_glucose_PO | | 0.996393 | DISCARD |
| 2-year summary (Laboratory tests) - lowest value of Glucose^2H_post_75_g_glucose_PO | | 0.996393 | DISCARD |
| 2-year summary (Laboratory tests) - average value of Glucose^2H_post_75_g_glucose_PO | | 0.996393 | DISCARD |
| 2-year summary (Laboratory tests) - median value of Glucose^2H_post_75_g_glucose_PO | | 0.996393 | DISCARD |
| 2-year summary (Laboratory tests) - count of normal tests for Glucose^2H_post_75_g_glucose_PO | | 0.996393 | DISCARD |
| 1-year summary (Doctor visits) - NURSE PRACTITIONERS | | 0.996421 | DISCARD |
| 2-year summary (Drug prescriptions) - VALPROIC ACID DERIVATIVES Days since first prescription | | 0.996449 | DISCARD |
| 2-year summary (Drug prescriptions) - VALPROIC ACID DERIVATIVES Days since last prescription | | 0.996449 | DISCARD |
| 2-year summary (Drug prescriptions) - VALPROIC ACID DERIVATIVES Prescription count | | 0.996449 | DISCARD |
| 1-year summary (Drug prescriptions) - GABA DERIVATIVES Average frequency (days) | | 0.996476 | DISCARD |
| 1-year summary (Drug prescriptions) - GABA DERIVATIVES Max frequency (days) | | 0.996476 | DISCARD |
| 1-year summary (Drug prescriptions) - GABA DERIVATIVES Min frequency (days) | | 0.996476 | DISCARD |
| 1-year summary (Drug prescriptions) - GLAUCOMA THERAPY: OTHER Average frequency (days) | | 0.996559 | DISCARD |
| 1-year summary (Drug prescriptions) - GLAUCOMA THERAPY: OTHER Max frequency (days) | | 0.996559 | DISCARD |
| 1-year summary (Drug prescriptions) - GLAUCOMA THERAPY: OTHER Min frequency (days) | | 0.996559 | DISCARD |

| | | |
|---|---|---|
| 2-year summary (Drug prescriptions) - OTHER ANTI-INFLMMATORY AGENTS Average frequency (days) | 0.996586 | DISCARD |
| 2-year summary (Drug prescriptions) - OTHER ANTI-INFLMMATORY AGENTS Max frequency (days) | 0.996586 | DISCARD |
| 2-year summary (Drug prescriptions) - OTHER ANTI-INFLMMATORY AGENTS Min frequency (days) | 0.996586 | DISCARD |
| 1-year summary (Drug prescriptions) - VITAMIN B COMPLEX Days since first prescription | 0.996641 | DISCARD |
| 1-year summary (Drug prescriptions) - VITAMIN B COMPLEX Days since last prescription | 0.996641 | DISCARD |
| 1-year summary (Drug prescriptions) - VITAMIN B COMPLEX Prescription count | 0.996641 | DISCARD |
| 1-year summary (Drug prescriptions) - VITAMIN D Average frequency (days) | 0.996641 | DISCARD |
| 1-year summary (Drug prescriptions) - VITAMIN D Max frequency (days) | 0.996641 | DISCARD |
| 1-year summary (Drug prescriptions) - VITAMIN D Min frequency (days) | 0.996641 | DISCARD |
| 2-year summary (Drug prescriptions) - CARBONIC ANHYDRASE INHIBITORS Average frequency (days) | 0.996641 | DISCARD |
| 2-year summary (Drug prescriptions) - CARBONIC ANHYDRASE INHIBITORS Max frequency (days) | 0.996641 | DISCARD |
| 2-year summary (Drug prescriptions) - CARBONIC ANHYDRASE INHIBITORS Min frequency (days) | 0.996641 | DISCARD |
| 2-year summary (Drug prescriptions) - MONOCLONAL ANTIBODIES Days since first prescription | 0.996641 | DISCARD |
| 2-year summary (Drug prescriptions) - MONOCLONAL ANTIBODIES Days since last prescription | 0.996641 | DISCARD |
| 2-year summary (Drug prescriptions) - MONOCLONAL ANTIBODIES Prescription count | 0.996641 | DISCARD |
| 2-year summary (Drug prescriptions) - VITAMIN B COMPLEX Average frequency (days) | 0.996641 | DISCARD |
| 2-year summary (Drug prescriptions) - VITAMIN B COMPLEX Max frequency (days) | 0.996641 | DISCARD |
| 2-year summary (Drug prescriptions) - VITAMIN B COMPLEX Min frequency (days) | 0.996641 | DISCARD |
| 1-year summary (Drug prescriptions) - REPLACEMENT AGENTS Days since first prescription | 0.996669 | DISCARD |
| 1-year summary (Drug prescriptions) - REPLACEMENT AGENTS Days since last prescription | 0.996669 | DISCARD |
| 1-year summary (Drug prescriptions) - REPLACEMENT AGENTS Prescription count | 0.996669 | DISCARD |
| 1-year summary (Drug prescriptions) - ANTIDIABETIC AGENTS: MISCELLANEOUS Days since first prescription | 0.996696 | DISCARD |
| 1-year summary (Drug prescriptions) - ANTIDIABETIC AGENTS: MISCELLANEOUS Days since last prescription | 0.996696 | DISCARD |
| 1-year summary (Drug prescriptions) - ANTIDIABETIC AGENTS: MISCELLANEOUS Prescription count | 0.996696 | DISCARD |
| 1-year summary (Drug prescriptions) - CARBONIC ANHYDRASE INHIBITORS Days since first prescription | 0.996696 | DISCARD |
| 1-year summary (Drug prescriptions) - CARBONIC ANHYDRASE INHIBITORS Days since last prescription | 0.996696 | DISCARD |
| 1-year summary (Drug prescriptions) - CARBONIC ANHYDRASE INHIBITORS Prescription count | 0.996696 | DISCARD |
| 1-year summary (Drug prescriptions) - VALPROIC ACID DERIVATIVES Days since first prescription | 0.996696 | DISCARD |
| 1-year summary (Drug prescriptions) - VALPROIC ACID DERIVATIVES Days since last prescription | 0.996696 | DISCARD |
| 1-year summary (Drug prescriptions) - VALPROIC ACID DERIVATIVES Prescription count | 0.996696 | DISCARD |
| 2-year summary (Hospitalizations) - Class - Unavailable | 0.996696 | DISCARD |
| 2-year summary (Drug prescriptions) - ANTIDIABETIC AGENTS: MISCELLANEOUS Days since first prescription | 0.996696 | DISCARD |
| 2-year summary (Drug prescriptions) - ANTIDIABETIC AGENTS: MISCELLANEOUS Days since last prescription | 0.996696 | DISCARD |
| 2-year summary (Drug prescriptions) - ANTIDIABETIC AGENTS: MISCELLANEOUS Prescription count | 0.996696 | DISCARD |
| 2-year summary (Doctor visits) - THORACIC SURGERY | 0.996696 | DISCARD |
| 1-year summary (Drug prescriptions) - ANTIMETABOLITES Days since first prescription | 0.996724 | DISCARD |
| 1-year summary (Drug prescriptions) - ANTIMETABOLITES Days since last prescription | 0.996724 | DISCARD |

| | | | |
|---|---|---|---|
| 1-year summary (Drug prescriptions) - ANTIMETABOLITES Prescription count | | 0.996724 | DISCARD |
| 1-year summary (Drug prescriptions) - CORONARY VASODILATORS Average frequency (days) | | 0.996724 | DISCARD |
| 1-year summary (Drug prescriptions) - CORONARY VASODILATORS Max frequency (days) | | 0.996724 | DISCARD |
| 1-year summary (Drug prescriptions) - CORONARY VASODILATORS Min frequency (days) | | 0.996724 | DISCARD |
| 1-year summary (Drug prescriptions) - ESTROGENS, PLAIN Average frequency (days) | | 0.996724 | DISCARD |
| 1-year summary (Drug prescriptions) - ESTROGENS, PLAIN Max frequency (days) | | 0.996724 | DISCARD |
| 1-year summary (Drug prescriptions) - ESTROGENS, PLAIN Min frequency (days) | | 0.996724 | DISCARD |
| 2-year summary (Drug prescriptions) - VALPROIC ACID DERIVATIVES Average frequency (days) | | 0.996724 | DISCARD |
| 2-year summary (Drug prescriptions) - VALPROIC ACID DERIVATIVES Max frequency (days) | | 0.996724 | DISCARD |
| 2-year summary (Drug prescriptions) - VALPROIC ACID DERIVATIVES Min frequency (days) | | 0.996724 | DISCARD |
| 1-year summary (Drug prescriptions) - OVER-ACTIVE BLADDER Days since first prescription | | 0.996751 | DISCARD |
| 1-year summary (Drug prescriptions) - OVER-ACTIVE BLADDER Days since last prescription | | 0.996751 | DISCARD |
| 1-year summary (Drug prescriptions) - OVER-ACTIVE BLADDER Prescription count | | 0.996751 | DISCARD |
| 2-year summary (Drug prescriptions) - ANTIVIRAL AGENTS Average frequency (days) | | 0.996806 | DISCARD |
| 2-year summary (Drug prescriptions) - ANTIVIRAL AGENTS Max frequency (days) | | 0.996806 | DISCARD |
| 2-year summary (Drug prescriptions) - ANTIVIRAL AGENTS Min frequency (days) | | 0.996806 | DISCARD |
| 2-year summary (Drug prescriptions) - ANTIVIRALS Days since first prescription | | 0.996834 | DISCARD |
| 2-year summary (Drug prescriptions) - ANTIVIRALS Days since last prescription | | 0.996834 | DISCARD |
| 2-year summary (Drug prescriptions) - ANTIVIRALS Prescription count | | 0.996834 | DISCARD |
| 1-year summary (Drug prescriptions) - VALPROIC ACID DERIVATIVES Average frequency (days) | | 0.996972 | DISCARD |
| 1-year summary (Drug prescriptions) - VALPROIC ACID DERIVATIVES Max frequency (days) | | 0.996972 | DISCARD |
| 1-year summary (Drug prescriptions) - VALPROIC ACID DERIVATIVES Min frequency (days) | | 0.996972 | DISCARD |
| 2-year summary (Drug prescriptions) - VASODILATOR ANTIHYPERTENSIVE DRUGS Days since first prescription | | 0.996972 | DISCARD |
| 2-year summary (Drug prescriptions) - VASODILATOR ANTIHYPERTENSIVE DRUGS Days since last prescription | | 0.996972 | DISCARD |
| 2-year summary (Drug prescriptions) - VASODILATOR ANTIHYPERTENSIVE DRUGS Prescription count | | 0.996972 | DISCARD |
| 2-year summary (Drug prescriptions) - IMMUNOSUPPRESSIVE AGENTS Days since first prescription | | 0.996999 | DISCARD |
| 2-year summary (Drug prescriptions) - IMMUNOSUPPRESSIVE AGENTS Days since last prescription | | 0.996999 | DISCARD |
| 2-year summary (Drug prescriptions) - IMMUNOSUPPRESSIVE AGENTS Prescription count | | 0.996999 | DISCARD |
| 2-year summary (Drug prescriptions) - ANTIMETABOLITES Average frequency (days) | | 0.997082 | DISCARD |
| 2-year summary (Drug prescriptions) - ANTIMETABOLITES Max frequency (days) | | 0.997082 | DISCARD |
| 2-year summary (Drug prescriptions) - ANTIMETABOLITES Min frequency (days) | | 0.997082 | DISCARD |
| 2-year summary (Drug prescriptions) - CORTICOSTEROIDS WITH OTHER COMBINATIONS Days since first prescription | | 0.997082 | DISCARD |
| 2-year summary (Drug prescriptions) - CORTICOSTEROIDS WITH OTHER COMBINATIONS Days since last prescription | | 0.997082 | DISCARD |
| 2-year summary (Drug prescriptions) - CORTICOSTEROIDS WITH OTHER COMBINATIONS Prescription count | | 0.997082 | DISCARD |
| 2-year summary (Drug prescriptions) - MONOCLONAL ANTIBODIES Average frequency (days) | | 0.997109 | DISCARD |
| 2-year summary (Drug prescriptions) - MONOCLONAL ANTIBODIES Max frequency (days) | | 0.997109 | DISCARD |
| 2-year summary (Drug prescriptions) - MONOCLONAL ANTIBODIES Min frequency (days) | | 0.997109 | DISCARD |

| | | | |
|---|---|---|---|
| 1-year summary (Drug prescriptions) - ANTI-HERPESVIRUS INFECTION Days since first prescription | | 0.997137 | DISCARD |
| 1-year summary (Drug prescriptions) - ANTI-HERPESVIRUS INFECTION Days since last prescription | | 0.997137 | DISCARD |
| 1-year summary (Drug prescriptions) - ANTI-HERPESVIRUS INFECTION Prescription count | | 0.997137 | DISCARD |
| 1-year summary (Drug prescriptions) - CONTACT LENS SOLN, TEARS AND LUBRICANTS Average frequency (days) | | 0.997137 | DISCARD |
| 1-year summary (Drug prescriptions) - CONTACT LENS SOLN, TEARS AND LUBRICANTS Max frequency (days) | | 0.997137 | DISCARD |
| 1-year summary (Drug prescriptions) - CONTACT LENS SOLN, TEARS AND LUBRICANTS Min frequency (days) | | 0.997137 | DISCARD |
| 2-year summary (Drug prescriptions) - IMMUNOSUPPRESSIVE AGENTS Average frequency (days) | | 0.997164 | DISCARD |
| 2-year summary (Drug prescriptions) - IMMUNOSUPPRESSIVE AGENTS Max frequency (days) | | 0.997164 | DISCARD |
| 2-year summary (Drug prescriptions) - IMMUNOSUPPRESSIVE AGENTS Min frequency (days) | | 0.997164 | DISCARD |
| 2-year summary (Laboratory tests) - number of tests for Prostate_specific_Ag.free/Prostate_specific_Ag.total | | 0.997164 | DISCARD |
| 2-year summary (Laboratory tests) - days since earliest test for Prostate_specific_Ag.free/Prostate_specific_Ag.total | | 0.997164 | DISCARD |
| 2-year summary (Laboratory tests) - days since latest test for Prostate_specific_Ag.free/Prostate_specific_Ag.total | | 0.997164 | DISCARD |
| 2-year summary (Laboratory tests) - number of high values of Prostate_specific_Ag.free/Prostate_specific_Ag.total | | 0.997164 | DISCARD |
| 2-year summary (Laboratory tests) - highest value of Prostate_specific_Ag.free/Prostate_specific_Ag.total | | 0.997164 | DISCARD |
| 2-year summary (Laboratory tests) - number of low values of Prostate_specific_Ag.free/Prostate_specific_Ag.total | | 0.997164 | DISCARD |
| 2-year summary (Laboratory tests) - lowest value of Prostate_specific_Ag.free/Prostate_specific_Ag.total | | 0.997164 | DISCARD |
| 2-year summary (Laboratory tests) - average value of Prostate_specific_Ag.free/Prostate_specific_Ag.total | | 0.997164 | DISCARD |
| 2-year summary (Laboratory tests) - median value of Prostate_specific_Ag.free/Prostate_specific_Ag.total | | 0.997164 | DISCARD |
| 2-year summary (Laboratory tests) - count of normal tests for Prostate_specific_Ag.free/Prostate_specific_Ag.total | | 0.997164 | DISCARD |
| 1-year summary (Hospitalizations) - Class - Unavailable | | 0.997192 | DISCARD |
| 1-year summary (Drug prescriptions) - C.N.S. STIMULANTS Days since first prescription | | 0.997192 | DISCARD |
| 1-year summary (Drug prescriptions) - C.N.S. STIMULANTS Days since last prescription | | 0.997192 | DISCARD |
| 1-year summary (Drug prescriptions) - C.N.S. STIMULANTS Prescription count | | 0.997192 | DISCARD |
| 2-year summary (Drug prescriptions) - VASODILATOR ANTIHYPERTENSIVE DRUGS Average frequency (days) | | 0.997192 | DISCARD |
| 2-year summary (Drug prescriptions) - VASODILATOR ANTIHYPERTENSIVE DRUGS Max frequency (days) | | 0.997192 | DISCARD |
| 2-year summary (Drug prescriptions) - VASODILATOR ANTIHYPERTENSIVE DRUGS Min frequency (days) | | 0.997192 | DISCARD |
| 1-year summary (Drug prescriptions) - VITAMIN B COMPLEX Average frequency (days) | | 0.997219 | DISCARD |
| 1-year summary (Drug prescriptions) - VITAMIN B COMPLEX Max frequency (days) | | 0.997219 | DISCARD |
| 1-year summary (Drug prescriptions) - VITAMIN B COMPLEX Min frequency (days) | | 0.997219 | DISCARD |
| 1-year summary (Drug prescriptions) - ANTIDIARRHEA Days since first prescription | | 0.997274 | DISCARD |
| 1-year summary (Drug prescriptions) - ANTIDIARRHEA Days since last prescription | | 0.997274 | DISCARD |
| 1-year summary (Drug prescriptions) - ANTIDIARRHEA Prescription count | | 0.997274 | DISCARD |
| 2-year summary (Drug prescriptions) - OPIOID DEPENDENCE Days since first prescription | | 0.997274 | DISCARD |
| 2-year summary (Drug prescriptions) - OPIOID DEPENDENCE Days since last prescription | | 0.997274 | DISCARD |
| 2-year summary (Drug prescriptions) - OPIOID DEPENDENCE Prescription count | | 0.997274 | DISCARD |
| 1-year summary (Drug prescriptions) - VASODILATOR ANTIHYPERTENSIVE DRUGS Days since first prescription | | 0.997302 | DISCARD |
| 1-year summary (Drug prescriptions) - VASODILATOR ANTIHYPERTENSIVE DRUGS Days since last prescription | | 0.997302 | DISCARD |

| | | |
|---|---|---|
| 1-year summary (Drug prescriptions) - VASODILATOR ANTIHYPERTENSIVE DRUGS Prescription count | 0.997302 | DISCARD |
| 1-year summary (Drug prescriptions) - IMMUNOSUPPRESSIVE AGENTS Days since first prescription | 0.997330 | DISCARD |
| 1-year summary (Drug prescriptions) - IMMUNOSUPPRESSIVE AGENTS Days since last prescription | 0.997330 | DISCARD |
| 1-year summary (Drug prescriptions) - IMMUNOSUPPRESSIVE AGENTS Prescription count | 0.997330 | DISCARD |
| 1-year summary (Drug prescriptions) - OVER-ACTIVE BLADDER Average frequency (days) | 0.997330 | DISCARD |
| 1-year summary (Drug prescriptions) - OVER-ACTIVE BLADDER Max frequency (days) | 0.997330 | DISCARD |
| 1-year summary (Drug prescriptions) - OVER-ACTIVE BLADDER Min frequency (days) | 0.997330 | DISCARD |
| 1-year summary (Drug prescriptions) - CARBONIC ANHYDRASE INHIBITORS Average frequency (days) | 0.997357 | DISCARD |
| 1-year summary (Drug prescriptions) - CARBONIC ANHYDRASE INHIBITORS Max frequency (days) | 0.997357 | DISCARD |
| 1-year summary (Drug prescriptions) - CARBONIC ANHYDRASE INHIBITORS Min frequency (days) | 0.997357 | DISCARD |
| 1-year summary (Drug prescriptions) - VASOPRESSORS Days since first prescription | 0.997357 | DISCARD |
| 1-year summary (Drug prescriptions) - VASOPRESSORS Days since last prescription | 0.997357 | DISCARD |
| 1-year summary (Drug prescriptions) - VASOPRESSORS Prescription count | 0.997357 | DISCARD |
| 2-year summary (Drug prescriptions) - OPIOID DEPENDENCE Average frequency (days) | 0.997385 | DISCARD |
| 2-year summary (Drug prescriptions) - OPIOID DEPENDENCE Max frequency (days) | 0.997385 | DISCARD |
| 2-year summary (Drug prescriptions) - OPIOID DEPENDENCE Min frequency (days) | 0.997385 | DISCARD |
| 2-year summary (Drug prescriptions) - TOPICAL IMMUNOSUPRESSIVE AGENTS Days since first prescription | 0.997385 | DISCARD |
| 2-year summary (Drug prescriptions) - TOPICAL IMMUNOSUPRESSIVE AGENTS Days since last prescription | 0.997385 | DISCARD |
| 2-year summary (Drug prescriptions) - TOPICAL IMMUNOSUPRESSIVE AGENTS Prescription count | 0.997385 | DISCARD |
| 1-year summary (Drug prescriptions) - ANTIDIABETIC AGENTS: MISCELLANEOUS Average frequency (days) | 0.997412 | DISCARD |
| 1-year summary (Drug prescriptions) - ANTIDIABETIC AGENTS: MISCELLANEOUS Max frequency (days) | 0.997412 | DISCARD |
| 1-year summary (Drug prescriptions) - ANTIDIABETIC AGENTS: MISCELLANEOUS Min frequency (days) | 0.997412 | DISCARD |
| 1-year summary (Drug prescriptions) - MONOCLONAL ANTIBODIES Days since first prescription | 0.997412 | DISCARD |
| 1-year summary (Drug prescriptions) - MONOCLONAL ANTIBODIES Days since last prescription | 0.997412 | DISCARD |
| 1-year summary (Drug prescriptions) - MONOCLONAL ANTIBODIES Prescription count | 0.997412 | DISCARD |
| 2-year summary (Drug prescriptions) - ANTIDIABETIC AGENTS: MISCELLANEOUS Average frequency (days) | 0.997412 | DISCARD |
| 2-year summary (Drug prescriptions) - ANTIDIABETIC AGENTS: MISCELLANEOUS Max frequency (days) | 0.997412 | DISCARD |
| 2-year summary (Drug prescriptions) - ANTIDIABETIC AGENTS: MISCELLANEOUS Min frequency (days) | 0.997412 | DISCARD |
| 1-year summary (Drug prescriptions) - ANTIEMETICS AND ANTINAUSEANTS Average frequency (days) | 0.997440 | DISCARD |
| 1-year summary (Drug prescriptions) - ANTIEMETICS AND ANTINAUSEANTS Max frequency (days) | 0.997440 | DISCARD |
| 1-year summary (Drug prescriptions) - ANTIEMETICS AND ANTINAUSEANTS Min frequency (days) | 0.997440 | DISCARD |
| 1-year summary (Drug prescriptions) - ANTIVIRAL AGENTS - INFLUENZA VIRUS SPECIFIC Days since first prescription | 0.997440 | DISCARD |
| 1-year summary (Drug prescriptions) - ANTIVIRAL AGENTS - INFLUENZA VIRUS SPECIFIC Days since last prescription | 0.997440 | DISCARD |
| 1-year summary (Drug prescriptions) - ANTIVIRAL AGENTS - INFLUENZA VIRUS SPECIFIC Prescription count | 0.997440 | DISCARD |
| 2-year summary (Drug prescriptions) - MISCELLANEOUS SKIN AND MUCOUS MEMBRANE AGENTS Days since first prescription | 0.997467 | DISCARD |
| 2-year summary (Drug prescriptions) - MISCELLANEOUS SKIN AND MUCOUS MEMBRANE AGENTS Days since last prescription | 0.997467 | DISCARD |
| 2-year summary (Drug prescriptions) - MISCELLANEOUS SKIN AND MUCOUS MEMBRANE AGENTS Prescription count | 0.997467 | DISCARD |

| | | | |
|---|---|---|---|
| 2-year summary (Drug prescriptions) - MULTIVITAMIN PREPARATIONS Days since first prescription | | 0.997467 | DISCARD |
| 2-year summary (Drug prescriptions) - MULTIVITAMIN PREPARATIONS Days since last prescription | | 0.997467 | DISCARD |
| 2-year summary (Drug prescriptions) - MULTIVITAMIN PREPARATIONS Prescription count | | 0.997467 | DISCARD |
| 1-year summary (Drug prescriptions) - NEEDLE/SYRINGE/LANCET Days since first prescription | | 0.997495 | DISCARD |
| 1-year summary (Drug prescriptions) - NEEDLE/SYRINGE/LANCET Days since last prescription | | 0.997495 | DISCARD |
| 1-year summary (Drug prescriptions) - NEEDLE/SYRINGE/LANCET Prescription count | | 0.997495 | DISCARD |
| 1-year summary (Drug prescriptions) - SULFONAMIDES, TRIMETROPRIM AND COMBINATION Average frequency (days) | | 0.997522 | DISCARD |
| 1-year summary (Drug prescriptions) - SULFONAMIDES, TRIMETROPRIM AND COMBINATION Max frequency (days) | | 0.997522 | DISCARD |
| 1-year summary (Drug prescriptions) - SULFONAMIDES, TRIMETROPRIM AND COMBINATION Min frequency (days) | | 0.997522 | DISCARD |
| 2-year summary (Doctor visits) - GENETICS | | 0.997522 | DISCARD |
| 1-year summary (Drug prescriptions) - ANTI-HEMORRHOIDAL AGENTS WITHOUT CORTICOSTEROIDS Days since first prescriptic | | 0.997605 | DISCARD |
| 1-year summary (Drug prescriptions) - ANTI-HEMORRHOIDAL AGENTS WITHOUT CORTICOSTEROIDS Days since last prescriptic | | 0.997605 | DISCARD |
| 1-year summary (Drug prescriptions) - ANTI-HEMORRHOIDAL AGENTS WITHOUT CORTICOSTEROIDS Prescription count | | 0.997605 | DISCARD |
| 1-year summary (Drug prescriptions) - IMMUNOSUPPRESSIVE AGENTS Average frequency (days) | | 0.997632 | DISCARD |
| 1-year summary (Drug prescriptions) - IMMUNOSUPPRESSIVE AGENTS Max frequency (days) | | 0.997632 | DISCARD |
| 1-year summary (Drug prescriptions) - IMMUNOSUPPRESSIVE AGENTS Min frequency (days) | | 0.997632 | DISCARD |
| 1-year summary (Drug prescriptions) - ANTIMETABOLITES Average frequency (days) | | 0.997660 | DISCARD |
| 1-year summary (Drug prescriptions) - ANTIMETABOLITES Max frequency (days) | | 0.997660 | DISCARD |
| 1-year summary (Drug prescriptions) - ANTIMETABOLITES Min frequency (days) | | 0.997660 | DISCARD |
| 2-year summary (Drug prescriptions) - MULTIVITAMIN PREPARATIONS Average frequency (days) | | 0.997660 | DISCARD |
| 2-year summary (Drug prescriptions) - MULTIVITAMIN PREPARATIONS Max frequency (days) | | 0.997660 | DISCARD |
| 2-year summary (Drug prescriptions) - MULTIVITAMIN PREPARATIONS Min frequency (days) | | 0.997660 | DISCARD |
| 1-year summary (Drug prescriptions) - VASODILATOR ANTIHYPERTENSIVE DRUGS Average frequency (days) | | 0.997687 | DISCARD |
| 1-year summary (Drug prescriptions) - VASODILATOR ANTIHYPERTENSIVE DRUGS Max frequency (days) | | 0.997687 | DISCARD |
| 1-year summary (Drug prescriptions) - VASODILATOR ANTIHYPERTENSIVE DRUGS Min frequency (days) | | 0.997687 | DISCARD |
| 2-year summary (Drug prescriptions) - NUTRITIONAL PRODUCTS Days since first prescription | | 0.997687 | DISCARD |
| 2-year summary (Drug prescriptions) - NUTRITIONAL PRODUCTS Days since last prescription | | 0.997687 | DISCARD |
| 2-year summary (Drug prescriptions) - NUTRITIONAL PRODUCTS Prescription count | | 0.997687 | DISCARD |
| 2-year summary (Drug prescriptions) - OTHER HORMONAL CONTRACEPTIVES Days since first prescription | | 0.997687 | DISCARD |
| 2-year summary (Drug prescriptions) - OTHER HORMONAL CONTRACEPTIVES Days since last prescription | | 0.997687 | DISCARD |
| 2-year summary (Drug prescriptions) - OTHER HORMONAL CONTRACEPTIVES Prescription count | | 0.997687 | DISCARD |
| 1-year summary (Drug prescriptions) - MULTIVITAMIN PREPARATIONS Days since first prescription | | 0.997715 | DISCARD |
| 1-year summary (Drug prescriptions) - MULTIVITAMIN PREPARATIONS Days since last prescription | | 0.997715 | DISCARD |
| 1-year summary (Drug prescriptions) - MULTIVITAMIN PREPARATIONS Prescription count | | 0.997715 | DISCARD |
| 1-year summary (Drug prescriptions) - ORAL CONTRACEPTIVES Average frequency (days) | | 0.997715 | DISCARD |
| 1-year summary (Drug prescriptions) - ORAL CONTRACEPTIVES Max frequency (days) | | 0.997715 | DISCARD |
| 1-year summary (Drug prescriptions) - ORAL CONTRACEPTIVES Min frequency (days) | | 0.997715 | DISCARD |

| | | | |
|---|---|---|---|
| 2-year summary (Drug prescriptions) - ANTIVIRAL AGENTS - INFLUENZA VIRUS SPECIFIC Average frequency (days) | | 0.997715 | DISCARD |
| 2-year summary (Drug prescriptions) - ANTIVIRAL AGENTS - INFLUENZA VIRUS SPECIFIC Max frequency (days) | | 0.997715 | DISCARD |
| 2-year summary (Drug prescriptions) - ANTIVIRAL AGENTS - INFLUENZA VIRUS SPECIFIC Min frequency (days) | | 0.997715 | DISCARD |
| 2-year summary (Drug prescriptions) - CARBOXAMIDE DERIVATIVES Days since first prescription | | 0.997715 | DISCARD |
| 2-year summary (Drug prescriptions) - CARBOXAMIDE DERIVATIVES Days since last prescription | | 0.997715 | DISCARD |
| 2-year summary (Drug prescriptions) - CARBOXAMIDE DERIVATIVES Prescription count | | 0.997715 | DISCARD |
| 1-year summary (Drug prescriptions) - C.N.S. STIMULANTS Average frequency (days) | | 0.997742 | DISCARD |
| 1-year summary (Drug prescriptions) - C.N.S. STIMULANTS Max frequency (days) | | 0.997742 | DISCARD |
| 1-year summary (Drug prescriptions) - C.N.S. STIMULANTS Min frequency (days) | | 0.997742 | DISCARD |
| 1-year summary (Drug prescriptions) - OTHER ANTI-INFLMMATORY AGENTS Average frequency (days) | | 0.997742 | DISCARD |
| 1-year summary (Drug prescriptions) - OTHER ANTI-INFLMMATORY AGENTS Max frequency (days) | | 0.997742 | DISCARD |
| 1-year summary (Drug prescriptions) - OTHER ANTI-INFLMMATORY AGENTS Min frequency (days) | | 0.997742 | DISCARD |
| 2-year summary (Drug prescriptions) - PROGESTOGENS Average frequency (days) | | 0.997742 | DISCARD |
| 2-year summary (Drug prescriptions) - PROGESTOGENS Max frequency (days) | | 0.997742 | DISCARD |
| 2-year summary (Drug prescriptions) - PROGESTOGENS Min frequency (days) | | 0.997742 | DISCARD |
| 1-year summary (Drug prescriptions) - ANTIVIRAL AGENTS Average frequency (days) | | 0.997770 | DISCARD |
| 1-year summary (Drug prescriptions) - ANTIVIRAL AGENTS Max frequency (days) | | 0.997770 | DISCARD |
| 1-year summary (Drug prescriptions) - ANTIVIRAL AGENTS Min frequency (days) | | 0.997770 | DISCARD |
| 1-year summary (Doctor visits) - THORACIC SURGERY | | 0.997770 | DISCARD |
| 2-year summary (Drug prescriptions) - LEVODOPA AND COMBINATION Days since first prescription | | 0.997770 | DISCARD |
| 2-year summary (Drug prescriptions) - LEVODOPA AND COMBINATION Days since last prescription | | 0.997770 | DISCARD |
| 2-year summary (Drug prescriptions) - LEVODOPA AND COMBINATION Prescription count | | 0.997770 | DISCARD |
| 1-year summary (Drug prescriptions) - MONOCLONAL ANTIBODIES Average frequency (days) | | 0.997798 | DISCARD |
| 1-year summary (Drug prescriptions) - MONOCLONAL ANTIBODIES Max frequency (days) | | 0.997798 | DISCARD |
| 1-year summary (Drug prescriptions) - MONOCLONAL ANTIBODIES Min frequency (days) | | 0.997798 | DISCARD |
| 2-year summary (Drug prescriptions) - DIGITALIS PREPARATIONS Days since first prescription | | 0.997798 | DISCARD |
| 2-year summary (Drug prescriptions) - DIGITALIS PREPARATIONS Days since last prescription | | 0.997798 | DISCARD |
| 2-year summary (Drug prescriptions) - DIGITALIS PREPARATIONS Prescription count | | 0.997798 | DISCARD |
| 2-year summary (Drug prescriptions) - PLASMODICIDES (ANTIMALARIALS) Days since first prescription | | 0.997798 | DISCARD |
| 2-year summary (Drug prescriptions) - PLASMODICIDES (ANTIMALARIALS) Days since last prescription | | 0.997798 | DISCARD |
| 2-year summary (Drug prescriptions) - PLASMODICIDES (ANTIMALARIALS) Prescription count | | 0.997798 | DISCARD |
| 2-year summary (Drug prescriptions) - ANTILIPEMIC: FIBRATES Days since first prescription | | 0.997825 | DISCARD |
| 2-year summary (Drug prescriptions) - ANTILIPEMIC: FIBRATES Days since last prescription | | 0.997825 | DISCARD |
| 2-year summary (Drug prescriptions) - ANTILIPEMIC: FIBRATES Prescription count | | 0.997825 | DISCARD |
| 1-year summary (Drug prescriptions) - UNCLASSIFIED THERAPEUTIC AGENTS: OTHER Days since first prescription | | 0.997853 | DISCARD |
| 1-year summary (Drug prescriptions) - UNCLASSIFIED THERAPEUTIC AGENTS: OTHER Days since last prescription | | 0.997853 | DISCARD |
| 1-year summary (Drug prescriptions) - UNCLASSIFIED THERAPEUTIC AGENTS: OTHER Prescription count | | 0.997853 | DISCARD |

| | | |
|---|---|---|
| 2-year summary (Drug prescriptions) - LEVODOPA AND COMBINATION Average frequency (days) | 0.997880 | DISCARD |
| 2-year summary (Drug prescriptions) - LEVODOPA AND COMBINATION Max frequency (days) | 0.997880 | DISCARD |
| 2-year summary (Drug prescriptions) - LEVODOPA AND COMBINATION Min frequency (days) | 0.997880 | DISCARD |
| 1-year summary (Drug prescriptions) - LEVODOPA AND COMBINATION Days since first prescription | 0.997908 | DISCARD |
| 1-year summary (Drug prescriptions) - LEVODOPA AND COMBINATION Days since last prescription | 0.997908 | DISCARD |
| 1-year summary (Drug prescriptions) - LEVODOPA AND COMBINATION Prescription count | 0.997908 | DISCARD |
| 1-year summary (Drug prescriptions) - MULTIVITAMIN PREPARATIONS Average frequency (days) | 0.997908 | DISCARD |
| 1-year summary (Drug prescriptions) - MULTIVITAMIN PREPARATIONS Max frequency (days) | 0.997908 | DISCARD |
| 1-year summary (Drug prescriptions) - MULTIVITAMIN PREPARATIONS Min frequency (days) | 0.997908 | DISCARD |
| 2-year summary (Drug prescriptions) - ANTIDIARRHEA Average frequency (days) | 0.997908 | DISCARD |
| 2-year summary (Drug prescriptions) - ANTIDIARRHEA Max frequency (days) | 0.997908 | DISCARD |
| 2-year summary (Drug prescriptions) - ANTIDIARRHEA Min frequency (days) | 0.997908 | DISCARD |
| 2-year summary (Drug prescriptions) - CARBOXAMIDE DERIVATIVES Average frequency (days) | 0.997908 | DISCARD |
| 2-year summary (Drug prescriptions) - CARBOXAMIDE DERIVATIVES Max frequency (days) | 0.997908 | DISCARD |
| 2-year summary (Drug prescriptions) - CARBOXAMIDE DERIVATIVES Min frequency (days) | 0.997908 | DISCARD |
| 2-year summary (Drug prescriptions) - DIGITALIS PREPARATIONS Average frequency (days) | 0.997908 | DISCARD |
| 2-year summary (Drug prescriptions) - DIGITALIS PREPARATIONS Max frequency (days) | 0.997908 | DISCARD |
| 2-year summary (Drug prescriptions) - DIGITALIS PREPARATIONS Min frequency (days) | 0.997908 | DISCARD |
| 2-year summary (Drug prescriptions) - NUTRITIONAL PRODUCTS Average frequency (days) | 0.997908 | DISCARD |
| 2-year summary (Drug prescriptions) - NUTRITIONAL PRODUCTS Max frequency (days) | 0.997908 | DISCARD |
| 2-year summary (Drug prescriptions) - NUTRITIONAL PRODUCTS Min frequency (days) | 0.997908 | DISCARD |
| 2-year summary (Drug prescriptions) - OPHTHALMOLOGICALS Days since first prescription | 0.997908 | DISCARD |
| 2-year summary (Drug prescriptions) - OPHTHALMOLOGICALS Days since last prescription | 0.997908 | DISCARD |
| 2-year summary (Drug prescriptions) - OPHTHALMOLOGICALS Prescription count | 0.997908 | DISCARD |
| 2-year summary (Drug prescriptions) - CONTRACEPTIVES Days since first prescription | 0.997935 | DISCARD |
| 2-year summary (Drug prescriptions) - CONTRACEPTIVES Days since last prescription | 0.997935 | DISCARD |
| 2-year summary (Drug prescriptions) - CONTRACEPTIVES Prescription count | 0.997935 | DISCARD |
| 2-year summary (Drug prescriptions) - NEEDLE/SYRINGE/LANCET Average frequency (days) | 0.997935 | DISCARD |
| 2-year summary (Drug prescriptions) - NEEDLE/SYRINGE/LANCET Max frequency (days) | 0.997935 | DISCARD |
| 2-year summary (Drug prescriptions) - NEEDLE/SYRINGE/LANCET Min frequency (days) | 0.997935 | DISCARD |
| 2-year summary (Drug prescriptions) - NON-STEROIDAL ANTI-INFLAMMATORY Average frequency (days) | 0.997935 | DISCARD |
| 2-year summary (Drug prescriptions) - NON-STEROIDAL ANTI-INFLAMMATORY Max frequency (days) | 0.997935 | DISCARD |
| 2-year summary (Drug prescriptions) - NON-STEROIDAL ANTI-INFLAMMATORY Min frequency (days) | 0.997935 | DISCARD |
| 1-year summary (Drug prescriptions) - OPIOID DEPENDENCE Days since first prescription | 0.997963 | DISCARD |
| 1-year summary (Drug prescriptions) - OPIOID DEPENDENCE Days since last prescription | 0.997963 | DISCARD |
| 1-year summary (Drug prescriptions) - OPIOID DEPENDENCE Prescription count | 0.997963 | DISCARD |
| 2-year summary (Drug prescriptions) - UNCLASSIFIED THERAPEUTIC AGENTS: OTHER Average frequency (days) | 0.997963 | DISCARD |

| | | | |
|---|---|---|---|
| 2-year summary (Drug prescriptions) - UNCLASSIFIED THERAPEUTIC AGENTS: OTHER Max frequency (days) | | 0.997963 | DISCARD |
| 2-year summary (Drug prescriptions) - UNCLASSIFIED THERAPEUTIC AGENTS: OTHER Min frequency (days) | | 0.997963 | DISCARD |
| 1-year summary (Drug prescriptions) - NUTRITIONAL PRODUCTS Days since first prescription | | 0.997990 | DISCARD |
| 1-year summary (Drug prescriptions) - NUTRITIONAL PRODUCTS Days since last prescription | | 0.997990 | DISCARD |
| 1-year summary (Drug prescriptions) - NUTRITIONAL PRODUCTS Prescription count | | 0.997990 | DISCARD |
| 2-year summary (Drug prescriptions) - ANTICHOLINERGIC AGENTS Days since first prescription | | 0.997990 | DISCARD |
| 2-year summary (Drug prescriptions) - ANTICHOLINERGIC AGENTS Days since last prescription | | 0.997990 | DISCARD |
| 2-year summary (Drug prescriptions) - ANTICHOLINERGIC AGENTS Prescription count | | 0.997990 | DISCARD |
| 1-year summary (Drug prescriptions) - ANTIBACTERIAL/ANTIPROTOZOAL Average frequency (days) | | 0.998018 | DISCARD |
| 1-year summary (Drug prescriptions) - ANTIBACTERIAL/ANTIPROTOZOAL Max frequency (days) | | 0.998018 | DISCARD |
| 1-year summary (Drug prescriptions) - ANTIBACTERIAL/ANTIPROTOZOAL Min frequency (days) | | 0.998018 | DISCARD |
| 1-year summary (Drug prescriptions) - DIGITALIS PREPARATIONS Days since first prescription | | 0.998018 | DISCARD |
| 1-year summary (Drug prescriptions) - DIGITALIS PREPARATIONS Days since last prescription | | 0.998018 | DISCARD |
| 1-year summary (Drug prescriptions) - DIGITALIS PREPARATIONS Prescription count | | 0.998018 | DISCARD |
| 1-year summary (Drug prescriptions) - NON-STEROIDAL ANTI-INFLAMMATORY Days since first prescription | | 0.998018 | DISCARD |
| 1-year summary (Drug prescriptions) - NON-STEROIDAL ANTI-INFLAMMATORY Days since last prescription | | 0.998018 | DISCARD |
| 1-year summary (Drug prescriptions) - NON-STEROIDAL ANTI-INFLAMMATORY Prescription count | | 0.998018 | DISCARD |
| 1-year summary (Drug prescriptions) - PROGESTOGENS Days since first prescription | | 0.998018 | DISCARD |
| 1-year summary (Drug prescriptions) - PROGESTOGENS Days since last prescription | | 0.998018 | DISCARD |
| 1-year summary (Drug prescriptions) - PROGESTOGENS Prescription count | | 0.998018 | DISCARD |
| 2-year summary (Drug prescriptions) - ANTI-HERPESVIRUS INFECTION Average frequency (days) | | 0.998018 | DISCARD |
| 2-year summary (Drug prescriptions) - ANTI-HERPESVIRUS INFECTION Max frequency (days) | | 0.998018 | DISCARD |
| 2-year summary (Drug prescriptions) - ANTI-HERPESVIRUS INFECTION Min frequency (days) | | 0.998018 | DISCARD |
| 1-year summary (Drug prescriptions) - CARBOXAMIDE DERIVATIVES Days since first prescription | | 0.998045 | DISCARD |
| 1-year summary (Drug prescriptions) - CARBOXAMIDE DERIVATIVES Days since last prescription | | 0.998045 | DISCARD |
| 1-year summary (Drug prescriptions) - CARBOXAMIDE DERIVATIVES Prescription count | | 0.998045 | DISCARD |
| 1-year summary (Drug prescriptions) - OPIOID DEPENDENCE Average frequency (days) | | 0.998045 | DISCARD |
| 1-year summary (Drug prescriptions) - OPIOID DEPENDENCE Max frequency (days) | | 0.998045 | DISCARD |
| 1-year summary (Drug prescriptions) - OPIOID DEPENDENCE Min frequency (days) | | 0.998045 | DISCARD |
| 2-year summary (Drug prescriptions) - AMYNOGLYCOSIDES Average frequency (days) | | 0.998045 | DISCARD |
| 2-year summary (Drug prescriptions) - AMYNOGLYCOSIDES Max frequency (days) | | 0.998045 | DISCARD |
| 2-year summary (Drug prescriptions) - AMYNOGLYCOSIDES Min frequency (days) | | 0.998045 | DISCARD |
| 2-year summary (Drug prescriptions) - POTASSIUM-REMOVING RESINS Days since first prescription | | 0.998045 | DISCARD |
| 2-year summary (Drug prescriptions) - POTASSIUM-REMOVING RESINS Days since last prescription | | 0.998045 | DISCARD |
| 2-year summary (Drug prescriptions) - POTASSIUM-REMOVING RESINS Prescription count | | 0.998045 | DISCARD |
| 1-year summary (Drug prescriptions) - ANTIFUNGALS Average frequency (days) | | 0.998073 | DISCARD |
| 1-year summary (Drug prescriptions) - ANTIFUNGALS Max frequency (days) | | 0.998073 | DISCARD |

| | | | |
|---|---|---|---|
| 1-year summary (Drug prescriptions) - ANTIFUNGALS Min frequency (days) | | 0.998073 | DISCARD |
| 1-year summary (Drug prescriptions) - CARBOXAMIDE DERIVATIVES Average frequency (days) | | 0.998100 | DISCARD |
| 1-year summary (Drug prescriptions) - CARBOXAMIDE DERIVATIVES Max frequency (days) | | 0.998100 | DISCARD |
| 1-year summary (Drug prescriptions) - CARBOXAMIDE DERIVATIVES Min frequency (days) | | 0.998100 | DISCARD |
| 1-year summary (Drug prescriptions) - CORTICOSTEROIDS WITH OTHER COMBINATIONS Days since first prescription | | 0.998100 | DISCARD |
| 1-year summary (Drug prescriptions) - CORTICOSTEROIDS WITH OTHER COMBINATIONS Days since last prescription | | 0.998100 | DISCARD |
| 1-year summary (Drug prescriptions) - CORTICOSTEROIDS WITH OTHER COMBINATIONS Prescription count | | 0.998100 | DISCARD |
| 1-year summary (Drug prescriptions) - DIGITALIS PREPARATIONS Average frequency (days) | | 0.998100 | DISCARD |
| 1-year summary (Drug prescriptions) - DIGITALIS PREPARATIONS Max frequency (days) | | 0.998100 | DISCARD |
| 1-year summary (Drug prescriptions) - DIGITALIS PREPARATIONS Min frequency (days) | | 0.998100 | DISCARD |
| 1-year summary (Drug prescriptions) - LEVODOPA AND COMBINATION Average frequency (days) | | 0.998100 | DISCARD |
| 1-year summary (Drug prescriptions) - LEVODOPA AND COMBINATION Max frequency (days) | | 0.998100 | DISCARD |
| 1-year summary (Drug prescriptions) - LEVODOPA AND COMBINATION Min frequency (days) | | 0.998100 | DISCARD |
| 1-year summary (Drug prescriptions) - TETRACYCLINES Average frequency (days) | | 0.998100 | DISCARD |
| 1-year summary (Drug prescriptions) - TETRACYCLINES Max frequency (days) | | 0.998100 | DISCARD |
| 1-year summary (Drug prescriptions) - TETRACYCLINES Min frequency (days) | | 0.998100 | DISCARD |
| 2-year summary (Drug prescriptions) - ANTILIPEMIC: FIBRATES Average frequency (days) | | 0.998100 | DISCARD |
| 2-year summary (Drug prescriptions) - ANTILIPEMIC: FIBRATES Max frequency (days) | | 0.998100 | DISCARD |
| 2-year summary (Drug prescriptions) - ANTILIPEMIC: FIBRATES Min frequency (days) | | 0.998100 | DISCARD |
| 2-year summary (Drug prescriptions) - PLASMODICIDES (ANTIMALARIALS) Average frequency (days) | | 0.998100 | DISCARD |
| 2-year summary (Drug prescriptions) - PLASMODICIDES (ANTIMALARIALS) Max frequency (days) | | 0.998100 | DISCARD |
| 2-year summary (Drug prescriptions) - PLASMODICIDES (ANTIMALARIALS) Min frequency (days) | | 0.998100 | DISCARD |
| 1-year summary (Drug prescriptions) - ACNE THERAPY Average frequency (days) | | 0.998128 | DISCARD |
| 1-year summary (Drug prescriptions) - ACNE THERAPY Max frequency (days) | | 0.998128 | DISCARD |
| 1-year summary (Drug prescriptions) - ACNE THERAPY Min frequency (days) | | 0.998128 | DISCARD |
| 1-year summary (Drug prescriptions) - NUTRITIONAL PRODUCTS Average frequency (days) | | 0.998128 | DISCARD |
| 1-year summary (Drug prescriptions) - NUTRITIONAL PRODUCTS Max frequency (days) | | 0.998128 | DISCARD |
| 1-year summary (Drug prescriptions) - NUTRITIONAL PRODUCTS Min frequency (days) | | 0.998128 | DISCARD |
| 2-year summary (Drug prescriptions) - PLATELET AGGREGATION INHIBITORS Days since first prescription | | 0.998128 | DISCARD |
| 2-year summary (Drug prescriptions) - PLATELET AGGREGATION INHIBITORS Days since last prescription | | 0.998128 | DISCARD |
| 2-year summary (Drug prescriptions) - PLATELET AGGREGATION INHIBITORS Prescription count | | 0.998128 | DISCARD |
| 2-year summary (Drug prescriptions) - OPHTHALMOLOGICALS Average frequency (days) | | 0.998183 | DISCARD |
| 2-year summary (Drug prescriptions) - OPHTHALMOLOGICALS Max frequency (days) | | 0.998183 | DISCARD |
| 2-year summary (Drug prescriptions) - OPHTHALMOLOGICALS Min frequency (days) | | 0.998183 | DISCARD |
| 2-year summary (Drug prescriptions) - PARASITICIDES Days since first prescription | | 0.998183 | DISCARD |
| 2-year summary (Drug prescriptions) - PARASITICIDES Days since last prescription | | 0.998183 | DISCARD |
| 2-year summary (Drug prescriptions) - PARASITICIDES Prescription count | | 0.998183 | DISCARD |

| | | | |
|---|---|---|---|
| 2-year summary (Drug prescriptions) - DOPAMINE AGONISTS Days since first prescription | | 0.998211 | DISCARD |
| 2-year summary (Drug prescriptions) - DOPAMINE AGONISTS Days since last prescription | | 0.998211 | DISCARD |
| 2-year summary (Drug prescriptions) - DOPAMINE AGONISTS Prescription count | | 0.998211 | DISCARD |
| 2-year summary (Drug prescriptions) - VASOPRESSORS Average frequency (days) | | 0.998211 | DISCARD |
| 2-year summary (Drug prescriptions) - VASOPRESSORS Max frequency (days) | | 0.998211 | DISCARD |
| 2-year summary (Drug prescriptions) - VASOPRESSORS Min frequency (days) | | 0.998211 | DISCARD |
| 1-year summary (Drug prescriptions) - PLASMODICIDES (ANTIMALARIALS) Days since first prescription | | 0.998238 | DISCARD |
| 1-year summary (Drug prescriptions) - PLASMODICIDES (ANTIMALARIALS) Days since last prescription | | 0.998238 | DISCARD |
| 1-year summary (Drug prescriptions) - PLASMODICIDES (ANTIMALARIALS) Prescription count | | 0.998238 | DISCARD |
| 1-year summary (Drug prescriptions) - VITAMIN B12 PREP Average frequency (days) | | 0.998238 | DISCARD |
| 1-year summary (Drug prescriptions) - VITAMIN B12 PREP Max frequency (days) | | 0.998238 | DISCARD |
| 1-year summary (Drug prescriptions) - VITAMIN B12 PREP Min frequency (days) | | 0.998238 | DISCARD |
| 2-year summary (Drug prescriptions) - PLATELET AGGREGATION INHIBITORS Average frequency (days) | | 0.998238 | DISCARD |
| 2-year summary (Drug prescriptions) - PLATELET AGGREGATION INHIBITORS Max frequency (days) | | 0.998238 | DISCARD |
| 2-year summary (Drug prescriptions) - PLATELET AGGREGATION INHIBITORS Min frequency (days) | | 0.998238 | DISCARD |
| 1-year summary (Drug prescriptions) - OPHTHALMOLOGICALS Days since first prescription | | 0.998266 | DISCARD |
| 1-year summary (Drug prescriptions) - OPHTHALMOLOGICALS Days since last prescription | | 0.998266 | DISCARD |
| 1-year summary (Drug prescriptions) - OPHTHALMOLOGICALS Prescription count | | 0.998266 | DISCARD |
| 1-year summary (Drug prescriptions) - ANTIVIRALS Days since first prescription | | 0.998293 | DISCARD |
| 1-year summary (Drug prescriptions) - ANTIVIRALS Days since last prescription | | 0.998293 | DISCARD |
| 1-year summary (Drug prescriptions) - ANTIVIRALS Prescription count | | 0.998293 | DISCARD |
| 2-year summary (Drug prescriptions) - ANTICOAGULANTS MISCELLANEOUS Average frequency (days) | | 0.998293 | DISCARD |
| 2-year summary (Drug prescriptions) - ANTICOAGULANTS MISCELLANEOUS Days since first prescription | | 0.998293 | DISCARD |
| 2-year summary (Drug prescriptions) - ANTICOAGULANTS MISCELLANEOUS Days since last prescription | | 0.998293 | DISCARD |
| 2-year summary (Drug prescriptions) - ANTICOAGULANTS MISCELLANEOUS Max frequency (days) | | 0.998293 | DISCARD |
| 2-year summary (Drug prescriptions) - ANTICOAGULANTS MISCELLANEOUS Min frequency (days) | | 0.998293 | DISCARD |
| 2-year summary (Drug prescriptions) - ANTICOAGULANTS MISCELLANEOUS Prescription count | | 0.998293 | DISCARD |
| 2-year summary (Drug prescriptions) - CALCIUM BLOCKERS ANTILIPEMIC COMBINATIONS Days since first prescription | | 0.998293 | DISCARD |
| 2-year summary (Drug prescriptions) - CALCIUM BLOCKERS ANTILIPEMIC COMBINATIONS Days since last prescription | | 0.998293 | DISCARD |
| 2-year summary (Drug prescriptions) - CALCIUM BLOCKERS ANTILIPEMIC COMBINATIONS Prescription count | | 0.998293 | DISCARD |
| 2-year summary (Drug prescriptions) - LINCOSAMIDES Average frequency (days) | | 0.998321 | DISCARD |
| 2-year summary (Drug prescriptions) - LINCOSAMIDES Max frequency (days) | | 0.998321 | DISCARD |
| 2-year summary (Drug prescriptions) - LINCOSAMIDES Min frequency (days) | | 0.998321 | DISCARD |
| 2-year summary (Laboratory tests) - number of tests for GLUCOSE^1H_POST_75_G_GLUCOSE_PO | | 0.998321 | DISCARD |
| 2-year summary (Laboratory tests) - days since earliest test for GLUCOSE^1H_POST_75_G_GLUCOSE_PO | | 0.998321 | DISCARD |
| 2-year summary (Laboratory tests) - days since latest test for GLUCOSE^1H_POST_75_G_GLUCOSE_PO | | 0.998321 | DISCARD |
| 2-year summary (Laboratory tests) - number of high values of GLUCOSE^1H_POST_75_G_GLUCOSE_PO | | 0.998321 | DISCARD |

| | | |
|---|---|---|
| 2-year summary (Laboratory tests) - highest value of GLUCOSE^1H_POST_75_G_GLUCOSE_PO | 0.998321 | DISCARD |
| 2-year summary (Laboratory tests) - number of low values of GLUCOSE^1H_POST_75_G_GLUCOSE_PO | 0.998321 | DISCARD |
| 2-year summary (Laboratory tests) - lowest value of GLUCOSE^1H_POST_75_G_GLUCOSE_PO | 0.998321 | DISCARD |
| 2-year summary (Laboratory tests) - average value of GLUCOSE^1H_POST_75_G_GLUCOSE_PO | 0.998321 | DISCARD |
| 2-year summary (Laboratory tests) - median value of GLUCOSE^1H_POST_75_G_GLUCOSE_PO | 0.998321 | DISCARD |
| 2-year summary (Laboratory tests) - count of normal tests for GLUCOSE^1H_POST_75_G_GLUCOSE_PO | 0.998321 | DISCARD |
| 1-year summary (Drug prescriptions) - ANTILIPEMIC: FIBRATES Days since first prescription | 0.998348 | DISCARD |
| 1-year summary (Drug prescriptions) - ANTILIPEMIC: FIBRATES Days since last prescription | 0.998348 | DISCARD |
| 1-year summary (Drug prescriptions) - ANTILIPEMIC: FIBRATES Prescription count | 0.998348 | DISCARD |
| 1-year summary (Drug prescriptions) - CORTICOSTEROIDS AND ANTI-INFECTIVE COMBINATIONS Average frequency (days) | 0.998348 | DISCARD |
| 1-year summary (Drug prescriptions) - CORTICOSTEROIDS AND ANTI-INFECTIVE COMBINATIONS Max frequency (days) | 0.998348 | DISCARD |
| 1-year summary (Drug prescriptions) - CORTICOSTEROIDS AND ANTI-INFECTIVE COMBINATIONS Min frequency (days) | 0.998348 | DISCARD |
| 2-year summary (Drug prescriptions) - CALCIUM BLOCKERS ANTILIPEMIC COMBINATIONS Average frequency (days) | 0.998348 | DISCARD |
| 2-year summary (Drug prescriptions) - CALCIUM BLOCKERS ANTILIPEMIC COMBINATIONS Max frequency (days) | 0.998348 | DISCARD |
| 2-year summary (Drug prescriptions) - CALCIUM BLOCKERS ANTILIPEMIC COMBINATIONS Min frequency (days) | 0.998348 | DISCARD |
| 2-year summary (Drug prescriptions) - LUTEINIZING HORMONE-RELEASING HORMONE (LHRH) ANALOG Days since first prescri | 0.998348 | DISCARD |
| 2-year summary (Drug prescriptions) - LUTEINIZING HORMONE-RELEASING HORMONE (LHRH) ANALOG Days since last prescri | 0.998348 | DISCARD |
| 2-year summary (Drug prescriptions) - LUTEINIZING HORMONE-RELEASING HORMONE (LHRH) ANALOG Prescription count | 0.998348 | DISCARD |
| 2-year summary (Drug prescriptions) - ADRENERGIC AGONISTS Days since first prescription | 0.998376 | DISCARD |
| 2-year summary (Drug prescriptions) - ADRENERGIC AGONISTS Days since last prescription | 0.998376 | DISCARD |
| 2-year summary (Drug prescriptions) - ADRENERGIC AGONISTS Prescription count | 0.998376 | DISCARD |
| 2-year summary (Drug prescriptions) - TESTOSTERONES AND RELATED HORMONES Days since first prescription | 0.998376 | DISCARD |
| 2-year summary (Drug prescriptions) - TESTOSTERONES AND RELATED HORMONES Days since last prescription | 0.998376 | DISCARD |
| 2-year summary (Drug prescriptions) - TESTOSTERONES AND RELATED HORMONES Prescription count | 0.998376 | DISCARD |
| 1-year summary (Laboratory tests) - number of tests for Cholesterol.total/Cholesterol.in_HDL | 0.998403 | DISCARD |
| 1-year summary (Laboratory tests) - days since earliest test for Cholesterol.total/Cholesterol.in_HDL | 0.998403 | DISCARD |
| 1-year summary (Laboratory tests) - days since latest test for Cholesterol.total/Cholesterol.in_HDL | 0.998403 | DISCARD |
| 1-year summary (Laboratory tests) - number of high values of Cholesterol.total/Cholesterol.in_HDL | 0.998403 | DISCARD |
| 1-year summary (Laboratory tests) - highest value of Cholesterol.total/Cholesterol.in_HDL | 0.998403 | DISCARD |
| 1-year summary (Laboratory tests) - number of low values of Cholesterol.total/Cholesterol.in_HDL | 0.998403 | DISCARD |
| 1-year summary (Laboratory tests) - lowest value of Cholesterol.total/Cholesterol.in_HDL | 0.998403 | DISCARD |
| 1-year summary (Laboratory tests) - average value of Cholesterol.total/Cholesterol.in_HDL | 0.998403 | DISCARD |
| 1-year summary (Laboratory tests) - median value of Cholesterol.total/Cholesterol.in_HDL | 0.998403 | DISCARD |
| 1-year summary (Laboratory tests) - count of normal tests for Cholesterol.total/Cholesterol.in_HDL | 0.998403 | DISCARD |
| 2-year summary (Drug prescriptions) - ANTI-HEMORRHOIDAL AGENTS WITHOUT CORTICOSTEROIDS Average frequency (days) | 0.998403 | DISCARD |
| 2-year summary (Drug prescriptions) - ANTI-HEMORRHOIDAL AGENTS WITHOUT CORTICOSTEROIDS Max frequency (days) | 0.998403 | DISCARD |
| 2-year summary (Drug prescriptions) - ANTI-HEMORRHOIDAL AGENTS WITHOUT CORTICOSTEROIDS Min frequency (days) | 0.998403 | DISCARD |

| | | | |
|---|---|---|---|
| 2-year summary (Drug prescriptions) - CENTRALLY ACTING ANTIADRENERGIC Days since first prescription | | 0.998403 | DISCARD |
| 2-year summary (Drug prescriptions) - CENTRALLY ACTING ANTIADRENERGIC Days since last prescription | | 0.998403 | DISCARD |
| 2-year summary (Drug prescriptions) - CENTRALLY ACTING ANTIADRENERGIC Prescription count | | 0.998403 | DISCARD |
| 2-year summary (Drug prescriptions) - MISCELLANEOUS LOCAL ANTI-INFECTIVES Days since first prescription | | 0.998403 | DISCARD |
| 2-year summary (Drug prescriptions) - MISCELLANEOUS LOCAL ANTI-INFECTIVES Days since last prescription | | 0.998403 | DISCARD |
| 2-year summary (Drug prescriptions) - MISCELLANEOUS LOCAL ANTI-INFECTIVES Prescription count | | 0.998403 | DISCARD |
| 2-year summary (Drug prescriptions) - ANTICHOLINERGIC AGENTS Average frequency (days) | | 0.998431 | DISCARD |
| 2-year summary (Drug prescriptions) - ANTICHOLINERGIC AGENTS Max frequency (days) | | 0.998431 | DISCARD |
| 2-year summary (Drug prescriptions) - ANTICHOLINERGIC AGENTS Min frequency (days) | | 0.998431 | DISCARD |
| 2-year summary (Drug prescriptions) - CLASS III ANTIARRHYTHMIC DRUGS Days since first prescription | | 0.998431 | DISCARD |
| 2-year summary (Drug prescriptions) - CLASS III ANTIARRHYTHMIC DRUGS Days since last prescription | | 0.998431 | DISCARD |
| 2-year summary (Drug prescriptions) - CLASS III ANTIARRHYTHMIC DRUGS Prescription count | | 0.998431 | DISCARD |
| 2-year summary (Drug prescriptions) - LOW MOLECULAR WEIGHT HEPARINS (LMWH) Days since first prescription | | 0.998431 | DISCARD |
| 2-year summary (Drug prescriptions) - LOW MOLECULAR WEIGHT HEPARINS (LMWH) Days since last prescription | | 0.998431 | DISCARD |
| 2-year summary (Drug prescriptions) - LOW MOLECULAR WEIGHT HEPARINS (LMWH) Prescription count | | 0.998431 | DISCARD |
| 1-year summary (Drug prescriptions) - PLASMODICIDES (ANTIMALARIALS) Average frequency (days) | | 0.998458 | DISCARD |
| 1-year summary (Drug prescriptions) - PLASMODICIDES (ANTIMALARIALS) Max frequency (days) | | 0.998458 | DISCARD |
| 1-year summary (Drug prescriptions) - PLASMODICIDES (ANTIMALARIALS) Min frequency (days) | | 0.998458 | DISCARD |
| 1-year summary (Drug prescriptions) - CALCIUM BLOCKERS ANTILIPEMIC COMBINATIONS Average frequency (days) | | 0.998486 | DISCARD |
| 1-year summary (Drug prescriptions) - CALCIUM BLOCKERS ANTILIPEMIC COMBINATIONS Days since first prescription | | 0.998486 | DISCARD |
| 1-year summary (Drug prescriptions) - CALCIUM BLOCKERS ANTILIPEMIC COMBINATIONS Days since last prescription | | 0.998486 | DISCARD |
| 1-year summary (Drug prescriptions) - CALCIUM BLOCKERS ANTILIPEMIC COMBINATIONS Max frequency (days) | | 0.998486 | DISCARD |
| 1-year summary (Drug prescriptions) - CALCIUM BLOCKERS ANTILIPEMIC COMBINATIONS Min frequency (days) | | 0.998486 | DISCARD |
| 1-year summary (Drug prescriptions) - CALCIUM BLOCKERS ANTILIPEMIC COMBINATIONS Prescription count | | 0.998486 | DISCARD |
| 1-year summary (Drug prescriptions) - TOPICAL IMMUNOSUPRESSIVE AGENTS Days since first prescription | | 0.998486 | DISCARD |
| 1-year summary (Drug prescriptions) - TOPICAL IMMUNOSUPRESSIVE AGENTS Days since last prescription | | 0.998486 | DISCARD |
| 1-year summary (Drug prescriptions) - TOPICAL IMMUNOSUPRESSIVE AGENTS Prescription count | | 0.998486 | DISCARD |
| 1-year summary (Drug prescriptions) - ANTICHOLINERGIC AGENTS Days since first prescription | | 0.998513 | DISCARD |
| 1-year summary (Drug prescriptions) - ANTICHOLINERGIC AGENTS Days since last prescription | | 0.998513 | DISCARD |
| 1-year summary (Drug prescriptions) - ANTICHOLINERGIC AGENTS Prescription count | | 0.998513 | DISCARD |
| 1-year summary (Drug prescriptions) - PLATELET AGGREGATION INHIBITORS Days since first prescription | | 0.998513 | DISCARD |
| 1-year summary (Drug prescriptions) - PLATELET AGGREGATION INHIBITORS Days since last prescription | | 0.998513 | DISCARD |
| 1-year summary (Drug prescriptions) - PLATELET AGGREGATION INHIBITORS Prescription count | | 0.998513 | DISCARD |
| 2-year summary (Drug prescriptions) - DOPAMINE AGONISTS Average frequency (days) | | 0.998513 | DISCARD |
| 2-year summary (Drug prescriptions) - DOPAMINE AGONISTS Max frequency (days) | | 0.998513 | DISCARD |
| 2-year summary (Drug prescriptions) - DOPAMINE AGONISTS Min frequency (days) | | 0.998513 | DISCARD |
| 1-year summary (Drug prescriptions) - DOPAMINE AGONISTS Days since first prescription | | 0.998541 | DISCARD |

| | | | |
|---|---|---|---|
| 1-year summary (Drug prescriptions) - DOPAMINE AGONISTS Days since last prescription | | 0.998541 | DISCARD |
| 1-year summary (Drug prescriptions) - DOPAMINE AGONISTS Prescription count | | 0.998541 | DISCARD |
| 1-year summary (Drug prescriptions) - LUTEINIZING HORMONE-RELEASING HORMONE (LHRH) ANALOG Days since first prescri | | 0.998541 | DISCARD |
| 1-year summary (Drug prescriptions) - LUTEINIZING HORMONE-RELEASING HORMONE (LHRH) ANALOG Days since last prescri | | 0.998541 | DISCARD |
| 1-year summary (Drug prescriptions) - LUTEINIZING HORMONE-RELEASING HORMONE (LHRH) ANALOG Prescription count | | 0.998541 | DISCARD |
| 1-year summary (Drug prescriptions) - OPHTHALMOLOGICALS Average frequency (days) | | 0.998541 | DISCARD |
| 1-year summary (Drug prescriptions) - OPHTHALMOLOGICALS Max frequency (days) | | 0.998541 | DISCARD |
| 1-year summary (Drug prescriptions) - OPHTHALMOLOGICALS Min frequency (days) | | 0.998541 | DISCARD |
| 2-year summary (Drug prescriptions) - BIOLOGIC RESPONSE MODIFYING AGENTS Days since first prescription | | 0.998541 | DISCARD |
| 2-year summary (Drug prescriptions) - BIOLOGIC RESPONSE MODIFYING AGENTS Days since last prescription | | 0.998541 | DISCARD |
| 2-year summary (Drug prescriptions) - BIOLOGIC RESPONSE MODIFYING AGENTS Prescription count | | 0.998541 | DISCARD |
| 2-year summary (Drug prescriptions) - HYDANTOIN DERIVATIVES Days since first prescription | | 0.998541 | DISCARD |
| 2-year summary (Drug prescriptions) - HYDANTOIN DERIVATIVES Days since last prescription | | 0.998541 | DISCARD |
| 2-year summary (Drug prescriptions) - HYDANTOIN DERIVATIVES Prescription count | | 0.998541 | DISCARD |
| 2-year summary (Doctor visits) - ORAL SURGERY | | 0.998541 | DISCARD |
| 2-year summary (Drug prescriptions) - ANTACIDS & ADSORBENTS Days since first prescription | | 0.998568 | DISCARD |
| 2-year summary (Drug prescriptions) - ANTACIDS & ADSORBENTS Days since last prescription | | 0.998568 | DISCARD |
| 2-year summary (Drug prescriptions) - ANTACIDS & ADSORBENTS Prescription count | | 0.998568 | DISCARD |
| 1-year summary (Drug prescriptions) - ANTILIPEMIC: FIBRATES Average frequency (days) | | 0.998596 | DISCARD |
| 1-year summary (Drug prescriptions) - ANTILIPEMIC: FIBRATES Max frequency (days) | | 0.998596 | DISCARD |
| 1-year summary (Drug prescriptions) - ANTILIPEMIC: FIBRATES Min frequency (days) | | 0.998596 | DISCARD |
| 1-year summary (Drug prescriptions) - POTASSIUM-REMOVING RESINS Days since first prescription | | 0.998596 | DISCARD |
| 1-year summary (Drug prescriptions) - POTASSIUM-REMOVING RESINS Days since last prescription | | 0.998596 | DISCARD |
| 1-year summary (Drug prescriptions) - POTASSIUM-REMOVING RESINS Prescription count | | 0.998596 | DISCARD |
| 2-year summary (Drug prescriptions) - BIOLOGIC RESPONSE MODIFYING AGENTS Average frequency (days) | | 0.998596 | DISCARD |
| 2-year summary (Drug prescriptions) - BIOLOGIC RESPONSE MODIFYING AGENTS Max frequency (days) | | 0.998596 | DISCARD |
| 2-year summary (Drug prescriptions) - BIOLOGIC RESPONSE MODIFYING AGENTS Min frequency (days) | | 0.998596 | DISCARD |
| 2-year summary (Drug prescriptions) - LUTEINIZING HORMONE-RELEASING HORMONE (LHRH) ANALOG Average frequency (da | | 0.998596 | DISCARD |
| 2-year summary (Drug prescriptions) - LUTEINIZING HORMONE-RELEASING HORMONE (LHRH) ANALOG Max frequency (days) | | 0.998596 | DISCARD |
| 2-year summary (Drug prescriptions) - LUTEINIZING HORMONE-RELEASING HORMONE (LHRH) ANALOG Min frequency (days) | | 0.998596 | DISCARD |
| 2-year summary (Drug prescriptions) - SMOKING CESSATION AIDS Days since first prescription | | 0.998596 | DISCARD |
| 2-year summary (Drug prescriptions) - SMOKING CESSATION AIDS Days since last prescription | | 0.998596 | DISCARD |
| 2-year summary (Drug prescriptions) - SMOKING CESSATION AIDS Prescription count | | 0.998596 | DISCARD |
| 1-year summary (Drug prescriptions) - MISCELLANEOUS SKIN AND MUCOUS MEMBRANE AGENTS Days since first prescription | | 0.998623 | DISCARD |
| 1-year summary (Drug prescriptions) - MISCELLANEOUS SKIN AND MUCOUS MEMBRANE AGENTS Days since last prescription | | 0.998623 | DISCARD |
| 1-year summary (Drug prescriptions) - MISCELLANEOUS SKIN AND MUCOUS MEMBRANE AGENTS Prescription count | | 0.998623 | DISCARD |
| 1-year summary (Laboratory tests) - number of tests for Glucose^1H_post_50_g_glucose_PO | | 0.998623 | DISCARD |

| | | | |
|---|---|---|---|
| 1-year summary (Laboratory tests) - days since earliest test for Glucose^1H_post_50_g_glucose_PO | | 0.998623 | DISCARD |
| 1-year summary (Laboratory tests) - days since latest test for Glucose^1H_post_50_g_glucose_PO | | 0.998623 | DISCARD |
| 1-year summary (Laboratory tests) - number of high values of Glucose^1H_post_50_g_glucose_PO | | 0.998623 | DISCARD |
| 1-year summary (Laboratory tests) - highest value of Glucose^1H_post_50_g_glucose_PO | | 0.998623 | DISCARD |
| 1-year summary (Laboratory tests) - number of low values of Glucose^1H_post_50_g_glucose_PO | | 0.998623 | DISCARD |
| 1-year summary (Laboratory tests) - lowest value of Glucose^1H_post_50_g_glucose_PO | | 0.998623 | DISCARD |
| 1-year summary (Laboratory tests) - average value of Glucose^1H_post_50_g_glucose_PO | | 0.998623 | DISCARD |
| 1-year summary (Laboratory tests) - median value of Glucose^1H_post_50_g_glucose_PO | | 0.998623 | DISCARD |
| 1-year summary (Laboratory tests) - count of normal tests for Glucose^1H_post_50_g_glucose_PO | | 0.998623 | DISCARD |
| 2-year summary (Drug prescriptions) - CHOLINERGIC AGONISTS Days since first prescription | | 0.998623 | DISCARD |
| 2-year summary (Drug prescriptions) - CHOLINERGIC AGONISTS Days since last prescription | | 0.998623 | DISCARD |
| 2-year summary (Drug prescriptions) - CHOLINERGIC AGONISTS Prescription count | | 0.998623 | DISCARD |
| 2-year summary (Drug prescriptions) - CORTICOSTEROIDS WITH OTHER COMBINATIONS Average frequency (days) | | 0.998623 | DISCARD |
| 2-year summary (Drug prescriptions) - CORTICOSTEROIDS WITH OTHER COMBINATIONS Max frequency (days) | | 0.998623 | DISCARD |
| 2-year summary (Drug prescriptions) - CORTICOSTEROIDS WITH OTHER COMBINATIONS Min frequency (days) | | 0.998623 | DISCARD |
| 1-year summary (Drug prescriptions) - ANTICHOLINERGIC AGENTS Average frequency (days) | | 0.998651 | DISCARD |
| 1-year summary (Drug prescriptions) - ANTICHOLINERGIC AGENTS Max frequency (days) | | 0.998651 | DISCARD |
| 1-year summary (Drug prescriptions) - ANTICHOLINERGIC AGENTS Min frequency (days) | | 0.998651 | DISCARD |
| 1-year summary (Drug prescriptions) - HYDANTOIN DERIVATIVES Days since first prescription | | 0.998651 | DISCARD |
| 1-year summary (Drug prescriptions) - HYDANTOIN DERIVATIVES Days since last prescription | | 0.998651 | DISCARD |
| 1-year summary (Drug prescriptions) - HYDANTOIN DERIVATIVES Prescription count | | 0.998651 | DISCARD |
| 1-year summary (Drug prescriptions) - UNCLASSIFIED THERAPEUTIC AGENTS: OTHER Average frequency (days) | | 0.998651 | DISCARD |
| 1-year summary (Drug prescriptions) - UNCLASSIFIED THERAPEUTIC AGENTS: OTHER Max frequency (days) | | 0.998651 | DISCARD |
| 1-year summary (Drug prescriptions) - UNCLASSIFIED THERAPEUTIC AGENTS: OTHER Min frequency (days) | | 0.998651 | DISCARD |
| 1-year summary (Drug prescriptions) - ANTIDIARRHEA Average frequency (days) | | 0.998679 | DISCARD |
| 1-year summary (Drug prescriptions) - ANTIDIARRHEA Max frequency (days) | | 0.998679 | DISCARD |
| 1-year summary (Drug prescriptions) - ANTIDIARRHEA Min frequency (days) | | 0.998679 | DISCARD |
| 1-year summary (Drug prescriptions) - DOPAMINE AGONISTS Average frequency (days) | | 0.998679 | DISCARD |
| 1-year summary (Drug prescriptions) - DOPAMINE AGONISTS Max frequency (days) | | 0.998679 | DISCARD |
| 1-year summary (Drug prescriptions) - DOPAMINE AGONISTS Min frequency (days) | | 0.998679 | DISCARD |
| 1-year summary (Drug prescriptions) - PLATELET AGGREGATION INHIBITORS Average frequency (days) | | 0.998679 | DISCARD |
| 1-year summary (Drug prescriptions) - PLATELET AGGREGATION INHIBITORS Max frequency (days) | | 0.998679 | DISCARD |
| 1-year summary (Drug prescriptions) - PLATELET AGGREGATION INHIBITORS Min frequency (days) | | 0.998679 | DISCARD |
| 2-year summary (Drug prescriptions) - ANTIGLAUCOMA AGENTS Days since first prescription | | 0.998679 | DISCARD |
| 2-year summary (Drug prescriptions) - ANTIGLAUCOMA AGENTS Days since last prescription | | 0.998679 | DISCARD |
| 2-year summary (Drug prescriptions) - ANTIGLAUCOMA AGENTS Prescription count | | 0.998679 | DISCARD |
| 1-year summary (Drug prescriptions) - ANTACIDS & ADSORBENTS Days since first prescription | | 0.998706 | DISCARD |

| | | |
|---|---|---|
| 1-year summary (Drug prescriptions) - ANTACIDS & ADSORBENTS Days since last prescription | 0.998706 | DISCARD |
| 1-year summary (Drug prescriptions) - ANTACIDS & ADSORBENTS Prescription count | 0.998706 | DISCARD |
| 1-year summary (Drug prescriptions) - NEEDLE/SYRINGE/LANCET Average frequency (days) | 0.998706 | DISCARD |
| 1-year summary (Drug prescriptions) - NEEDLE/SYRINGE/LANCET Max frequency (days) | 0.998706 | DISCARD |
| 1-year summary (Drug prescriptions) - NEEDLE/SYRINGE/LANCET Min frequency (days) | 0.998706 | DISCARD |
| 2-year summary (Drug prescriptions) - AROMATASE INHIBITORS Days since first prescription | 0.998706 | DISCARD |
| 2-year summary (Drug prescriptions) - AROMATASE INHIBITORS Days since last prescription | 0.998706 | DISCARD |
| 2-year summary (Drug prescriptions) - AROMATASE INHIBITORS Prescription count | 0.998706 | DISCARD |
| 2-year summary (Drug prescriptions) - CLASS III ANTIARRHYTHMIC DRUGS Average frequency (days) | 0.998706 | DISCARD |
| 2-year summary (Drug prescriptions) - CLASS III ANTIARRHYTHMIC DRUGS Max frequency (days) | 0.998706 | DISCARD |
| 2-year summary (Drug prescriptions) - CLASS III ANTIARRHYTHMIC DRUGS Min frequency (days) | 0.998706 | DISCARD |
| 2-year summary (Drug prescriptions) - HYDANTOIN DERIVATIVES Average frequency (days) | 0.998706 | DISCARD |
| 2-year summary (Drug prescriptions) - HYDANTOIN DERIVATIVES Max frequency (days) | 0.998706 | DISCARD |
| 2-year summary (Drug prescriptions) - HYDANTOIN DERIVATIVES Min frequency (days) | 0.998706 | DISCARD |
| 1-year summary (Laboratory tests) - number of tests for Glucose^2H_post_75_g_glucose_PO | 0.998734 | DISCARD |
| 1-year summary (Laboratory tests) - days since earliest test for Glucose^2H_post_75_g_glucose_PO | 0.998734 | DISCARD |
| 1-year summary (Laboratory tests) - days since latest test for Glucose^2H_post_75_g_glucose_PO | 0.998734 | DISCARD |
| 1-year summary (Laboratory tests) - number of high values of Glucose^2H_post_75_g_glucose_PO | 0.998734 | DISCARD |
| 1-year summary (Laboratory tests) - highest value of Glucose^2H_post_75_g_glucose_PO | 0.998734 | DISCARD |
| 1-year summary (Laboratory tests) - number of low values of Glucose^2H_post_75_g_glucose_PO | 0.998734 | DISCARD |
| 1-year summary (Laboratory tests) - lowest value of Glucose^2H_post_75_g_glucose_PO | 0.998734 | DISCARD |
| 1-year summary (Laboratory tests) - average value of Glucose^2H_post_75_g_glucose_PO | 0.998734 | DISCARD |
| 1-year summary (Laboratory tests) - median value of Glucose^2H_post_75_g_glucose_PO | 0.998734 | DISCARD |
| 1-year summary (Laboratory tests) - count of normal tests for Glucose^2H_post_75_g_glucose_PO | 0.998734 | DISCARD |
| 2-year summary (Hospitalizations) - Class - Congenital malformations, deformations and chromosomal abnormalities (Q00-Q99) | 0.998734 | DISCARD |
| 2-year summary (Drug prescriptions) - AROMATASE INHIBITORS Average frequency (days) | 0.998734 | DISCARD |
| 2-year summary (Drug prescriptions) - AROMATASE INHIBITORS Max frequency (days) | 0.998734 | DISCARD |
| 2-year summary (Drug prescriptions) - AROMATASE INHIBITORS Min frequency (days) | 0.998734 | DISCARD |
| 2-year summary (Drug prescriptions) - IMMUNOMODULATING AGENTS Days since first prescription | 0.998734 | DISCARD |
| 2-year summary (Drug prescriptions) - IMMUNOMODULATING AGENTS Days since last prescription | 0.998734 | DISCARD |
| 2-year summary (Drug prescriptions) - IMMUNOMODULATING AGENTS Prescription count | 0.998734 | DISCARD |
| 2-year summary (Drug prescriptions) - REPLACEMENT AGENTS Average frequency (days) | 0.998734 | DISCARD |
| 2-year summary (Drug prescriptions) - REPLACEMENT AGENTS Max frequency (days) | 0.998734 | DISCARD |
| 2-year summary (Drug prescriptions) - REPLACEMENT AGENTS Min frequency (days) | 0.998734 | DISCARD |
| 2-year summary (Drug prescriptions) - TESTOSTERONES AND RELATED HORMONES Average frequency (days) | 0.998734 | DISCARD |
| 2-year summary (Drug prescriptions) - TESTOSTERONES AND RELATED HORMONES Max frequency (days) | 0.998734 | DISCARD |
| 2-year summary (Drug prescriptions) - TESTOSTERONES AND RELATED HORMONES Min frequency (days) | 0.998734 | DISCARD |

| | | | |
|---|---|---|---|
| 2-year summary (Doctor visits) - DENTAL SURGERY | | 0.998734 | DISCARD |
| 1-year summary (Drug prescriptions) - ADRENERGIC AGONISTS Days since first prescription | | 0.998761 | DISCARD |
| 1-year summary (Drug prescriptions) - ADRENERGIC AGONISTS Days since last prescription | | 0.998761 | DISCARD |
| 1-year summary (Drug prescriptions) - ADRENERGIC AGONISTS Prescription count | | 0.998761 | DISCARD |
| 1-year summary (Drug prescriptions) - PROGESTOGENS Average frequency (days) | | 0.998761 | DISCARD |
| 1-year summary (Drug prescriptions) - PROGESTOGENS Max frequency (days) | | 0.998761 | DISCARD |
| 1-year summary (Drug prescriptions) - PROGESTOGENS Min frequency (days) | | 0.998761 | DISCARD |
| 2-year summary (Drug prescriptions) - ADRENERGIC AGONISTS Average frequency (days) | | 0.998761 | DISCARD |
| 2-year summary (Drug prescriptions) - ADRENERGIC AGONISTS Max frequency (days) | | 0.998761 | DISCARD |
| 2-year summary (Drug prescriptions) - ADRENERGIC AGONISTS Min frequency (days) | | 0.998761 | DISCARD |
| 2-year summary (Drug prescriptions) - IMMUNOMODULATING AGENTS Average frequency (days) | | 0.998761 | DISCARD |
| 2-year summary (Drug prescriptions) - IMMUNOMODULATING AGENTS Max frequency (days) | | 0.998761 | DISCARD |
| 2-year summary (Drug prescriptions) - IMMUNOMODULATING AGENTS Min frequency (days) | | 0.998761 | DISCARD |
| 1-year summary (Drug prescriptions) - CHOLINERGIC AGONISTS Days since first prescription | | 0.998789 | DISCARD |
| 1-year summary (Drug prescriptions) - CHOLINERGIC AGONISTS Days since last prescription | | 0.998789 | DISCARD |
| 1-year summary (Drug prescriptions) - CHOLINERGIC AGONISTS Prescription count | | 0.998789 | DISCARD |
| 1-year summary (Drug prescriptions) - CLASS III ANTIARRHYTHMIC DRUGS Days since first prescription | | 0.998789 | DISCARD |
| 1-year summary (Drug prescriptions) - CLASS III ANTIARRHYTHMIC DRUGS Days since last prescription | | 0.998789 | DISCARD |
| 1-year summary (Drug prescriptions) - CLASS III ANTIARRHYTHMIC DRUGS Prescription count | | 0.998789 | DISCARD |
| 2-year summary (Hospitalizations) - Class - Diseases of the eye and adnexa (H00-H59) | | 0.998789 | DISCARD |
| 1-year summary (Drug prescriptions) - ANTI-HERPESVIRUS INFECTION Average frequency (days) | | 0.998816 | DISCARD |
| 1-year summary (Drug prescriptions) - ANTI-HERPESVIRUS INFECTION Max frequency (days) | | 0.998816 | DISCARD |
| 1-year summary (Drug prescriptions) - ANTI-HERPESVIRUS INFECTION Min frequency (days) | | 0.998816 | DISCARD |
| 2-year summary (Drug prescriptions) - ANTACIDS & ADSORBENTS Average frequency (days) | | 0.998816 | DISCARD |
| 2-year summary (Drug prescriptions) - ANTACIDS & ADSORBENTS Max frequency (days) | | 0.998816 | DISCARD |
| 2-year summary (Drug prescriptions) - ANTACIDS & ADSORBENTS Min frequency (days) | | 0.998816 | DISCARD |
| 2-year summary (Drug prescriptions) - ANTIGLAUCOMA AGENTS Average frequency (days) | | 0.998816 | DISCARD |
| 2-year summary (Drug prescriptions) - ANTIGLAUCOMA AGENTS Max frequency (days) | | 0.998816 | DISCARD |
| 2-year summary (Drug prescriptions) - ANTIGLAUCOMA AGENTS Min frequency (days) | | 0.998816 | DISCARD |
| 2-year summary (Drug prescriptions) - POTASSIUM-REMOVING RESINS Average frequency (days) | | 0.998816 | DISCARD |
| 2-year summary (Drug prescriptions) - POTASSIUM-REMOVING RESINS Max frequency (days) | | 0.998816 | DISCARD |
| 2-year summary (Drug prescriptions) - POTASSIUM-REMOVING RESINS Min frequency (days) | | 0.998816 | DISCARD |
| 1-year summary (Drug prescriptions) - ANTICOAGULANTS MISCELLANEOUS Days since first prescription | | 0.998844 | DISCARD |
| 1-year summary (Drug prescriptions) - ANTICOAGULANTS MISCELLANEOUS Days since last prescription | | 0.998844 | DISCARD |
| 1-year summary (Drug prescriptions) - ANTICOAGULANTS MISCELLANEOUS Prescription count | | 0.998844 | DISCARD |
| 1-year summary (Drug prescriptions) - AROMATASE INHIBITORS Average frequency (days) | | 0.998844 | DISCARD |
| 1-year summary (Drug prescriptions) - AROMATASE INHIBITORS Days since first prescription | | 0.998844 | DISCARD |

| | | | |
|---|---|---|---|
| 1-year summary (Drug prescriptions) - AROMATASE INHIBITORS Days since last prescription | | 0.998844 | DISCARD |
| 1-year summary (Drug prescriptions) - AROMATASE INHIBITORS Max frequency (days) | | 0.998844 | DISCARD |
| 1-year summary (Drug prescriptions) - AROMATASE INHIBITORS Min frequency (days) | | 0.998844 | DISCARD |
| 1-year summary (Drug prescriptions) - AROMATASE INHIBITORS Prescription count | | 0.998844 | DISCARD |
| 1-year summary (Drug prescriptions) - HYDANTOIN DERIVATIVES Average frequency (days) | | 0.998844 | DISCARD |
| 1-year summary (Drug prescriptions) - HYDANTOIN DERIVATIVES Max frequency (days) | | 0.998844 | DISCARD |
| 1-year summary (Drug prescriptions) - HYDANTOIN DERIVATIVES Min frequency (days) | | 0.998844 | DISCARD |
| 1-year summary (Doctor visits) - GENETICS | | 0.998844 | DISCARD |
| 2-year summary (Drug prescriptions) - ANION EXCHANGE RESINS Days since first prescription | | 0.998844 | DISCARD |
| 2-year summary (Drug prescriptions) - ANION EXCHANGE RESINS Days since last prescription | | 0.998844 | DISCARD |
| 2-year summary (Drug prescriptions) - ANION EXCHANGE RESINS Prescription count | | 0.998844 | DISCARD |
| 2-year summary (Drug prescriptions) - CENTRALLY ACTING ANTIADRENERGIC Average frequency (days) | | 0.998844 | DISCARD |
| 2-year summary (Drug prescriptions) - CENTRALLY ACTING ANTIADRENERGIC Max frequency (days) | | 0.998844 | DISCARD |
| 2-year summary (Drug prescriptions) - CENTRALLY ACTING ANTIADRENERGIC Min frequency (days) | | 0.998844 | DISCARD |
| 2-year summary (Drug prescriptions) - HEMATOPOIETIC AGENTS Days since first prescription | | 0.998844 | DISCARD |
| 2-year summary (Drug prescriptions) - HEMATOPOIETIC AGENTS Days since last prescription | | 0.998844 | DISCARD |
| 2-year summary (Drug prescriptions) - HEMATOPOIETIC AGENTS Prescription count | | 0.998844 | DISCARD |
| 2-year summary (Drug prescriptions) - NON-SELECTIVE BETA-ADRENERGIC AGONISTS Days since first prescription | | 0.998844 | DISCARD |
| 2-year summary (Drug prescriptions) - NON-SELECTIVE BETA-ADRENERGIC AGONISTS Days since last prescription | | 0.998844 | DISCARD |
| 2-year summary (Drug prescriptions) - NON-SELECTIVE BETA-ADRENERGIC AGONISTS Prescription count | | 0.998844 | DISCARD |
| 1-year summary (Drug prescriptions) - BIOLOGIC RESPONSE MODIFYING AGENTS Days since first prescription | | 0.998871 | DISCARD |
| 1-year summary (Drug prescriptions) - BIOLOGIC RESPONSE MODIFYING AGENTS Days since last prescription | | 0.998871 | DISCARD |
| 1-year summary (Drug prescriptions) - BIOLOGIC RESPONSE MODIFYING AGENTS Prescription count | | 0.998871 | DISCARD |
| 1-year summary (Drug prescriptions) - CENTRALLY ACTING ANTIADRENERGIC Days since first prescription | | 0.998871 | DISCARD |
| 1-year summary (Drug prescriptions) - CENTRALLY ACTING ANTIADRENERGIC Days since last prescription | | 0.998871 | DISCARD |
| 1-year summary (Drug prescriptions) - CENTRALLY ACTING ANTIADRENERGIC Prescription count | | 0.998871 | DISCARD |
| 1-year summary (Drug prescriptions) - LUTEINIZING HORMONE-RELEASING HORMONE (LHRH) ANALOG Average frequency (da | | 0.998871 | DISCARD |
| 1-year summary (Drug prescriptions) - LUTEINIZING HORMONE-RELEASING HORMONE (LHRH) ANALOG Max frequency (days) | | 0.998871 | DISCARD |
| 1-year summary (Drug prescriptions) - LUTEINIZING HORMONE-RELEASING HORMONE (LHRH) ANALOG Min frequency (days) | | 0.998871 | DISCARD |
| 2-year summary (Drug prescriptions) - PSYCHOTHERAPEUTIC AGENTS Days since first prescription | | 0.998871 | DISCARD |
| 2-year summary (Drug prescriptions) - PSYCHOTHERAPEUTIC AGENTS Days since last prescription | | 0.998871 | DISCARD |
| 2-year summary (Drug prescriptions) - PSYCHOTHERAPEUTIC AGENTS Prescription count | | 0.998871 | DISCARD |
| 1-year summary (Drug prescriptions) - BIOLOGIC RESPONSE MODIFYING AGENTS Average frequency (days) | | 0.998899 | DISCARD |
| 1-year summary (Drug prescriptions) - BIOLOGIC RESPONSE MODIFYING AGENTS Max frequency (days) | | 0.998899 | DISCARD |
| 1-year summary (Drug prescriptions) - BIOLOGIC RESPONSE MODIFYING AGENTS Min frequency (days) | | 0.998899 | DISCARD |
| 1-year summary (Drug prescriptions) - TESTOSTERONES AND RELATED HORMONES Days since first prescription | | 0.998899 | DISCARD |
| 1-year summary (Drug prescriptions) - TESTOSTERONES AND RELATED HORMONES Days since last prescription | | 0.998899 | DISCARD |

| | | | |
|---|---|---|---|
| 1-year summary (Drug prescriptions) - TESTOSTERONES AND RELATED HORMONES Prescription count | | 0.998899 | DISCARD |
| 1-year summary (Drug prescriptions) - IMMUNOMODULATING AGENTS Days since first prescription | | 0.998926 | DISCARD |
| 1-year summary (Drug prescriptions) - IMMUNOMODULATING AGENTS Days since last prescription | | 0.998926 | DISCARD |
| 1-year summary (Drug prescriptions) - IMMUNOMODULATING AGENTS Prescription count | | 0.998926 | DISCARD |
| 2-year summary (Drug prescriptions) - CHOLINERGIC AGONISTS Average frequency (days) | | 0.998926 | DISCARD |
| 2-year summary (Drug prescriptions) - CHOLINERGIC AGONISTS Max frequency (days) | | 0.998926 | DISCARD |
| 2-year summary (Drug prescriptions) - CHOLINERGIC AGONISTS Min frequency (days) | | 0.998926 | DISCARD |
| 1-year summary (Drug prescriptions) - CLASS III ANTIARRHYTHMIC DRUGS Average frequency (days) | | 0.998954 | DISCARD |
| 1-year summary (Drug prescriptions) - CLASS III ANTIARRHYTHMIC DRUGS Max frequency (days) | | 0.998954 | DISCARD |
| 1-year summary (Drug prescriptions) - CLASS III ANTIARRHYTHMIC DRUGS Min frequency (days) | | 0.998954 | DISCARD |
| 1-year summary (Drug prescriptions) - LOW MOLECULAR WEIGHT HEPARINS (LMWH) Days since first prescription | | 0.998954 | DISCARD |
| 1-year summary (Drug prescriptions) - LOW MOLECULAR WEIGHT HEPARINS (LMWH) Days since last prescription | | 0.998954 | DISCARD |
| 1-year summary (Drug prescriptions) - LOW MOLECULAR WEIGHT HEPARINS (LMWH) Prescription count | | 0.998954 | DISCARD |
| 2-year summary (Drug prescriptions) - ANTIMANIC AGENTS Days since first prescription | | 0.998954 | DISCARD |
| 2-year summary (Drug prescriptions) - ANTIMANIC AGENTS Days since last prescription | | 0.998954 | DISCARD |
| 2-year summary (Drug prescriptions) - ANTIMANIC AGENTS Prescription count | | 0.998954 | DISCARD |
| 1-year summary (Drug prescriptions) - ANTIGLAUCOMA AGENTS Days since first prescription | | 0.998981 | DISCARD |
| 1-year summary (Drug prescriptions) - ANTIGLAUCOMA AGENTS Days since last prescription | | 0.998981 | DISCARD |
| 1-year summary (Drug prescriptions) - ANTIGLAUCOMA AGENTS Prescription count | | 0.998981 | DISCARD |
| 1-year summary (Drug prescriptions) - IMMUNOMODULATING AGENTS Average frequency (days) | | 0.998981 | DISCARD |
| 1-year summary (Drug prescriptions) - IMMUNOMODULATING AGENTS Max frequency (days) | | 0.998981 | DISCARD |
| 1-year summary (Drug prescriptions) - IMMUNOMODULATING AGENTS Min frequency (days) | | 0.998981 | DISCARD |
| 2-year summary (Drug prescriptions) - ANTIANDROGENS Days since first prescription | | 0.998981 | DISCARD |
| 2-year summary (Drug prescriptions) - ANTIANDROGENS Days since last prescription | | 0.998981 | DISCARD |
| 2-year summary (Drug prescriptions) - ANTIANDROGENS Prescription count | | 0.998981 | DISCARD |
| 2-year summary (Drug prescriptions) - SEX HORMONES Days since first prescription | | 0.998981 | DISCARD |
| 2-year summary (Drug prescriptions) - SEX HORMONES Days since last prescription | | 0.998981 | DISCARD |
| 2-year summary (Drug prescriptions) - SEX HORMONES Prescription count | | 0.998981 | DISCARD |
| 2-year summary (Drug prescriptions) - TOPICAL IMMUNOSUPRESSIVE AGENTS Average frequency (days) | | 0.998981 | DISCARD |
| 2-year summary (Drug prescriptions) - TOPICAL IMMUNOSUPRESSIVE AGENTS Max frequency (days) | | 0.998981 | DISCARD |
| 2-year summary (Drug prescriptions) - TOPICAL IMMUNOSUPRESSIVE AGENTS Min frequency (days) | | 0.998981 | DISCARD |
| 1-year summary (Drug prescriptions) - ANTACIDS & ADSORBENTS Average frequency (days) | | 0.999009 | DISCARD |
| 1-year summary (Drug prescriptions) - ANTACIDS & ADSORBENTS Max frequency (days) | | 0.999009 | DISCARD |
| 1-year summary (Drug prescriptions) - ANTACIDS & ADSORBENTS Min frequency (days) | | 0.999009 | DISCARD |
| 1-year summary (Drug prescriptions) - AMYNOGLYCOSIDES Average frequency (days) | | 0.999036 | DISCARD |
| 1-year summary (Drug prescriptions) - AMYNOGLYCOSIDES Max frequency (days) | | 0.999036 | DISCARD |
| 1-year summary (Drug prescriptions) - AMYNOGLYCOSIDES Min frequency (days) | | 0.999036 | DISCARD |

| | | |
|---|---|---|
| 1-year summary (Drug prescriptions) - ANTI-HEMORRHOIDAL AGENTS WITHOUT CORTICOSTEROIDS Average frequency (days) | 0.999064 | DISCARD |
| 1-year summary (Drug prescriptions) - ANTI-HEMORRHOIDAL AGENTS WITHOUT CORTICOSTEROIDS Max frequency (days) | 0.999064 | DISCARD |
| 1-year summary (Drug prescriptions) - ANTI-HEMORRHOIDAL AGENTS WITHOUT CORTICOSTEROIDS Min frequency (days) | 0.999064 | DISCARD |
| 1-year summary (Drug prescriptions) - ANTIVIRAL AGENTS - INFLUENZA VIRUS SPECIFIC Average frequency (days) | 0.999064 | DISCARD |
| 1-year summary (Drug prescriptions) - ANTIVIRAL AGENTS - INFLUENZA VIRUS SPECIFIC Max frequency (days) | 0.999064 | DISCARD |
| 1-year summary (Drug prescriptions) - ANTIVIRAL AGENTS - INFLUENZA VIRUS SPECIFIC Min frequency (days) | 0.999064 | DISCARD |
| 1-year summary (Drug prescriptions) - PARASITICIDES Days since first prescription | 0.999064 | DISCARD |
| 1-year summary (Drug prescriptions) - PARASITICIDES Days since last prescription | 0.999064 | DISCARD |
| 1-year summary (Drug prescriptions) - PARASITICIDES Prescription count | 0.999064 | DISCARD |
| 2-year summary (Drug prescriptions) - ANTIMANIC AGENTS Average frequency (days) | 0.999064 | DISCARD |
| 2-year summary (Drug prescriptions) - ANTIMANIC AGENTS Max frequency (days) | 0.999064 | DISCARD |
| 2-year summary (Drug prescriptions) - ANTIMANIC AGENTS Min frequency (days) | 0.999064 | DISCARD |
| 2-year summary (Drug prescriptions) - LOW MOLECULAR WEIGHT HEPARINS (LMWH) Average frequency (days) | 0.999064 | DISCARD |
| 2-year summary (Drug prescriptions) - LOW MOLECULAR WEIGHT HEPARINS (LMWH) Max frequency (days) | 0.999064 | DISCARD |
| 2-year summary (Drug prescriptions) - LOW MOLECULAR WEIGHT HEPARINS (LMWH) Min frequency (days) | 0.999064 | DISCARD |
| 2-year summary (Drug prescriptions) - PSYCHOTHERAPEUTIC AGENTS Average frequency (days) | 0.999064 | DISCARD |
| 2-year summary (Drug prescriptions) - PSYCHOTHERAPEUTIC AGENTS Max frequency (days) | 0.999064 | DISCARD |
| 2-year summary (Drug prescriptions) - PSYCHOTHERAPEUTIC AGENTS Min frequency (days) | 0.999064 | DISCARD |
| 1-year summary (Drug prescriptions) - ANTICOAGULANTS MISCELLANEOUS Average frequency (days) | 0.999091 | DISCARD |
| 1-year summary (Drug prescriptions) - ANTICOAGULANTS MISCELLANEOUS Max frequency (days) | 0.999091 | DISCARD |
| 1-year summary (Drug prescriptions) - ANTICOAGULANTS MISCELLANEOUS Min frequency (days) | 0.999091 | DISCARD |
| 1-year summary (Drug prescriptions) - ANTIGLAUCOMA AGENTS Average frequency (days) | 0.999091 | DISCARD |
| 1-year summary (Drug prescriptions) - ANTIGLAUCOMA AGENTS Max frequency (days) | 0.999091 | DISCARD |
| 1-year summary (Drug prescriptions) - ANTIGLAUCOMA AGENTS Min frequency (days) | 0.999091 | DISCARD |
| 1-year summary (Drug prescriptions) - CHOLINERGIC AGONISTS Average frequency (days) | 0.999091 | DISCARD |
| 1-year summary (Drug prescriptions) - CHOLINERGIC AGONISTS Max frequency (days) | 0.999091 | DISCARD |
| 1-year summary (Drug prescriptions) - CHOLINERGIC AGONISTS Min frequency (days) | 0.999091 | DISCARD |
| 2-year summary (Drug prescriptions) - MISCELLANEOUS SKIN AND MUCOUS MEMBRANE AGENTS Average frequency (days) | 0.999091 | DISCARD |
| 2-year summary (Drug prescriptions) - MISCELLANEOUS SKIN AND MUCOUS MEMBRANE AGENTS Max frequency (days) | 0.999091 | DISCARD |
| 2-year summary (Drug prescriptions) - MISCELLANEOUS SKIN AND MUCOUS MEMBRANE AGENTS Min frequency (days) | 0.999091 | DISCARD |
| 2-year summary (Doctor visits) - ORAL PATHOLOGY | 0.999091 | DISCARD |
| 1-year summary (Drug prescriptions) - ADRENERGIC AGONISTS Average frequency (days) | 0.999119 | DISCARD |
| 1-year summary (Drug prescriptions) - ADRENERGIC AGONISTS Max frequency (days) | 0.999119 | DISCARD |
| 1-year summary (Drug prescriptions) - ADRENERGIC AGONISTS Min frequency (days) | 0.999119 | DISCARD |
| 2-year summary (Hospitalizations) - Class - Diseases of the ear and mastoid process (H60-H95) | 0.999119 | DISCARD |
| 2-year summary (Drug prescriptions) - CARDIAC DRUGS OTHER Days since first prescription | 0.999119 | DISCARD |
| 2-year summary (Drug prescriptions) - CARDIAC DRUGS OTHER Days since last prescription | 0.999119 | DISCARD |

| | | | |
|---|---|---|---|
| 2-year summary (Drug prescriptions) - CARDIAC DRUGS OTHER Prescription count | | 0.999119 | DISCARD |
| 1-year summary (Drug prescriptions) - ANTIMANIC AGENTS Average frequency (days) | | 0.999147 | DISCARD |
| 1-year summary (Drug prescriptions) - ANTIMANIC AGENTS Days since first prescription | | 0.999147 | DISCARD |
| 1-year summary (Drug prescriptions) - ANTIMANIC AGENTS Days since last prescription | | 0.999147 | DISCARD |
| 1-year summary (Drug prescriptions) - ANTIMANIC AGENTS Max frequency (days) | | 0.999147 | DISCARD |
| 1-year summary (Drug prescriptions) - ANTIMANIC AGENTS Min frequency (days) | | 0.999147 | DISCARD |
| 1-year summary (Drug prescriptions) - ANTIMANIC AGENTS Prescription count | | 0.999147 | DISCARD |
| 1-year summary (Drug prescriptions) - NON-STEROIDAL ANTI-INFLAMMATORY Average frequency (days) | | 0.999147 | DISCARD |
| 1-year summary (Drug prescriptions) - NON-STEROIDAL ANTI-INFLAMMATORY Max frequency (days) | | 0.999147 | DISCARD |
| 1-year summary (Drug prescriptions) - NON-STEROIDAL ANTI-INFLAMMATORY Min frequency (days) | | 0.999147 | DISCARD |
| 2-year summary (Drug prescriptions) - ANTINEOPLASTIC AGENTS Average frequency (days) | | 0.999147 | DISCARD |
| 2-year summary (Drug prescriptions) - ANTINEOPLASTIC AGENTS Days since first prescription | | 0.999147 | DISCARD |
| 2-year summary (Drug prescriptions) - ANTINEOPLASTIC AGENTS Days since last prescription | | 0.999147 | DISCARD |
| 2-year summary (Drug prescriptions) - ANTINEOPLASTIC AGENTS Max frequency (days) | | 0.999147 | DISCARD |
| 2-year summary (Drug prescriptions) - ANTINEOPLASTIC AGENTS Min frequency (days) | | 0.999147 | DISCARD |
| 2-year summary (Drug prescriptions) - ANTINEOPLASTIC AGENTS Prescription count | | 0.999147 | DISCARD |
| 1-year summary (Drug prescriptions) - POTASSIUM-REMOVING RESINS Average frequency (days) | | 0.999174 | DISCARD |
| 1-year summary (Drug prescriptions) - POTASSIUM-REMOVING RESINS Max frequency (days) | | 0.999174 | DISCARD |
| 1-year summary (Drug prescriptions) - POTASSIUM-REMOVING RESINS Min frequency (days) | | 0.999174 | DISCARD |
| 1-year summary (Drug prescriptions) - SMOKING CESSATION AIDS Days since first prescription | | 0.999174 | DISCARD |
| 1-year summary (Drug prescriptions) - SMOKING CESSATION AIDS Days since last prescription | | 0.999174 | DISCARD |
| 1-year summary (Drug prescriptions) - SMOKING CESSATION AIDS Prescription count | | 0.999174 | DISCARD |
| 2-year summary (Drug prescriptions) - CARDIAC DRUGS OTHER Average frequency (days) | | 0.999174 | DISCARD |
| 2-year summary (Drug prescriptions) - CARDIAC DRUGS OTHER Max frequency (days) | | 0.999174 | DISCARD |
| 2-year summary (Drug prescriptions) - CARDIAC DRUGS OTHER Min frequency (days) | | 0.999174 | DISCARD |
| 2-year summary (Drug prescriptions) - NON-DEPOLARIZING AGENTS Days since first prescription | | 0.999174 | DISCARD |
| 2-year summary (Drug prescriptions) - NON-DEPOLARIZING AGENTS Days since last prescription | | 0.999174 | DISCARD |
| 2-year summary (Drug prescriptions) - NON-DEPOLARIZING AGENTS Prescription count | | 0.999174 | DISCARD |
| 1-year summary (Drug prescriptions) - CENTRALLY ACTING ANTIADRENERGIC Average frequency (days) | | 0.999202 | DISCARD |
| 1-year summary (Drug prescriptions) - CENTRALLY ACTING ANTIADRENERGIC Max frequency (days) | | 0.999202 | DISCARD |
| 1-year summary (Drug prescriptions) - CENTRALLY ACTING ANTIADRENERGIC Min frequency (days) | | 0.999202 | DISCARD |
| 1-year summary (Drug prescriptions) - HEMATOPOIETIC AGENTS Days since first prescription | | 0.999202 | DISCARD |
| 1-year summary (Drug prescriptions) - HEMATOPOIETIC AGENTS Days since last prescription | | 0.999202 | DISCARD |
| 1-year summary (Drug prescriptions) - HEMATOPOIETIC AGENTS Prescription count | | 0.999202 | DISCARD |
| 1-year summary (Drug prescriptions) - MISCELLANEOUS LOCAL ANTI-INFECTIVES Days since first prescription | | 0.999202 | DISCARD |
| 1-year summary (Drug prescriptions) - MISCELLANEOUS LOCAL ANTI-INFECTIVES Days since last prescription | | 0.999202 | DISCARD |
| 1-year summary (Drug prescriptions) - MISCELLANEOUS LOCAL ANTI-INFECTIVES Prescription count | | 0.999202 | DISCARD |

| | | | |
|---|---|---|---|
| 2-year summary (Drug prescriptions) - ANTIVIRALS Average frequency (days) | | 0.999202 | DISCARD |
| 2-year summary (Drug prescriptions) - ANTIVIRALS Max frequency (days) | | 0.999202 | DISCARD |
| 2-year summary (Drug prescriptions) - ANTIVIRALS Min frequency (days) | | 0.999202 | DISCARD |
| 2-year summary (Drug prescriptions) - HEMATOPOIETIC AGENTS Average frequency (days) | | 0.999202 | DISCARD |
| 2-year summary (Drug prescriptions) - HEMATOPOIETIC AGENTS Max frequency (days) | | 0.999202 | DISCARD |
| 2-year summary (Drug prescriptions) - HEMATOPOIETIC AGENTS Min frequency (days) | | 0.999202 | DISCARD |
| 1-year summary (Drug prescriptions) - CARDIAC DRUGS OTHER Days since first prescription | | 0.999229 | DISCARD |
| 1-year summary (Drug prescriptions) - CARDIAC DRUGS OTHER Days since last prescription | | 0.999229 | DISCARD |
| 1-year summary (Drug prescriptions) - CARDIAC DRUGS OTHER Prescription count | | 0.999229 | DISCARD |
| 1-year summary (Drug prescriptions) - CORTICOSTEROIDS WITH OTHER COMBINATIONS Average frequency (days) | | 0.999229 | DISCARD |
| 1-year summary (Drug prescriptions) - CORTICOSTEROIDS WITH OTHER COMBINATIONS Max frequency (days) | | 0.999229 | DISCARD |
| 1-year summary (Drug prescriptions) - CORTICOSTEROIDS WITH OTHER COMBINATIONS Min frequency (days) | | 0.999229 | DISCARD |
| 1-year summary (Drug prescriptions) - TESTOSTERONES AND RELATED HORMONES Average frequency (days) | | 0.999229 | DISCARD |
| 1-year summary (Drug prescriptions) - TESTOSTERONES AND RELATED HORMONES Max frequency (days) | | 0.999229 | DISCARD |
| 1-year summary (Drug prescriptions) - TESTOSTERONES AND RELATED HORMONES Min frequency (days) | | 0.999229 | DISCARD |
| 2-year summary (Drug prescriptions) - NUCLEOSIDE REVERSE TRANSCRIPTASE INHIBITOR Days since first prescription | | 0.999229 | DISCARD |
| 2-year summary (Drug prescriptions) - NUCLEOSIDE REVERSE TRANSCRIPTASE INHIBITOR Days since last prescription | | 0.999229 | DISCARD |
| 2-year summary (Drug prescriptions) - NUCLEOSIDE REVERSE TRANSCRIPTASE INHIBITOR Prescription count | | 0.999229 | DISCARD |
| 1-year summary (Hospitalizations) - Class - Congenital malformations, deformations and chromosomal abnormalities (Q00-Q99) | | 0.999257 | DISCARD |
| 2-year summary (Drug prescriptions) - BARBITURATES AND DERIVATIVES Days since first prescription | | 0.999257 | DISCARD |
| 2-year summary (Drug prescriptions) - BARBITURATES AND DERIVATIVES Days since last prescription | | 0.999257 | DISCARD |
| 2-year summary (Drug prescriptions) - BARBITURATES AND DERIVATIVES Prescription count | | 0.999257 | DISCARD |
| 1-year summary (Drug prescriptions) - ANTINEOPLASTIC AGENTS Average frequency (days) | | 0.999284 | DISCARD |
| 1-year summary (Drug prescriptions) - ANTINEOPLASTIC AGENTS Days since first prescription | | 0.999284 | DISCARD |
| 1-year summary (Drug prescriptions) - ANTINEOPLASTIC AGENTS Days since last prescription | | 0.999284 | DISCARD |
| 1-year summary (Drug prescriptions) - ANTINEOPLASTIC AGENTS Max frequency (days) | | 0.999284 | DISCARD |
| 1-year summary (Drug prescriptions) - ANTINEOPLASTIC AGENTS Min frequency (days) | | 0.999284 | DISCARD |
| 1-year summary (Drug prescriptions) - ANTINEOPLASTIC AGENTS Prescription count | | 0.999284 | DISCARD |
| 1-year summary (Drug prescriptions) - CARDIAC DRUGS OTHER Average frequency (days) | | 0.999284 | DISCARD |
| 1-year summary (Drug prescriptions) - CARDIAC DRUGS OTHER Max frequency (days) | | 0.999284 | DISCARD |
| 1-year summary (Drug prescriptions) - CARDIAC DRUGS OTHER Min frequency (days) | | 0.999284 | DISCARD |
| 1-year summary (Drug prescriptions) - NON-SELECTIVE BETA-ADRENERGIC AGONISTS Days since first prescription | | 0.999284 | DISCARD |
| 1-year summary (Drug prescriptions) - NON-SELECTIVE BETA-ADRENERGIC AGONISTS Days since last prescription | | 0.999284 | DISCARD |
| 1-year summary (Drug prescriptions) - NON-SELECTIVE BETA-ADRENERGIC AGONISTS Prescription count | | 0.999284 | DISCARD |
| 1-year summary (Drug prescriptions) - VASOPRESSORS Average frequency (days) | | 0.999284 | DISCARD |
| 1-year summary (Drug prescriptions) - VASOPRESSORS Max frequency (days) | | 0.999284 | DISCARD |
| 1-year summary (Drug prescriptions) - VASOPRESSORS Min frequency (days) | | 0.999284 | DISCARD |

| | | |
|---|---|---|
| 2-year summary (Drug prescriptions) - PARASITICIDES Average frequency (days) | 0.999284 | DISCARD |
| 2-year summary (Drug prescriptions) - PARASITICIDES Max frequency (days) | 0.999284 | DISCARD |
| 2-year summary (Drug prescriptions) - PARASITICIDES Min frequency (days) | 0.999284 | DISCARD |
| 2-year summary (Drug prescriptions) - WOUND CARE AND SCAR MANAGEMENT AGENTS Days since first prescription | 0.999284 | DISCARD |
| 2-year summary (Drug prescriptions) - WOUND CARE AND SCAR MANAGEMENT AGENTS Days since last prescription | 0.999284 | DISCARD |
| 2-year summary (Drug prescriptions) - WOUND CARE AND SCAR MANAGEMENT AGENTS Prescription count | 0.999284 | DISCARD |
| 1-year summary (Hospitalizations) - Class - Diseases of the eye and adnexa (H00-H59) | 0.999312 | DISCARD |
| 1-year summary (Drug prescriptions) - LINCOSAMIDES Average frequency (days) | 0.999312 | DISCARD |
| 1-year summary (Drug prescriptions) - LINCOSAMIDES Max frequency (days) | 0.999312 | DISCARD |
| 1-year summary (Drug prescriptions) - LINCOSAMIDES Min frequency (days) | 0.999312 | DISCARD |
| 1-year summary (Drug prescriptions) - LOW MOLECULAR WEIGHT HEPARINS (LMWH) Average frequency (days) | 0.999312 | DISCARD |
| 1-year summary (Drug prescriptions) - LOW MOLECULAR WEIGHT HEPARINS (LMWH) Max frequency (days) | 0.999312 | DISCARD |
| 1-year summary (Drug prescriptions) - LOW MOLECULAR WEIGHT HEPARINS (LMWH) Min frequency (days) | 0.999312 | DISCARD |
| 2-year summary (Drug prescriptions) - ANTIESTROGENS Days since first prescription | 0.999312 | DISCARD |
| 2-year summary (Drug prescriptions) - ANTIESTROGENS Days since last prescription | 0.999312 | DISCARD |
| 2-year summary (Drug prescriptions) - ANTIESTROGENS Prescription count | 0.999312 | DISCARD |
| 2-year summary (Drug prescriptions) - BARBITURATES AND DERIVATIVES Average frequency (days) | 0.999312 | DISCARD |
| 2-year summary (Drug prescriptions) - BARBITURATES AND DERIVATIVES Max frequency (days) | 0.999312 | DISCARD |
| 2-year summary (Drug prescriptions) - BARBITURATES AND DERIVATIVES Min frequency (days) | 0.999312 | DISCARD |
| 2-year summary (Drug prescriptions) - HYPERTHYROIDISM THERAPY Days since first prescription | 0.999312 | DISCARD |
| 2-year summary (Drug prescriptions) - HYPERTHYROIDISM THERAPY Days since last prescription | 0.999312 | DISCARD |
| 2-year summary (Drug prescriptions) - HYPERTHYROIDISM THERAPY Prescription count | 0.999312 | DISCARD |
| 1-year summary (Drug prescriptions) - HYPERTHYROIDISM THERAPY Days since first prescription | 0.999339 | DISCARD |
| 1-year summary (Drug prescriptions) - HYPERTHYROIDISM THERAPY Days since last prescription | 0.999339 | DISCARD |
| 1-year summary (Drug prescriptions) - HYPERTHYROIDISM THERAPY Prescription count | 0.999339 | DISCARD |
| 1-year summary (Drug prescriptions) - OTHER HORMONAL CONTRACEPTIVES Days since first prescription | 0.999339 | DISCARD |
| 1-year summary (Drug prescriptions) - OTHER HORMONAL CONTRACEPTIVES Days since last prescription | 0.999339 | DISCARD |
| 1-year summary (Drug prescriptions) - OTHER HORMONAL CONTRACEPTIVES Prescription count | 0.999339 | DISCARD |
| 2-year summary (Drug prescriptions) - HYPERTHYROIDISM THERAPY Average frequency (days) | 0.999339 | DISCARD |
| 2-year summary (Drug prescriptions) - HYPERTHYROIDISM THERAPY Max frequency (days) | 0.999339 | DISCARD |
| 2-year summary (Drug prescriptions) - HYPERTHYROIDISM THERAPY Min frequency (days) | 0.999339 | DISCARD |
| 1-year summary (Drug prescriptions) - BARBITURATES AND DERIVATIVES Average frequency (days) | 0.999367 | DISCARD |
| 1-year summary (Drug prescriptions) - BARBITURATES AND DERIVATIVES Days since first prescription | 0.999367 | DISCARD |
| 1-year summary (Drug prescriptions) - BARBITURATES AND DERIVATIVES Days since last prescription | 0.999367 | DISCARD |
| 1-year summary (Drug prescriptions) - BARBITURATES AND DERIVATIVES Max frequency (days) | 0.999367 | DISCARD |
| 1-year summary (Drug prescriptions) - BARBITURATES AND DERIVATIVES Min frequency (days) | 0.999367 | DISCARD |
| 1-year summary (Drug prescriptions) - BARBITURATES AND DERIVATIVES Prescription count | 0.999367 | DISCARD |

| | | | |
|---|---|---|---|
| 1-year summary (Drug prescriptions) - PSYCHOTHERAPEUTIC AGENTS Days since first prescription | | 0.999367 | DISCARD |
| 1-year summary (Drug prescriptions) - PSYCHOTHERAPEUTIC AGENTS Days since last prescription | | 0.999367 | DISCARD |
| 1-year summary (Drug prescriptions) - PSYCHOTHERAPEUTIC AGENTS Prescription count | | 0.999367 | DISCARD |
| 2-year summary (Drug prescriptions) - ANTIESTROGENS Average frequency (days) | | 0.999367 | DISCARD |
| 2-year summary (Drug prescriptions) - ANTIESTROGENS Max frequency (days) | | 0.999367 | DISCARD |
| 2-year summary (Drug prescriptions) - ANTIESTROGENS Min frequency (days) | | 0.999367 | DISCARD |
| 2-year summary (Drug prescriptions) - MYDRIATICS Days since first prescription | | 0.999367 | DISCARD |
| 2-year summary (Drug prescriptions) - MYDRIATICS Days since last prescription | | 0.999367 | DISCARD |
| 2-year summary (Drug prescriptions) - MYDRIATICS Prescription count | | 0.999367 | DISCARD |
| 2-year summary (Drug prescriptions) - NUCLEOSIDE REVERSE TRANSCRIPTASE INHIBITOR Average frequency (days) | | 0.999367 | DISCARD |
| 2-year summary (Drug prescriptions) - NUCLEOSIDE REVERSE TRANSCRIPTASE INHIBITOR Max frequency (days) | | 0.999367 | DISCARD |
| 2-year summary (Drug prescriptions) - NUCLEOSIDE REVERSE TRANSCRIPTASE INHIBITOR Min frequency (days) | | 0.999367 | DISCARD |
| 1-year summary (Drug prescriptions) - HEMATOPOIETIC AGENTS Average frequency (days) | | 0.999394 | DISCARD |
| 1-year summary (Drug prescriptions) - HEMATOPOIETIC AGENTS Max frequency (days) | | 0.999394 | DISCARD |
| 1-year summary (Drug prescriptions) - HEMATOPOIETIC AGENTS Min frequency (days) | | 0.999394 | DISCARD |
| 1-year summary (Drug prescriptions) - NON-DEPOLARIZING AGENTS Days since first prescription | | 0.999394 | DISCARD |
| 1-year summary (Drug prescriptions) - NON-DEPOLARIZING AGENTS Days since last prescription | | 0.999394 | DISCARD |
| 1-year summary (Drug prescriptions) - NON-DEPOLARIZING AGENTS Prescription count | | 0.999394 | DISCARD |
| 1-year summary (Doctor visits) - ORAL SURGERY | | 0.999394 | DISCARD |
| 1-year summary (Laboratory tests) - number of tests for GLUCOSE^1H_POST_75_G_GLUCOSE_PO | | 0.999394 | DISCARD |
| 1-year summary (Laboratory tests) - days since earliest test for GLUCOSE^1H_POST_75_G_GLUCOSE_PO | | 0.999394 | DISCARD |
| 1-year summary (Laboratory tests) - days since latest test for GLUCOSE^1H_POST_75_G_GLUCOSE_PO | | 0.999394 | DISCARD |
| 1-year summary (Laboratory tests) - number of high values of GLUCOSE^1H_POST_75_G_GLUCOSE_PO | | 0.999394 | DISCARD |
| 1-year summary (Laboratory tests) - highest value of GLUCOSE^1H_POST_75_G_GLUCOSE_PO | | 0.999394 | DISCARD |
| 1-year summary (Laboratory tests) - number of low values of GLUCOSE^1H_POST_75_G_GLUCOSE_PO | | 0.999394 | DISCARD |
| 1-year summary (Laboratory tests) - lowest value of GLUCOSE^1H_POST_75_G_GLUCOSE_PO | | 0.999394 | DISCARD |
| 1-year summary (Laboratory tests) - average value of GLUCOSE^1H_POST_75_G_GLUCOSE_PO | | 0.999394 | DISCARD |
| 1-year summary (Laboratory tests) - median value of GLUCOSE^1H_POST_75_G_GLUCOSE_PO | | 0.999394 | DISCARD |
| 1-year summary (Laboratory tests) - count of normal tests for GLUCOSE^1H_POST_75_G_GLUCOSE_PO | | 0.999394 | DISCARD |
| 2-year summary (Drug prescriptions) - ANION EXCHANGE RESINS Average frequency (days) | | 0.999394 | DISCARD |
| 2-year summary (Drug prescriptions) - ANION EXCHANGE RESINS Max frequency (days) | | 0.999394 | DISCARD |
| 2-year summary (Drug prescriptions) - ANION EXCHANGE RESINS Min frequency (days) | | 0.999394 | DISCARD |
| 2-year summary (Drug prescriptions) - ERYTHROPOIESIS REGULATING HORMONE Days since first prescription | | 0.999394 | DISCARD |
| 2-year summary (Drug prescriptions) - ERYTHROPOIESIS REGULATING HORMONE Days since last prescription | | 0.999394 | DISCARD |
| 2-year summary (Drug prescriptions) - ERYTHROPOIESIS REGULATING HORMONE Prescription count | | 0.999394 | DISCARD |
| 2-year summary (Drug prescriptions) - NON-DEPOLARIZING AGENTS Average frequency (days) | | 0.999394 | DISCARD |
| 2-year summary (Drug prescriptions) - NON-DEPOLARIZING AGENTS Max frequency (days) | | 0.999394 | DISCARD |

| | | |
|---|---|---|
| 2-year summary (Drug prescriptions) - NON-DEPOLARIZING AGENTS Min frequency (days) | 0.999394 | DISCARD |
| 2-year summary (Drug prescriptions) - SMOKING CESSATION AIDS Average frequency (days) | 0.999394 | DISCARD |
| 2-year summary (Drug prescriptions) - SMOKING CESSATION AIDS Max frequency (days) | 0.999394 | DISCARD |
| 2-year summary (Drug prescriptions) - SMOKING CESSATION AIDS Min frequency (days) | 0.999394 | DISCARD |
| 1-year summary (Drug prescriptions) - ANION EXCHANGE RESINS Days since first prescription | 0.999422 | DISCARD |
| 1-year summary (Drug prescriptions) - ANION EXCHANGE RESINS Days since last prescription | 0.999422 | DISCARD |
| 1-year summary (Drug prescriptions) - ANION EXCHANGE RESINS Prescription count | 0.999422 | DISCARD |
| 1-year summary (Drug prescriptions) - NUCLEOSIDE REVERSE TRANSCRIPTASE INHIBITOR Days since first prescription | 0.999422 | DISCARD |
| 1-year summary (Drug prescriptions) - NUCLEOSIDE REVERSE TRANSCRIPTASE INHIBITOR Days since last prescription | 0.999422 | DISCARD |
| 1-year summary (Drug prescriptions) - NUCLEOSIDE REVERSE TRANSCRIPTASE INHIBITOR Prescription count | 0.999422 | DISCARD |
| 2-year summary (Drug prescriptions) - ANTHELMINTICS Days since first prescription | 0.999422 | DISCARD |
| 2-year summary (Drug prescriptions) - ANTHELMINTICS Days since last prescription | 0.999422 | DISCARD |
| 2-year summary (Drug prescriptions) - ANTHELMINTICS Prescription count | 0.999422 | DISCARD |
| 2-year summary (Drug prescriptions) - BETA-BLOCKERS COMBINATION Average frequency (days) | 0.999422 | DISCARD |
| 2-year summary (Drug prescriptions) - BETA-BLOCKERS COMBINATION Days since first prescription | 0.999422 | DISCARD |
| 2-year summary (Drug prescriptions) - BETA-BLOCKERS COMBINATION Days since last prescription | 0.999422 | DISCARD |
| 2-year summary (Drug prescriptions) - BETA-BLOCKERS COMBINATION Max frequency (days) | 0.999422 | DISCARD |
| 2-year summary (Drug prescriptions) - BETA-BLOCKERS COMBINATION Min frequency (days) | 0.999422 | DISCARD |
| 2-year summary (Drug prescriptions) - BETA-BLOCKERS COMBINATION Prescription count | 0.999422 | DISCARD |
| 2-year summary (Drug prescriptions) - DIGESTANTS Days since first prescription | 0.999422 | DISCARD |
| 2-year summary (Drug prescriptions) - DIGESTANTS Days since last prescription | 0.999422 | DISCARD |
| 2-year summary (Drug prescriptions) - DIGESTANTS Prescription count | 0.999422 | DISCARD |
| 1-year summary (Drug prescriptions) - ANTIANDROGENS Days since first prescription | 0.999449 | DISCARD |
| 1-year summary (Drug prescriptions) - ANTIANDROGENS Days since last prescription | 0.999449 | DISCARD |
| 1-year summary (Drug prescriptions) - ANTIANDROGENS Prescription count | 0.999449 | DISCARD |
| 1-year summary (Drug prescriptions) - ANTIESTROGENS Days since first prescription | 0.999449 | DISCARD |
| 1-year summary (Drug prescriptions) - ANTIESTROGENS Days since last prescription | 0.999449 | DISCARD |
| 1-year summary (Drug prescriptions) - ANTIESTROGENS Prescription count | 0.999449 | DISCARD |
| 1-year summary (Drug prescriptions) - DIGESTANTS Days since first prescription | 0.999449 | DISCARD |
| 1-year summary (Drug prescriptions) - DIGESTANTS Days since last prescription | 0.999449 | DISCARD |
| 1-year summary (Drug prescriptions) - DIGESTANTS Prescription count | 0.999449 | DISCARD |
| 1-year summary (Drug prescriptions) - REPLACEMENT AGENTS Average frequency (days) | 0.999449 | DISCARD |
| 1-year summary (Drug prescriptions) - REPLACEMENT AGENTS Max frequency (days) | 0.999449 | DISCARD |
| 1-year summary (Drug prescriptions) - REPLACEMENT AGENTS Min frequency (days) | 0.999449 | DISCARD |
| 1-year summary (Doctor visits) - DENTAL SURGERY | 0.999449 | DISCARD |
| 1-year summary (Doctor visits) - ORAL PATHOLOGY | 0.999449 | DISCARD |
| 2-year summary (Drug prescriptions) - ALKYLATING AGENTS Days since first prescription | 0.999449 | DISCARD |

| | | | |
|---|---|---|---|
| 2-year summary (Drug prescriptions) - ALKYLATING AGENTS Days since last prescription | | 0.999449 | DISCARD |
| 2-year summary (Drug prescriptions) - ALKYLATING AGENTS Prescription count | | 0.999449 | DISCARD |
| 1-year summary (Drug prescriptions) - CONTRACEPTIVES Days since first prescription | | 0.999477 | DISCARD |
| 1-year summary (Drug prescriptions) - CONTRACEPTIVES Days since last prescription | | 0.999477 | DISCARD |
| 1-year summary (Drug prescriptions) - CONTRACEPTIVES Prescription count | | 0.999477 | DISCARD |
| 1-year summary (Drug prescriptions) - MISCELLANEOUS SKIN AND MUCOUS MEMBRANE AGENTS Average frequency (days) | | 0.999477 | DISCARD |
| 1-year summary (Drug prescriptions) - MISCELLANEOUS SKIN AND MUCOUS MEMBRANE AGENTS Max frequency (days) | | 0.999477 | DISCARD |
| 1-year summary (Drug prescriptions) - MISCELLANEOUS SKIN AND MUCOUS MEMBRANE AGENTS Min frequency (days) | | 0.999477 | DISCARD |
| 1-year summary (Drug prescriptions) - NUCLEOSIDE REVERSE TRANSCRIPTASE INHIBITOR Average frequency (days) | | 0.999477 | DISCARD |
| 1-year summary (Drug prescriptions) - NUCLEOSIDE REVERSE TRANSCRIPTASE INHIBITOR Max frequency (days) | | 0.999477 | DISCARD |
| 1-year summary (Drug prescriptions) - NUCLEOSIDE REVERSE TRANSCRIPTASE INHIBITOR Min frequency (days) | | 0.999477 | DISCARD |
| 2-year summary (Drug prescriptions) - ANTIDIURETICS Days since first prescription | | 0.999477 | DISCARD |
| 2-year summary (Drug prescriptions) - ANTIDIURETICS Days since last prescription | | 0.999477 | DISCARD |
| 2-year summary (Drug prescriptions) - ANTIDIURETICS Prescription count | | 0.999477 | DISCARD |
| 2-year summary (Drug prescriptions) - EMERGENCY CONTRACEPTIVES Days since first prescription | | 0.999477 | DISCARD |
| 2-year summary (Drug prescriptions) - EMERGENCY CONTRACEPTIVES Days since last prescription | | 0.999477 | DISCARD |
| 2-year summary (Drug prescriptions) - EMERGENCY CONTRACEPTIVES Prescription count | | 0.999477 | DISCARD |
| 2-year summary (Drug prescriptions) - NON-STEROIDAL ANTI-INFLAMMATORY: ASA BASE Days since first prescription | | 0.999477 | DISCARD |
| 2-year summary (Drug prescriptions) - NON-STEROIDAL ANTI-INFLAMMATORY: ASA BASE Days since last prescription | | 0.999477 | DISCARD |
| 2-year summary (Drug prescriptions) - NON-STEROIDAL ANTI-INFLAMMATORY: ASA BASE Prescription count | | 0.999477 | DISCARD |
| 2-year summary (Drug prescriptions) - PANCREATIC HORMONES Days since first prescription | | 0.999477 | DISCARD |
| 2-year summary (Drug prescriptions) - PANCREATIC HORMONES Days since last prescription | | 0.999477 | DISCARD |
| 2-year summary (Drug prescriptions) - PANCREATIC HORMONES Prescription count | | 0.999477 | DISCARD |
| 2-year summary (Drug prescriptions) - SEX HORMONES Average frequency (days) | | 0.999477 | DISCARD |
| 2-year summary (Drug prescriptions) - SEX HORMONES Max frequency (days) | | 0.999477 | DISCARD |
| 2-year summary (Drug prescriptions) - SEX HORMONES Min frequency (days) | | 0.999477 | DISCARD |
| 1-year summary (Drug prescriptions) - HYPERTHYROIDISM THERAPY Average frequency (days) | | 0.999504 | DISCARD |
| 1-year summary (Drug prescriptions) - HYPERTHYROIDISM THERAPY Max frequency (days) | | 0.999504 | DISCARD |
| 1-year summary (Drug prescriptions) - HYPERTHYROIDISM THERAPY Min frequency (days) | | 0.999504 | DISCARD |
| 1-year summary (Drug prescriptions) - PSYCHOTHERAPEUTIC AGENTS Average frequency (days) | | 0.999504 | DISCARD |
| 1-year summary (Drug prescriptions) - PSYCHOTHERAPEUTIC AGENTS Max frequency (days) | | 0.999504 | DISCARD |
| 1-year summary (Drug prescriptions) - PSYCHOTHERAPEUTIC AGENTS Min frequency (days) | | 0.999504 | DISCARD |
| 1-year summary (Drug prescriptions) - TOPICAL IMMUNOSUPRESSIVE AGENTS Average frequency (days) | | 0.999504 | DISCARD |
| 1-year summary (Drug prescriptions) - TOPICAL IMMUNOSUPRESSIVE AGENTS Max frequency (days) | | 0.999504 | DISCARD |
| 1-year summary (Drug prescriptions) - TOPICAL IMMUNOSUPRESSIVE AGENTS Min frequency (days) | | 0.999504 | DISCARD |
| 2-year summary (Drug prescriptions) - DIGESTANTS Average frequency (days) | | 0.999504 | DISCARD |
| 2-year summary (Drug prescriptions) - DIGESTANTS Max frequency (days) | | 0.999504 | DISCARD |

| | | |
|---|---|---|
| 2-year summary (Drug prescriptions) - DIGESTANTS Min frequency (days) | 0.999504 | DISCARD |
| 1-year summary (Drug prescriptions) - ANTIESTROGENS Average frequency (days) | 0.999532 | DISCARD |
| 1-year summary (Drug prescriptions) - ANTIESTROGENS Max frequency (days) | 0.999532 | DISCARD |
| 1-year summary (Drug prescriptions) - ANTIESTROGENS Min frequency (days) | 0.999532 | DISCARD |
| 1-year summary (Drug prescriptions) - DIGESTANTS Average frequency (days) | 0.999532 | DISCARD |
| 1-year summary (Drug prescriptions) - DIGESTANTS Max frequency (days) | 0.999532 | DISCARD |
| 1-year summary (Drug prescriptions) - DIGESTANTS Min frequency (days) | 0.999532 | DISCARD |
| 1-year summary (Drug prescriptions) - WOUND CARE AND SCAR MANAGEMENT AGENTS Days since first prescription | 0.999532 | DISCARD |
| 1-year summary (Drug prescriptions) - WOUND CARE AND SCAR MANAGEMENT AGENTS Days since last prescription | 0.999532 | DISCARD |
| 1-year summary (Drug prescriptions) - WOUND CARE AND SCAR MANAGEMENT AGENTS Prescription count | 0.999532 | DISCARD |
| 2-year summary (Drug prescriptions) - ALKYLATING AGENTS Average frequency (days) | 0.999532 | DISCARD |
| 2-year summary (Drug prescriptions) - ALKYLATING AGENTS Max frequency (days) | 0.999532 | DISCARD |
| 2-year summary (Drug prescriptions) - ALKYLATING AGENTS Min frequency (days) | 0.999532 | DISCARD |
| 2-year summary (Drug prescriptions) - ANTIANDROGENS Average frequency (days) | 0.999532 | DISCARD |
| 2-year summary (Drug prescriptions) - ANTIANDROGENS Max frequency (days) | 0.999532 | DISCARD |
| 2-year summary (Drug prescriptions) - ANTIANDROGENS Min frequency (days) | 0.999532 | DISCARD |
| 2-year summary (Drug prescriptions) - MISCELLANEOUS LOCAL ANTI-INFECTIVES Average frequency (days) | 0.999532 | DISCARD |
| 2-year summary (Drug prescriptions) - MISCELLANEOUS LOCAL ANTI-INFECTIVES Max frequency (days) | 0.999532 | DISCARD |
| 2-year summary (Drug prescriptions) - MISCELLANEOUS LOCAL ANTI-INFECTIVES Min frequency (days) | 0.999532 | DISCARD |
| 1-year summary (Drug prescriptions) - MYDRIATICS Days since first prescription | 0.999560 | DISCARD |
| 1-year summary (Drug prescriptions) - MYDRIATICS Days since last prescription | 0.999560 | DISCARD |
| 1-year summary (Drug prescriptions) - MYDRIATICS Prescription count | 0.999560 | DISCARD |
| 2-year summary (Drug prescriptions) - ANTILIPEMIC AND ATHEROSCLEROSIS INHIBITING AGENTS Days since first prescription | 0.999560 | DISCARD |
| 2-year summary (Drug prescriptions) - ANTILIPEMIC AND ATHEROSCLEROSIS INHIBITING AGENTS Days since last prescription | 0.999560 | DISCARD |
| 2-year summary (Drug prescriptions) - ANTILIPEMIC AND ATHEROSCLEROSIS INHIBITING AGENTS Prescription count | 0.999560 | DISCARD |
| 2-year summary (Drug prescriptions) - MEMBRANE STABILISING DRUGS TYPE IC Days since first prescription | 0.999560 | DISCARD |
| 2-year summary (Drug prescriptions) - MEMBRANE STABILISING DRUGS TYPE IC Days since last prescription | 0.999560 | DISCARD |
| 2-year summary (Drug prescriptions) - MEMBRANE STABILISING DRUGS TYPE IC Prescription count | 0.999560 | DISCARD |
| 2-year summary (Laboratory tests) - number of tests for Glucose^1H_post_dose_glucose | 0.999560 | DISCARD |
| 2-year summary (Laboratory tests) - days since earliest test for Glucose^1H_post_dose_glucose | 0.999560 | DISCARD |
| 2-year summary (Laboratory tests) - days since latest test for Glucose^1H_post_dose_glucose | 0.999560 | DISCARD |
| 2-year summary (Laboratory tests) - number of high values of Glucose^1H_post_dose_glucose | 0.999560 | DISCARD |
| 2-year summary (Laboratory tests) - highest value of Glucose^1H_post_dose_glucose | 0.999560 | DISCARD |
| 2-year summary (Laboratory tests) - number of low values of Glucose^1H_post_dose_glucose | 0.999560 | DISCARD |
| 2-year summary (Laboratory tests) - lowest value of Glucose^1H_post_dose_glucose | 0.999560 | DISCARD |
| 2-year summary (Laboratory tests) - average value of Glucose^1H_post_dose_glucose | 0.999560 | DISCARD |
| 2-year summary (Laboratory tests) - median value of Glucose^1H_post_dose_glucose | 0.999560 | DISCARD |

| | | | |
|---|---|---|---|
| 2-year summary (Laboratory tests) - count of normal tests for Glucose^1H_post_dose_glucose | | 0.999560 | DISCARD |
| 1-year summary (Hospitalizations) - Class - Diseases of the ear and mastoid process (H60-H95) | | 0.999587 | DISCARD |
| 1-year summary (Drug prescriptions) - BETA-BLOCKERS COMBINATION Days since first prescription | | 0.999587 | DISCARD |
| 1-year summary (Drug prescriptions) - BETA-BLOCKERS COMBINATION Days since last prescription | | 0.999587 | DISCARD |
| 1-year summary (Drug prescriptions) - BETA-BLOCKERS COMBINATION Prescription count | | 0.999587 | DISCARD |
| 2-year summary (Drug prescriptions) - ERYTHROPOIESIS REGULATING HORMONE Average frequency (days) | | 0.999587 | DISCARD |
| 2-year summary (Drug prescriptions) - ERYTHROPOIESIS REGULATING HORMONE Max frequency (days) | | 0.999587 | DISCARD |
| 2-year summary (Drug prescriptions) - ERYTHROPOIESIS REGULATING HORMONE Min frequency (days) | | 0.999587 | DISCARD |
| 2-year summary (Drug prescriptions) - XANTHINES Days since first prescription | | 0.999587 | DISCARD |
| 2-year summary (Drug prescriptions) - XANTHINES Days since last prescription | | 0.999587 | DISCARD |
| 2-year summary (Drug prescriptions) - XANTHINES Prescription count | | 0.999587 | DISCARD |
| 1-year summary (Drug prescriptions) - ANTIDIURETICS Days since first prescription | | 0.999615 | DISCARD |
| 1-year summary (Drug prescriptions) - ANTIDIURETICS Days since last prescription | | 0.999615 | DISCARD |
| 1-year summary (Drug prescriptions) - ANTIDIURETICS Prescription count | | 0.999615 | DISCARD |
| 1-year summary (Drug prescriptions) - BETA-BLOCKERS COMBINATION Average frequency (days) | | 0.999615 | DISCARD |
| 1-year summary (Drug prescriptions) - BETA-BLOCKERS COMBINATION Max frequency (days) | | 0.999615 | DISCARD |
| 1-year summary (Drug prescriptions) - BETA-BLOCKERS COMBINATION Min frequency (days) | | 0.999615 | DISCARD |
| 1-year summary (Drug prescriptions) - MEMBRANE STABILISING DRUGS TYPE IC Days since first prescription | | 0.999615 | DISCARD |
| 1-year summary (Drug prescriptions) - MEMBRANE STABILISING DRUGS TYPE IC Days since last prescription | | 0.999615 | DISCARD |
| 1-year summary (Drug prescriptions) - MEMBRANE STABILISING DRUGS TYPE IC Prescription count | | 0.999615 | DISCARD |
| 1-year summary (Drug prescriptions) - NON-DEPOLARIZING AGENTS Average frequency (days) | | 0.999615 | DISCARD |
| 1-year summary (Drug prescriptions) - NON-DEPOLARIZING AGENTS Max frequency (days) | | 0.999615 | DISCARD |
| 1-year summary (Drug prescriptions) - NON-DEPOLARIZING AGENTS Min frequency (days) | | 0.999615 | DISCARD |
| 2-year summary (Drug prescriptions) - ALL OTHER THERAPEUTIC PRODUCTS Days since first prescription | | 0.999615 | DISCARD |
| 2-year summary (Drug prescriptions) - ALL OTHER THERAPEUTIC PRODUCTS Days since last prescription | | 0.999615 | DISCARD |
| 2-year summary (Drug prescriptions) - ALL OTHER THERAPEUTIC PRODUCTS Prescription count | | 0.999615 | DISCARD |
| 2-year summary (Drug prescriptions) - ANTIDIURETICS Average frequency (days) | | 0.999615 | DISCARD |
| 2-year summary (Drug prescriptions) - ANTIDIURETICS Max frequency (days) | | 0.999615 | DISCARD |
| 2-year summary (Drug prescriptions) - ANTIDIURETICS Min frequency (days) | | 0.999615 | DISCARD |
| 2-year summary (Drug prescriptions) - MEMBRANE STABILISING DRUGS TYPE IC Average frequency (days) | | 0.999615 | DISCARD |
| 2-year summary (Drug prescriptions) - MEMBRANE STABILISING DRUGS TYPE IC Max frequency (days) | | 0.999615 | DISCARD |
| 2-year summary (Drug prescriptions) - MEMBRANE STABILISING DRUGS TYPE IC Min frequency (days) | | 0.999615 | DISCARD |
| 2-year summary (Laboratory tests) - number of tests for Glucose^2H_post_meal | | 0.999615 | DISCARD |
| 2-year summary (Laboratory tests) - days since earliest test for Glucose^2H_post_meal | | 0.999615 | DISCARD |
| 2-year summary (Laboratory tests) - days since latest test for Glucose^2H_post_meal | | 0.999615 | DISCARD |
| 2-year summary (Laboratory tests) - number of high values of Glucose^2H_post_meal | | 0.999615 | DISCARD |
| 2-year summary (Laboratory tests) - highest value of Glucose^2H_post_meal | | 0.999615 | DISCARD |

| | | |
|---|---|---|
| 2-year summary (Laboratory tests) - number of low values of Glucose^2H_post_meal | 0.999615 | DISCARD |
| 2-year summary (Laboratory tests) - lowest value of Glucose^2H_post_meal | 0.999615 | DISCARD |
| 2-year summary (Laboratory tests) - average value of Glucose^2H_post_meal | 0.999615 | DISCARD |
| 2-year summary (Laboratory tests) - median value of Glucose^2H_post_meal | 0.999615 | DISCARD |
| 2-year summary (Laboratory tests) - count of normal tests for Glucose^2H_post_meal | 0.999615 | DISCARD |
| 1-year summary (Drug prescriptions) - ANTIVIRALS Average frequency (days) | 0.999642 | DISCARD |
| 1-year summary (Drug prescriptions) - ANTIVIRALS Max frequency (days) | 0.999642 | DISCARD |
| 1-year summary (Drug prescriptions) - ANTIVIRALS Min frequency (days) | 0.999642 | DISCARD |
| 1-year summary (Drug prescriptions) - SEX HORMONES Days since first prescription | 0.999642 | DISCARD |
| 1-year summary (Drug prescriptions) - SEX HORMONES Days since last prescription | 0.999642 | DISCARD |
| 1-year summary (Drug prescriptions) - SEX HORMONES Prescription count | 0.999642 | DISCARD |
| 1-year summary (Drug prescriptions) - XANTHINES Days since first prescription | 0.999642 | DISCARD |
| 1-year summary (Drug prescriptions) - XANTHINES Days since last prescription | 0.999642 | DISCARD |
| 1-year summary (Drug prescriptions) - XANTHINES Prescription count | 0.999642 | DISCARD |
| 2-year summary (Drug prescriptions) - ANTIBIOTICS: OTHER Days since first prescription | 0.999642 | DISCARD |
| 2-year summary (Drug prescriptions) - ANTIBIOTICS: OTHER Days since last prescription | 0.999642 | DISCARD |
| 2-year summary (Drug prescriptions) - ANTIBIOTICS: OTHER Prescription count | 0.999642 | DISCARD |
| 2-year summary (Drug prescriptions) - GLAUCOMA THERAPY Days since first prescription | 0.999642 | DISCARD |
| 2-year summary (Drug prescriptions) - GLAUCOMA THERAPY Days since last prescription | 0.999642 | DISCARD |
| 2-year summary (Drug prescriptions) - GLAUCOMA THERAPY Prescription count | 0.999642 | DISCARD |
| 2-year summary (Drug prescriptions) - HERBS AND NATURAL PRODUCTS Days since first prescription | 0.999642 | DISCARD |
| 2-year summary (Drug prescriptions) - HERBS AND NATURAL PRODUCTS Days since last prescription | 0.999642 | DISCARD |
| 2-year summary (Drug prescriptions) - HERBS AND NATURAL PRODUCTS Prescription count | 0.999642 | DISCARD |
| 2-year summary (Drug prescriptions) - MINERALS Days since first prescription | 0.999642 | DISCARD |
| 2-year summary (Drug prescriptions) - MINERALS Days since last prescription | 0.999642 | DISCARD |
| 2-year summary (Drug prescriptions) - MINERALS Prescription count | 0.999642 | DISCARD |
| 2-year summary (Drug prescriptions) - NON-NUCLEOSIDE REVERSE TRANSCRIPTASE INHIBITOR Average frequency (days) | 0.999642 | DISCARD |
| 2-year summary (Drug prescriptions) - NON-NUCLEOSIDE REVERSE TRANSCRIPTASE INHIBITOR Days since first prescription | 0.999642 | DISCARD |
| 2-year summary (Drug prescriptions) - NON-NUCLEOSIDE REVERSE TRANSCRIPTASE INHIBITOR Days since last prescription | 0.999642 | DISCARD |
| 2-year summary (Drug prescriptions) - NON-NUCLEOSIDE REVERSE TRANSCRIPTASE INHIBITOR Max frequency (days) | 0.999642 | DISCARD |
| 2-year summary (Drug prescriptions) - NON-NUCLEOSIDE REVERSE TRANSCRIPTASE INHIBITOR Min frequency (days) | 0.999642 | DISCARD |
| 2-year summary (Drug prescriptions) - NON-NUCLEOSIDE REVERSE TRANSCRIPTASE INHIBITOR Prescription count | 0.999642 | DISCARD |
| 2-year summary (Drug prescriptions) - PERIPHERAL VASODILATORS Days since first prescription | 0.999642 | DISCARD |
| 2-year summary (Drug prescriptions) - PERIPHERAL VASODILATORS Days since last prescription | 0.999642 | DISCARD |
| 2-year summary (Drug prescriptions) - PERIPHERAL VASODILATORS Prescription count | 0.999642 | DISCARD |
| 1-year summary (Drug prescriptions) - ALKYLATING AGENTS Days since first prescription | 0.999670 | DISCARD |
| 1-year summary (Drug prescriptions) - ALKYLATING AGENTS Days since last prescription | 0.999670 | DISCARD |

| | | | |
|---|---|---|---|
| 1-year summary (Drug prescriptions) - ALKYLATING AGENTS Prescription count | | 0.999670 | DISCARD |
| 1-year summary (Drug prescriptions) - ANTIANDROGENS Average frequency (days) | | 0.999670 | DISCARD |
| 1-year summary (Drug prescriptions) - ANTIANDROGENS Max frequency (days) | | 0.999670 | DISCARD |
| 1-year summary (Drug prescriptions) - ANTIANDROGENS Min frequency (days) | | 0.999670 | DISCARD |
| 1-year summary (Drug prescriptions) - ERYTHROPOIESIS REGULATING HORMONE Days since first prescription | | 0.999670 | DISCARD |
| 1-year summary (Drug prescriptions) - ERYTHROPOIESIS REGULATING HORMONE Days since last prescription | | 0.999670 | DISCARD |
| 1-year summary (Drug prescriptions) - ERYTHROPOIESIS REGULATING HORMONE Prescription count | | 0.999670 | DISCARD |
| 1-year summary (Drug prescriptions) - MEMBRANE STABILISING DRUGS TYPE IC Average frequency (days) | | 0.999670 | DISCARD |
| 1-year summary (Drug prescriptions) - MEMBRANE STABILISING DRUGS TYPE IC Max frequency (days) | | 0.999670 | DISCARD |
| 1-year summary (Drug prescriptions) - MEMBRANE STABILISING DRUGS TYPE IC Min frequency (days) | | 0.999670 | DISCARD |
| 2-year summary (Drug prescriptions) - ANTILIPEMIC AND ATHEROSCLEROSIS INHIBITING AGENTS Average frequency (days) | | 0.999670 | DISCARD |
| 2-year summary (Drug prescriptions) - ANTILIPEMIC AND ATHEROSCLEROSIS INHIBITING AGENTS Max frequency (days) | | 0.999670 | DISCARD |
| 2-year summary (Drug prescriptions) - ANTILIPEMIC AND ATHEROSCLEROSIS INHIBITING AGENTS Min frequency (days) | | 0.999670 | DISCARD |
| 2-year summary (Drug prescriptions) - ANTINEOPLASTIC MISCELLANEOUS Days since first prescription | | 0.999670 | DISCARD |
| 2-year summary (Drug prescriptions) - ANTINEOPLASTIC MISCELLANEOUS Days since last prescription | | 0.999670 | DISCARD |
| 2-year summary (Drug prescriptions) - ANTINEOPLASTIC MISCELLANEOUS Prescription count | | 0.999670 | DISCARD |
| 2-year summary (Drug prescriptions) - ANTIVIRAL AGENTS - HIV/AIDS SPECIFIC Days since first prescription | | 0.999670 | DISCARD |
| 2-year summary (Drug prescriptions) - ANTIVIRAL AGENTS - HIV/AIDS SPECIFIC Days since last prescription | | 0.999670 | DISCARD |
| 2-year summary (Drug prescriptions) - ANTIVIRAL AGENTS - HIV/AIDS SPECIFIC Prescription count | | 0.999670 | DISCARD |
| 2-year summary (Drug prescriptions) - GLYCOPEPTIDES Days since first prescription | | 0.999670 | DISCARD |
| 2-year summary (Drug prescriptions) - GLYCOPEPTIDES Days since last prescription | | 0.999670 | DISCARD |
| 2-year summary (Drug prescriptions) - GLYCOPEPTIDES Prescription count | | 0.999670 | DISCARD |
| 2-year summary (Drug prescriptions) - MYASTHENIA GRAVIS Days since first prescription | | 0.999670 | DISCARD |
| 2-year summary (Drug prescriptions) - MYASTHENIA GRAVIS Days since last prescription | | 0.999670 | DISCARD |
| 2-year summary (Drug prescriptions) - MYASTHENIA GRAVIS Prescription count | | 0.999670 | DISCARD |
| 2-year summary (Drug prescriptions) - NON-SELECTIVE BETA-ADRENERGIC AGONISTS Average frequency (days) | | 0.999670 | DISCARD |
| 2-year summary (Drug prescriptions) - NON-SELECTIVE BETA-ADRENERGIC AGONISTS Max frequency (days) | | 0.999670 | DISCARD |
| 2-year summary (Drug prescriptions) - NON-SELECTIVE BETA-ADRENERGIC AGONISTS Min frequency (days) | | 0.999670 | DISCARD |
| 2-year summary (Drug prescriptions) - NON-STEROIDAL ANTI-INFLAMMATORY: ASA BASE Average frequency (days) | | 0.999670 | DISCARD |
| 2-year summary (Drug prescriptions) - NON-STEROIDAL ANTI-INFLAMMATORY: ASA BASE Max frequency (days) | | 0.999670 | DISCARD |
| 2-year summary (Drug prescriptions) - NON-STEROIDAL ANTI-INFLAMMATORY: ASA BASE Min frequency (days) | | 0.999670 | DISCARD |
| 2-year summary (Drug prescriptions) - XANTHINES Average frequency (days) | | 0.999670 | DISCARD |
| 2-year summary (Drug prescriptions) - XANTHINES Max frequency (days) | | 0.999670 | DISCARD |
| 2-year summary (Drug prescriptions) - XANTHINES Min frequency (days) | | 0.999670 | DISCARD |
| 1-year summary (Drug prescriptions) - ALL OTHER THERAPEUTIC PRODUCTS Days since first prescription | | 0.999697 | DISCARD |
| 1-year summary (Drug prescriptions) - ALL OTHER THERAPEUTIC PRODUCTS Days since last prescription | | 0.999697 | DISCARD |
| 1-year summary (Drug prescriptions) - ALL OTHER THERAPEUTIC PRODUCTS Prescription count | | 0.999697 | DISCARD |

| | | | |
|---|---|---|---|
| 1-year summary (Drug prescriptions) - ANION EXCHANGE RESINS Average frequency (days) | | 0.999697 | DISCARD |
| 1-year summary (Drug prescriptions) - ANION EXCHANGE RESINS Max frequency (days) | | 0.999697 | DISCARD |
| 1-year summary (Drug prescriptions) - ANION EXCHANGE RESINS Min frequency (days) | | 0.999697 | DISCARD |
| 1-year summary (Drug prescriptions) - ANTIDIURETICS Average frequency (days) | | 0.999697 | DISCARD |
| 1-year summary (Drug prescriptions) - ANTIDIURETICS Max frequency (days) | | 0.999697 | DISCARD |
| 1-year summary (Drug prescriptions) - ANTIDIURETICS Min frequency (days) | | 0.999697 | DISCARD |
| 1-year summary (Drug prescriptions) - MYASTHENIA GRAVIS Average frequency (days) | | 0.999697 | DISCARD |
| 1-year summary (Drug prescriptions) - MYASTHENIA GRAVIS Days since first prescription | | 0.999697 | DISCARD |
| 1-year summary (Drug prescriptions) - MYASTHENIA GRAVIS Days since last prescription | | 0.999697 | DISCARD |
| 1-year summary (Drug prescriptions) - MYASTHENIA GRAVIS Max frequency (days) | | 0.999697 | DISCARD |
| 1-year summary (Drug prescriptions) - MYASTHENIA GRAVIS Min frequency (days) | | 0.999697 | DISCARD |
| 1-year summary (Drug prescriptions) - MYASTHENIA GRAVIS Prescription count | | 0.999697 | DISCARD |
| 1-year summary (Drug prescriptions) - NON-NUCLEOSIDE REVERSE TRANSCRIPTASE INHIBITOR Days since first prescription | | 0.999697 | DISCARD |
| 1-year summary (Drug prescriptions) - NON-NUCLEOSIDE REVERSE TRANSCRIPTASE INHIBITOR Days since last prescription | | 0.999697 | DISCARD |
| 1-year summary (Drug prescriptions) - NON-NUCLEOSIDE REVERSE TRANSCRIPTASE INHIBITOR Prescription count | | 0.999697 | DISCARD |
| 1-year summary (Drug prescriptions) - PARASITICIDES Average frequency (days) | | 0.999697 | DISCARD |
| 1-year summary (Drug prescriptions) - PARASITICIDES Max frequency (days) | | 0.999697 | DISCARD |
| 1-year summary (Drug prescriptions) - PARASITICIDES Min frequency (days) | | 0.999697 | DISCARD |
| 1-year summary (Drug prescriptions) - PERIPHERAL VASODILATORS Days since first prescription | | 0.999697 | DISCARD |
| 1-year summary (Drug prescriptions) - PERIPHERAL VASODILATORS Days since last prescription | | 0.999697 | DISCARD |
| 1-year summary (Drug prescriptions) - PERIPHERAL VASODILATORS Prescription count | | 0.999697 | DISCARD |
| 1-year summary (Drug prescriptions) - SMOKING CESSATION AIDS Average frequency (days) | | 0.999697 | DISCARD |
| 1-year summary (Drug prescriptions) - SMOKING CESSATION AIDS Max frequency (days) | | 0.999697 | DISCARD |
| 1-year summary (Drug prescriptions) - SMOKING CESSATION AIDS Min frequency (days) | | 0.999697 | DISCARD |
| 2-year summary (Drug prescriptions) - ALKALINIZING AGENTS Days since first prescription | | 0.999697 | DISCARD |
| 2-year summary (Drug prescriptions) - ALKALINIZING AGENTS Days since last prescription | | 0.999697 | DISCARD |
| 2-year summary (Drug prescriptions) - ALKALINIZING AGENTS Prescription count | | 0.999697 | DISCARD |
| 2-year summary (Drug prescriptions) - HERBS AND NATURAL PRODUCTS Average frequency (days) | | 0.999697 | DISCARD |
| 2-year summary (Drug prescriptions) - HERBS AND NATURAL PRODUCTS Max frequency (days) | | 0.999697 | DISCARD |
| 2-year summary (Drug prescriptions) - HERBS AND NATURAL PRODUCTS Min frequency (days) | | 0.999697 | DISCARD |
| 2-year summary (Drug prescriptions) - MINERALS Average frequency (days) | | 0.999697 | DISCARD |
| 2-year summary (Drug prescriptions) - MINERALS Max frequency (days) | | 0.999697 | DISCARD |
| 2-year summary (Drug prescriptions) - MINERALS Min frequency (days) | | 0.999697 | DISCARD |
| 2-year summary (Drug prescriptions) - MYASTHENIA GRAVIS Average frequency (days) | | 0.999697 | DISCARD |
| 2-year summary (Drug prescriptions) - MYASTHENIA GRAVIS Max frequency (days) | | 0.999697 | DISCARD |
| 2-year summary (Drug prescriptions) - MYASTHENIA GRAVIS Min frequency (days) | | 0.999697 | DISCARD |
| 1-year summary (Drug prescriptions) - ALKYLATING AGENTS Average frequency (days) | | 0.999725 | DISCARD |

| | | | |
|---|---|---|---|
| 1-year summary (Drug prescriptions) - ALKYLATING AGENTS Max frequency (days) | | 0.999725 | DISCARD |
| 1-year summary (Drug prescriptions) - ALKYLATING AGENTS Min frequency (days) | | 0.999725 | DISCARD |
| 1-year summary (Drug prescriptions) - ANTILIPEMIC AND ATHEROSCLEROSIS INHIBITING AGENTS Days since first prescription | | 0.999725 | DISCARD |
| 1-year summary (Drug prescriptions) - ANTILIPEMIC AND ATHEROSCLEROSIS INHIBITING AGENTS Days since last prescription | | 0.999725 | DISCARD |
| 1-year summary (Drug prescriptions) - ANTILIPEMIC AND ATHEROSCLEROSIS INHIBITING AGENTS Prescription count | | 0.999725 | DISCARD |
| 1-year summary (Drug prescriptions) - EMERGENCY CONTRACEPTIVES Days since first prescription | | 0.999725 | DISCARD |
| 1-year summary (Drug prescriptions) - EMERGENCY CONTRACEPTIVES Days since last prescription | | 0.999725 | DISCARD |
| 1-year summary (Drug prescriptions) - EMERGENCY CONTRACEPTIVES Prescription count | | 0.999725 | DISCARD |
| 1-year summary (Drug prescriptions) - ERYTHROPOIESIS REGULATING HORMONE Average frequency (days) | | 0.999725 | DISCARD |
| 1-year summary (Drug prescriptions) - ERYTHROPOIESIS REGULATING HORMONE Max frequency (days) | | 0.999725 | DISCARD |
| 1-year summary (Drug prescriptions) - ERYTHROPOIESIS REGULATING HORMONE Min frequency (days) | | 0.999725 | DISCARD |
| 1-year summary (Drug prescriptions) - GLAUCOMA THERAPY Days since first prescription | | 0.999725 | DISCARD |
| 1-year summary (Drug prescriptions) - GLAUCOMA THERAPY Days since last prescription | | 0.999725 | DISCARD |
| 1-year summary (Drug prescriptions) - GLAUCOMA THERAPY Prescription count | | 0.999725 | DISCARD |
| 1-year summary (Drug prescriptions) - HERBS AND NATURAL PRODUCTS Days since first prescription | | 0.999725 | DISCARD |
| 1-year summary (Drug prescriptions) - HERBS AND NATURAL PRODUCTS Days since last prescription | | 0.999725 | DISCARD |
| 1-year summary (Drug prescriptions) - HERBS AND NATURAL PRODUCTS Prescription count | | 0.999725 | DISCARD |
| 1-year summary (Drug prescriptions) - PANCREATIC HORMONES Days since first prescription | | 0.999725 | DISCARD |
| 1-year summary (Drug prescriptions) - PANCREATIC HORMONES Days since last prescription | | 0.999725 | DISCARD |
| 1-year summary (Drug prescriptions) - PANCREATIC HORMONES Prescription count | | 0.999725 | DISCARD |
| 2-year summary (Drug prescriptions) - ALL OTHER THERAPEUTIC PRODUCTS Average frequency (days) | | 0.999725 | DISCARD |
| 2-year summary (Drug prescriptions) - ALL OTHER THERAPEUTIC PRODUCTS Max frequency (days) | | 0.999725 | DISCARD |
| 2-year summary (Drug prescriptions) - ALL OTHER THERAPEUTIC PRODUCTS Min frequency (days) | | 0.999725 | DISCARD |
| 2-year summary (Drug prescriptions) - ANALGESICS AND ANTIPYRETICS Days since first prescription | | 0.999725 | DISCARD |
| 2-year summary (Drug prescriptions) - ANALGESICS AND ANTIPYRETICS Days since last prescription | | 0.999725 | DISCARD |
| 2-year summary (Drug prescriptions) - ANALGESICS AND ANTIPYRETICS Prescription count | | 0.999725 | DISCARD |
| 2-year summary (Drug prescriptions) - ANTINEOPLASTIC MISCELLANEOUS Average frequency (days) | | 0.999725 | DISCARD |
| 2-year summary (Drug prescriptions) - ANTINEOPLASTIC MISCELLANEOUS Max frequency (days) | | 0.999725 | DISCARD |
| 2-year summary (Drug prescriptions) - ANTINEOPLASTIC MISCELLANEOUS Min frequency (days) | | 0.999725 | DISCARD |
| 2-year summary (Drug prescriptions) - ANTISEBORRHEIC AND ANTIPSORIATIC AGENTS Days since first prescription | | 0.999725 | DISCARD |
| 2-year summary (Drug prescriptions) - ANTISEBORRHEIC AND ANTIPSORIATIC AGENTS Days since last prescription | | 0.999725 | DISCARD |
| 2-year summary (Drug prescriptions) - ANTISEBORRHEIC AND ANTIPSORIATIC AGENTS Prescription count | | 0.999725 | DISCARD |
| 2-year summary (Drug prescriptions) - ANTITUBERCULAR AGENTS Days since first prescription | | 0.999725 | DISCARD |
| 2-year summary (Drug prescriptions) - ANTITUBERCULAR AGENTS Days since last prescription | | 0.999725 | DISCARD |
| 2-year summary (Drug prescriptions) - ANTITUBERCULAR AGENTS Prescription count | | 0.999725 | DISCARD |
| 2-year summary (Drug prescriptions) - ANTIVIRAL AGENTS - HIV/AIDS SPECIFIC Average frequency (days) | | 0.999725 | DISCARD |
| 2-year summary (Drug prescriptions) - ANTIVIRAL AGENTS - HIV/AIDS SPECIFIC Max frequency (days) | | 0.999725 | DISCARD |

| | | | |
|---|---|---|---|
| 2-year summary (Drug prescriptions) - ANTIVIRAL AGENTS - HIV/AIDS SPECIFIC Min frequency (days) | | 0.999725 | DISCARD |
| 2-year summary (Drug prescriptions) - PERIPHERAL VASODILATORS Average frequency (days) | | 0.999725 | DISCARD |
| 2-year summary (Drug prescriptions) - PERIPHERAL VASODILATORS Max frequency (days) | | 0.999725 | DISCARD |
| 2-year summary (Drug prescriptions) - PERIPHERAL VASODILATORS Min frequency (days) | | 0.999725 | DISCARD |
| 2-year summary (Laboratory tests) - number of tests for Prostate_specific_Ag.free | | 0.999725 | DISCARD |
| 2-year summary (Laboratory tests) - days since earliest test for Prostate_specific_Ag.free | | 0.999725 | DISCARD |
| 2-year summary (Laboratory tests) - days since latest test for Prostate_specific_Ag.free | | 0.999725 | DISCARD |
| 2-year summary (Laboratory tests) - number of high values of Prostate_specific_Ag.free | | 0.999725 | DISCARD |
| 2-year summary (Laboratory tests) - highest value of Prostate_specific_Ag.free | | 0.999725 | DISCARD |
| 2-year summary (Laboratory tests) - number of low values of Prostate_specific_Ag.free | | 0.999725 | DISCARD |
| 2-year summary (Laboratory tests) - lowest value of Prostate_specific_Ag.free | | 0.999725 | DISCARD |
| 2-year summary (Laboratory tests) - average value of Prostate_specific_Ag.free | | 0.999725 | DISCARD |
| 2-year summary (Laboratory tests) - median value of Prostate_specific_Ag.free | | 0.999725 | DISCARD |
| 2-year summary (Laboratory tests) - count of normal tests for Prostate_specific_Ag.free | | 0.999725 | DISCARD |
| 1-year summary (Drug prescriptions) - ALL OTHER THERAPEUTIC PRODUCTS Average frequency (days) | | 0.999752 | DISCARD |
| 1-year summary (Drug prescriptions) - ALL OTHER THERAPEUTIC PRODUCTS Max frequency (days) | | 0.999752 | DISCARD |
| 1-year summary (Drug prescriptions) - ALL OTHER THERAPEUTIC PRODUCTS Min frequency (days) | | 0.999752 | DISCARD |
| 1-year summary (Drug prescriptions) - MINERALS Average frequency (days) | | 0.999752 | DISCARD |
| 1-year summary (Drug prescriptions) - MINERALS Days since first prescription | | 0.999752 | DISCARD |
| 1-year summary (Drug prescriptions) - MINERALS Days since last prescription | | 0.999752 | DISCARD |
| 1-year summary (Drug prescriptions) - MINERALS Max frequency (days) | | 0.999752 | DISCARD |
| 1-year summary (Drug prescriptions) - MINERALS Min frequency (days) | | 0.999752 | DISCARD |
| 1-year summary (Drug prescriptions) - MINERALS Prescription count | | 0.999752 | DISCARD |
| 1-year summary (Drug prescriptions) - NON-NUCLEOSIDE REVERSE TRANSCRIPTASE INHIBITOR Average frequency (days) | | 0.999752 | DISCARD |
| 1-year summary (Drug prescriptions) - NON-NUCLEOSIDE REVERSE TRANSCRIPTASE INHIBITOR Max frequency (days) | | 0.999752 | DISCARD |
| 1-year summary (Drug prescriptions) - NON-NUCLEOSIDE REVERSE TRANSCRIPTASE INHIBITOR Min frequency (days) | | 0.999752 | DISCARD |
| 1-year summary (Drug prescriptions) - PERIPHERAL VASODILATORS Average frequency (days) | | 0.999752 | DISCARD |
| 1-year summary (Drug prescriptions) - PERIPHERAL VASODILATORS Max frequency (days) | | 0.999752 | DISCARD |
| 1-year summary (Drug prescriptions) - PERIPHERAL VASODILATORS Min frequency (days) | | 0.999752 | DISCARD |
| 1-year summary (Drug prescriptions) - PROTEASE INHIBITOR Average frequency (days) | | 0.999752 | DISCARD |
| 1-year summary (Drug prescriptions) - PROTEASE INHIBITOR Days since first prescription | | 0.999752 | DISCARD |
| 1-year summary (Drug prescriptions) - PROTEASE INHIBITOR Days since last prescription | | 0.999752 | DISCARD |
| 1-year summary (Drug prescriptions) - PROTEASE INHIBITOR Max frequency (days) | | 0.999752 | DISCARD |
| 1-year summary (Drug prescriptions) - PROTEASE INHIBITOR Min frequency (days) | | 0.999752 | DISCARD |
| 1-year summary (Drug prescriptions) - PROTEASE INHIBITOR Prescription count | | 0.999752 | DISCARD |
| 1-year summary (Drug prescriptions) - XANTHINES Average frequency (days) | | 0.999752 | DISCARD |
| 1-year summary (Drug prescriptions) - XANTHINES Max frequency (days) | | 0.999752 | DISCARD |

| | | |
|---|---|---|
| 1-year summary (Drug prescriptions) - XANTHINES Min frequency (days) | 0.999752 | DISCARD |
| 2-year summary (Drug prescriptions) - ALLERGENIC EXTRACTS Days since first prescription | 0.999752 | DISCARD |
| 2-year summary (Drug prescriptions) - ALLERGENIC EXTRACTS Days since last prescription | 0.999752 | DISCARD |
| 2-year summary (Drug prescriptions) - ALLERGENIC EXTRACTS Prescription count | 0.999752 | DISCARD |
| 2-year summary (Drug prescriptions) - EMOLLIENTS, DEMULCENTS AND PROTECTANTS Days since first prescription | 0.999752 | DISCARD |
| 2-year summary (Drug prescriptions) - EMOLLIENTS, DEMULCENTS AND PROTECTANTS Days since last prescription | 0.999752 | DISCARD |
| 2-year summary (Drug prescriptions) - EMOLLIENTS, DEMULCENTS AND PROTECTANTS Prescription count | 0.999752 | DISCARD |
| 2-year summary (Drug prescriptions) - PROGESTERON ANALOGUES Days since first prescription | 0.999752 | DISCARD |
| 2-year summary (Drug prescriptions) - PROGESTERON ANALOGUES Days since last prescription | 0.999752 | DISCARD |
| 2-year summary (Drug prescriptions) - PROGESTERON ANALOGUES Prescription count | 0.999752 | DISCARD |
| 2-year summary (Drug prescriptions) - PROTEASE INHIBITOR Average frequency (days) | 0.999752 | DISCARD |
| 2-year summary (Drug prescriptions) - PROTEASE INHIBITOR Days since first prescription | 0.999752 | DISCARD |
| 2-year summary (Drug prescriptions) - PROTEASE INHIBITOR Days since last prescription | 0.999752 | DISCARD |
| 2-year summary (Drug prescriptions) - PROTEASE INHIBITOR Max frequency (days) | 0.999752 | DISCARD |
| 2-year summary (Drug prescriptions) - PROTEASE INHIBITOR Min frequency (days) | 0.999752 | DISCARD |
| 2-year summary (Drug prescriptions) - PROTEASE INHIBITOR Prescription count | 0.999752 | DISCARD |
| 2-year summary (Drug prescriptions) - SEROTONIN (5-HT) RECEPTOR AGONIST Average frequency (days) | 0.999752 | DISCARD |
| 2-year summary (Drug prescriptions) - SEROTONIN (5-HT) RECEPTOR AGONIST Days since first prescription | 0.999752 | DISCARD |
| 2-year summary (Drug prescriptions) - SEROTONIN (5-HT) RECEPTOR AGONIST Days since last prescription | 0.999752 | DISCARD |
| 2-year summary (Drug prescriptions) - SEROTONIN (5-HT) RECEPTOR AGONIST Max frequency (days) | 0.999752 | DISCARD |
| 2-year summary (Drug prescriptions) - SEROTONIN (5-HT) RECEPTOR AGONIST Min frequency (days) | 0.999752 | DISCARD |
| 2-year summary (Drug prescriptions) - SEROTONIN (5-HT) RECEPTOR AGONIST Prescription count | 0.999752 | DISCARD |
| 1-year summary (Drug prescriptions) - ANALGESICS AND ANTIPYRETICS Days since first prescription | 0.999780 | DISCARD |
| 1-year summary (Drug prescriptions) - ANALGESICS AND ANTIPYRETICS Days since last prescription | 0.999780 | DISCARD |
| 1-year summary (Drug prescriptions) - ANALGESICS AND ANTIPYRETICS Prescription count | 0.999780 | DISCARD |
| 1-year summary (Drug prescriptions) - ANTHELMINTICS Days since first prescription | 0.999780 | DISCARD |
| 1-year summary (Drug prescriptions) - ANTHELMINTICS Days since last prescription | 0.999780 | DISCARD |
| 1-year summary (Drug prescriptions) - ANTHELMINTICS Prescription count | 0.999780 | DISCARD |
| 1-year summary (Drug prescriptions) - ANTISEBORRHEIC AND ANTIPSORIATIC AGENTS Days since first prescription | 0.999780 | DISCARD |
| 1-year summary (Drug prescriptions) - ANTISEBORRHEIC AND ANTIPSORIATIC AGENTS Days since last prescription | 0.999780 | DISCARD |
| 1-year summary (Drug prescriptions) - ANTISEBORRHEIC AND ANTIPSORIATIC AGENTS Prescription count | 0.999780 | DISCARD |
| 1-year summary (Drug prescriptions) - ANTIVIRAL AGENTS - HIV/AIDS SPECIFIC Days since first prescription | 0.999780 | DISCARD |
| 1-year summary (Drug prescriptions) - ANTIVIRAL AGENTS - HIV/AIDS SPECIFIC Days since last prescription | 0.999780 | DISCARD |
| 1-year summary (Drug prescriptions) - ANTIVIRAL AGENTS - HIV/AIDS SPECIFIC Prescription count | 0.999780 | DISCARD |
| 1-year summary (Drug prescriptions) - HERBS AND NATURAL PRODUCTS Average frequency (days) | 0.999780 | DISCARD |
| 1-year summary (Drug prescriptions) - HERBS AND NATURAL PRODUCTS Max frequency (days) | 0.999780 | DISCARD |
| 1-year summary (Drug prescriptions) - HERBS AND NATURAL PRODUCTS Min frequency (days) | 0.999780 | DISCARD |

| | | | |
|---|---|---|---|
| 1-year summary (Drug prescriptions) - MYDRIATICS Average frequency (days) | | 0.999780 | DISCARD |
| 1-year summary (Drug prescriptions) - MYDRIATICS Max frequency (days) | | 0.999780 | DISCARD |
| 1-year summary (Drug prescriptions) - MYDRIATICS Min frequency (days) | | 0.999780 | DISCARD |
| 2-year summary (Drug prescriptions) - ANALGESICS AND ANTIPYRETICS Average frequency (days) | | 0.999780 | DISCARD |
| 2-year summary (Drug prescriptions) - ANALGESICS AND ANTIPYRETICS Max frequency (days) | | 0.999780 | DISCARD |
| 2-year summary (Drug prescriptions) - ANALGESICS AND ANTIPYRETICS Min frequency (days) | | 0.999780 | DISCARD |
| 2-year summary (Drug prescriptions) - HORMONE ANTAGONISTS Days since first prescription | | 0.999780 | DISCARD |
| 2-year summary (Drug prescriptions) - HORMONE ANTAGONISTS Days since last prescription | | 0.999780 | DISCARD |
| 2-year summary (Drug prescriptions) - HORMONE ANTAGONISTS Prescription count | | 0.999780 | DISCARD |
| 2-year summary (Drug prescriptions) - MULTIPLE SCLEROSIS Days since first prescription | | 0.999780 | DISCARD |
| 2-year summary (Drug prescriptions) - MULTIPLE SCLEROSIS Days since last prescription | | 0.999780 | DISCARD |
| 2-year summary (Drug prescriptions) - MULTIPLE SCLEROSIS Prescription count | | 0.999780 | DISCARD |
| 2-year summary (Drug prescriptions) - MYDRIATICS Average frequency (days) | | 0.999780 | DISCARD |
| 2-year summary (Drug prescriptions) - MYDRIATICS Max frequency (days) | | 0.999780 | DISCARD |
| 2-year summary (Drug prescriptions) - MYDRIATICS Min frequency (days) | | 0.999780 | DISCARD |
| 2-year summary (Drug prescriptions) - NICOTINIC ACID GROUP Days since first prescription | | 0.999780 | DISCARD |
| 2-year summary (Drug prescriptions) - NICOTINIC ACID GROUP Days since last prescription | | 0.999780 | DISCARD |
| 2-year summary (Drug prescriptions) - NICOTINIC ACID GROUP Prescription count | | 0.999780 | DISCARD |
| 2-year summary (Drug prescriptions) - VITAMIN A Days since first prescription | | 0.999780 | DISCARD |
| 2-year summary (Drug prescriptions) - VITAMIN A Days since last prescription | | 0.999780 | DISCARD |
| 2-year summary (Drug prescriptions) - VITAMIN A Prescription count | | 0.999780 | DISCARD |
| 2-year summary (Drug prescriptions) - WOUND CARE AND SCAR MANAGEMENT AGENTS Average frequency (days) | | 0.999780 | DISCARD |
| 2-year summary (Drug prescriptions) - WOUND CARE AND SCAR MANAGEMENT AGENTS Max frequency (days) | | 0.999780 | DISCARD |
| 2-year summary (Drug prescriptions) - WOUND CARE AND SCAR MANAGEMENT AGENTS Min frequency (days) | | 0.999780 | DISCARD |
| 1-year summary (Drug prescriptions) - ALKALINIZING AGENTS Days since first prescription | | 0.999807 | DISCARD |
| 1-year summary (Drug prescriptions) - ALKALINIZING AGENTS Days since last prescription | | 0.999807 | DISCARD |
| 1-year summary (Drug prescriptions) - ALKALINIZING AGENTS Prescription count | | 0.999807 | DISCARD |
| 1-year summary (Drug prescriptions) - ALLERGENIC EXTRACTS Days since first prescription | | 0.999807 | DISCARD |
| 1-year summary (Drug prescriptions) - ALLERGENIC EXTRACTS Days since last prescription | | 0.999807 | DISCARD |
| 1-year summary (Drug prescriptions) - ALLERGENIC EXTRACTS Prescription count | | 0.999807 | DISCARD |
| 1-year summary (Drug prescriptions) - ANALGESICS AND ANTIPYRETICS Average frequency (days) | | 0.999807 | DISCARD |
| 1-year summary (Drug prescriptions) - ANALGESICS AND ANTIPYRETICS Max frequency (days) | | 0.999807 | DISCARD |
| 1-year summary (Drug prescriptions) - ANALGESICS AND ANTIPYRETICS Min frequency (days) | | 0.999807 | DISCARD |
| 1-year summary (Drug prescriptions) - ANTIBIOTICS: OTHER Days since first prescription | | 0.999807 | DISCARD |
| 1-year summary (Drug prescriptions) - ANTIBIOTICS: OTHER Days since last prescription | | 0.999807 | DISCARD |
| 1-year summary (Drug prescriptions) - ANTIBIOTICS: OTHER Prescription count | | 0.999807 | DISCARD |
| 1-year summary (Drug prescriptions) - ANTICONVULSANTS Days since first prescription | | 0.999807 | DISCARD |

| | | | |
|---|---|---|---|
| 1-year summary (Drug prescriptions) - ANTICONVULSANTS Days since last prescription | | 0.999807 | DISCARD |
| 1-year summary (Drug prescriptions) - ANTICONVULSANTS Prescription count | | 0.999807 | DISCARD |
| 1-year summary (Drug prescriptions) - ANTINEOPLASTIC MISCELLANEOUS Average frequency (days) | | 0.999807 | DISCARD |
| 1-year summary (Drug prescriptions) - ANTINEOPLASTIC MISCELLANEOUS Days since first prescription | | 0.999807 | DISCARD |
| 1-year summary (Drug prescriptions) - ANTINEOPLASTIC MISCELLANEOUS Days since last prescription | | 0.999807 | DISCARD |
| 1-year summary (Drug prescriptions) - ANTINEOPLASTIC MISCELLANEOUS Max frequency (days) | | 0.999807 | DISCARD |
| 1-year summary (Drug prescriptions) - ANTINEOPLASTIC MISCELLANEOUS Min frequency (days) | | 0.999807 | DISCARD |
| 1-year summary (Drug prescriptions) - ANTINEOPLASTIC MISCELLANEOUS Prescription count | | 0.999807 | DISCARD |
| 1-year summary (Drug prescriptions) - ANTITUBERCULAR AGENTS Days since first prescription | | 0.999807 | DISCARD |
| 1-year summary (Drug prescriptions) - ANTITUBERCULAR AGENTS Days since last prescription | | 0.999807 | DISCARD |
| 1-year summary (Drug prescriptions) - ANTITUBERCULAR AGENTS Prescription count | | 0.999807 | DISCARD |
| 1-year summary (Drug prescriptions) - ANTIVIRAL AGENTS - HIV/AIDS SPECIFIC Average frequency (days) | | 0.999807 | DISCARD |
| 1-year summary (Drug prescriptions) - ANTIVIRAL AGENTS - HIV/AIDS SPECIFIC Max frequency (days) | | 0.999807 | DISCARD |
| 1-year summary (Drug prescriptions) - ANTIVIRAL AGENTS - HIV/AIDS SPECIFIC Min frequency (days) | | 0.999807 | DISCARD |
| 1-year summary (Drug prescriptions) - BENZODIAZEPINE RELATED DRUGS Days since first prescription | | 0.999807 | DISCARD |
| 1-year summary (Drug prescriptions) - BENZODIAZEPINE RELATED DRUGS Days since last prescription | | 0.999807 | DISCARD |
| 1-year summary (Drug prescriptions) - BENZODIAZEPINE RELATED DRUGS Prescription count | | 0.999807 | DISCARD |
| 1-year summary (Drug prescriptions) - CHOLELITHOLYTIC AGENTS Days since first prescription | | 0.999807 | DISCARD |
| 1-year summary (Drug prescriptions) - CHOLELITHOLYTIC AGENTS Days since last prescription | | 0.999807 | DISCARD |
| 1-year summary (Drug prescriptions) - CHOLELITHOLYTIC AGENTS Prescription count | | 0.999807 | DISCARD |
| 1-year summary (Drug prescriptions) - GASTROINTESTINAL ANTISPASMODICS Days since first prescription | | 0.999807 | DISCARD |
| 1-year summary (Drug prescriptions) - GASTROINTESTINAL ANTISPASMODICS Days since last prescription | | 0.999807 | DISCARD |
| 1-year summary (Drug prescriptions) - GASTROINTESTINAL ANTISPASMODICS Prescription count | | 0.999807 | DISCARD |
| 1-year summary (Drug prescriptions) - HORMONE ANTAGONISTS Days since first prescription | | 0.999807 | DISCARD |
| 1-year summary (Drug prescriptions) - HORMONE ANTAGONISTS Days since last prescription | | 0.999807 | DISCARD |
| 1-year summary (Drug prescriptions) - HORMONE ANTAGONISTS Prescription count | | 0.999807 | DISCARD |
| 1-year summary (Drug prescriptions) - MULTIPLE SCLEROSIS Days since first prescription | | 0.999807 | DISCARD |
| 1-year summary (Drug prescriptions) - MULTIPLE SCLEROSIS Days since last prescription | | 0.999807 | DISCARD |
| 1-year summary (Drug prescriptions) - MULTIPLE SCLEROSIS Prescription count | | 0.999807 | DISCARD |
| 1-year summary (Drug prescriptions) - PROGESTERON ANALOGUES Days since first prescription | | 0.999807 | DISCARD |
| 1-year summary (Drug prescriptions) - PROGESTERON ANALOGUES Days since last prescription | | 0.999807 | DISCARD |
| 1-year summary (Drug prescriptions) - PROGESTERON ANALOGUES Prescription count | | 0.999807 | DISCARD |
| 1-year summary (Laboratory tests) - number of tests for Glucose^2H_post_meal | | 0.999807 | DISCARD |
| 1-year summary (Laboratory tests) - number of tests for Prostate_specific_Ag | | 0.999807 | DISCARD |
| 1-year summary (Laboratory tests) - days since earliest test for Glucose^2H_post_meal | | 0.999807 | DISCARD |
| 1-year summary (Laboratory tests) - days since earliest test for Prostate_specific_Ag | | 0.999807 | DISCARD |
| 1-year summary (Laboratory tests) - days since latest test for Glucose^2H_post_meal | | 0.999807 | DISCARD |

| | | | |
|---|---|---|---|
| 1-year summary (Laboratory tests) - days since latest test for Prostate_specific_Ag | | 0.999807 | DISCARD |
| 1-year summary (Laboratory tests) - number of high values of Glucose^2H_post_meal | | 0.999807 | DISCARD |
| 1-year summary (Laboratory tests) - number of high values of Prostate_specific_Ag | | 0.999807 | DISCARD |
| 1-year summary (Laboratory tests) - highest value of Glucose^2H_post_meal | | 0.999807 | DISCARD |
| 1-year summary (Laboratory tests) - highest value of Prostate_specific_Ag | | 0.999807 | DISCARD |
| 1-year summary (Laboratory tests) - number of low values of Glucose^2H_post_meal | | 0.999807 | DISCARD |
| 1-year summary (Laboratory tests) - number of low values of Prostate_specific_Ag | | 0.999807 | DISCARD |
| 1-year summary (Laboratory tests) - lowest value of Glucose^2H_post_meal | | 0.999807 | DISCARD |
| 1-year summary (Laboratory tests) - lowest value of Prostate_specific_Ag | | 0.999807 | DISCARD |
| 1-year summary (Laboratory tests) - average value of Glucose^2H_post_meal | | 0.999807 | DISCARD |
| 1-year summary (Laboratory tests) - average value of Prostate_specific_Ag | | 0.999807 | DISCARD |
| 1-year summary (Laboratory tests) - median value of Glucose^2H_post_meal | | 0.999807 | DISCARD |
| 1-year summary (Laboratory tests) - median value of Prostate_specific_Ag | | 0.999807 | DISCARD |
| 1-year summary (Laboratory tests) - count of normal tests for Glucose^2H_post_meal | | 0.999807 | DISCARD |
| 1-year summary (Laboratory tests) - count of normal tests for Prostate_specific_Ag | | 0.999807 | DISCARD |
| 2-year summary (Drug prescriptions) - ANTHELMINTICS Average frequency (days) | | 0.999807 | DISCARD |
| 2-year summary (Drug prescriptions) - ANTHELMINTICS Max frequency (days) | | 0.999807 | DISCARD |
| 2-year summary (Drug prescriptions) - ANTHELMINTICS Min frequency (days) | | 0.999807 | DISCARD |
| 2-year summary (Drug prescriptions) - ANTI-CYCLOMEGALOVIRUS (CMV) INFECTION Days since first prescription | | 0.999807 | DISCARD |
| 2-year summary (Drug prescriptions) - ANTI-CYCLOMEGALOVIRUS (CMV) INFECTION Days since last prescription | | 0.999807 | DISCARD |
| 2-year summary (Drug prescriptions) - ANTI-CYCLOMEGALOVIRUS (CMV) INFECTION Prescription count | | 0.999807 | DISCARD |
| 2-year summary (Drug prescriptions) - ANTICONVULSANTS Days since first prescription | | 0.999807 | DISCARD |
| 2-year summary (Drug prescriptions) - ANTICONVULSANTS Days since last prescription | | 0.999807 | DISCARD |
| 2-year summary (Drug prescriptions) - ANTICONVULSANTS Prescription count | | 0.999807 | DISCARD |
| 2-year summary (Drug prescriptions) - ANTISEBORRHEIC AND ANTIPSORIATIC AGENTS Average frequency (days) | | 0.999807 | DISCARD |
| 2-year summary (Drug prescriptions) - ANTISEBORRHEIC AND ANTIPSORIATIC AGENTS Max frequency (days) | | 0.999807 | DISCARD |
| 2-year summary (Drug prescriptions) - ANTISEBORRHEIC AND ANTIPSORIATIC AGENTS Min frequency (days) | | 0.999807 | DISCARD |
| 2-year summary (Drug prescriptions) - BENZODIAZEPINE RELATED DRUGS Days since first prescription | | 0.999807 | DISCARD |
| 2-year summary (Drug prescriptions) - BENZODIAZEPINE RELATED DRUGS Days since last prescription | | 0.999807 | DISCARD |
| 2-year summary (Drug prescriptions) - BENZODIAZEPINE RELATED DRUGS Prescription count | | 0.999807 | DISCARD |
| 2-year summary (Drug prescriptions) - CHOLELITHOLYTIC AGENTS Days since first prescription | | 0.999807 | DISCARD |
| 2-year summary (Drug prescriptions) - CHOLELITHOLYTIC AGENTS Days since last prescription | | 0.999807 | DISCARD |
| 2-year summary (Drug prescriptions) - CHOLELITHOLYTIC AGENTS Prescription count | | 0.999807 | DISCARD |
| 2-year summary (Drug prescriptions) - EMERGENCY CONTRACEPTIVES Average frequency (days) | | 0.999807 | DISCARD |
| 2-year summary (Drug prescriptions) - EMERGENCY CONTRACEPTIVES Max frequency (days) | | 0.999807 | DISCARD |
| 2-year summary (Drug prescriptions) - EMERGENCY CONTRACEPTIVES Min frequency (days) | | 0.999807 | DISCARD |
| 2-year summary (Drug prescriptions) - GASTROINTESTINAL ANTISPASMODICS Days since first prescription | | 0.999807 | DISCARD |

| | | |
|---|---|---|
| 2-year summary (Drug prescriptions) - GASTROINTESTINAL ANTISPASMODICS Days since last prescription | 0.999807 | DISCARD |
| 2-year summary (Drug prescriptions) - GASTROINTESTINAL ANTISPASMODICS Prescription count | 0.999807 | DISCARD |
| 2-year summary (Drug prescriptions) - MULTIPLE SCLEROSIS Average frequency (days) | 0.999807 | DISCARD |
| 2-year summary (Drug prescriptions) - MULTIPLE SCLEROSIS Max frequency (days) | 0.999807 | DISCARD |
| 2-year summary (Drug prescriptions) - MULTIPLE SCLEROSIS Min frequency (days) | 0.999807 | DISCARD |
| 2-year summary (Drug prescriptions) - PROGESTERON ANALOGUES Average frequency (days) | 0.999807 | DISCARD |
| 2-year summary (Drug prescriptions) - PROGESTERON ANALOGUES Max frequency (days) | 0.999807 | DISCARD |
| 2-year summary (Drug prescriptions) - PROGESTERON ANALOGUES Min frequency (days) | 0.999807 | DISCARD |
| 1-year summary (Drug prescriptions) - ANTICONVULSANTS Average frequency (days) | 0.999835 | DISCARD |
| 1-year summary (Drug prescriptions) - ANTICONVULSANTS Max frequency (days) | 0.999835 | DISCARD |
| 1-year summary (Drug prescriptions) - ANTICONVULSANTS Min frequency (days) | 0.999835 | DISCARD |
| 1-year summary (Drug prescriptions) - BENZODIAZEPINE RELATED DRUGS Average frequency (days) | 0.999835 | DISCARD |
| 1-year summary (Drug prescriptions) - BENZODIAZEPINE RELATED DRUGS Max frequency (days) | 0.999835 | DISCARD |
| 1-year summary (Drug prescriptions) - BENZODIAZEPINE RELATED DRUGS Min frequency (days) | 0.999835 | DISCARD |
| 1-year summary (Drug prescriptions) - CHOLELITHOLYTIC AGENTS Average frequency (days) | 0.999835 | DISCARD |
| 1-year summary (Drug prescriptions) - CHOLELITHOLYTIC AGENTS Max frequency (days) | 0.999835 | DISCARD |
| 1-year summary (Drug prescriptions) - CHOLELITHOLYTIC AGENTS Min frequency (days) | 0.999835 | DISCARD |
| 1-year summary (Drug prescriptions) - MISCELLANEOUS LOCAL ANTI-INFECTIVES Average frequency (days) | 0.999835 | DISCARD |
| 1-year summary (Drug prescriptions) - MISCELLANEOUS LOCAL ANTI-INFECTIVES Max frequency (days) | 0.999835 | DISCARD |
| 1-year summary (Drug prescriptions) - MISCELLANEOUS LOCAL ANTI-INFECTIVES Min frequency (days) | 0.999835 | DISCARD |
| 1-year summary (Drug prescriptions) - NON-SELECTIVE BETA-ADRENERGIC AGONISTS Average frequency (days) | 0.999835 | DISCARD |
| 1-year summary (Drug prescriptions) - NON-SELECTIVE BETA-ADRENERGIC AGONISTS Max frequency (days) | 0.999835 | DISCARD |
| 1-year summary (Drug prescriptions) - NON-SELECTIVE BETA-ADRENERGIC AGONISTS Min frequency (days) | 0.999835 | DISCARD |
| 1-year summary (Drug prescriptions) - NON-STEROIDAL ANTI-INFLAMMATORY: ASA BASE Days since first prescription | 0.999835 | DISCARD |
| 1-year summary (Drug prescriptions) - NON-STEROIDAL ANTI-INFLAMMATORY: ASA BASE Days since last prescription | 0.999835 | DISCARD |
| 1-year summary (Drug prescriptions) - NON-STEROIDAL ANTI-INFLAMMATORY: ASA BASE Prescription count | 0.999835 | DISCARD |
| 1-year summary (Drug prescriptions) - SEROTONIN (5-HT) RECEPTOR AGONIST Days since first prescription | 0.999835 | DISCARD |
| 1-year summary (Drug prescriptions) - SEROTONIN (5-HT) RECEPTOR AGONIST Days since last prescription | 0.999835 | DISCARD |
| 1-year summary (Drug prescriptions) - SEROTONIN (5-HT) RECEPTOR AGONIST Prescription count | 0.999835 | DISCARD |
| 1-year summary (Drug prescriptions) - SEX HORMONES Average frequency (days) | 0.999835 | DISCARD |
| 1-year summary (Drug prescriptions) - SEX HORMONES Max frequency (days) | 0.999835 | DISCARD |
| 1-year summary (Drug prescriptions) - SEX HORMONES Min frequency (days) | 0.999835 | DISCARD |
| 2-year summary (Drug prescriptions) - ADRENALS Days since first prescription | 0.999835 | DISCARD |
| 2-year summary (Drug prescriptions) - ADRENALS Days since last prescription | 0.999835 | DISCARD |
| 2-year summary (Drug prescriptions) - ADRENALS Prescription count | 0.999835 | DISCARD |
| 2-year summary (Drug prescriptions) - ADRENERGIC AND ANTICHOLINERGIC BRONCHODILATORS Days since first prescription | 0.999835 | DISCARD |
| 2-year summary (Drug prescriptions) - ADRENERGIC AND ANTICHOLINERGIC BRONCHODILATORS Days since last prescription | 0.999835 | DISCARD |

| | | |
|---|---|---|
| 2-year summary (Drug prescriptions) - ADRENERGIC AND ANTICHOLINERGIC BRONCHODILATORS Prescription count | 0.999835 | DISCARD |
| 2-year summary (Drug prescriptions) - ALKALINIZING AGENTS Average frequency (days) | 0.999835 | DISCARD |
| 2-year summary (Drug prescriptions) - ALKALINIZING AGENTS Max frequency (days) | 0.999835 | DISCARD |
| 2-year summary (Drug prescriptions) - ALKALINIZING AGENTS Min frequency (days) | 0.999835 | DISCARD |
| 2-year summary (Drug prescriptions) - ANTI- HEPATITIS Days since first prescription | 0.999835 | DISCARD |
| 2-year summary (Drug prescriptions) - ANTI- HEPATITIS Days since last prescription | 0.999835 | DISCARD |
| 2-year summary (Drug prescriptions) - ANTI- HEPATITIS Prescription count | 0.999835 | DISCARD |
| 2-year summary (Drug prescriptions) - ANTI-CYCLOMEGALOVIRUS (CMV) INFECTION Average frequency (days) | 0.999835 | DISCARD |
| 2-year summary (Drug prescriptions) - ANTI-CYCLOMEGALOVIRUS (CMV) INFECTION Max frequency (days) | 0.999835 | DISCARD |
| 2-year summary (Drug prescriptions) - ANTI-CYCLOMEGALOVIRUS (CMV) INFECTION Min frequency (days) | 0.999835 | DISCARD |
| 2-year summary (Drug prescriptions) - ANTICONVULSANTS Average frequency (days) | 0.999835 | DISCARD |
| 2-year summary (Drug prescriptions) - ANTICONVULSANTS Max frequency (days) | 0.999835 | DISCARD |
| 2-year summary (Drug prescriptions) - ANTICONVULSANTS Min frequency (days) | 0.999835 | DISCARD |
| 2-year summary (Drug prescriptions) - ANTITUBERCULAR AGENTS Average frequency (days) | 0.999835 | DISCARD |
| 2-year summary (Drug prescriptions) - ANTITUBERCULAR AGENTS Max frequency (days) | 0.999835 | DISCARD |
| 2-year summary (Drug prescriptions) - ANTITUBERCULAR AGENTS Min frequency (days) | 0.999835 | DISCARD |
| 2-year summary (Drug prescriptions) - BENZODIAZEPINE RELATED DRUGS Average frequency (days) | 0.999835 | DISCARD |
| 2-year summary (Drug prescriptions) - BENZODIAZEPINE RELATED DRUGS Max frequency (days) | 0.999835 | DISCARD |
| 2-year summary (Drug prescriptions) - BENZODIAZEPINE RELATED DRUGS Min frequency (days) | 0.999835 | DISCARD |
| 2-year summary (Drug prescriptions) - BIOLOGIC AGENT Days since first prescription | 0.999835 | DISCARD |
| 2-year summary (Drug prescriptions) - BIOLOGIC AGENT Days since last prescription | 0.999835 | DISCARD |
| 2-year summary (Drug prescriptions) - BIOLOGIC AGENT Prescription count | 0.999835 | DISCARD |
| 2-year summary (Drug prescriptions) - CHOLELITHOLYTIC AGENTS Average frequency (days) | 0.999835 | DISCARD |
| 2-year summary (Drug prescriptions) - CHOLELITHOLYTIC AGENTS Max frequency (days) | 0.999835 | DISCARD |
| 2-year summary (Drug prescriptions) - CHOLELITHOLYTIC AGENTS Min frequency (days) | 0.999835 | DISCARD |
| 2-year summary (Drug prescriptions) - GLAUCOMA THERAPY Average frequency (days) | 0.999835 | DISCARD |
| 2-year summary (Drug prescriptions) - GLAUCOMA THERAPY Max frequency (days) | 0.999835 | DISCARD |
| 2-year summary (Drug prescriptions) - GLAUCOMA THERAPY Min frequency (days) | 0.999835 | DISCARD |
| 2-year summary (Drug prescriptions) - NICOTINIC ACID GROUP Average frequency (days) | 0.999835 | DISCARD |
| 2-year summary (Drug prescriptions) - NICOTINIC ACID GROUP Max frequency (days) | 0.999835 | DISCARD |
| 2-year summary (Drug prescriptions) - NICOTINIC ACID GROUP Min frequency (days) | 0.999835 | DISCARD |
| 2-year summary (Drug prescriptions) - PANCREATIC HORMONES Average frequency (days) | 0.999835 | DISCARD |
| 2-year summary (Drug prescriptions) - PANCREATIC HORMONES Max frequency (days) | 0.999835 | DISCARD |
| 2-year summary (Drug prescriptions) - PANCREATIC HORMONES Min frequency (days) | 0.999835 | DISCARD |
| 2-year summary (Doctor visits) - COMMUNITY MEDICINE | 0.999835 | DISCARD |
| 2-year summary (Doctor visits) - ORAL RADIOLOGY | 0.999835 | DISCARD |
| 1-year summary (Drug prescriptions) - ADRENALS Average frequency (days) | 1.000000 | DISCARD |

| | | | |
|---|---|---|---|
| 1-year summary (Drug prescriptions) - ADRENALS Days since first prescription | | 1.000000 | DISCARD |
| 1-year summary (Drug prescriptions) - ADRENALS Days since last prescription | | 1.000000 | DISCARD |
| 1-year summary (Drug prescriptions) - ADRENALS Max frequency (days) | | 1.000000 | DISCARD |
| 1-year summary (Drug prescriptions) - ADRENALS Min frequency (days) | | 1.000000 | DISCARD |
| 1-year summary (Drug prescriptions) - ADRENALS Prescription count | | 1.000000 | DISCARD |
| 1-year summary (Drug prescriptions) - ADRENERGIC AND ANTICHOLINERGIC BRONCHODILATORS Average frequency (days) | | 1.000000 | DISCARD |
| 1-year summary (Drug prescriptions) - ADRENERGIC AND ANTICHOLINERGIC BRONCHODILATORS Days since first prescription | | 1.000000 | DISCARD |
| 1-year summary (Drug prescriptions) - ADRENERGIC AND ANTICHOLINERGIC BRONCHODILATORS Days since last prescription | | 1.000000 | DISCARD |
| 1-year summary (Drug prescriptions) - ADRENERGIC AND ANTICHOLINERGIC BRONCHODILATORS Max frequency (days) | | 1.000000 | DISCARD |
| 1-year summary (Drug prescriptions) - ADRENERGIC AND ANTICHOLINERGIC BRONCHODILATORS Min frequency (days) | | 1.000000 | DISCARD |
| 1-year summary (Drug prescriptions) - ADRENERGIC AND ANTICHOLINERGIC BRONCHODILATORS Prescription count | | 1.000000 | DISCARD |
| 1-year summary (Drug prescriptions) - ALCOHOL DEPENDENCE Days since first prescription | | 1.000000 | DISCARD |
| 1-year summary (Drug prescriptions) - ALCOHOL DEPENDENCE Days since last prescription | | 1.000000 | DISCARD |
| 1-year summary (Drug prescriptions) - ALCOHOL DEPENDENCE Prescription count | | 1.000000 | DISCARD |
| 1-year summary (Drug prescriptions) - ALKALINIZING AGENTS Average frequency (days) | | 1.000000 | DISCARD |
| 1-year summary (Drug prescriptions) - ALKALINIZING AGENTS Max frequency (days) | | 1.000000 | DISCARD |
| 1-year summary (Drug prescriptions) - ALKALINIZING AGENTS Min frequency (days) | | 1.000000 | DISCARD |
| 1-year summary (Drug prescriptions) - ALKALOIDS AND DERIVATIVES Average frequency (days) | | 1.000000 | DISCARD |
| 1-year summary (Drug prescriptions) - ALKALOIDS AND DERIVATIVES Days since first prescription | | 1.000000 | DISCARD |
| 1-year summary (Drug prescriptions) - ALKALOIDS AND DERIVATIVES Days since last prescription | | 1.000000 | DISCARD |
| 1-year summary (Drug prescriptions) - ALKALOIDS AND DERIVATIVES Max frequency (days) | | 1.000000 | DISCARD |
| 1-year summary (Drug prescriptions) - ALKALOIDS AND DERIVATIVES Min frequency (days) | | 1.000000 | DISCARD |
| 1-year summary (Drug prescriptions) - ALKALOIDS AND DERIVATIVES Prescription count | | 1.000000 | DISCARD |
| 1-year summary (Drug prescriptions) - ALLERGENIC EXTRACTS Average frequency (days) | | 1.000000 | DISCARD |
| 1-year summary (Drug prescriptions) - ALLERGENIC EXTRACTS Max frequency (days) | | 1.000000 | DISCARD |
| 1-year summary (Drug prescriptions) - ALLERGENIC EXTRACTS Min frequency (days) | | 1.000000 | DISCARD |
| 1-year summary (Drug prescriptions) - ALLERGY PREPARATIONS Days since first prescription | | 1.000000 | DISCARD |
| 1-year summary (Drug prescriptions) - ALLERGY PREPARATIONS Days since last prescription | | 1.000000 | DISCARD |
| 1-year summary (Drug prescriptions) - ALLERGY PREPARATIONS Prescription count | | 1.000000 | DISCARD |
| 1-year summary (Drug prescriptions) - ALPHA-2-ADRENERGIC AGONIST Average frequency (days) | | 1.000000 | DISCARD |
| 1-year summary (Drug prescriptions) - ALPHA-2-ADRENERGIC AGONIST Days since first prescription | | 1.000000 | DISCARD |
| 1-year summary (Drug prescriptions) - ALPHA-2-ADRENERGIC AGONIST Days since last prescription | | 1.000000 | DISCARD |
| 1-year summary (Drug prescriptions) - ALPHA-2-ADRENERGIC AGONIST Max frequency (days) | | 1.000000 | DISCARD |
| 1-year summary (Drug prescriptions) - ALPHA-2-ADRENERGIC AGONIST Min frequency (days) | | 1.000000 | DISCARD |
| 1-year summary (Drug prescriptions) - ALPHA-2-ADRENERGIC AGONIST Prescription count | | 1.000000 | DISCARD |
| 1-year summary (Drug prescriptions) - ANTHELMINTICS Average frequency (days) | | 1.000000 | DISCARD |
| 1-year summary (Drug prescriptions) - ANTHELMINTICS Max frequency (days) | | 1.000000 | DISCARD |

| | | |
|---|---|---|
| 1-year summary (Drug prescriptions) - ANTHELMINTICS Min frequency (days) | 1.000000 | DISCARD |
| 1-year summary (Drug prescriptions) - ANTI- HEPATITIS Average frequency (days) | 1.000000 | DISCARD |
| 1-year summary (Drug prescriptions) - ANTI- HEPATITIS Days since first prescription | 1.000000 | DISCARD |
| 1-year summary (Drug prescriptions) - ANTI- HEPATITIS Days since last prescription | 1.000000 | DISCARD |
| 1-year summary (Drug prescriptions) - ANTI- HEPATITIS Max frequency (days) | 1.000000 | DISCARD |
| 1-year summary (Drug prescriptions) - ANTI- HEPATITIS Min frequency (days) | 1.000000 | DISCARD |
| 1-year summary (Drug prescriptions) - ANTI- HEPATITIS Prescription count | 1.000000 | DISCARD |
| 1-year summary (Drug prescriptions) - ANTI-CYCLOMEGALOVIRUS (CMV) INFECTION Average frequency (days) | 1.000000 | DISCARD |
| 1-year summary (Drug prescriptions) - ANTI-CYCLOMEGALOVIRUS (CMV) INFECTION Days since first prescription | 1.000000 | DISCARD |
| 1-year summary (Drug prescriptions) - ANTI-CYCLOMEGALOVIRUS (CMV) INFECTION Days since last prescription | 1.000000 | DISCARD |
| 1-year summary (Drug prescriptions) - ANTI-CYCLOMEGALOVIRUS (CMV) INFECTION Max frequency (days) | 1.000000 | DISCARD |
| 1-year summary (Drug prescriptions) - ANTI-CYCLOMEGALOVIRUS (CMV) INFECTION Min frequency (days) | 1.000000 | DISCARD |
| 1-year summary (Drug prescriptions) - ANTI-CYCLOMEGALOVIRUS (CMV) INFECTION Prescription count | 1.000000 | DISCARD |
| 1-year summary (Drug prescriptions) - ANTIBIOTICS: OTHER Average frequency (days) | 1.000000 | DISCARD |
| 1-year summary (Drug prescriptions) - ANTIBIOTICS: OTHER Max frequency (days) | 1.000000 | DISCARD |
| 1-year summary (Drug prescriptions) - ANTIBIOTICS: OTHER Min frequency (days) | 1.000000 | DISCARD |
| 1-year summary (Drug prescriptions) - ANTIDOTES Average frequency (days) | 1.000000 | DISCARD |
| 1-year summary (Drug prescriptions) - ANTIDOTES Days since first prescription | 1.000000 | DISCARD |
| 1-year summary (Drug prescriptions) - ANTIDOTES Days since last prescription | 1.000000 | DISCARD |
| 1-year summary (Drug prescriptions) - ANTIDOTES Max frequency (days) | 1.000000 | DISCARD |
| 1-year summary (Drug prescriptions) - ANTIDOTES Min frequency (days) | 1.000000 | DISCARD |
| 1-year summary (Drug prescriptions) - ANTIDOTES Prescription count | 1.000000 | DISCARD |
| 1-year summary (Drug prescriptions) - ANTIFIBRINOLYTIC AGENTS Days since first prescription | 1.000000 | DISCARD |
| 1-year summary (Drug prescriptions) - ANTIFIBRINOLYTIC AGENTS Days since last prescription | 1.000000 | DISCARD |
| 1-year summary (Drug prescriptions) - ANTIFIBRINOLYTIC AGENTS Prescription count | 1.000000 | DISCARD |
| 1-year summary (Drug prescriptions) - ANTIHISTAMINE Average frequency (days) | 1.000000 | DISCARD |
| 1-year summary (Drug prescriptions) - ANTIHISTAMINE Days since first prescription | 1.000000 | DISCARD |
| 1-year summary (Drug prescriptions) - ANTIHISTAMINE Days since last prescription | 1.000000 | DISCARD |
| 1-year summary (Drug prescriptions) - ANTIHISTAMINE Max frequency (days) | 1.000000 | DISCARD |
| 1-year summary (Drug prescriptions) - ANTIHISTAMINE Min frequency (days) | 1.000000 | DISCARD |
| 1-year summary (Drug prescriptions) - ANTIHISTAMINE Prescription count | 1.000000 | DISCARD |
| 1-year summary (Drug prescriptions) - ANTIHISTAMINICS Average frequency (days) | 1.000000 | DISCARD |
| 1-year summary (Drug prescriptions) - ANTIHISTAMINICS Days since first prescription | 1.000000 | DISCARD |
| 1-year summary (Drug prescriptions) - ANTIHISTAMINICS Days since last prescription | 1.000000 | DISCARD |
| 1-year summary (Drug prescriptions) - ANTIHISTAMINICS Max frequency (days) | 1.000000 | DISCARD |
| 1-year summary (Drug prescriptions) - ANTIHISTAMINICS Min frequency (days) | 1.000000 | DISCARD |
| 1-year summary (Drug prescriptions) - ANTIHISTAMINICS Prescription count | 1.000000 | DISCARD |

| | | |
|---|---|---|
| 1-year summary (Drug prescriptions) - ANTILIPEMIC AND ATHEROSCLEROSIS INHIBITING AGENTS Average frequency (days) | 1.000000 | DISCARD |
| 1-year summary (Drug prescriptions) - ANTILIPEMIC AND ATHEROSCLEROSIS INHIBITING AGENTS Max frequency (days) | 1.000000 | DISCARD |
| 1-year summary (Drug prescriptions) - ANTILIPEMIC AND ATHEROSCLEROSIS INHIBITING AGENTS Min frequency (days) | 1.000000 | DISCARD |
| 1-year summary (Drug prescriptions) - ANTINEOPLASTIC AND IMMUNOMODULATING AGENTS Average frequency (days) | 1.000000 | DISCARD |
| 1-year summary (Drug prescriptions) - ANTINEOPLASTIC AND IMMUNOMODULATING AGENTS Days since first prescription | 1.000000 | DISCARD |
| 1-year summary (Drug prescriptions) - ANTINEOPLASTIC AND IMMUNOMODULATING AGENTS Days since last prescription | 1.000000 | DISCARD |
| 1-year summary (Drug prescriptions) - ANTINEOPLASTIC AND IMMUNOMODULATING AGENTS Max frequency (days) | 1.000000 | DISCARD |
| 1-year summary (Drug prescriptions) - ANTINEOPLASTIC AND IMMUNOMODULATING AGENTS Min frequency (days) | 1.000000 | DISCARD |
| 1-year summary (Drug prescriptions) - ANTINEOPLASTIC AND IMMUNOMODULATING AGENTS Prescription count | 1.000000 | DISCARD |
| 1-year summary (Drug prescriptions) - ANTIPARKINSONIAN AGENTS Average frequency (days) | 1.000000 | DISCARD |
| 1-year summary (Drug prescriptions) - ANTIPARKINSONIAN AGENTS Days since first prescription | 1.000000 | DISCARD |
| 1-year summary (Drug prescriptions) - ANTIPARKINSONIAN AGENTS Days since last prescription | 1.000000 | DISCARD |
| 1-year summary (Drug prescriptions) - ANTIPARKINSONIAN AGENTS Max frequency (days) | 1.000000 | DISCARD |
| 1-year summary (Drug prescriptions) - ANTIPARKINSONIAN AGENTS Min frequency (days) | 1.000000 | DISCARD |
| 1-year summary (Drug prescriptions) - ANTIPARKINSONIAN AGENTS Prescription count | 1.000000 | DISCARD |
| 1-year summary (Drug prescriptions) - ANTISEBORRHEIC AND ANTIPSORIATIC AGENTS Average frequency (days) | 1.000000 | DISCARD |
| 1-year summary (Drug prescriptions) - ANTISEBORRHEIC AND ANTIPSORIATIC AGENTS Max frequency (days) | 1.000000 | DISCARD |
| 1-year summary (Drug prescriptions) - ANTISEBORRHEIC AND ANTIPSORIATIC AGENTS Min frequency (days) | 1.000000 | DISCARD |
| 1-year summary (Drug prescriptions) - ANTITUBERCULAR AGENTS Average frequency (days) | 1.000000 | DISCARD |
| 1-year summary (Drug prescriptions) - ANTITUBERCULAR AGENTS Max frequency (days) | 1.000000 | DISCARD |
| 1-year summary (Drug prescriptions) - ANTITUBERCULAR AGENTS Min frequency (days) | 1.000000 | DISCARD |
| 1-year summary (Drug prescriptions) - AZASPIRODECANEDIONE DERIVATIVES Average frequency (days) | 1.000000 | DISCARD |
| 1-year summary (Drug prescriptions) - AZASPIRODECANEDIONE DERIVATIVES Days since first prescription | 1.000000 | DISCARD |
| 1-year summary (Drug prescriptions) - AZASPIRODECANEDIONE DERIVATIVES Days since last prescription | 1.000000 | DISCARD |
| 1-year summary (Drug prescriptions) - AZASPIRODECANEDIONE DERIVATIVES Max frequency (days) | 1.000000 | DISCARD |
| 1-year summary (Drug prescriptions) - AZASPIRODECANEDIONE DERIVATIVES Min frequency (days) | 1.000000 | DISCARD |
| 1-year summary (Drug prescriptions) - AZASPIRODECANEDIONE DERIVATIVES Prescription count | 1.000000 | DISCARD |
| 1-year summary (Drug prescriptions) - BIOLOGIC AGENT Average frequency (days) | 1.000000 | DISCARD |
| 1-year summary (Drug prescriptions) - BIOLOGIC AGENT Days since first prescription | 1.000000 | DISCARD |
| 1-year summary (Drug prescriptions) - BIOLOGIC AGENT Days since last prescription | 1.000000 | DISCARD |
| 1-year summary (Drug prescriptions) - BIOLOGIC AGENT Max frequency (days) | 1.000000 | DISCARD |
| 1-year summary (Drug prescriptions) - BIOLOGIC AGENT Min frequency (days) | 1.000000 | DISCARD |
| 1-year summary (Drug prescriptions) - BIOLOGIC AGENT Prescription count | 1.000000 | DISCARD |
| 1-year summary (Drug prescriptions) - BLOOD COAGULATION FACTORS Days since first prescription | 1.000000 | DISCARD |
| 1-year summary (Drug prescriptions) - BLOOD COAGULATION FACTORS Days since last prescription | 1.000000 | DISCARD |
| 1-year summary (Drug prescriptions) - BLOOD COAGULATION FACTORS Prescription count | 1.000000 | DISCARD |
| 1-year summary (Drug prescriptions) - CARDIOVASCULAR DRUGS: OTHER Average frequency (days) | 1.000000 | DISCARD |

| | | | |
|---|---|---|---|
| 1-year summary (Drug prescriptions) - CARDIOVASCULAR DRUGS: OTHER Days since first prescription | | 1.000000 | DISCARD |
| 1-year summary (Drug prescriptions) - CARDIOVASCULAR DRUGS: OTHER Days since last prescription | | 1.000000 | DISCARD |
| 1-year summary (Drug prescriptions) - CARDIOVASCULAR DRUGS: OTHER Max frequency (days) | | 1.000000 | DISCARD |
| 1-year summary (Drug prescriptions) - CARDIOVASCULAR DRUGS: OTHER Min frequency (days) | | 1.000000 | DISCARD |
| 1-year summary (Drug prescriptions) - CARDIOVASCULAR DRUGS: OTHER Prescription count | | 1.000000 | DISCARD |
| 1-year summary (Drug prescriptions) - CENTRAL NERVOUS SYSTEM DRUGS: MISCELLANEOUS Average frequency (days) | | 1.000000 | DISCARD |
| 1-year summary (Drug prescriptions) - CENTRAL NERVOUS SYSTEM DRUGS: MISCELLANEOUS Days since first prescription | | 1.000000 | DISCARD |
| 1-year summary (Drug prescriptions) - CENTRAL NERVOUS SYSTEM DRUGS: MISCELLANEOUS Days since last prescription | | 1.000000 | DISCARD |
| 1-year summary (Drug prescriptions) - CENTRAL NERVOUS SYSTEM DRUGS: MISCELLANEOUS Max frequency (days) | | 1.000000 | DISCARD |
| 1-year summary (Drug prescriptions) - CENTRAL NERVOUS SYSTEM DRUGS: MISCELLANEOUS Min frequency (days) | | 1.000000 | DISCARD |
| 1-year summary (Drug prescriptions) - CENTRAL NERVOUS SYSTEM DRUGS: MISCELLANEOUS Prescription count | | 1.000000 | DISCARD |
| 1-year summary (Drug prescriptions) - CONTRACEPTIVES Average frequency (days) | | 1.000000 | DISCARD |
| 1-year summary (Drug prescriptions) - CONTRACEPTIVES Max frequency (days) | | 1.000000 | DISCARD |
| 1-year summary (Drug prescriptions) - CONTRACEPTIVES Min frequency (days) | | 1.000000 | DISCARD |
| 1-year summary (Drug prescriptions) - DIRECT SKELETAL MUSCLE RELAXANT Average frequency (days) | | 1.000000 | DISCARD |
| 1-year summary (Drug prescriptions) - DIRECT SKELETAL MUSCLE RELAXANT Days since first prescription | | 1.000000 | DISCARD |
| 1-year summary (Drug prescriptions) - DIRECT SKELETAL MUSCLE RELAXANT Days since last prescription | | 1.000000 | DISCARD |
| 1-year summary (Drug prescriptions) - DIRECT SKELETAL MUSCLE RELAXANT Max frequency (days) | | 1.000000 | DISCARD |
| 1-year summary (Drug prescriptions) - DIRECT SKELETAL MUSCLE RELAXANT Min frequency (days) | | 1.000000 | DISCARD |
| 1-year summary (Drug prescriptions) - DIRECT SKELETAL MUSCLE RELAXANT Prescription count | | 1.000000 | DISCARD |
| 1-year summary (Drug prescriptions) - EMERGENCY CONTRACEPTIVES Average frequency (days) | | 1.000000 | DISCARD |
| 1-year summary (Drug prescriptions) - EMERGENCY CONTRACEPTIVES Max frequency (days) | | 1.000000 | DISCARD |
| 1-year summary (Drug prescriptions) - EMERGENCY CONTRACEPTIVES Min frequency (days) | | 1.000000 | DISCARD |
| 1-year summary (Drug prescriptions) - EMETICS Average frequency (days) | | 1.000000 | DISCARD |
| 1-year summary (Drug prescriptions) - EMETICS Days since first prescription | | 1.000000 | DISCARD |
| 1-year summary (Drug prescriptions) - EMETICS Days since last prescription | | 1.000000 | DISCARD |
| 1-year summary (Drug prescriptions) - EMETICS Max frequency (days) | | 1.000000 | DISCARD |
| 1-year summary (Drug prescriptions) - EMETICS Min frequency (days) | | 1.000000 | DISCARD |
| 1-year summary (Drug prescriptions) - EMETICS Prescription count | | 1.000000 | DISCARD |
| 1-year summary (Drug prescriptions) - EMOLLIENTS, DEMULCENTS AND PROTECTANTS Average frequency (days) | | 1.000000 | DISCARD |
| 1-year summary (Drug prescriptions) - EMOLLIENTS, DEMULCENTS AND PROTECTANTS Days since first prescription | | 1.000000 | DISCARD |
| 1-year summary (Drug prescriptions) - EMOLLIENTS, DEMULCENTS AND PROTECTANTS Days since last prescription | | 1.000000 | DISCARD |
| 1-year summary (Drug prescriptions) - EMOLLIENTS, DEMULCENTS AND PROTECTANTS Max frequency (days) | | 1.000000 | DISCARD |
| 1-year summary (Drug prescriptions) - EMOLLIENTS, DEMULCENTS AND PROTECTANTS Min frequency (days) | | 1.000000 | DISCARD |
| 1-year summary (Drug prescriptions) - EMOLLIENTS, DEMULCENTS AND PROTECTANTS Prescription count | | 1.000000 | DISCARD |
| 1-year summary (Drug prescriptions) - FIBRINOGEN-PLATELET BINDING INHIBITORS Average frequency (days) | | 1.000000 | DISCARD |
| 1-year summary (Drug prescriptions) - FIBRINOGEN-PLATELET BINDING INHIBITORS Days since first prescription | | 1.000000 | DISCARD |

| | | |
|---|---|---|
| 1-year summary (Drug prescriptions) - FIBRINOGEN-PLATELET BINDING INHIBITORS Days since last prescription | 1.000000 | DISCARD |
| 1-year summary (Drug prescriptions) - FIBRINOGEN-PLATELET BINDING INHIBITORS Max frequency (days) | 1.000000 | DISCARD |
| 1-year summary (Drug prescriptions) - FIBRINOGEN-PLATELET BINDING INHIBITORS Min frequency (days) | 1.000000 | DISCARD |
| 1-year summary (Drug prescriptions) - FIBRINOGEN-PLATELET BINDING INHIBITORS Prescription count | 1.000000 | DISCARD |
| 1-year summary (Drug prescriptions) - GASTROINTESTINAL ANTISPASMODICS Average frequency (days) | 1.000000 | DISCARD |
| 1-year summary (Drug prescriptions) - GASTROINTESTINAL ANTISPASMODICS Max frequency (days) | 1.000000 | DISCARD |
| 1-year summary (Drug prescriptions) - GASTROINTESTINAL ANTISPASMODICS Min frequency (days) | 1.000000 | DISCARD |
| 1-year summary (Drug prescriptions) - GLAUCOMA THERAPY Average frequency (days) | 1.000000 | DISCARD |
| 1-year summary (Drug prescriptions) - GLAUCOMA THERAPY Max frequency (days) | 1.000000 | DISCARD |
| 1-year summary (Drug prescriptions) - GLAUCOMA THERAPY Min frequency (days) | 1.000000 | DISCARD |
| 1-year summary (Drug prescriptions) - GLYCOPEPTIDES Average frequency (days) | 1.000000 | DISCARD |
| 1-year summary (Drug prescriptions) - GLYCOPEPTIDES Days since first prescription | 1.000000 | DISCARD |
| 1-year summary (Drug prescriptions) - GLYCOPEPTIDES Days since last prescription | 1.000000 | DISCARD |
| 1-year summary (Drug prescriptions) - GLYCOPEPTIDES Max frequency (days) | 1.000000 | DISCARD |
| 1-year summary (Drug prescriptions) - GLYCOPEPTIDES Min frequency (days) | 1.000000 | DISCARD |
| 1-year summary (Drug prescriptions) - GLYCOPEPTIDES Prescription count | 1.000000 | DISCARD |
| 1-year summary (Drug prescriptions) - GROWTH STIMULANT HORMONE Average frequency (days) | 1.000000 | DISCARD |
| 1-year summary (Drug prescriptions) - GROWTH STIMULANT HORMONE Days since first prescription | 1.000000 | DISCARD |
| 1-year summary (Drug prescriptions) - GROWTH STIMULANT HORMONE Days since last prescription | 1.000000 | DISCARD |
| 1-year summary (Drug prescriptions) - GROWTH STIMULANT HORMONE Max frequency (days) | 1.000000 | DISCARD |
| 1-year summary (Drug prescriptions) - GROWTH STIMULANT HORMONE Min frequency (days) | 1.000000 | DISCARD |
| 1-year summary (Drug prescriptions) - GROWTH STIMULANT HORMONE Prescription count | 1.000000 | DISCARD |
| 1-year summary (Drug prescriptions) - HEAVY METAL ANTAGONISTS Average frequency (days) | 1.000000 | DISCARD |
| 1-year summary (Drug prescriptions) - HEAVY METAL ANTAGONISTS Days since first prescription | 1.000000 | DISCARD |
| 1-year summary (Drug prescriptions) - HEAVY METAL ANTAGONISTS Days since last prescription | 1.000000 | DISCARD |
| 1-year summary (Drug prescriptions) - HEAVY METAL ANTAGONISTS Max frequency (days) | 1.000000 | DISCARD |
| 1-year summary (Drug prescriptions) - HEAVY METAL ANTAGONISTS Min frequency (days) | 1.000000 | DISCARD |
| 1-year summary (Drug prescriptions) - HEAVY METAL ANTAGONISTS Prescription count | 1.000000 | DISCARD |
| 1-year summary (Drug prescriptions) - HORMONE ANTAGONISTS Average frequency (days) | 1.000000 | DISCARD |
| 1-year summary (Drug prescriptions) - HORMONE ANTAGONISTS Max frequency (days) | 1.000000 | DISCARD |
| 1-year summary (Drug prescriptions) - HORMONE ANTAGONISTS Min frequency (days) | 1.000000 | DISCARD |
| 1-year summary (Drug prescriptions) - HORMONES ANALOGUES Average frequency (days) | 1.000000 | DISCARD |
| 1-year summary (Drug prescriptions) - HORMONES ANALOGUES Days since first prescription | 1.000000 | DISCARD |
| 1-year summary (Drug prescriptions) - HORMONES ANALOGUES Days since last prescription | 1.000000 | DISCARD |
| 1-year summary (Drug prescriptions) - HORMONES ANALOGUES Max frequency (days) | 1.000000 | DISCARD |
| 1-year summary (Drug prescriptions) - HORMONES ANALOGUES Min frequency (days) | 1.000000 | DISCARD |
| 1-year summary (Drug prescriptions) - HORMONES ANALOGUES Prescription count | 1.000000 | DISCARD |

| | | |
|---|---|---|
| 1-year summary (Drug prescriptions) - INTERLEUKIN INHIBITOR Average frequency (days) | 1.000000 | DISCARD |
| 1-year summary (Drug prescriptions) - INTERLEUKIN INHIBITOR Days since first prescription | 1.000000 | DISCARD |
| 1-year summary (Drug prescriptions) - INTERLEUKIN INHIBITOR Days since last prescription | 1.000000 | DISCARD |
| 1-year summary (Drug prescriptions) - INTERLEUKIN INHIBITOR Max frequency (days) | 1.000000 | DISCARD |
| 1-year summary (Drug prescriptions) - INTERLEUKIN INHIBITOR Min frequency (days) | 1.000000 | DISCARD |
| 1-year summary (Drug prescriptions) - INTERLEUKIN INHIBITOR Prescription count | 1.000000 | DISCARD |
| 1-year summary (Drug prescriptions) - IRREVERSIBLE MAO Average frequency (days) | 1.000000 | DISCARD |
| 1-year summary (Drug prescriptions) - IRREVERSIBLE MAO Days since first prescription | 1.000000 | DISCARD |
| 1-year summary (Drug prescriptions) - IRREVERSIBLE MAO Days since last prescription | 1.000000 | DISCARD |
| 1-year summary (Drug prescriptions) - IRREVERSIBLE MAO Max frequency (days) | 1.000000 | DISCARD |
| 1-year summary (Drug prescriptions) - IRREVERSIBLE MAO Min frequency (days) | 1.000000 | DISCARD |
| 1-year summary (Drug prescriptions) - IRREVERSIBLE MAO Prescription count | 1.000000 | DISCARD |
| 1-year summary (Drug prescriptions) - LEUKOTRIENE RECEPTOR ANTAGONISTS Average frequency (days) | 1.000000 | DISCARD |
| 1-year summary (Drug prescriptions) - LEUKOTRIENE RECEPTOR ANTAGONISTS Days since first prescription | 1.000000 | DISCARD |
| 1-year summary (Drug prescriptions) - LEUKOTRIENE RECEPTOR ANTAGONISTS Days since last prescription | 1.000000 | DISCARD |
| 1-year summary (Drug prescriptions) - LEUKOTRIENE RECEPTOR ANTAGONISTS Max frequency (days) | 1.000000 | DISCARD |
| 1-year summary (Drug prescriptions) - LEUKOTRIENE RECEPTOR ANTAGONISTS Min frequency (days) | 1.000000 | DISCARD |
| 1-year summary (Drug prescriptions) - LEUKOTRIENE RECEPTOR ANTAGONISTS Prescription count | 1.000000 | DISCARD |
| 1-year summary (Drug prescriptions) - LOCAL ANESTHETICS INJ Average frequency (days) | 1.000000 | DISCARD |
| 1-year summary (Drug prescriptions) - LOCAL ANESTHETICS INJ Days since first prescription | 1.000000 | DISCARD |
| 1-year summary (Drug prescriptions) - LOCAL ANESTHETICS INJ Days since last prescription | 1.000000 | DISCARD |
| 1-year summary (Drug prescriptions) - LOCAL ANESTHETICS INJ Max frequency (days) | 1.000000 | DISCARD |
| 1-year summary (Drug prescriptions) - LOCAL ANESTHETICS INJ Min frequency (days) | 1.000000 | DISCARD |
| 1-year summary (Drug prescriptions) - LOCAL ANESTHETICS INJ Prescription count | 1.000000 | DISCARD |
| 1-year summary (Drug prescriptions) - MEMBRANE STABILISING DRUGS TYPE A Average frequency (days) | 1.000000 | DISCARD |
| 1-year summary (Drug prescriptions) - MEMBRANE STABILISING DRUGS TYPE A Days since first prescription | 1.000000 | DISCARD |
| 1-year summary (Drug prescriptions) - MEMBRANE STABILISING DRUGS TYPE A Days since last prescription | 1.000000 | DISCARD |
| 1-year summary (Drug prescriptions) - MEMBRANE STABILISING DRUGS TYPE A Max frequency (days) | 1.000000 | DISCARD |
| 1-year summary (Drug prescriptions) - MEMBRANE STABILISING DRUGS TYPE A Min frequency (days) | 1.000000 | DISCARD |
| 1-year summary (Drug prescriptions) - MEMBRANE STABILISING DRUGS TYPE A Prescription count | 1.000000 | DISCARD |
| 1-year summary (Drug prescriptions) - MULTIPLE SCLEROSIS Average frequency (days) | 1.000000 | DISCARD |
| 1-year summary (Drug prescriptions) - MULTIPLE SCLEROSIS Max frequency (days) | 1.000000 | DISCARD |
| 1-year summary (Drug prescriptions) - MULTIPLE SCLEROSIS Min frequency (days) | 1.000000 | DISCARD |
| 1-year summary (Drug prescriptions) - MULTIVITAMINS Average frequency (days) | 1.000000 | DISCARD |
| 1-year summary (Drug prescriptions) - MULTIVITAMINS Days since first prescription | 1.000000 | DISCARD |
| 1-year summary (Drug prescriptions) - MULTIVITAMINS Days since last prescription | 1.000000 | DISCARD |
| 1-year summary (Drug prescriptions) - MULTIVITAMINS Max frequency (days) | 1.000000 | DISCARD |

| | | |
|---|---|---|
| 1-year summary (Drug prescriptions) - MULTIVITAMINS Min frequency (days) | 1.000000 | DISCARD |
| 1-year summary (Drug prescriptions) - MULTIVITAMINS Prescription count | 1.000000 | DISCARD |
| 1-year summary (Drug prescriptions) - NEUROMUSCULAR BLOCKING AGENT Average frequency (days) | 1.000000 | DISCARD |
| 1-year summary (Drug prescriptions) - NEUROMUSCULAR BLOCKING AGENT Days since first prescription | 1.000000 | DISCARD |
| 1-year summary (Drug prescriptions) - NEUROMUSCULAR BLOCKING AGENT Days since last prescription | 1.000000 | DISCARD |
| 1-year summary (Drug prescriptions) - NEUROMUSCULAR BLOCKING AGENT Max frequency (days) | 1.000000 | DISCARD |
| 1-year summary (Drug prescriptions) - NEUROMUSCULAR BLOCKING AGENT Min frequency (days) | 1.000000 | DISCARD |
| 1-year summary (Drug prescriptions) - NEUROMUSCULAR BLOCKING AGENT Prescription count | 1.000000 | DISCARD |
| 1-year summary (Drug prescriptions) - NICOTINIC ACID GROUP Average frequency (days) | 1.000000 | DISCARD |
| 1-year summary (Drug prescriptions) - NICOTINIC ACID GROUP Days since first prescription | 1.000000 | DISCARD |
| 1-year summary (Drug prescriptions) - NICOTINIC ACID GROUP Days since last prescription | 1.000000 | DISCARD |
| 1-year summary (Drug prescriptions) - NICOTINIC ACID GROUP Max frequency (days) | 1.000000 | DISCARD |
| 1-year summary (Drug prescriptions) - NICOTINIC ACID GROUP Min frequency (days) | 1.000000 | DISCARD |
| 1-year summary (Drug prescriptions) - NICOTINIC ACID GROUP Prescription count | 1.000000 | DISCARD |
| 1-year summary (Drug prescriptions) - NON-STEROIDAL ANTI-INFLAMMATORY: ASA BASE Average frequency (days) | 1.000000 | DISCARD |
| 1-year summary (Drug prescriptions) - NON-STEROIDAL ANTI-INFLAMMATORY: ASA BASE Max frequency (days) | 1.000000 | DISCARD |
| 1-year summary (Drug prescriptions) - NON-STEROIDAL ANTI-INFLAMMATORY: ASA BASE Min frequency (days) | 1.000000 | DISCARD |
| 1-year summary (Drug prescriptions) - OTHER ANTIPARKINSONIAN AGENTS Average frequency (days) | 1.000000 | DISCARD |
| 1-year summary (Drug prescriptions) - OTHER ANTIPARKINSONIAN AGENTS Days since first prescription | 1.000000 | DISCARD |
| 1-year summary (Drug prescriptions) - OTHER ANTIPARKINSONIAN AGENTS Days since last prescription | 1.000000 | DISCARD |
| 1-year summary (Drug prescriptions) - OTHER ANTIPARKINSONIAN AGENTS Max frequency (days) | 1.000000 | DISCARD |
| 1-year summary (Drug prescriptions) - OTHER ANTIPARKINSONIAN AGENTS Min frequency (days) | 1.000000 | DISCARD |
| 1-year summary (Drug prescriptions) - OTHER ANTIPARKINSONIAN AGENTS Prescription count | 1.000000 | DISCARD |
| 1-year summary (Drug prescriptions) - OTHER HORMONAL CONTRACEPTIVES Average frequency (days) | 1.000000 | DISCARD |
| 1-year summary (Drug prescriptions) - OTHER HORMONAL CONTRACEPTIVES Max frequency (days) | 1.000000 | DISCARD |
| 1-year summary (Drug prescriptions) - OTHER HORMONAL CONTRACEPTIVES Min frequency (days) | 1.000000 | DISCARD |
| 1-year summary (Drug prescriptions) - OTHER MIGRAINES THERAPY Average frequency (days) | 1.000000 | DISCARD |
| 1-year summary (Drug prescriptions) - OTHER MIGRAINES THERAPY Days since first prescription | 1.000000 | DISCARD |
| 1-year summary (Drug prescriptions) - OTHER MIGRAINES THERAPY Days since last prescription | 1.000000 | DISCARD |
| 1-year summary (Drug prescriptions) - OTHER MIGRAINES THERAPY Max frequency (days) | 1.000000 | DISCARD |
| 1-year summary (Drug prescriptions) - OTHER MIGRAINES THERAPY Min frequency (days) | 1.000000 | DISCARD |
| 1-year summary (Drug prescriptions) - OTHER MIGRAINES THERAPY Prescription count | 1.000000 | DISCARD |
| 1-year summary (Drug prescriptions) - PANCREATIC HORMONES Average frequency (days) | 1.000000 | DISCARD |
| 1-year summary (Drug prescriptions) - PANCREATIC HORMONES Max frequency (days) | 1.000000 | DISCARD |
| 1-year summary (Drug prescriptions) - PANCREATIC HORMONES Min frequency (days) | 1.000000 | DISCARD |
| 1-year summary (Drug prescriptions) - PARASYMPATHOMIMETIC (CHOLINERGIC) Average frequency (days) | 1.000000 | DISCARD |
| 1-year summary (Drug prescriptions) - PARASYMPATHOMIMETIC (CHOLINERGIC) Days since first prescription | 1.000000 | DISCARD |

| | | |
|---|---|---|
| 1-year summary (Drug prescriptions) - PARASYMPATHOMIMETIC (CHOLINERGIC) Days since last prescription | 1.000000 | DISCARD |
| 1-year summary (Drug prescriptions) - PARASYMPATHOMIMETIC (CHOLINERGIC) Max frequency (days) | 1.000000 | DISCARD |
| 1-year summary (Drug prescriptions) - PARASYMPATHOMIMETIC (CHOLINERGIC) Min frequency (days) | 1.000000 | DISCARD |
| 1-year summary (Drug prescriptions) - PARASYMPATHOMIMETIC (CHOLINERGIC) Prescription count | 1.000000 | DISCARD |
| 1-year summary (Drug prescriptions) - PARASYMPATHOMIMETIC AGENTS Average frequency (days) | 1.000000 | DISCARD |
| 1-year summary (Drug prescriptions) - PARASYMPATHOMIMETIC AGENTS Days since first prescription | 1.000000 | DISCARD |
| 1-year summary (Drug prescriptions) - PARASYMPATHOMIMETIC AGENTS Days since last prescription | 1.000000 | DISCARD |
| 1-year summary (Drug prescriptions) - PARASYMPATHOMIMETIC AGENTS Max frequency (days) | 1.000000 | DISCARD |
| 1-year summary (Drug prescriptions) - PARASYMPATHOMIMETIC AGENTS Min frequency (days) | 1.000000 | DISCARD |
| 1-year summary (Drug prescriptions) - PARASYMPATHOMIMETIC AGENTS Prescription count | 1.000000 | DISCARD |
| 1-year summary (Drug prescriptions) - PARATHYROID AGENTS Average frequency (days) | 1.000000 | DISCARD |
| 1-year summary (Drug prescriptions) - PARATHYROID AGENTS Days since first prescription | 1.000000 | DISCARD |
| 1-year summary (Drug prescriptions) - PARATHYROID AGENTS Days since last prescription | 1.000000 | DISCARD |
| 1-year summary (Drug prescriptions) - PARATHYROID AGENTS Max frequency (days) | 1.000000 | DISCARD |
| 1-year summary (Drug prescriptions) - PARATHYROID AGENTS Min frequency (days) | 1.000000 | DISCARD |
| 1-year summary (Drug prescriptions) - PARATHYROID AGENTS Prescription count | 1.000000 | DISCARD |
| 1-year summary (Drug prescriptions) - PITUITARY GONADOTROPIN INHIBITOR Average frequency (days) | 1.000000 | DISCARD |
| 1-year summary (Drug prescriptions) - PITUITARY GONADOTROPIN INHIBITOR Days since first prescription | 1.000000 | DISCARD |
| 1-year summary (Drug prescriptions) - PITUITARY GONADOTROPIN INHIBITOR Days since last prescription | 1.000000 | DISCARD |
| 1-year summary (Drug prescriptions) - PITUITARY GONADOTROPIN INHIBITOR Max frequency (days) | 1.000000 | DISCARD |
| 1-year summary (Drug prescriptions) - PITUITARY GONADOTROPIN INHIBITOR Min frequency (days) | 1.000000 | DISCARD |
| 1-year summary (Drug prescriptions) - PITUITARY GONADOTROPIN INHIBITOR Prescription count | 1.000000 | DISCARD |
| 1-year summary (Drug prescriptions) - PLATELET-REDUCING AGENT Average frequency (days) | 1.000000 | DISCARD |
| 1-year summary (Drug prescriptions) - PLATELET-REDUCING AGENT Days since first prescription | 1.000000 | DISCARD |
| 1-year summary (Drug prescriptions) - PLATELET-REDUCING AGENT Days since last prescription | 1.000000 | DISCARD |
| 1-year summary (Drug prescriptions) - PLATELET-REDUCING AGENT Max frequency (days) | 1.000000 | DISCARD |
| 1-year summary (Drug prescriptions) - PLATELET-REDUCING AGENT Min frequency (days) | 1.000000 | DISCARD |
| 1-year summary (Drug prescriptions) - PLATELET-REDUCING AGENT Prescription count | 1.000000 | DISCARD |
| 1-year summary (Drug prescriptions) - PROGESTERON ANALOGUES Average frequency (days) | 1.000000 | DISCARD |
| 1-year summary (Drug prescriptions) - PROGESTERON ANALOGUES Max frequency (days) | 1.000000 | DISCARD |
| 1-year summary (Drug prescriptions) - PROGESTERON ANALOGUES Min frequency (days) | 1.000000 | DISCARD |
| 1-year summary (Drug prescriptions) - PROLACTINE INHIBITORS Average frequency (days) | 1.000000 | DISCARD |
| 1-year summary (Drug prescriptions) - PROLACTINE INHIBITORS Days since first prescription | 1.000000 | DISCARD |
| 1-year summary (Drug prescriptions) - PROLACTINE INHIBITORS Days since last prescription | 1.000000 | DISCARD |
| 1-year summary (Drug prescriptions) - PROLACTINE INHIBITORS Max frequency (days) | 1.000000 | DISCARD |
| 1-year summary (Drug prescriptions) - PROLACTINE INHIBITORS Min frequency (days) | 1.000000 | DISCARD |
| 1-year summary (Drug prescriptions) - PROLACTINE INHIBITORS Prescription count | 1.000000 | DISCARD |

| | | |
|---|---|---|
| 1-year summary (Drug prescriptions) - REVERSIBLE MAO Average frequency (days) | 1.000000 | DISCARD |
| 1-year summary (Drug prescriptions) - REVERSIBLE MAO Days since first prescription | 1.000000 | DISCARD |
| 1-year summary (Drug prescriptions) - REVERSIBLE MAO Days since last prescription | 1.000000 | DISCARD |
| 1-year summary (Drug prescriptions) - REVERSIBLE MAO Max frequency (days) | 1.000000 | DISCARD |
| 1-year summary (Drug prescriptions) - REVERSIBLE MAO Min frequency (days) | 1.000000 | DISCARD |
| 1-year summary (Drug prescriptions) - REVERSIBLE MAO Prescription count | 1.000000 | DISCARD |
| 1-year summary (Drug prescriptions) - SELECTIVE MAO INHIBITORS Average frequency (days) | 1.000000 | DISCARD |
| 1-year summary (Drug prescriptions) - SELECTIVE MAO INHIBITORS Days since first prescription | 1.000000 | DISCARD |
| 1-year summary (Drug prescriptions) - SELECTIVE MAO INHIBITORS Days since last prescription | 1.000000 | DISCARD |
| 1-year summary (Drug prescriptions) - SELECTIVE MAO INHIBITORS Max frequency (days) | 1.000000 | DISCARD |
| 1-year summary (Drug prescriptions) - SELECTIVE MAO INHIBITORS Min frequency (days) | 1.000000 | DISCARD |
| 1-year summary (Drug prescriptions) - SELECTIVE MAO INHIBITORS Prescription count | 1.000000 | DISCARD |
| 1-year summary (Drug prescriptions) - SELECTIVE ESTROGEN RECEPTOR MODULATOR Average frequency (days) | 1.000000 | DISCARD |
| 1-year summary (Drug prescriptions) - SELECTIVE ESTROGEN RECEPTOR MODULATOR Days since first prescription | 1.000000 | DISCARD |
| 1-year summary (Drug prescriptions) - SELECTIVE ESTROGEN RECEPTOR MODULATOR Days since last prescription | 1.000000 | DISCARD |
| 1-year summary (Drug prescriptions) - SELECTIVE ESTROGEN RECEPTOR MODULATOR Max frequency (days) | 1.000000 | DISCARD |
| 1-year summary (Drug prescriptions) - SELECTIVE ESTROGEN RECEPTOR MODULATOR Min frequency (days) | 1.000000 | DISCARD |
| 1-year summary (Drug prescriptions) - SELECTIVE ESTROGEN RECEPTOR MODULATOR Prescription count | 1.000000 | DISCARD |
| 1-year summary (Drug prescriptions) - SEROTONIN (5-HT) RECEPTOR AGONIST Average frequency (days) | 1.000000 | DISCARD |
| 1-year summary (Drug prescriptions) - SEROTONIN (5-HT) RECEPTOR AGONIST Max frequency (days) | 1.000000 | DISCARD |
| 1-year summary (Drug prescriptions) - SEROTONIN (5-HT) RECEPTOR AGONIST Min frequency (days) | 1.000000 | DISCARD |
| 1-year summary (Drug prescriptions) - SOMATOSTATIN ANALOGUES Average frequency (days) | 1.000000 | DISCARD |
| 1-year summary (Drug prescriptions) - SOMATOSTATIN ANALOGUES Days since first prescription | 1.000000 | DISCARD |
| 1-year summary (Drug prescriptions) - SOMATOSTATIN ANALOGUES Days since last prescription | 1.000000 | DISCARD |
| 1-year summary (Drug prescriptions) - SOMATOSTATIN ANALOGUES Max frequency (days) | 1.000000 | DISCARD |
| 1-year summary (Drug prescriptions) - SOMATOSTATIN ANALOGUES Min frequency (days) | 1.000000 | DISCARD |
| 1-year summary (Drug prescriptions) - SOMATOSTATIN ANALOGUES Prescription count | 1.000000 | DISCARD |
| 1-year summary (Drug prescriptions) - SUCCINIMIDE DERIVATIVES Average frequency (days) | 1.000000 | DISCARD |
| 1-year summary (Drug prescriptions) - SUCCINIMIDE DERIVATIVES Days since first prescription | 1.000000 | DISCARD |
| 1-year summary (Drug prescriptions) - SUCCINIMIDE DERIVATIVES Days since last prescription | 1.000000 | DISCARD |
| 1-year summary (Drug prescriptions) - SUCCINIMIDE DERIVATIVES Max frequency (days) | 1.000000 | DISCARD |
| 1-year summary (Drug prescriptions) - SUCCINIMIDE DERIVATIVES Min frequency (days) | 1.000000 | DISCARD |
| 1-year summary (Drug prescriptions) - SUCCINIMIDE DERIVATIVES Prescription count | 1.000000 | DISCARD |
| 1-year summary (Drug prescriptions) - SULFONES Average frequency (days) | 1.000000 | DISCARD |
| 1-year summary (Drug prescriptions) - SULFONES Days since first prescription | 1.000000 | DISCARD |
| 1-year summary (Drug prescriptions) - SULFONES Days since last prescription | 1.000000 | DISCARD |
| 1-year summary (Drug prescriptions) - SULFONES Max frequency (days) | 1.000000 | DISCARD |

| | | | |
|---|---|---|---|
| 1-year summary (Drug prescriptions) - SULFONES Min frequency (days) | | 1.000000 | DISCARD |
| 1-year summary (Drug prescriptions) - SULFONES Prescription count | | 1.000000 | DISCARD |
| 1-year summary (Drug prescriptions) - SYMPATHOMIMETIC Average frequency (days) | | 1.000000 | DISCARD |
| 1-year summary (Drug prescriptions) - SYMPATHOMIMETIC Days since first prescription | | 1.000000 | DISCARD |
| 1-year summary (Drug prescriptions) - SYMPATHOMIMETIC Days since last prescription | | 1.000000 | DISCARD |
| 1-year summary (Drug prescriptions) - SYMPATHOMIMETIC Max frequency (days) | | 1.000000 | DISCARD |
| 1-year summary (Drug prescriptions) - SYMPATHOMIMETIC Min frequency (days) | | 1.000000 | DISCARD |
| 1-year summary (Drug prescriptions) - SYMPATHOMIMETIC Prescription count | | 1.000000 | DISCARD |
| 1-year summary (Drug prescriptions) - SYNTHETIC ANTITHROMBOTIC AGENTS Days since first prescription | | 1.000000 | DISCARD |
| 1-year summary (Drug prescriptions) - SYNTHETIC ANTITHROMBOTIC AGENTS Days since last prescription | | 1.000000 | DISCARD |
| 1-year summary (Drug prescriptions) - SYNTHETIC ANTITHROMBOTIC AGENTS Prescription count | | 1.000000 | DISCARD |
| 1-year summary (Drug prescriptions) - URICOSURIC DRUGS Average frequency (days) | | 1.000000 | DISCARD |
| 1-year summary (Drug prescriptions) - URICOSURIC DRUGS Days since first prescription | | 1.000000 | DISCARD |
| 1-year summary (Drug prescriptions) - URICOSURIC DRUGS Days since last prescription | | 1.000000 | DISCARD |
| 1-year summary (Drug prescriptions) - URICOSURIC DRUGS Max frequency (days) | | 1.000000 | DISCARD |
| 1-year summary (Drug prescriptions) - URICOSURIC DRUGS Min frequency (days) | | 1.000000 | DISCARD |
| 1-year summary (Drug prescriptions) - URICOSURIC DRUGS Prescription count | | 1.000000 | DISCARD |
| 1-year summary (Drug prescriptions) - VACCINES Average frequency (days) | | 1.000000 | DISCARD |
| 1-year summary (Drug prescriptions) - VACCINES Max frequency (days) | | 1.000000 | DISCARD |
| 1-year summary (Drug prescriptions) - VACCINES Min frequency (days) | | 1.000000 | DISCARD |
| 1-year summary (Drug prescriptions) - VITAMIN A Average frequency (days) | | 1.000000 | DISCARD |
| 1-year summary (Drug prescriptions) - VITAMIN A Days since first prescription | | 1.000000 | DISCARD |
| 1-year summary (Drug prescriptions) - VITAMIN A Days since last prescription | | 1.000000 | DISCARD |
| 1-year summary (Drug prescriptions) - VITAMIN A Max frequency (days) | | 1.000000 | DISCARD |
| 1-year summary (Drug prescriptions) - VITAMIN A Min frequency (days) | | 1.000000 | DISCARD |
| 1-year summary (Drug prescriptions) - VITAMIN A Prescription count | | 1.000000 | DISCARD |
| 1-year summary (Drug prescriptions) - VITAMIN C Average frequency (days) | | 1.000000 | DISCARD |
| 1-year summary (Drug prescriptions) - VITAMIN C Days since first prescription | | 1.000000 | DISCARD |
| 1-year summary (Drug prescriptions) - VITAMIN C Days since last prescription | | 1.000000 | DISCARD |
| 1-year summary (Drug prescriptions) - VITAMIN C Max frequency (days) | | 1.000000 | DISCARD |
| 1-year summary (Drug prescriptions) - VITAMIN C Min frequency (days) | | 1.000000 | DISCARD |
| 1-year summary (Drug prescriptions) - VITAMIN C Prescription count | | 1.000000 | DISCARD |
| 1-year summary (Drug prescriptions) - VITAMIN E Average frequency (days) | | 1.000000 | DISCARD |
| 1-year summary (Drug prescriptions) - VITAMIN E Days since first prescription | | 1.000000 | DISCARD |
| 1-year summary (Drug prescriptions) - VITAMIN E Days since last prescription | | 1.000000 | DISCARD |
| 1-year summary (Drug prescriptions) - VITAMIN E Max frequency (days) | | 1.000000 | DISCARD |
| 1-year summary (Drug prescriptions) - VITAMIN E Min frequency (days) | | 1.000000 | DISCARD |

| | | | |
|---|---|---|---|
| 1-year summary (Drug prescriptions) - VITAMIN E Prescription count | | 1.000000 | DISCARD |
| 1-year summary (Drug prescriptions) - VITAMIN K ANALOGUES Average frequency (days) | | 1.000000 | DISCARD |
| 1-year summary (Drug prescriptions) - VITAMIN K ANALOGUES Days since first prescription | | 1.000000 | DISCARD |
| 1-year summary (Drug prescriptions) - VITAMIN K ANALOGUES Days since last prescription | | 1.000000 | DISCARD |
| 1-year summary (Drug prescriptions) - VITAMIN K ANALOGUES Max frequency (days) | | 1.000000 | DISCARD |
| 1-year summary (Drug prescriptions) - VITAMIN K ANALOGUES Min frequency (days) | | 1.000000 | DISCARD |
| 1-year summary (Drug prescriptions) - VITAMIN K ANALOGUES Prescription count | | 1.000000 | DISCARD |
| 1-year summary (Drug prescriptions) - WART AND ANTI-CORN AGENTS Days since first prescription | | 1.000000 | DISCARD |
| 1-year summary (Drug prescriptions) - WART AND ANTI-CORN AGENTS Days since last prescription | | 1.000000 | DISCARD |
| 1-year summary (Drug prescriptions) - WART AND ANTI-CORN AGENTS Prescription count | | 1.000000 | DISCARD |
| 1-year summary (Drug prescriptions) - WOUND CARE AND SCAR MANAGEMENT AGENTS Average frequency (days) | | 1.000000 | DISCARD |
| 1-year summary (Drug prescriptions) - WOUND CARE AND SCAR MANAGEMENT AGENTS Max frequency (days) | | 1.000000 | DISCARD |
| 1-year summary (Drug prescriptions) - WOUND CARE AND SCAR MANAGEMENT AGENTS Min frequency (days) | | 1.000000 | DISCARD |
| 1-year summary (Doctor visits) - COMMUNITY MEDICINE | | 1.000000 | DISCARD |
| 1-year summary (Doctor visits) - ORAL RADIOLOGY | | 1.000000 | DISCARD |
| 1-year summary (Doctor visits) - PAEDODONTICS | | 1.000000 | DISCARD |
| 1-year summary (Doctor visits) - PERIODONTICS | | 1.000000 | DISCARD |
| 1-year summary (Doctor visits) - PROSTHODONTICS | | 1.000000 | DISCARD |
| 1-year summary (Laboratory tests) - number of tests for Glucose^1H_post_dose_glucose | | 1.000000 | DISCARD |
| 1-year summary (Laboratory tests) - number of tests for Glucose^2H_post_dose_glucose | | 1.000000 | DISCARD |
| 1-year summary (Laboratory tests) - days since earliest test for Glucose^1H_post_dose_glucose | | 1.000000 | DISCARD |
| 1-year summary (Laboratory tests) - days since earliest test for Glucose^2H_post_dose_glucose | | 1.000000 | DISCARD |
| 1-year summary (Laboratory tests) - days since latest test for Glucose^1H_post_dose_glucose | | 1.000000 | DISCARD |
| 1-year summary (Laboratory tests) - days since latest test for Glucose^2H_post_dose_glucose | | 1.000000 | DISCARD |
| 1-year summary (Laboratory tests) - number of high values of Glucose^1H_post_dose_glucose | | 1.000000 | DISCARD |
| 1-year summary (Laboratory tests) - number of high values of Glucose^2H_post_dose_glucose | | 1.000000 | DISCARD |
| 1-year summary (Laboratory tests) - highest value of Glucose^1H_post_dose_glucose | | 1.000000 | DISCARD |
| 1-year summary (Laboratory tests) - highest value of Glucose^2H_post_dose_glucose | | 1.000000 | DISCARD |
| 1-year summary (Laboratory tests) - number of low values of Glucose^1H_post_dose_glucose | | 1.000000 | DISCARD |
| 1-year summary (Laboratory tests) - number of low values of Glucose^2H_post_dose_glucose | | 1.000000 | DISCARD |
| 1-year summary (Laboratory tests) - lowest value of Glucose^1H_post_dose_glucose | | 1.000000 | DISCARD |
| 1-year summary (Laboratory tests) - lowest value of Glucose^2H_post_dose_glucose | | 1.000000 | DISCARD |
| 1-year summary (Laboratory tests) - average value of Glucose^1H_post_dose_glucose | | 1.000000 | DISCARD |
| 1-year summary (Laboratory tests) - average value of Glucose^2H_post_dose_glucose | | 1.000000 | DISCARD |
| 1-year summary (Laboratory tests) - median value of Glucose^1H_post_dose_glucose | | 1.000000 | DISCARD |
| 1-year summary (Laboratory tests) - median value of Glucose^2H_post_dose_glucose | | 1.000000 | DISCARD |
| 1-year summary (Laboratory tests) - count of normal tests for Glucose^1H_post_dose_glucose | | 1.000000 | DISCARD |

| | | | |
|---|---|---|---|
| 1-year summary (Laboratory tests) - count of normal tests for Glucose^2H_post_dose_glucose | | 1.000000 | DISCARD |
| 2-year summary (Drug prescriptions) - ADRENALS Average frequency (days) | | 1.000000 | DISCARD |
| 2-year summary (Drug prescriptions) - ADRENALS Max frequency (days) | | 1.000000 | DISCARD |
| 2-year summary (Drug prescriptions) - ADRENALS Min frequency (days) | | 1.000000 | DISCARD |
| 2-year summary (Drug prescriptions) - ADRENERGIC AND ANTICHOLINERGIC BRONCHODILATORS Average frequency (days) | | 1.000000 | DISCARD |
| 2-year summary (Drug prescriptions) - ADRENERGIC AND ANTICHOLINERGIC BRONCHODILATORS Max frequency (days) | | 1.000000 | DISCARD |
| 2-year summary (Drug prescriptions) - ADRENERGIC AND ANTICHOLINERGIC BRONCHODILATORS Min frequency (days) | | 1.000000 | DISCARD |
| 2-year summary (Drug prescriptions) - ALCOHOL DEPENDENCE Average frequency (days) | | 1.000000 | DISCARD |
| 2-year summary (Drug prescriptions) - ALCOHOL DEPENDENCE Days since first prescription | | 1.000000 | DISCARD |
| 2-year summary (Drug prescriptions) - ALCOHOL DEPENDENCE Days since last prescription | | 1.000000 | DISCARD |
| 2-year summary (Drug prescriptions) - ALCOHOL DEPENDENCE Max frequency (days) | | 1.000000 | DISCARD |
| 2-year summary (Drug prescriptions) - ALCOHOL DEPENDENCE Min frequency (days) | | 1.000000 | DISCARD |
| 2-year summary (Drug prescriptions) - ALCOHOL DEPENDENCE Prescription count | | 1.000000 | DISCARD |
| 2-year summary (Drug prescriptions) - ALKALOIDS AND DERIVATIVES Average frequency (days) | | 1.000000 | DISCARD |
| 2-year summary (Drug prescriptions) - ALKALOIDS AND DERIVATIVES Days since first prescription | | 1.000000 | DISCARD |
| 2-year summary (Drug prescriptions) - ALKALOIDS AND DERIVATIVES Days since last prescription | | 1.000000 | DISCARD |
| 2-year summary (Drug prescriptions) - ALKALOIDS AND DERIVATIVES Max frequency (days) | | 1.000000 | DISCARD |
| 2-year summary (Drug prescriptions) - ALKALOIDS AND DERIVATIVES Min frequency (days) | | 1.000000 | DISCARD |
| 2-year summary (Drug prescriptions) - ALKALOIDS AND DERIVATIVES Prescription count | | 1.000000 | DISCARD |
| 2-year summary (Drug prescriptions) - ALLERGENIC EXTRACTS Average frequency (days) | | 1.000000 | DISCARD |
| 2-year summary (Drug prescriptions) - ALLERGENIC EXTRACTS Max frequency (days) | | 1.000000 | DISCARD |
| 2-year summary (Drug prescriptions) - ALLERGENIC EXTRACTS Min frequency (days) | | 1.000000 | DISCARD |
| 2-year summary (Drug prescriptions) - ALLERGY PREPARATIONS Average frequency (days) | | 1.000000 | DISCARD |
| 2-year summary (Drug prescriptions) - ALLERGY PREPARATIONS Days since first prescription | | 1.000000 | DISCARD |
| 2-year summary (Drug prescriptions) - ALLERGY PREPARATIONS Days since last prescription | | 1.000000 | DISCARD |
| 2-year summary (Drug prescriptions) - ALLERGY PREPARATIONS Max frequency (days) | | 1.000000 | DISCARD |
| 2-year summary (Drug prescriptions) - ALLERGY PREPARATIONS Min frequency (days) | | 1.000000 | DISCARD |
| 2-year summary (Drug prescriptions) - ALLERGY PREPARATIONS Prescription count | | 1.000000 | DISCARD |
| 2-year summary (Drug prescriptions) - ALPHA-2-ADRENERGIC AGONIST Average frequency (days) | | 1.000000 | DISCARD |
| 2-year summary (Drug prescriptions) - ALPHA-2-ADRENERGIC AGONIST Days since first prescription | | 1.000000 | DISCARD |
| 2-year summary (Drug prescriptions) - ALPHA-2-ADRENERGIC AGONIST Days since last prescription | | 1.000000 | DISCARD |
| 2-year summary (Drug prescriptions) - ALPHA-2-ADRENERGIC AGONIST Max frequency (days) | | 1.000000 | DISCARD |
| 2-year summary (Drug prescriptions) - ALPHA-2-ADRENERGIC AGONIST Min frequency (days) | | 1.000000 | DISCARD |
| 2-year summary (Drug prescriptions) - ALPHA-2-ADRENERGIC AGONIST Prescription count | | 1.000000 | DISCARD |
| 2-year summary (Drug prescriptions) - ANTI- HEPATITIS Average frequency (days) | | 1.000000 | DISCARD |
| 2-year summary (Drug prescriptions) - ANTI- HEPATITIS Max frequency (days) | | 1.000000 | DISCARD |
| 2-year summary (Drug prescriptions) - ANTI- HEPATITIS Min frequency (days) | | 1.000000 | DISCARD |

| | | |
|---|---|---|
| 2-year summary (Drug prescriptions) - ANTIBIOTICS: OTHER Average frequency (days) | 1.000000 | DISCARD |
| 2-year summary (Drug prescriptions) - ANTIBIOTICS: OTHER Max frequency (days) | 1.000000 | DISCARD |
| 2-year summary (Drug prescriptions) - ANTIBIOTICS: OTHER Min frequency (days) | 1.000000 | DISCARD |
| 2-year summary (Drug prescriptions) - ANTIDOTES Average frequency (days) | 1.000000 | DISCARD |
| 2-year summary (Drug prescriptions) - ANTIDOTES Days since first prescription | 1.000000 | DISCARD |
| 2-year summary (Drug prescriptions) - ANTIDOTES Days since last prescription | 1.000000 | DISCARD |
| 2-year summary (Drug prescriptions) - ANTIDOTES Max frequency (days) | 1.000000 | DISCARD |
| 2-year summary (Drug prescriptions) - ANTIDOTES Min frequency (days) | 1.000000 | DISCARD |
| 2-year summary (Drug prescriptions) - ANTIDOTES Prescription count | 1.000000 | DISCARD |
| 2-year summary (Drug prescriptions) - ANTIFIBRINOLYTIC AGENTS Days since first prescription | 1.000000 | DISCARD |
| 2-year summary (Drug prescriptions) - ANTIFIBRINOLYTIC AGENTS Days since last prescription | 1.000000 | DISCARD |
| 2-year summary (Drug prescriptions) - ANTIFIBRINOLYTIC AGENTS Prescription count | 1.000000 | DISCARD |
| 2-year summary (Drug prescriptions) - ANTIHISTAMINE Average frequency (days) | 1.000000 | DISCARD |
| 2-year summary (Drug prescriptions) - ANTIHISTAMINE Days since first prescription | 1.000000 | DISCARD |
| 2-year summary (Drug prescriptions) - ANTIHISTAMINE Days since last prescription | 1.000000 | DISCARD |
| 2-year summary (Drug prescriptions) - ANTIHISTAMINE Max frequency (days) | 1.000000 | DISCARD |
| 2-year summary (Drug prescriptions) - ANTIHISTAMINE Min frequency (days) | 1.000000 | DISCARD |
| 2-year summary (Drug prescriptions) - ANTIHISTAMINE Prescription count | 1.000000 | DISCARD |
| 2-year summary (Drug prescriptions) - ANTIHISTAMINICS Average frequency (days) | 1.000000 | DISCARD |
| 2-year summary (Drug prescriptions) - ANTIHISTAMINICS Days since first prescription | 1.000000 | DISCARD |
| 2-year summary (Drug prescriptions) - ANTIHISTAMINICS Days since last prescription | 1.000000 | DISCARD |
| 2-year summary (Drug prescriptions) - ANTIHISTAMINICS Max frequency (days) | 1.000000 | DISCARD |
| 2-year summary (Drug prescriptions) - ANTIHISTAMINICS Min frequency (days) | 1.000000 | DISCARD |
| 2-year summary (Drug prescriptions) - ANTIHISTAMINICS Prescription count | 1.000000 | DISCARD |
| 2-year summary (Drug prescriptions) - ANTINEOPLASTIC AND IMMUNOMODULATING AGENTS Average frequency (days) | 1.000000 | DISCARD |
| 2-year summary (Drug prescriptions) - ANTINEOPLASTIC AND IMMUNOMODULATING AGENTS Days since first prescription | 1.000000 | DISCARD |
| 2-year summary (Drug prescriptions) - ANTINEOPLASTIC AND IMMUNOMODULATING AGENTS Days since last prescription | 1.000000 | DISCARD |
| 2-year summary (Drug prescriptions) - ANTINEOPLASTIC AND IMMUNOMODULATING AGENTS Max frequency (days) | 1.000000 | DISCARD |
| 2-year summary (Drug prescriptions) - ANTINEOPLASTIC AND IMMUNOMODULATING AGENTS Min frequency (days) | 1.000000 | DISCARD |
| 2-year summary (Drug prescriptions) - ANTINEOPLASTIC AND IMMUNOMODULATING AGENTS Prescription count | 1.000000 | DISCARD |
| 2-year summary (Drug prescriptions) - ANTIPARKINSONIAN AGENTS Average frequency (days) | 1.000000 | DISCARD |
| 2-year summary (Drug prescriptions) - ANTIPARKINSONIAN AGENTS Days since first prescription | 1.000000 | DISCARD |
| 2-year summary (Drug prescriptions) - ANTIPARKINSONIAN AGENTS Days since last prescription | 1.000000 | DISCARD |
| 2-year summary (Drug prescriptions) - ANTIPARKINSONIAN AGENTS Max frequency (days) | 1.000000 | DISCARD |
| 2-year summary (Drug prescriptions) - ANTIPARKINSONIAN AGENTS Min frequency (days) | 1.000000 | DISCARD |
| 2-year summary (Drug prescriptions) - ANTIPARKINSONIAN AGENTS Prescription count | 1.000000 | DISCARD |
| 2-year summary (Drug prescriptions) - AZASPIRODECANEDIONE DERIVATIVES Average frequency (days) | 1.000000 | DISCARD |

| | | |
|---|---|---|
| 2-year summary (Drug prescriptions) - AZASPIRODECANEDIONE DERIVATIVES Days since first prescription | 1.000000 | DISCARD |
| 2-year summary (Drug prescriptions) - AZASPIRODECANEDIONE DERIVATIVES Days since last prescription | 1.000000 | DISCARD |
| 2-year summary (Drug prescriptions) - AZASPIRODECANEDIONE DERIVATIVES Max frequency (days) | 1.000000 | DISCARD |
| 2-year summary (Drug prescriptions) - AZASPIRODECANEDIONE DERIVATIVES Min frequency (days) | 1.000000 | DISCARD |
| 2-year summary (Drug prescriptions) - AZASPIRODECANEDIONE DERIVATIVES Prescription count | 1.000000 | DISCARD |
| 2-year summary (Drug prescriptions) - BIOLOGIC AGENT Average frequency (days) | 1.000000 | DISCARD |
| 2-year summary (Drug prescriptions) - BIOLOGIC AGENT Max frequency (days) | 1.000000 | DISCARD |
| 2-year summary (Drug prescriptions) - BIOLOGIC AGENT Min frequency (days) | 1.000000 | DISCARD |
| 2-year summary (Drug prescriptions) - BLOOD COAGULATION FACTORS Days since first prescription | 1.000000 | DISCARD |
| 2-year summary (Drug prescriptions) - BLOOD COAGULATION FACTORS Days since last prescription | 1.000000 | DISCARD |
| 2-year summary (Drug prescriptions) - BLOOD COAGULATION FACTORS Prescription count | 1.000000 | DISCARD |
| 2-year summary (Drug prescriptions) - CARDIOVASCULAR DRUGS: OTHER Average frequency (days) | 1.000000 | DISCARD |
| 2-year summary (Drug prescriptions) - CARDIOVASCULAR DRUGS: OTHER Days since first prescription | 1.000000 | DISCARD |
| 2-year summary (Drug prescriptions) - CARDIOVASCULAR DRUGS: OTHER Days since last prescription | 1.000000 | DISCARD |
| 2-year summary (Drug prescriptions) - CARDIOVASCULAR DRUGS: OTHER Max frequency (days) | 1.000000 | DISCARD |
| 2-year summary (Drug prescriptions) - CARDIOVASCULAR DRUGS: OTHER Min frequency (days) | 1.000000 | DISCARD |
| 2-year summary (Drug prescriptions) - CARDIOVASCULAR DRUGS: OTHER Prescription count | 1.000000 | DISCARD |
| 2-year summary (Drug prescriptions) - CENTRAL NERVOUS SYSTEM DRUGS: MISCELLANEOUS Average frequency (days) | 1.000000 | DISCARD |
| 2-year summary (Drug prescriptions) - CENTRAL NERVOUS SYSTEM DRUGS: MISCELLANEOUS Days since first prescription | 1.000000 | DISCARD |
| 2-year summary (Drug prescriptions) - CENTRAL NERVOUS SYSTEM DRUGS: MISCELLANEOUS Days since last prescription | 1.000000 | DISCARD |
| 2-year summary (Drug prescriptions) - CENTRAL NERVOUS SYSTEM DRUGS: MISCELLANEOUS Max frequency (days) | 1.000000 | DISCARD |
| 2-year summary (Drug prescriptions) - CENTRAL NERVOUS SYSTEM DRUGS: MISCELLANEOUS Min frequency (days) | 1.000000 | DISCARD |
| 2-year summary (Drug prescriptions) - CENTRAL NERVOUS SYSTEM DRUGS: MISCELLANEOUS Prescription count | 1.000000 | DISCARD |
| 2-year summary (Drug prescriptions) - CONTRACEPTIVES Average frequency (days) | 1.000000 | DISCARD |
| 2-year summary (Drug prescriptions) - CONTRACEPTIVES Max frequency (days) | 1.000000 | DISCARD |
| 2-year summary (Drug prescriptions) - CONTRACEPTIVES Min frequency (days) | 1.000000 | DISCARD |
| 2-year summary (Drug prescriptions) - DECONGESTANTS NASAL Days since first prescription | 1.000000 | DISCARD |
| 2-year summary (Drug prescriptions) - DECONGESTANTS NASAL Days since last prescription | 1.000000 | DISCARD |
| 2-year summary (Drug prescriptions) - DECONGESTANTS NASAL Prescription count | 1.000000 | DISCARD |
| 2-year summary (Drug prescriptions) - DIRECT SKELETAL MUSCLE RELAXANT Average frequency (days) | 1.000000 | DISCARD |
| 2-year summary (Drug prescriptions) - DIRECT SKELETAL MUSCLE RELAXANT Days since first prescription | 1.000000 | DISCARD |
| 2-year summary (Drug prescriptions) - DIRECT SKELETAL MUSCLE RELAXANT Days since last prescription | 1.000000 | DISCARD |
| 2-year summary (Drug prescriptions) - DIRECT SKELETAL MUSCLE RELAXANT Max frequency (days) | 1.000000 | DISCARD |
| 2-year summary (Drug prescriptions) - DIRECT SKELETAL MUSCLE RELAXANT Min frequency (days) | 1.000000 | DISCARD |
| 2-year summary (Drug prescriptions) - DIRECT SKELETAL MUSCLE RELAXANT Prescription count | 1.000000 | DISCARD |
| 2-year summary (Drug prescriptions) - EMETICS Average frequency (days) | 1.000000 | DISCARD |
| 2-year summary (Drug prescriptions) - EMETICS Days since first prescription | 1.000000 | DISCARD |

| | | |
|---|---|---|
| 2-year summary (Drug prescriptions) - EMETICS Days since last prescription | 1.000000 | DISCARD |
| 2-year summary (Drug prescriptions) - EMETICS Max frequency (days) | 1.000000 | DISCARD |
| 2-year summary (Drug prescriptions) - EMETICS Min frequency (days) | 1.000000 | DISCARD |
| 2-year summary (Drug prescriptions) - EMETICS Prescription count | 1.000000 | DISCARD |
| 2-year summary (Drug prescriptions) - EMOLLIENTS, DEMULCENTS AND PROTECTANTS Average frequency (days) | 1.000000 | DISCARD |
| 2-year summary (Drug prescriptions) - EMOLLIENTS, DEMULCENTS AND PROTECTANTS Max frequency (days) | 1.000000 | DISCARD |
| 2-year summary (Drug prescriptions) - EMOLLIENTS, DEMULCENTS AND PROTECTANTS Min frequency (days) | 1.000000 | DISCARD |
| 2-year summary (Drug prescriptions) - ERECTILE DYSFUNCTION Average frequency (days) | 1.000000 | DISCARD |
| 2-year summary (Drug prescriptions) - ERECTILE DYSFUNCTION Days since first prescription | 1.000000 | DISCARD |
| 2-year summary (Drug prescriptions) - ERECTILE DYSFUNCTION Days since last prescription | 1.000000 | DISCARD |
| 2-year summary (Drug prescriptions) - ERECTILE DYSFUNCTION Max frequency (days) | 1.000000 | DISCARD |
| 2-year summary (Drug prescriptions) - ERECTILE DYSFUNCTION Min frequency (days) | 1.000000 | DISCARD |
| 2-year summary (Drug prescriptions) - ERECTILE DYSFUNCTION Prescription count | 1.000000 | DISCARD |
| 2-year summary (Drug prescriptions) - FIBRINOGEN-PLATELET BINDING INHIBITORS Average frequency (days) | 1.000000 | DISCARD |
| 2-year summary (Drug prescriptions) - FIBRINOGEN-PLATELET BINDING INHIBITORS Days since first prescription | 1.000000 | DISCARD |
| 2-year summary (Drug prescriptions) - FIBRINOGEN-PLATELET BINDING INHIBITORS Days since last prescription | 1.000000 | DISCARD |
| 2-year summary (Drug prescriptions) - FIBRINOGEN-PLATELET BINDING INHIBITORS Max frequency (days) | 1.000000 | DISCARD |
| 2-year summary (Drug prescriptions) - FIBRINOGEN-PLATELET BINDING INHIBITORS Min frequency (days) | 1.000000 | DISCARD |
| 2-year summary (Drug prescriptions) - FIBRINOGEN-PLATELET BINDING INHIBITORS Prescription count | 1.000000 | DISCARD |
| 2-year summary (Drug prescriptions) - GASTROINTESTINAL ANTISPASMODICS Average frequency (days) | 1.000000 | DISCARD |
| 2-year summary (Drug prescriptions) - GASTROINTESTINAL ANTISPASMODICS Max frequency (days) | 1.000000 | DISCARD |
| 2-year summary (Drug prescriptions) - GASTROINTESTINAL ANTISPASMODICS Min frequency (days) | 1.000000 | DISCARD |
| 2-year summary (Drug prescriptions) - GLYCOPEPTIDES Average frequency (days) | 1.000000 | DISCARD |
| 2-year summary (Drug prescriptions) - GLYCOPEPTIDES Max frequency (days) | 1.000000 | DISCARD |
| 2-year summary (Drug prescriptions) - GLYCOPEPTIDES Min frequency (days) | 1.000000 | DISCARD |
| 2-year summary (Drug prescriptions) - GROWTH STIMULANT HORMONE Average frequency (days) | 1.000000 | DISCARD |
| 2-year summary (Drug prescriptions) - GROWTH STIMULANT HORMONE Days since first prescription | 1.000000 | DISCARD |
| 2-year summary (Drug prescriptions) - GROWTH STIMULANT HORMONE Days since last prescription | 1.000000 | DISCARD |
| 2-year summary (Drug prescriptions) - GROWTH STIMULANT HORMONE Max frequency (days) | 1.000000 | DISCARD |
| 2-year summary (Drug prescriptions) - GROWTH STIMULANT HORMONE Min frequency (days) | 1.000000 | DISCARD |
| 2-year summary (Drug prescriptions) - GROWTH STIMULANT HORMONE Prescription count | 1.000000 | DISCARD |
| 2-year summary (Drug prescriptions) - HEAVY METAL ANTAGONISTS Average frequency (days) | 1.000000 | DISCARD |
| 2-year summary (Drug prescriptions) - HEAVY METAL ANTAGONISTS Days since first prescription | 1.000000 | DISCARD |
| 2-year summary (Drug prescriptions) - HEAVY METAL ANTAGONISTS Days since last prescription | 1.000000 | DISCARD |
| 2-year summary (Drug prescriptions) - HEAVY METAL ANTAGONISTS Max frequency (days) | 1.000000 | DISCARD |
| 2-year summary (Drug prescriptions) - HEAVY METAL ANTAGONISTS Min frequency (days) | 1.000000 | DISCARD |
| 2-year summary (Drug prescriptions) - HEAVY METAL ANTAGONISTS Prescription count | 1.000000 | DISCARD |

| | | |
|---|---|---|
| 2-year summary (Drug prescriptions) - HORMONE ANTAGONISTS Average frequency (days) | 1.000000 | DISCARD |
| 2-year summary (Drug prescriptions) - HORMONE ANTAGONISTS Max frequency (days) | 1.000000 | DISCARD |
| 2-year summary (Drug prescriptions) - HORMONE ANTAGONISTS Min frequency (days) | 1.000000 | DISCARD |
| 2-year summary (Drug prescriptions) - HORMONES ANALOGUES Average frequency (days) | 1.000000 | DISCARD |
| 2-year summary (Drug prescriptions) - HORMONES ANALOGUES Days since first prescription | 1.000000 | DISCARD |
| 2-year summary (Drug prescriptions) - HORMONES ANALOGUES Days since last prescription | 1.000000 | DISCARD |
| 2-year summary (Drug prescriptions) - HORMONES ANALOGUES Max frequency (days) | 1.000000 | DISCARD |
| 2-year summary (Drug prescriptions) - HORMONES ANALOGUES Min frequency (days) | 1.000000 | DISCARD |
| 2-year summary (Drug prescriptions) - HORMONES ANALOGUES Prescription count | 1.000000 | DISCARD |
| 2-year summary (Drug prescriptions) - INTERLEUKIN INHIBITOR Average frequency (days) | 1.000000 | DISCARD |
| 2-year summary (Drug prescriptions) - INTERLEUKIN INHIBITOR Days since first prescription | 1.000000 | DISCARD |
| 2-year summary (Drug prescriptions) - INTERLEUKIN INHIBITOR Days since last prescription | 1.000000 | DISCARD |
| 2-year summary (Drug prescriptions) - INTERLEUKIN INHIBITOR Max frequency (days) | 1.000000 | DISCARD |
| 2-year summary (Drug prescriptions) - INTERLEUKIN INHIBITOR Min frequency (days) | 1.000000 | DISCARD |
| 2-year summary (Drug prescriptions) - INTERLEUKIN INHIBITOR Prescription count | 1.000000 | DISCARD |
| 2-year summary (Drug prescriptions) - IRREVERSIBLE MAO Average frequency (days) | 1.000000 | DISCARD |
| 2-year summary (Drug prescriptions) - IRREVERSIBLE MAO Days since first prescription | 1.000000 | DISCARD |
| 2-year summary (Drug prescriptions) - IRREVERSIBLE MAO Days since last prescription | 1.000000 | DISCARD |
| 2-year summary (Drug prescriptions) - IRREVERSIBLE MAO Max frequency (days) | 1.000000 | DISCARD |
| 2-year summary (Drug prescriptions) - IRREVERSIBLE MAO Min frequency (days) | 1.000000 | DISCARD |
| 2-year summary (Drug prescriptions) - IRREVERSIBLE MAO Prescription count | 1.000000 | DISCARD |
| 2-year summary (Drug prescriptions) - LEUKOTRIENE RECEPTOR ANTAGONISTS Average frequency (days) | 1.000000 | DISCARD |
| 2-year summary (Drug prescriptions) - LEUKOTRIENE RECEPTOR ANTAGONISTS Days since first prescription | 1.000000 | DISCARD |
| 2-year summary (Drug prescriptions) - LEUKOTRIENE RECEPTOR ANTAGONISTS Days since last prescription | 1.000000 | DISCARD |
| 2-year summary (Drug prescriptions) - LEUKOTRIENE RECEPTOR ANTAGONISTS Max frequency (days) | 1.000000 | DISCARD |
| 2-year summary (Drug prescriptions) - LEUKOTRIENE RECEPTOR ANTAGONISTS Min frequency (days) | 1.000000 | DISCARD |
| 2-year summary (Drug prescriptions) - LEUKOTRIENE RECEPTOR ANTAGONISTS Prescription count | 1.000000 | DISCARD |
| 2-year summary (Drug prescriptions) - LOCAL ANESTHETICS INJ Average frequency (days) | 1.000000 | DISCARD |
| 2-year summary (Drug prescriptions) - LOCAL ANESTHETICS INJ Days since first prescription | 1.000000 | DISCARD |
| 2-year summary (Drug prescriptions) - LOCAL ANESTHETICS INJ Days since last prescription | 1.000000 | DISCARD |
| 2-year summary (Drug prescriptions) - LOCAL ANESTHETICS INJ Max frequency (days) | 1.000000 | DISCARD |
| 2-year summary (Drug prescriptions) - LOCAL ANESTHETICS INJ Min frequency (days) | 1.000000 | DISCARD |
| 2-year summary (Drug prescriptions) - LOCAL ANESTHETICS INJ Prescription count | 1.000000 | DISCARD |
| 2-year summary (Drug prescriptions) - MEMBRANE STABILISING DRUGS TYPE A Average frequency (days) | 1.000000 | DISCARD |
| 2-year summary (Drug prescriptions) - MEMBRANE STABILISING DRUGS TYPE A Days since first prescription | 1.000000 | DISCARD |
| 2-year summary (Drug prescriptions) - MEMBRANE STABILISING DRUGS TYPE A Days since last prescription | 1.000000 | DISCARD |
| 2-year summary (Drug prescriptions) - MEMBRANE STABILISING DRUGS TYPE A Max frequency (days) | 1.000000 | DISCARD |

| | | | |
|---|---|---|---|
| 2-year summary (Drug prescriptions) - MEMBRANE STABILISING DRUGS TYPE A Min frequency (days) | | 1.000000 | DISCARD |
| 2-year summary (Drug prescriptions) - MEMBRANE STABILISING DRUGS TYPE A Prescription count | | 1.000000 | DISCARD |
| 2-year summary (Drug prescriptions) - MISCELLANEOUS ANTI-INFECTIVES Average frequency (days) | | 1.000000 | DISCARD |
| 2-year summary (Drug prescriptions) - MISCELLANEOUS ANTI-INFECTIVES Days since first prescription | | 1.000000 | DISCARD |
| 2-year summary (Drug prescriptions) - MISCELLANEOUS ANTI-INFECTIVES Days since last prescription | | 1.000000 | DISCARD |
| 2-year summary (Drug prescriptions) - MISCELLANEOUS ANTI-INFECTIVES Max frequency (days) | | 1.000000 | DISCARD |
| 2-year summary (Drug prescriptions) - MISCELLANEOUS ANTI-INFECTIVES Min frequency (days) | | 1.000000 | DISCARD |
| 2-year summary (Drug prescriptions) - MISCELLANEOUS ANTI-INFECTIVES Prescription count | | 1.000000 | DISCARD |
| 2-year summary (Drug prescriptions) - MULTIVITAMINS Average frequency (days) | | 1.000000 | DISCARD |
| 2-year summary (Drug prescriptions) - MULTIVITAMINS Days since first prescription | | 1.000000 | DISCARD |
| 2-year summary (Drug prescriptions) - MULTIVITAMINS Days since last prescription | | 1.000000 | DISCARD |
| 2-year summary (Drug prescriptions) - MULTIVITAMINS Max frequency (days) | | 1.000000 | DISCARD |
| 2-year summary (Drug prescriptions) - MULTIVITAMINS Min frequency (days) | | 1.000000 | DISCARD |
| 2-year summary (Drug prescriptions) - MULTIVITAMINS Prescription count | | 1.000000 | DISCARD |
| 2-year summary (Drug prescriptions) - NEUROMUSCULAR BLOCKING AGENT Average frequency (days) | | 1.000000 | DISCARD |
| 2-year summary (Drug prescriptions) - NEUROMUSCULAR BLOCKING AGENT Days since first prescription | | 1.000000 | DISCARD |
| 2-year summary (Drug prescriptions) - NEUROMUSCULAR BLOCKING AGENT Days since last prescription | | 1.000000 | DISCARD |
| 2-year summary (Drug prescriptions) - NEUROMUSCULAR BLOCKING AGENT Max frequency (days) | | 1.000000 | DISCARD |
| 2-year summary (Drug prescriptions) - NEUROMUSCULAR BLOCKING AGENT Min frequency (days) | | 1.000000 | DISCARD |
| 2-year summary (Drug prescriptions) - NEUROMUSCULAR BLOCKING AGENT Prescription count | | 1.000000 | DISCARD |
| 2-year summary (Drug prescriptions) - OTHER ANTIPARKINSONIAN AGENTS Average frequency (days) | | 1.000000 | DISCARD |
| 2-year summary (Drug prescriptions) - OTHER ANTIPARKINSONIAN AGENTS Days since first prescription | | 1.000000 | DISCARD |
| 2-year summary (Drug prescriptions) - OTHER ANTIPARKINSONIAN AGENTS Days since last prescription | | 1.000000 | DISCARD |
| 2-year summary (Drug prescriptions) - OTHER ANTIPARKINSONIAN AGENTS Max frequency (days) | | 1.000000 | DISCARD |
| 2-year summary (Drug prescriptions) - OTHER ANTIPARKINSONIAN AGENTS Min frequency (days) | | 1.000000 | DISCARD |
| 2-year summary (Drug prescriptions) - OTHER ANTIPARKINSONIAN AGENTS Prescription count | | 1.000000 | DISCARD |
| 2-year summary (Drug prescriptions) - OTHER HORMONAL CONTRACEPTIVES Average frequency (days) | | 1.000000 | DISCARD |
| 2-year summary (Drug prescriptions) - OTHER HORMONAL CONTRACEPTIVES Max frequency (days) | | 1.000000 | DISCARD |
| 2-year summary (Drug prescriptions) - OTHER HORMONAL CONTRACEPTIVES Min frequency (days) | | 1.000000 | DISCARD |
| 2-year summary (Drug prescriptions) - OTHER MIGRAINES THERAPY Average frequency (days) | | 1.000000 | DISCARD |
| 2-year summary (Drug prescriptions) - OTHER MIGRAINES THERAPY Days since first prescription | | 1.000000 | DISCARD |
| 2-year summary (Drug prescriptions) - OTHER MIGRAINES THERAPY Days since last prescription | | 1.000000 | DISCARD |
| 2-year summary (Drug prescriptions) - OTHER MIGRAINES THERAPY Max frequency (days) | | 1.000000 | DISCARD |
| 2-year summary (Drug prescriptions) - OTHER MIGRAINES THERAPY Min frequency (days) | | 1.000000 | DISCARD |
| 2-year summary (Drug prescriptions) - OTHER MIGRAINES THERAPY Prescription count | | 1.000000 | DISCARD |
| 2-year summary (Drug prescriptions) - PARASYMPATHOMIMETIC (CHOLINERGIC) Average frequency (days) | | 1.000000 | DISCARD |
| 2-year summary (Drug prescriptions) - PARASYMPATHOMIMETIC (CHOLINERGIC) Days since first prescription | | 1.000000 | DISCARD |

| | | |
|---|---|---|
| 2-year summary (Drug prescriptions) - PARASYMPATHOMIMETIC (CHOLINERGIC) Days since last prescription | 1.000000 | DISCARD |
| 2-year summary (Drug prescriptions) - PARASYMPATHOMIMETIC (CHOLINERGIC) Max frequency (days) | 1.000000 | DISCARD |
| 2-year summary (Drug prescriptions) - PARASYMPATHOMIMETIC (CHOLINERGIC) Min frequency (days) | 1.000000 | DISCARD |
| 2-year summary (Drug prescriptions) - PARASYMPATHOMIMETIC (CHOLINERGIC) Prescription count | 1.000000 | DISCARD |
| 2-year summary (Drug prescriptions) - PARASYMPATHOMIMETIC AGENTS Average frequency (days) | 1.000000 | DISCARD |
| 2-year summary (Drug prescriptions) - PARASYMPATHOMIMETIC AGENTS Days since first prescription | 1.000000 | DISCARD |
| 2-year summary (Drug prescriptions) - PARASYMPATHOMIMETIC AGENTS Days since last prescription | 1.000000 | DISCARD |
| 2-year summary (Drug prescriptions) - PARASYMPATHOMIMETIC AGENTS Max frequency (days) | 1.000000 | DISCARD |
| 2-year summary (Drug prescriptions) - PARASYMPATHOMIMETIC AGENTS Min frequency (days) | 1.000000 | DISCARD |
| 2-year summary (Drug prescriptions) - PARASYMPATHOMIMETIC AGENTS Prescription count | 1.000000 | DISCARD |
| 2-year summary (Drug prescriptions) - PARATHYROID AGENTS Average frequency (days) | 1.000000 | DISCARD |
| 2-year summary (Drug prescriptions) - PARATHYROID AGENTS Days since first prescription | 1.000000 | DISCARD |
| 2-year summary (Drug prescriptions) - PARATHYROID AGENTS Days since last prescription | 1.000000 | DISCARD |
| 2-year summary (Drug prescriptions) - PARATHYROID AGENTS Max frequency (days) | 1.000000 | DISCARD |
| 2-year summary (Drug prescriptions) - PARATHYROID AGENTS Min frequency (days) | 1.000000 | DISCARD |
| 2-year summary (Drug prescriptions) - PARATHYROID AGENTS Prescription count | 1.000000 | DISCARD |
| 2-year summary (Drug prescriptions) - PITUITARY FUNCTION DIAGNOSTIC Average frequency (days) | 1.000000 | DISCARD |
| 2-year summary (Drug prescriptions) - PITUITARY FUNCTION DIAGNOSTIC Days since first prescription | 1.000000 | DISCARD |
| 2-year summary (Drug prescriptions) - PITUITARY FUNCTION DIAGNOSTIC Days since last prescription | 1.000000 | DISCARD |
| 2-year summary (Drug prescriptions) - PITUITARY FUNCTION DIAGNOSTIC Max frequency (days) | 1.000000 | DISCARD |
| 2-year summary (Drug prescriptions) - PITUITARY FUNCTION DIAGNOSTIC Min frequency (days) | 1.000000 | DISCARD |
| 2-year summary (Drug prescriptions) - PITUITARY FUNCTION DIAGNOSTIC Prescription count | 1.000000 | DISCARD |
| 2-year summary (Drug prescriptions) - PITUITARY GONADOTROPIN INHIBITOR Average frequency (days) | 1.000000 | DISCARD |
| 2-year summary (Drug prescriptions) - PITUITARY GONADOTROPIN INHIBITOR Days since first prescription | 1.000000 | DISCARD |
| 2-year summary (Drug prescriptions) - PITUITARY GONADOTROPIN INHIBITOR Days since last prescription | 1.000000 | DISCARD |
| 2-year summary (Drug prescriptions) - PITUITARY GONADOTROPIN INHIBITOR Max frequency (days) | 1.000000 | DISCARD |
| 2-year summary (Drug prescriptions) - PITUITARY GONADOTROPIN INHIBITOR Min frequency (days) | 1.000000 | DISCARD |
| 2-year summary (Drug prescriptions) - PITUITARY GONADOTROPIN INHIBITOR Prescription count | 1.000000 | DISCARD |
| 2-year summary (Drug prescriptions) - PLATELET-REDUCING AGENT Average frequency (days) | 1.000000 | DISCARD |
| 2-year summary (Drug prescriptions) - PLATELET-REDUCING AGENT Days since first prescription | 1.000000 | DISCARD |
| 2-year summary (Drug prescriptions) - PLATELET-REDUCING AGENT Days since last prescription | 1.000000 | DISCARD |
| 2-year summary (Drug prescriptions) - PLATELET-REDUCING AGENT Max frequency (days) | 1.000000 | DISCARD |
| 2-year summary (Drug prescriptions) - PLATELET-REDUCING AGENT Min frequency (days) | 1.000000 | DISCARD |
| 2-year summary (Drug prescriptions) - PLATELET-REDUCING AGENT Prescription count | 1.000000 | DISCARD |
| 2-year summary (Drug prescriptions) - PROFESSIONAL CARE SERVICE Days since first prescription | 1.000000 | DISCARD |
| 2-year summary (Drug prescriptions) - PROFESSIONAL CARE SERVICE Days since last prescription | 1.000000 | DISCARD |
| 2-year summary (Drug prescriptions) - PROFESSIONAL CARE SERVICE Prescription count | 1.000000 | DISCARD |

| | | | |
|---|---|---|---|
| 2-year summary (Drug prescriptions) - PROLACTINE INHIBITORS Average frequency (days) | | 1.000000 | DISCARD |
| 2-year summary (Drug prescriptions) - PROLACTINE INHIBITORS Days since first prescription | | 1.000000 | DISCARD |
| 2-year summary (Drug prescriptions) - PROLACTINE INHIBITORS Days since last prescription | | 1.000000 | DISCARD |
| 2-year summary (Drug prescriptions) - PROLACTINE INHIBITORS Max frequency (days) | | 1.000000 | DISCARD |
| 2-year summary (Drug prescriptions) - PROLACTINE INHIBITORS Min frequency (days) | | 1.000000 | DISCARD |
| 2-year summary (Drug prescriptions) - PROLACTINE INHIBITORS Prescription count | | 1.000000 | DISCARD |
| 2-year summary (Drug prescriptions) - PSYCHOTROPICS: OTHER Days since first prescription | | 1.000000 | DISCARD |
| 2-year summary (Drug prescriptions) - PSYCHOTROPICS: OTHER Days since last prescription | | 1.000000 | DISCARD |
| 2-year summary (Drug prescriptions) - PSYCHOTROPICS: OTHER Prescription count | | 1.000000 | DISCARD |
| 2-year summary (Drug prescriptions) - RESPIRATORY DEVICE Days since first prescription | | 1.000000 | DISCARD |
| 2-year summary (Drug prescriptions) - RESPIRATORY DEVICE Days since last prescription | | 1.000000 | DISCARD |
| 2-year summary (Drug prescriptions) - RESPIRATORY DEVICE Prescription count | | 1.000000 | DISCARD |
| 2-year summary (Drug prescriptions) - REVERSIBLE MAO Average frequency (days) | | 1.000000 | DISCARD |
| 2-year summary (Drug prescriptions) - REVERSIBLE MAO Days since first prescription | | 1.000000 | DISCARD |
| 2-year summary (Drug prescriptions) - REVERSIBLE MAO Days since last prescription | | 1.000000 | DISCARD |
| 2-year summary (Drug prescriptions) - REVERSIBLE MAO Max frequency (days) | | 1.000000 | DISCARD |
| 2-year summary (Drug prescriptions) - REVERSIBLE MAO Min frequency (days) | | 1.000000 | DISCARD |
| 2-year summary (Drug prescriptions) - REVERSIBLE MAO Prescription count | | 1.000000 | DISCARD |
| 2-year summary (Drug prescriptions) - SELECTIVE MAO INHIBITORS Average frequency (days) |  | 1.000000 | DISCARD |
| 2-year summary (Drug prescriptions) - SELECTIVE MAO INHIBITORS Days since first prescription |  | 1.000000 | DISCARD |
| 2-year summary (Drug prescriptions) - SELECTIVE MAO INHIBITORS Days since last prescription |  | 1.000000 | DISCARD |
| 2-year summary (Drug prescriptions) - SELECTIVE MAO INHIBITORS Max frequency (days) |  | 1.000000 | DISCARD |
| 2-year summary (Drug prescriptions) - SELECTIVE MAO INHIBITORS Min frequency (days) |  | 1.000000 | DISCARD |
| 2-year summary (Drug prescriptions) - SELECTIVE MAO INHIBITORS Prescription count |  | 1.000000 | DISCARD |
| 2-year summary (Drug prescriptions) - SELECTIVE ESTROGEN RECEPTOR MODULATOR Average frequency (days) | | 1.000000 | DISCARD |
| 2-year summary (Drug prescriptions) - SELECTIVE ESTROGEN RECEPTOR MODULATOR Days since first prescription | | 1.000000 | DISCARD |
| 2-year summary (Drug prescriptions) - SELECTIVE ESTROGEN RECEPTOR MODULATOR Days since last prescription | | 1.000000 | DISCARD |
| 2-year summary (Drug prescriptions) - SELECTIVE ESTROGEN RECEPTOR MODULATOR Max frequency (days) | | 1.000000 | DISCARD |
| 2-year summary (Drug prescriptions) - SELECTIVE ESTROGEN RECEPTOR MODULATOR Min frequency (days) | | 1.000000 | DISCARD |
| 2-year summary (Drug prescriptions) - SELECTIVE ESTROGEN RECEPTOR MODULATOR Prescription count | | 1.000000 | DISCARD |
| 2-year summary (Drug prescriptions) - SOMATOSTATIN ANALOGUES Average frequency (days) | | 1.000000 | DISCARD |
| 2-year summary (Drug prescriptions) - SOMATOSTATIN ANALOGUES Days since first prescription | | 1.000000 | DISCARD |
| 2-year summary (Drug prescriptions) - SOMATOSTATIN ANALOGUES Days since last prescription | | 1.000000 | DISCARD |
| 2-year summary (Drug prescriptions) - SOMATOSTATIN ANALOGUES Max frequency (days) | | 1.000000 | DISCARD |
| 2-year summary (Drug prescriptions) - SOMATOSTATIN ANALOGUES Min frequency (days) | | 1.000000 | DISCARD |
| 2-year summary (Drug prescriptions) - SOMATOSTATIN ANALOGUES Prescription count | | 1.000000 | DISCARD |
| 2-year summary (Drug prescriptions) - SUCCINIMIDE DERIVATIVES Average frequency (days) | | 1.000000 | DISCARD |

| | | |
|---|---|---|
| 2-year summary (Drug prescriptions) - SUCCINIMIDE DERIVATIVES Days since first prescription | 1.000000 | DISCARD |
| 2-year summary (Drug prescriptions) - SUCCINIMIDE DERIVATIVES Days since last prescription | 1.000000 | DISCARD |
| 2-year summary (Drug prescriptions) - SUCCINIMIDE DERIVATIVES Max frequency (days) | 1.000000 | DISCARD |
| 2-year summary (Drug prescriptions) - SUCCINIMIDE DERIVATIVES Min frequency (days) | 1.000000 | DISCARD |
| 2-year summary (Drug prescriptions) - SUCCINIMIDE DERIVATIVES Prescription count | 1.000000 | DISCARD |
| 2-year summary (Drug prescriptions) - SULFONES Average frequency (days) | 1.000000 | DISCARD |
| 2-year summary (Drug prescriptions) - SULFONES Days since first prescription | 1.000000 | DISCARD |
| 2-year summary (Drug prescriptions) - SULFONES Days since last prescription | 1.000000 | DISCARD |
| 2-year summary (Drug prescriptions) - SULFONES Max frequency (days) | 1.000000 | DISCARD |
| 2-year summary (Drug prescriptions) - SULFONES Min frequency (days) | 1.000000 | DISCARD |
| 2-year summary (Drug prescriptions) - SULFONES Prescription count | 1.000000 | DISCARD |
| 2-year summary (Drug prescriptions) - SYMPATHOMIMETIC Average frequency (days) | 1.000000 | DISCARD |
| 2-year summary (Drug prescriptions) - SYMPATHOMIMETIC Days since first prescription | 1.000000 | DISCARD |
| 2-year summary (Drug prescriptions) - SYMPATHOMIMETIC Days since last prescription | 1.000000 | DISCARD |
| 2-year summary (Drug prescriptions) - SYMPATHOMIMETIC Max frequency (days) | 1.000000 | DISCARD |
| 2-year summary (Drug prescriptions) - SYMPATHOMIMETIC Min frequency (days) | 1.000000 | DISCARD |
| 2-year summary (Drug prescriptions) - SYMPATHOMIMETIC Prescription count | 1.000000 | DISCARD |
| 2-year summary (Drug prescriptions) - SYNTHETIC ANTITHROMBOTIC AGENTS Days since first prescription | 1.000000 | DISCARD |
| 2-year summary (Drug prescriptions) - SYNTHETIC ANTITHROMBOTIC AGENTS Days since last prescription | 1.000000 | DISCARD |
| 2-year summary (Drug prescriptions) - SYNTHETIC ANTITHROMBOTIC AGENTS Prescription count | 1.000000 | DISCARD |
| 2-year summary (Drug prescriptions) - URICOSURIC DRUGS Average frequency (days) | 1.000000 | DISCARD |
| 2-year summary (Drug prescriptions) - URICOSURIC DRUGS Days since first prescription | 1.000000 | DISCARD |
| 2-year summary (Drug prescriptions) - URICOSURIC DRUGS Days since last prescription | 1.000000 | DISCARD |
| 2-year summary (Drug prescriptions) - URICOSURIC DRUGS Max frequency (days) | 1.000000 | DISCARD |
| 2-year summary (Drug prescriptions) - URICOSURIC DRUGS Min frequency (days) | 1.000000 | DISCARD |
| 2-year summary (Drug prescriptions) - URICOSURIC DRUGS Prescription count | 1.000000 | DISCARD |
| 2-year summary (Drug prescriptions) - VITAMIN A Average frequency (days) | 1.000000 | DISCARD |
| 2-year summary (Drug prescriptions) - VITAMIN A Max frequency (days) | 1.000000 | DISCARD |
| 2-year summary (Drug prescriptions) - VITAMIN A Min frequency (days) | 1.000000 | DISCARD |
| 2-year summary (Drug prescriptions) - VITAMIN C Average frequency (days) | 1.000000 | DISCARD |
| 2-year summary (Drug prescriptions) - VITAMIN C Days since first prescription | 1.000000 | DISCARD |
| 2-year summary (Drug prescriptions) - VITAMIN C Days since last prescription | 1.000000 | DISCARD |
| 2-year summary (Drug prescriptions) - VITAMIN C Max frequency (days) | 1.000000 | DISCARD |
| 2-year summary (Drug prescriptions) - VITAMIN C Min frequency (days) | 1.000000 | DISCARD |
| 2-year summary (Drug prescriptions) - VITAMIN C Prescription count | 1.000000 | DISCARD |
| 2-year summary (Drug prescriptions) - VITAMIN E Average frequency (days) | 1.000000 | DISCARD |
| 2-year summary (Drug prescriptions) - VITAMIN E Days since first prescription | 1.000000 | DISCARD |

| | | | |
|---|---|---|---|
| 2-year summary (Drug prescriptions) - VITAMIN E Days since last prescription | | 1.000000 | DISCARD |
| 2-year summary (Drug prescriptions) - VITAMIN E Max frequency (days) | | 1.000000 | DISCARD |
| 2-year summary (Drug prescriptions) - VITAMIN E Min frequency (days) | | 1.000000 | DISCARD |
| 2-year summary (Drug prescriptions) - VITAMIN E Prescription count | | 1.000000 | DISCARD |
| 2-year summary (Drug prescriptions) - VITAMIN K ANALOGUES Average frequency (days) | | 1.000000 | DISCARD |
| 2-year summary (Drug prescriptions) - VITAMIN K ANALOGUES Days since first prescription | | 1.000000 | DISCARD |
| 2-year summary (Drug prescriptions) - VITAMIN K ANALOGUES Days since last prescription | | 1.000000 | DISCARD |
| 2-year summary (Drug prescriptions) - VITAMIN K ANALOGUES Max frequency (days) | | 1.000000 | DISCARD |
| 2-year summary (Drug prescriptions) - VITAMIN K ANALOGUES Min frequency (days) | | 1.000000 | DISCARD |
| 2-year summary (Drug prescriptions) - VITAMIN K ANALOGUES Prescription count | | 1.000000 | DISCARD |
| 2-year summary (Drug prescriptions) - WART AND ANTI-CORN AGENTS Average frequency (days) | | 1.000000 | DISCARD |
| 2-year summary (Drug prescriptions) - WART AND ANTI-CORN AGENTS Days since first prescription | | 1.000000 | DISCARD |
| 2-year summary (Drug prescriptions) - WART AND ANTI-CORN AGENTS Days since last prescription | | 1.000000 | DISCARD |
| 2-year summary (Drug prescriptions) - WART AND ANTI-CORN AGENTS Max frequency (days) | | 1.000000 | DISCARD |
| 2-year summary (Drug prescriptions) - WART AND ANTI-CORN AGENTS Min frequency (days) | | 1.000000 | DISCARD |
| 2-year summary (Drug prescriptions) - WART AND ANTI-CORN AGENTS Prescription count | | 1.000000 | DISCARD |
| 2-year summary (Doctor visits) - PAEDODONTICS | | 1.000000 | DISCARD |
| 2-year summary (Doctor visits) - PERIODONTICS | | 1.000000 | DISCARD |
| 2-year summary (Doctor visits) - PROSTHODONTICS | | 1.000000 | DISCARD |
| 2-year summary (Laboratory tests) - number of tests for Glucose^2H_post_dose_glucose | | 1.000000 | DISCARD |
| 2-year summary (Laboratory tests) - days since earliest test for Glucose^2H_post_dose_glucose | | 1.000000 | DISCARD |
| 2-year summary (Laboratory tests) - days since latest test for Glucose^2H_post_dose_glucose | | 1.000000 | DISCARD |
| 2-year summary (Laboratory tests) - number of high values of Glucose^2H_post_dose_glucose | | 1.000000 | DISCARD |
| 2-year summary (Laboratory tests) - highest value of Glucose^2H_post_dose_glucose | | 1.000000 | DISCARD |
| 2-year summary (Laboratory tests) - number of low values of Glucose^2H_post_dose_glucose | | 1.000000 | DISCARD |
| 2-year summary (Laboratory tests) - lowest value of Glucose^2H_post_dose_glucose | | 1.000000 | DISCARD |
| 2-year summary (Laboratory tests) - average value of Glucose^2H_post_dose_glucose | | 1.000000 | DISCARD |
| 2-year summary (Laboratory tests) - median value of Glucose^2H_post_dose_glucose | | 1.000000 | DISCARD |
| 2-year summary (Laboratory tests) - count of normal tests for Glucose^2H_post_dose_glucose | | 1.000000 | DISCARD |
